# Auditable cross-instrument detection of unusual multivariate psychiatric response configurations using a semantically aligned covariance subspace

**DOI:** 10.64898/2026.05.22.26353902

**Authors:** Vipul Periwal

## Abstract

**Background:** Conventional psychiatric screening instruments summarize symptoms within individual scales and prioritize cases with high single-instrument additive score severity. This design treats items as independent within instruments and ignores cross-instrument covariance structure, making it insensitive to respondents whose responses are distributed across multiple domains in unusual combinations that remain below threshold on every individual scale.

**Methods:** We analyzed two cohorts spanning older and younger adults. Item prompts from depression, stress, anxiety, and sleep instruments were embedded into a shared semantic space using a pretrained sentence encoder. Principal component analysis of the *item-prompt embeddings alone*—with no use of respondent data at this stage—was used to construct a low-dimensional subspace retaining 80% of variance in the item embedding matrix. Normalized participant responses were then projected into this subspace, with Jaccard-based stability analysis used as a check on dimensional robustness. Multivariate deviation from the cohort norm was quantified with Mahalanobis distance using Ledoit-Wolf covariance regularization. Candidate outliers were defined by the empirical 95th percentile of the cohort-specific distance distribution. To isolate response configurations not already captured by conventional single-instrument extreme-value logic, we excluded all outlier respondents who had endorsed any individual item at the maximum value of its Likert scale on any instrument. For the remaining outliers, anomalous components were backtracked to their original item loadings for interpretation.

**Results:** In the older-adult Health and Retirement Study (HRS) cohort, principal component analysis of 27 item-prompt embeddings showed that a 10-dimensional subspace provided a stable representation of cross-instrument semantic structure. In the younger-adult Xinxiang cohort the corresponding stable solution was 16-dimensional. In each cohort, seven respondents remained as multivariate outliers despite falling below every single-instrument extreme-value threshold. These cases were not characterized by uniformly severe symptom scores but by unusual cross-domain response configurations that became visible only in the shared semantic covariance subspace. The response structure of the retained configurations differed across cohorts: older-adult cases more often involved weak endorsement of mood-labeled items alongside nonzero body- and sleep-related responses, whereas younger-adult cases more often involved incomplete response configurations spanning mood, sleep, stress, and self-harm-related items.

**Conclusions:** A semantically aligned, auditable covariance subspace provides a practical tool for flagging unusual multivariate response configurations that single-instrument additive screening may not flag. The method is interpretable at the level of original item contributions. It should be understood as a hypothesis-generating screen for unusual response configurations requiring further clinical assessment, not as a diagnostic instrument. Outcome validity remains to be established by prospective study.

## Introduction

Multi-instrument psychiatric batteries are typically analyzed one scale at a time and summarized through additive total scores. This strategy is clinically practical but has a structural limitation: it treats items as independent *within* instruments and provides no framework for analyzing covariance *across* instruments. A respondent may look unremarkable on each instrument separately while occupying an unusual position in the joint response space. Detecting such cases requires a common representational frame for items drawn from instruments with different wording conventions, response formats, and construct boundaries—a frame that does not currently exist in routine screening practice.

The importance of cross-instrument structure has become more visible as dimensional and transdiagnostic models of psychopathology have gained traction. The DSM remains the dominant clinical framework, yet dissatisfaction with rigid disorder-defining boundaries has motivated alternatives emphasizing symptom heterogeneity, cross-diagnostic structure, and shared latent liabilities (***American Psychiatric Association, 2013***; ***Watson, 2005***; ***Krueger and Markon, 2006***; ***Kotov et al., 2017***). Network approaches make a related point from a different direction: symptoms may form interacting structures with meaningful covariance patterns across domains, rather than serving as independent indicators of a hidden disease entity (***Borsboom and Cramer, 2013***; ***Borsboom, 2017***; ***Cramer et al., 2010***). Empirically, even within established diagnoses, patients can express markedly different symptom constellations while obtaining similar total scores (***Fried and Nesse, 2015***).

These observations motivate cross-instrument analysis, but they do not resolve the alignment problem. Depression, perceived stress, anxiety, and sleep disruption instruments overlap in content but use different wording, different Likert formats, and different response anchors. Mapping items manually across instruments requires expert judgment, is difficult to audit, and does not generalize when new instruments are added. The tool introduced here addresses this problem by using pretrained sentence embeddings as an automated alignment layer. Items are placed into a shared representational space according to semantic proximity, responses are projected into that space, and Mahalanobis distance in the resulting low-dimensional subspace flags respondents whose joint configuration is globally unusual relative to cohort structure.

The present analysis applies this tool to two cohorts spanning distinct life stages and cultural milieus. The primary purpose is to demonstrate that the pipeline recovers a small coherent set of respondents whose response configurations are invisible to single-instrument screening. We refer to these as *pan-mild* cases—respondents whose responses are distributed across multiple domains at low-to-moderate intensity rather than concentrated in a single acute cluster—as a descriptive label for the geometric category the tool targets, not as a clinical category.

The subthreshold domain is not clinically trivial. Longitudinal work has shown that low-grade symptom burdens are not benign simply because they do not qualify as full disorder (***Judd et al., 1996, 1998, 2000***), and atypical or masked presentations are recognized sources of false negatives in routine care (***Katon and Schulberg, 1992***; ***Mitchell et al., 2009***). Whether the cases flagged by this tool fall into that clinically meaningful class is a question for prospective validation. Establishing that the tool can flag such cases at all, and that it flags them in a geometrically stable and auditable way, is the contribution of the present paper.

## Methods

Figure 1 depicts the steps described below.

**Figure 1.**
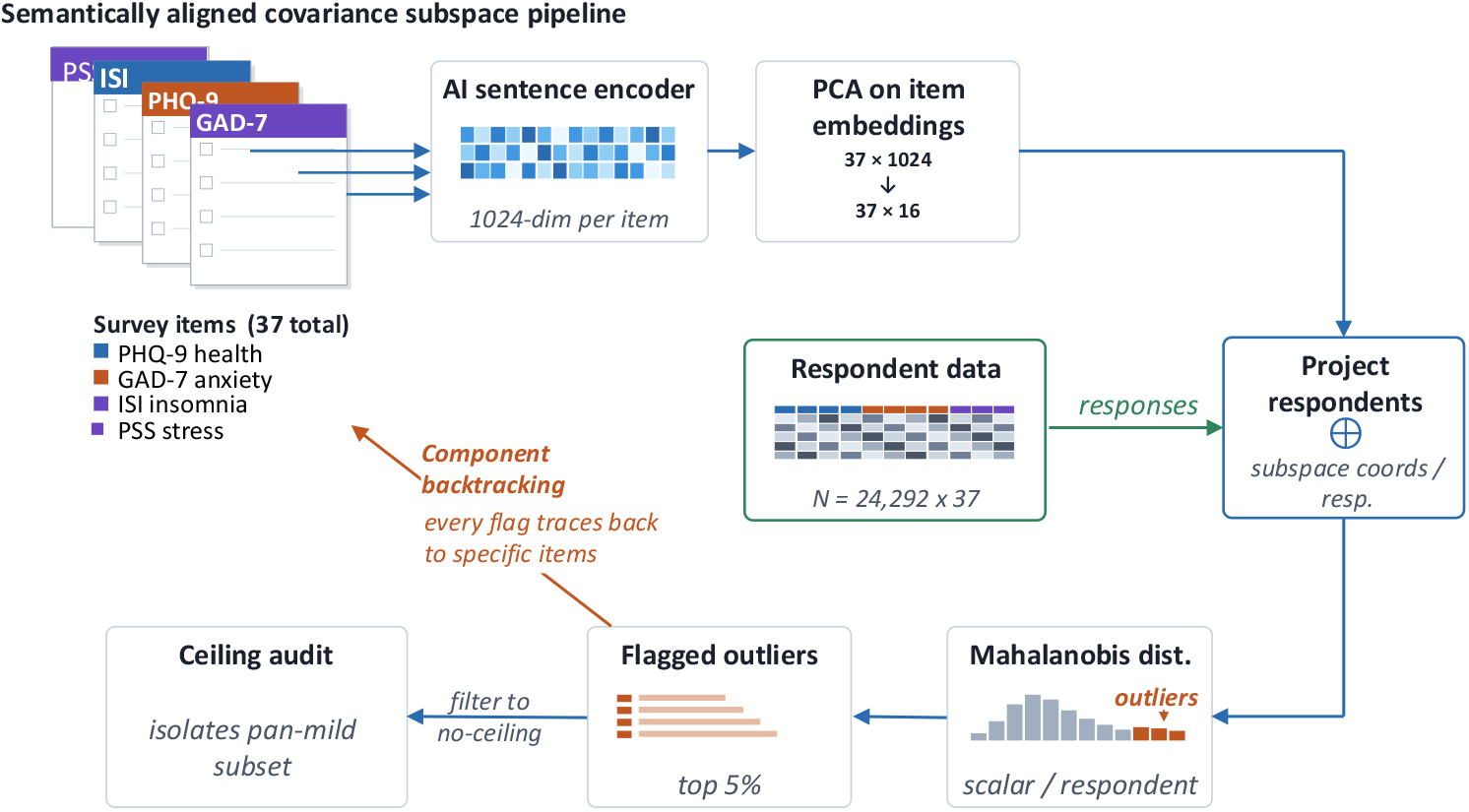
Schematic of analysis workflow

### Cohorts and measures

The primary cohort was drawn from the 2022 core wave of the HRS, a nationally representative longitudinal study of older adults in the United States (***Sonnega et al., 2014***). The analytic sample was created by deterministic linkage of respondents across available modules contributing mood-related, stress, anxiety, and sleep data. The final HRS matrix comprised 4,413 respondents and 27 aligned items drawn from four domains: the 8-item HRS form of the Center for Epidemiologic Studies Depression Scale (CES-D), the Perceived Stress Scale (PSS-10), a 5-item Beck Anxiety Inventory subset, and a 4-item sleep disruption inventory (***Radloff, 1977***; ***Steffick, 2000***; ***Cohen et al., 1983***; ***Beck et al., 1988***).

The comparison cohort was the Xinxiang sample (***Su et al., 2024***), consisting of 24,292 respondents and 37 aligned items spanning stress, insomnia, depression, anxiety, and related symptom domains (PSS-14 (***Cohen et al., 1983***), ISI-7 (***Morin et al., 2011***), GAD-7 (***Spitzer et al., 2006***), and PHQ-9 (***Kroenke et al., 2001***)). The purpose of the comparison cohort was to test whether the geometric structure of the tool’s output generalizes to a distinct measurement context and age range, not to provide external clinical validation.

### Preprocessing and score alignment

All item responses were normalized so that observed scale values lie in [−1, 1] before projection into the semantic space. Positively valenced items were reverse-scored so that higher values consistently indicated greater symptom burden. Missing values were imputed using distance-weighted *k*-nearest neighbors with *k* = 5 (recently surveyed in (***Li et al., 2024***)), chosen to preserve local response geometry more faithfully than mean imputation in heterogeneous multiscale data, as employed in analogous instrument-based measurement contexts (***Oğuztüzün et al., 2023***; ***Jiang et al., 2023***).

### Semantic alignment of questionnaire items

#### Embedding and dimensionality selection

Each questionnaire prompt was embedded using bge-m3 (***Xiao et al., 2023***), a state-of-the-art multilingual retrieval model based on a BERT-large architecture. Operating within the broader sentence-embedding framework, this encoder-only model uses contrastive learning to map natural language into a normalized 1024-dimensional semantic space following the Sentence-BERT framework and its MPNet/BERT backbone (***Devlin et al., 2019***; ***Reimers and Gurevych, 2019***; ***Song et al., 2020***). Two additional models, bge-large-en-v1.5 (***Xiao et al., 2023***) and all-mpnet-base-v2 (***Song et al., 2020***), were also used to evaluate the sensitivity of results to the choice of embedder.

Let *Q*_*r*_ ∈ ℝ^*p*×*m*^ denote the raw matrix of item embeddings for *p* items in an *m*-dimensional semantic space. We first subtracted a language-model bias vector *q* equal to the column mean over the *p* item embeddings, giving the centered matrix *Q* = *Q*_*r*_ − *q*. Principal component analysis (PCA) was then applied to the *p* rows of *Q*. This PCA operates entirely on the item-prompt embedding matrix. No respondent data are involved at this stage. The retained subspace dimensionality *D* was selected as the smallest number of principal components explaining at least 80% of the variance in the item embedding matrix. This criterion characterizes the semantic dimensionality of the question space: how many orthogonal conceptual directions are needed to represent the content of the *p* items to 80% fidelity. The result is a weight matrix *W*_*D*_ ∈ ℝ^*p*×*D*^ containing the projection of the *p* items on the *D* retained semantic components.

#### Role of the embedding layer

The embedding step is used solely to place questionnaire items into a shared representational space according to broad semantic proximity. It was not trained on the present outcomes and was not used to generate diagnoses or labels. Critically, this step does *not* perform natural language inference (NLI). NLI—the formal determination of entailment, contradiction, or neutrality between propositions—remains an open problem in computational linguistics, and pretrained sentence encoders do not solve it. Two items may occupy nearby regions of the embedding space because they concern the same conceptual domain even if they differ in polarity, response direction, or logical relationship. The embedding step should therefore be understood as a soft, data-driven substitute for manual item matching: it provides a common coordinate frame for cross-instrument analysis without requiring hand-built categorical mappings. The analysis does not interpret individual embedding dimensions, does not assume that semantic negation corresponds to vector negation, and does not perform vector arithmetic on item texts. All interpretive content derives from backtracking component scores to the original questionnaire items and observed responses.

### Respondent mapping

With the respondent-by-item matrix of normalized responses *X* ∈ ℝ^*n*×*p*^, respondents were projected into the shared semantic subspace by linear multiplication,

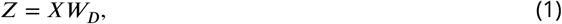

yielding a respondent-by-semantic-feature representation in which cross-instrument proximity was determined by item meaning rather than by instrument membership alone. Demographic nuisance structure (age and sex) was residualized from the manifold coordinates using ordinary least squares before multivariate distance estimation.

### Covariance-regularized multivariate deviation

To quantify deviation from the cohort norm, we computed Mahalanobis distance on the retained component scores (***Mahalanobis, 1936***). Because empirical covariance matrices become unstable in moderate- to high-dimensional settings, especially when item distributions differ in discreteness across instruments, covariance inversion was regularized using the Ledoit-Wolf shrinkage estimator (***Ledoit and Wolf, 2004, 2012***). For respondent *i* with retained component vector *y*_*i*_, distance was computed as

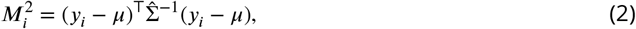

where *μ* is the cohort mean vector in retained component space and 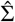 is the shrinkage-regularized covariance matrix. Dimensional stability was visualized by computing the Jaccard overlap of retained outlier sets across increasing component counts (***Ben-Hur et al., 2002***).

We did not use nominal 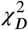 thresholds for outlier calling because bounded discrete data may be platykurtic or heavily skewed. Instead, candidate outliers were defined operationally using the empirical 95th percentile of the cohort-specific Mahalanobis distance distribution. In HRS, where the response structure included binary items, the empirical cutoff approximated the *χ*^2^(*D*, 0.99) reference for *D* = 10. In the Xinxiang cohort, composed of fully polytomous instruments, the empirical cutoff was more conservative than the nominal *χ*^2^(*D*, 0.99) reference. The 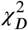 detected outlier numbers are provided in Fig. 2 for completeness. The empirical threshold (0.95) therefore served as a cohort-appropriate operational definition rather than an appeal to asymptotic idealization (***Brereton, 2015***).

**Figure 2.**
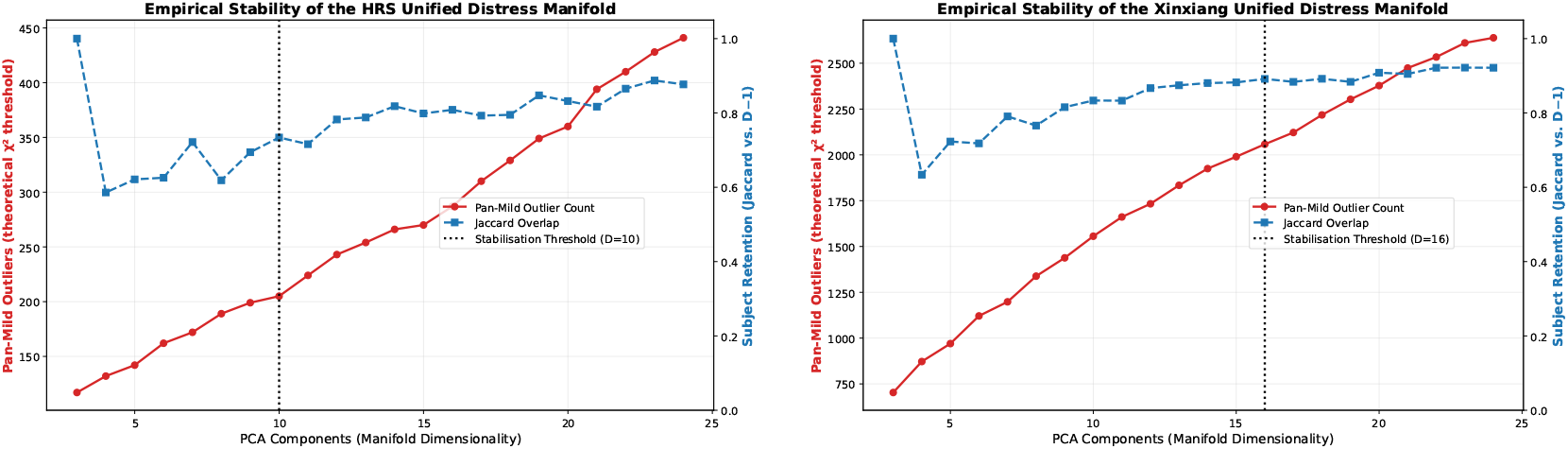
Empirical stability of the semantic psychological state subspace. Jaccard overlap between adjacent retained outlier sets and the corresponding growth in candidate outlier count across increasing PCA dimensionality applied to the item embedding matrix. The HRS cohort is on the left and the Xinxiang on the right.

### Audit against single-instrument extremes

The central aim of the analysis was not merely to identify multivariate outliers, but to isolate those whose anomaly could not be explained by isolated extreme scores on any single instrument. We therefore applied an audit layer after outlier detection.

The audit criterion is principled and uniform: a respondent is excluded if they endorsed *any* individual item at the **maximum value of the Likert scale for that instrument**. This is the most stringent single-item exclusion possible: it removes anyone who gave the most extreme available response on even one question across all four instruments. Respondents retained after this exclusion are exclusively those whose item-level profiles are nowhere at the scale ceiling, yet whose joint configuration is anomalous in the shared covariance subspace.

Concretely, for the Xinxiang cohort (PHQ-9 items 0–3; GAD-7 items 0–3; ISI items 0–4; PSS items 0–4), respondents were retained only if: max(PHQ-9) < 3, max(GAD-7) < 3, max(ISI) < 4, and max(PSS) < 4. For the HRS cohort, because the CES-D module uses binary scoring (0/1), an instrument-level sum was used (sum(CES-D) < 4), while item-level ceiling exclusions were applied to the remaining polytomous scales: max(SLEEP) < 3, max(PSS) < 5, and max(BAI) < 5.

This audit step is substantively important. Without it, multivariate outlier detection will inevitably recover many clinically obvious severe cases. The present study instead targets the residual class of cases that evade conventional scalar screening entirely. Interpretability enters only at this stage: unusually large positive or negative component scores are traced back to the specific questionnaire items and observed responses that jointly produced the respondent’s outlying position. The meaningful object is the response *configuration* associated with an outlier, not any standalone semantic meaning assigned to a direction in embedding space.

### Component backtracking and interpretability

For each retained respondent, interpretability was obtained by tracing anomalous principal component scores back to the original questionnaire items with the highest component loadings. This produced an auditable decomposition of each anomaly into the item clusters that contributed most strongly to its separation. Full item-to-component loading matrices are provided as machine-readable CSV files in the Supporting Information for all three embedding models evaluated.

### Uniform manifold approximation comparison

A standard approach to finding nonlinear features or groups in high-dimensional data is Uniform Manifold Approximation and Projection (UMAP) (***McInnes et al., 2018b***), which constructs a smooth manifold embedding using a local density objective with a neighbor-graph construction. We embedded the raw responses *X* using UMAP, as well as the semantically embedded responses *Z*, to understand the role of the semantic layer and the geometry of outlier detection in these nonlinear manifolds (***McInnes et al., 2018a***).

## Results

### Jaccard values attained stable values at the chosen *D* features in both cohorts

The semantic projection of *item prompts* produced low-dimensional subspaces in both cohorts with the 80% explained-variance criterion. Because this PCA operates on the *p* × *m* embedding matrix of item texts rather than on any respondent data, the dimensionality selected—*D* = 10 for HRS, *D* = 16 for Xinxiang—reflects the intrinsic semantic dimensionality of each instrument’s question set, not properties of the response distribution. The higher dimensionality for Xinxiang is expected: it contains more items (37 vs. 27) and a fully polytomous response structure, whereas HRS includes binary response elements.

The Jaccard overlap between outlier sets for the theoretical *χ*^2^(*D*, 0.99) threshold attained stable values at the chosen *D* for both cohorts, as shown in Fig. 2. To be more conservative and avoid inflation due to polytomous instrument usage, we applied covariance-regularized Mahalanobis distance and defined candidate outliers at the empirical 95th percentile of this distance distribution. The finalized dimensionalities *D* = 10 (HRS) and *D* = 16 (Xinxiang) are used in all downstream anomaly analyses.

### Retained cross-instrument pan-mild outliers in each cohort

After excluding all respondents who endorsed any item at the ceiling of its Likert scale, exactly seven pan-mild respondents (7/4413, 0.159%) remained in the HRS cohort and seven (7/24292, 0.029%) in the Xinxiang cohort. These fourteen cases constitute the demonstration sample for the tool. Figures 3 and 4 visualize these retained profiles as case-by-component z-score maps, with components labeled by their highest-loading item.

**Figure 3.**
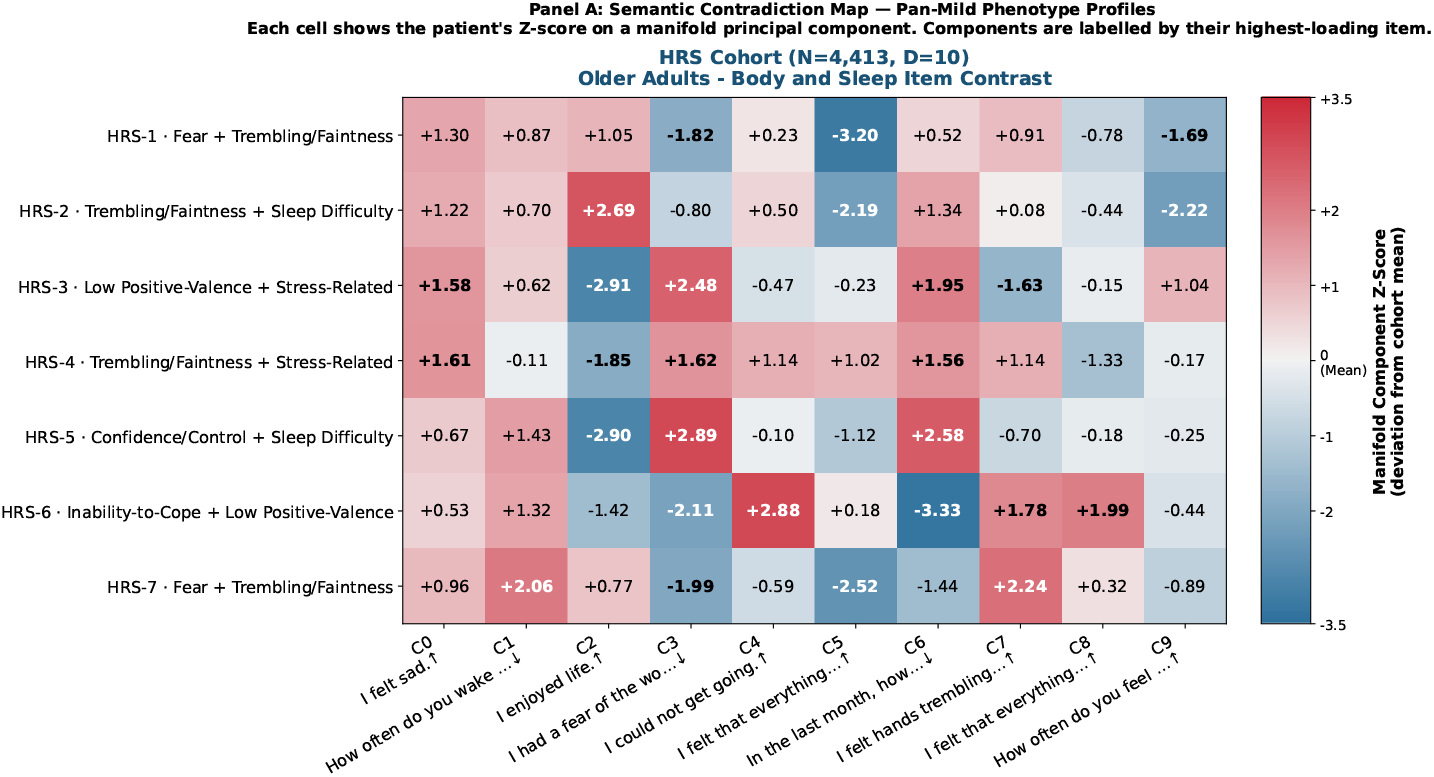
Semantic contradiction map of retained subthreshold response configurations. Heat map of respondent-level z-scores across retained subspace components. Components are labeled by their highest-loading questionnaire item. The HRS panel contains seven retained older-adult cases in a 10-dimensional subspace.

**Figure 4.**
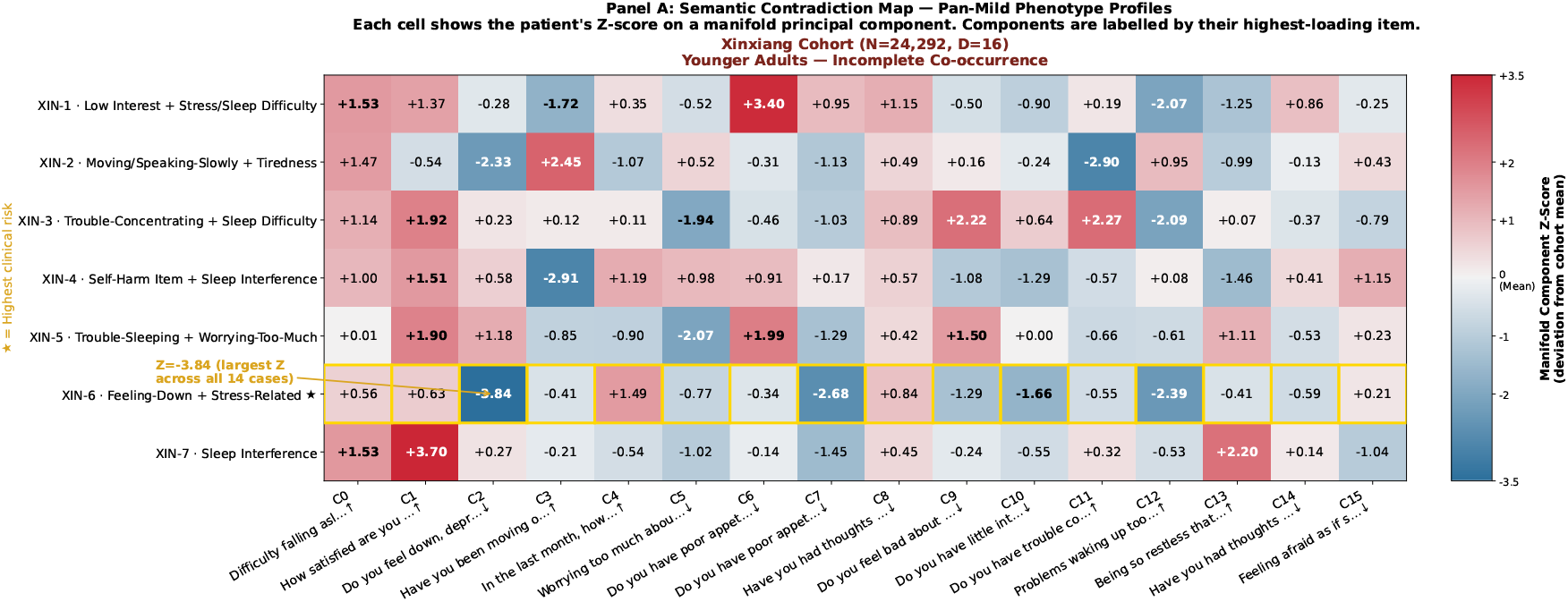
Semantic contradiction map of retained subthreshold response configurations. Heat map of respondent-level z-scores across retained subspace components. Components are labeled by their highest-loading questionnaire item. The Xinxiang panel contains seven retained younger-adult cases in a 16-dimensional subspace. The highlighted case shows the largest single-component deviation across both cohorts and is characterized by endorsement of the self-harm/better-off-dead item together with selected sleep-related items but without the broader co-occurrence pattern commonly seen alongside such items.

### Older-adult outliers: weak mood-item endorsement alongside somatic and sleep responses

The retained HRS profiles showed a consistent geometric motif: weak endorsement of items explicitly naming sadness, loneliness, or depression together with nonzero responses on body, sleep, control, or effort-related items. Fig. 3 summarizes these seven profiles as response configurations in which item clusters that often co-occur under additive scoring appear in less typical combinations. Across these cases, large component deviations tended to arise when low responses on mood-labeled items co-occurred with nonzero responses on other item groups.

The emphasis here is descriptive: the analysis identifies unusual covariance among item responses, not latent psychological states beyond those responses themselves.

### Younger-adult outliers: unusual selective co-occurrence of item groups

The Xinxiang profiles differed qualitatively. Fig. 4 summarizes this cohort as response configurations in which individual item groups were often present without the broader co-occurring pattern expected under additive scoring. Deviations were driven less by low responses on mood-labeled items and more by selective combinations of mood, sleep, stress, restlessness/bodily weakness, concentration, and self-harm-related items.

The clearest example is the highlighted Xinxiang case (XIN-6) with the largest single component deviation in the entire analysis, whose outlying position is driven by a response pattern that includes the self-harm/better-off-dead item together with low or absent responses on several restlessness/bodily-weakness and concentration items. More generally, the retained Xinxiang cases showed unusual combinations of self-harm-related, restlessness/bodily-weakness, sleep, appetite, concentration, and stress-related items—precisely the kind of response configuration that can be underweighted by additive screening thresholds.

### Cross-cohort geometric contrast

The geometric contrast is summarized in Fig. 5. In the older-adult HRS cohort, the defining structure is the co-occurrence of nonzero sleep-, body-, and control-related responses with weak responses on sadness-, loneliness-, or depression-labeled items. In the younger-adult Xinxiang cohort, the defining structure is the appearance of selected item groups—including self-harm-related, restlessness/bodily-weakness, sleep, appetite, and stress items—without the broader response pattern commonly seen alongside them under additive scoring.

**Figure 5.**
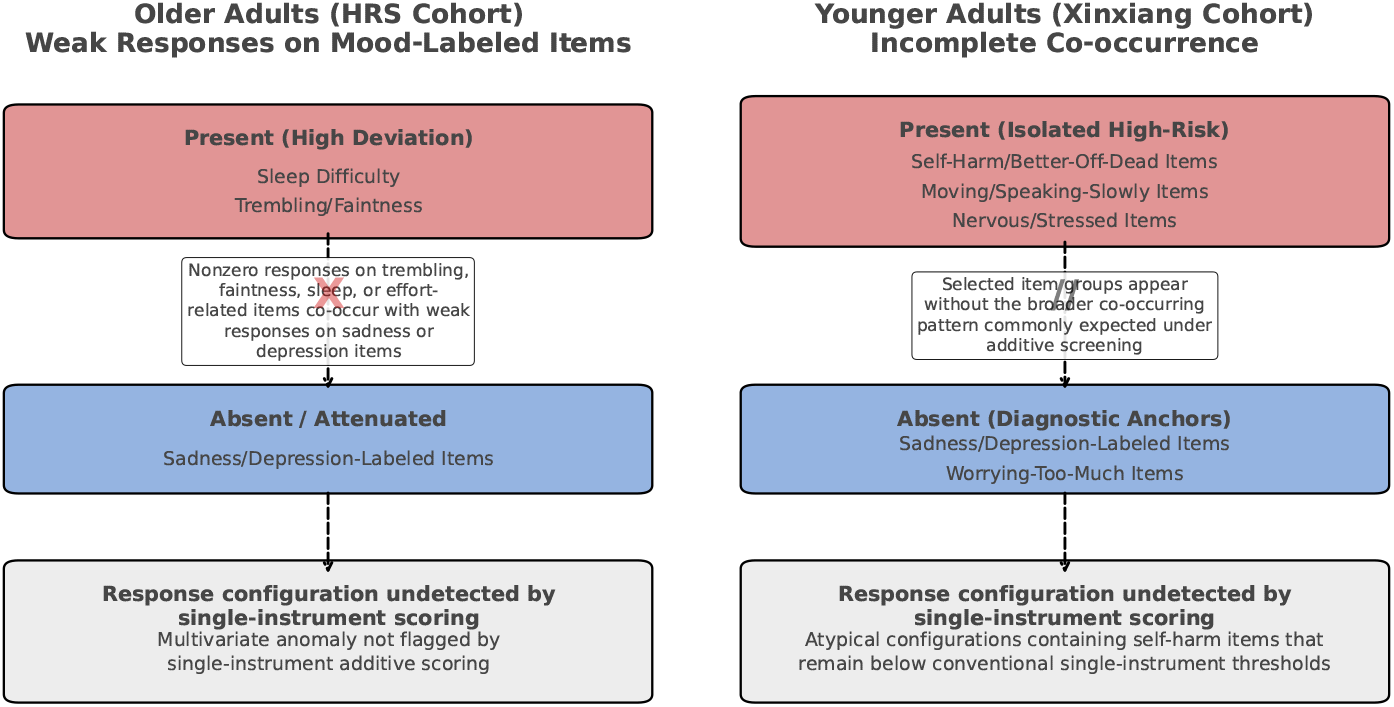
Cohort-level schematic of the geometric structure of retained response configurations. Older-adult cases are characterized by the co-occurrence of nonzero sleep- and body-related responses with weak responses on sadness-, loneliness-, or depression-labeled items. Younger-adult cases are characterized by partial co-occurrence among mood-, sleep-, stress-, restlessness/bodily weakness-, appetite-, and self-harm-related items rather than by a single dominant scale elevation. This contrast is descriptive and hypothesis-generating. Its replication in an independent sample with prospective outcomes is required before clinical interpretation is warranted.

This contrast is hypothesis-generating and should be interpreted with caution. The two cohorts differ simultaneously in age group, instrument set, response format (binary vs. fully polytomous), and cultural context. The observed structural difference cannot be attributed to any single factor.

### Visualizing semantic subspace via non-linear projection

To evaluate whether the linear semantic covariance framework captured topological features distinct from standard nonlinear dimensionality reduction, a baseline UMAP was computed directly on the raw quantitative item responses. Covariate residualization and Ledoit-Wolf regularized Mahalanobis thresholds (95th percentile) were applied to this nonlinear space. There was zero overlap between the cases flagged by the two methods for the bge-m3 model. Geometrically, the outliers defined within the UMAP projection collapsed into a single, localized cluster on the extreme topological boundary of the manifold (Fig. 6). In contrast, the cases identified by the semantic PCA pipeline were distributed across the sparse, interstitial regions of the main cohort projection for both cohorts. This structural divergence indicates that the nonlinear neighbor-graph approach prioritized local density and edge-case isolation—flagging a single, homogeneous behavioral subgroup—whereas the linear covariance framework remained sensitive to global multivariate deviations.

**Figure 6.**
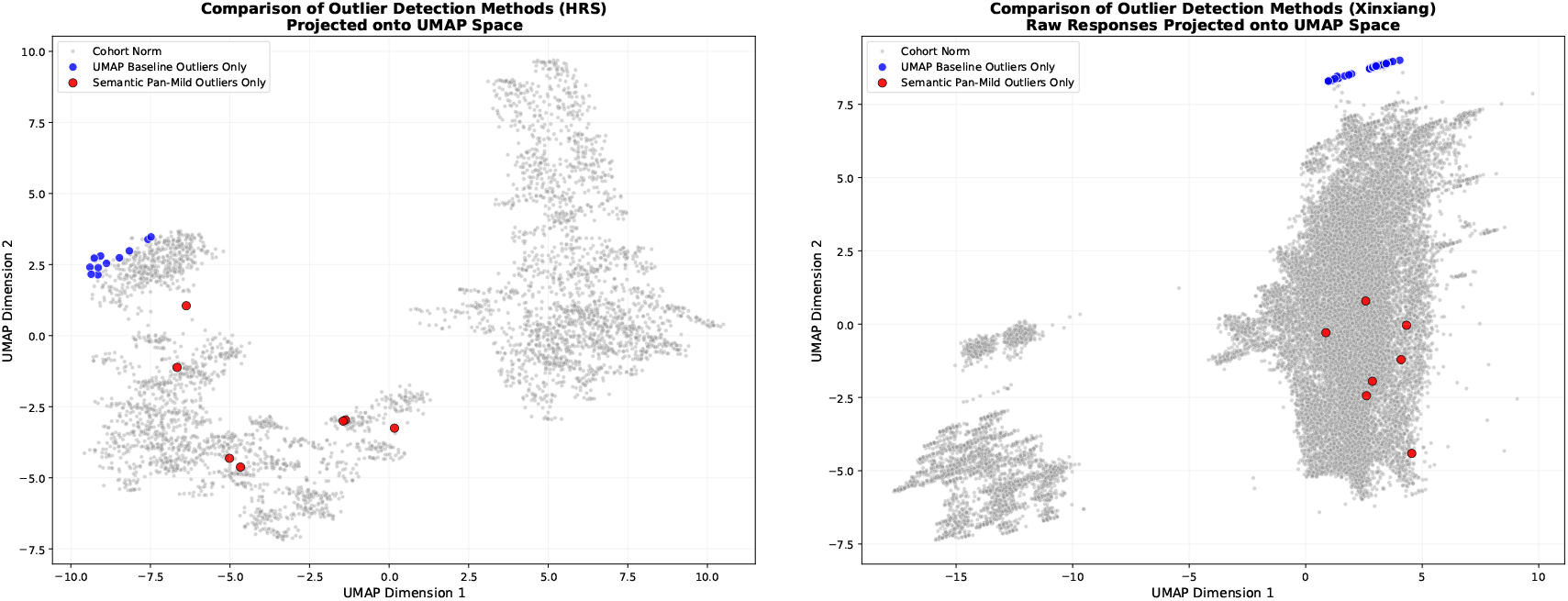
Nonlinear U MAP embeddings of raw responses show outliers which are completely distinct from the interstitial structural outliers captured by semantic alignment in the HRS (left) and Xinxiang (right) cohorts.

UMAP embeddings of the semantically embedded responses Z show a more condensed structure for both cohorts (Fig. 7): there are no Xinxiang UMAP outliers at all after semantic embedding, and only one for HRS. This compression of the nonlinear structure after semantic projection is consistent with the linear subspace having captured the dominant covariance structure of the item space.

**Figure 7.**
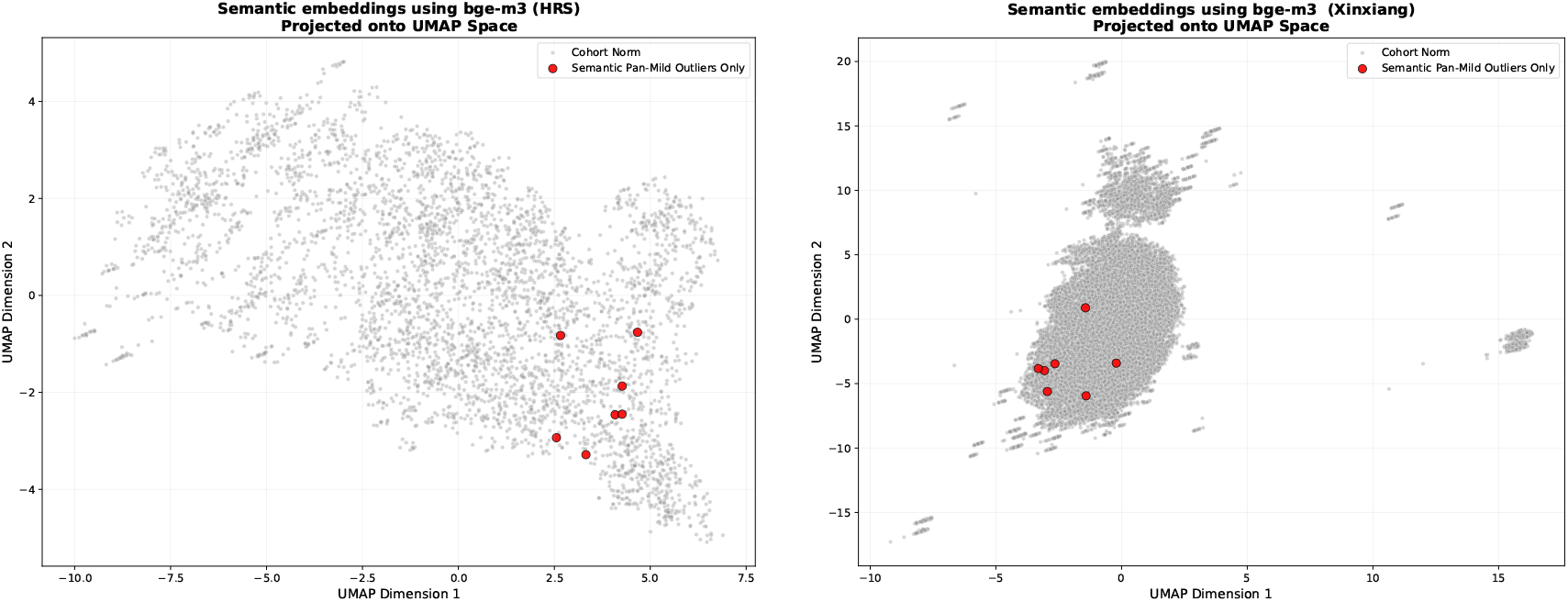
Nonlinear UMAP embeddings of semantically embedded responses with the bge-m3 embedder show one outlier for HRS and none for Xinxiang, in contrast to the interstitial structural outliers captured by semantic alignment in the HRS (left) and Xinxiang (right) cohorts.

### Comparison between different embedding models

For both cohorts and all three embedding models, the UMAP embeddings of semantically embedded responses have a smooth, similar structure with few or no UMAP-detected outliers, in contrast to the UMAP representations of the raw response data (Figs. 7,8, 9). The linear semantic covariance Mahalanobis detection produced comparable numbers of pan-mild outliers across all three embedders, with substantial pairwise overlap (Table 1).

**Table 1.** Pairwise Jaccard overlap of retained outlier sets across embedding models. Values are shown as intersection/union, with the Jaccard index in parentheses. Expected overlap under independence (hypergeometric null) is ≈ 0.01–0.002 for HRS and Xinxiang respectively, see Discussion.

| Cohort | bge-m3 vs. all-mpnet | bge-m3 vs. bge-large-en | all-mpnet vs. bge-large-en |
| --- | --- | --- | --- |
| HRS | 4/8 (0.500) | 2/10 (0.200) | 2/8 (0.250) |
| Xinxiang | 4/8 (0.500) | 5/7 (0.714) | 4/7 (0.571) |

**Figure 8.**
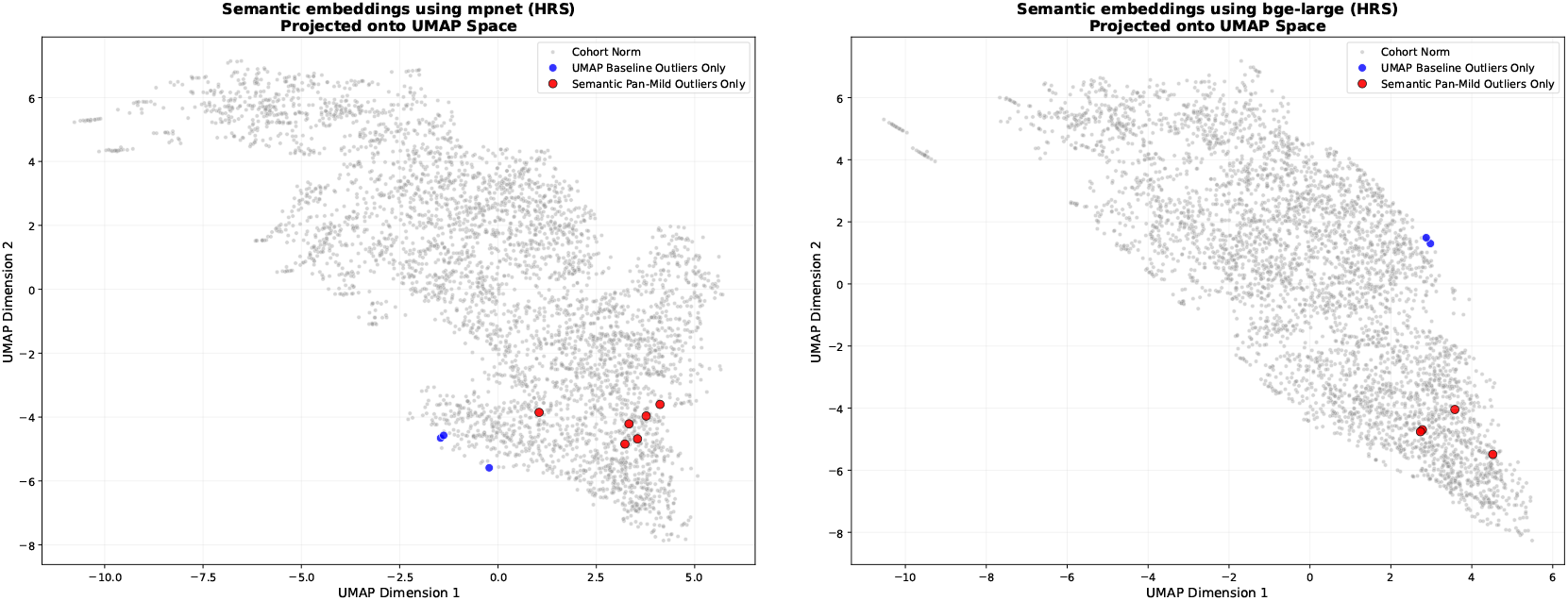
Nonlinear UMAP embeddings of semantically embedded responses with the mpnet (left) and bge-large-en-v1.5 (right) embedder show peripheral outliers (detected with UMAP) and one outlier in common with the interstitial structural outliers captured by semantic alignment in the HRS cohort using mpnet, and none using bge-large-en-v1.5.

**Figure 9.**
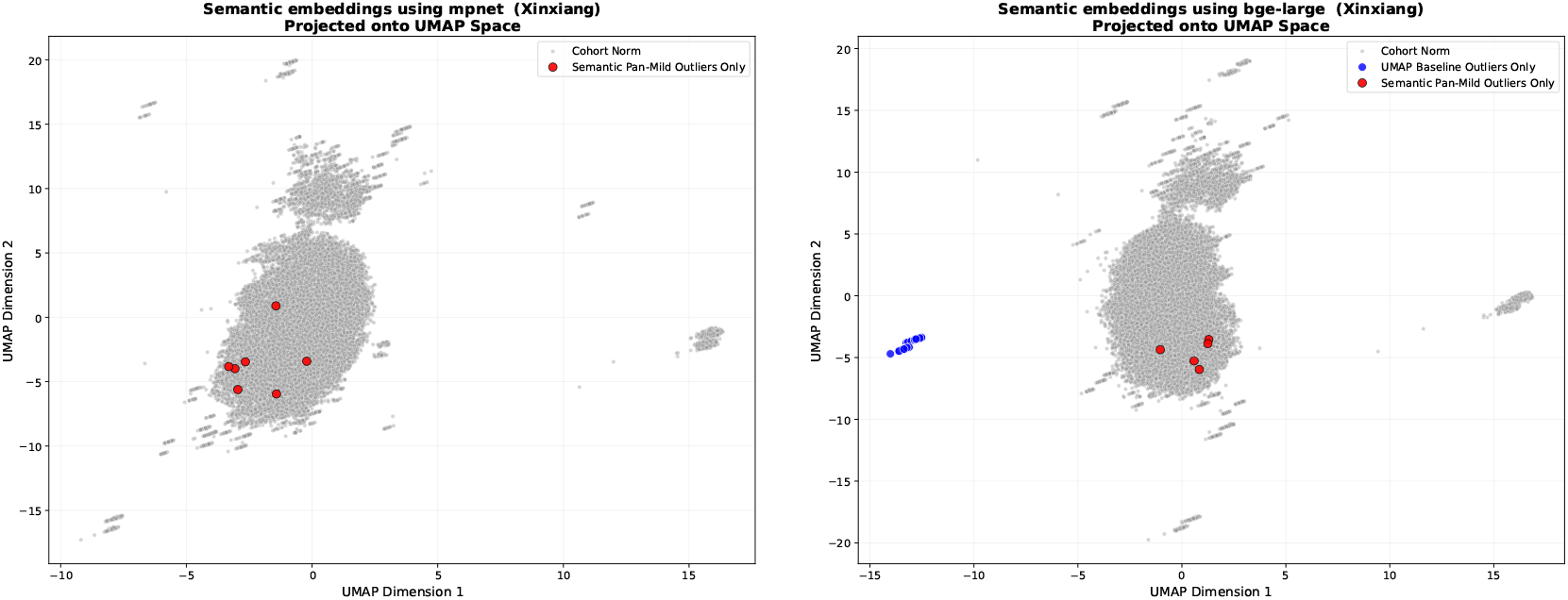
Nonlinear UMAP embeddings of semantically embedded responses with the mpnet (left) and bge-large-en-v1.5 (right) embedder show no outliers detected for the mpnet embedding and some peripheral outliers for bge-large-en-v1.5 (right), compared to the interstitial structural outliers captured by semantic alignment in the Xinxiang cohort.

The complete case profiles for the bge-m3 model are provided in Supplementary Appendix 1 (profiles for the other two models are given in Supporting Information). While the exact respondent identities fluctuate at the extreme distribution tails depending on the chosen embedding, these detailed profiles demonstrate that the qualitative response pattern—the cross-domain, uniformly sub-ceiling configuration—remains structurally consistent across all models.

## Discussion

We presented an auditable tool for cross-instrument multivariate outlier detection in multi-domain psychological assessments. The tool projects item prompts from heterogeneous instruments into a shared semantic subspace using a pretrained sentence encoder, projects respondent responses into that subspace, and applies Ledoit-Wolf regularized Mahalanobis distance to flag respondents whose joint response configuration is globally unusual relative to cohort structure. An audit layer then removes all cases whose anomaly is attributable to any single-item extreme value, isolating the residual class of respondents who evade conventional scalar screening. In both cohorts analyzed here, that residual class was non-empty and geometrically coherent.

### What the tool does

Routine psychiatric screening asks whether a person is severe on one scale. The present tool changes that question: it asks whether a person’s total response configuration is unusually structured relative to their cohort, even when nothing is extreme on any individual scale. This is a well-defined second-order question about multivariate covariance structure, and the answer to it is independent of the answer to the first-order scalar question.

### Role of the semantic layer

The semantic layer is important but its role should be understood precisely. Sentence embeddings provide a practical, automated alignment of item content across otherwise incompatible instruments—a data-driven alternative to manual item-matching—allowing joint analysis of depression, stress, anxiety, and sleep items without hand-built categorical mappings. The embedder is not a diagnostic classifier, does not perform NLI, and is not a theory of compositional meaning. Once the semantic alignment is in place, the rest of the pipeline is entirely classical: PCA for dimensional compression, Ledoit-Wolf shrinkage for stable covariance estimation, and Mahalanobis distance for multivariate deviation. This combination is interpretable at the level of original items, as required for responsible clinical AI applications (***Rudin, 2019***; ***Huys et al., 2016***).

### Dimensional selection

A feature of the present approach worth emphasizing is that dimensionality selection (*D* = 10 for HRS, *D* = 16 for Xinxiang) is determined entirely from the item embedding matrix and involves no respondent data. The 80% explained-variance criterion chooses the number of orthogonal semantic directions needed to represent the content of the item set. Respondents are then projected passively into this pre-defined space. This separation of the semantic and statistical stages avoids overfitting to the response distribution and ensures that the subspace structure is determined by question content rather than by the particular sample analyzed.

### Embedder stability: the hypergeometric argument

The pairwise Jaccard indices in Table 1 should be evaluated against the correct null expectation. Under the null hypothesis that two embedders independently select their outlier sets uniformly at random from the cohort, the expected intersection is *n*_1_*n*_2_/*N*, where *n*_1_ and *n*_2_ are the number of outliers found by each model and *N* is the cohort size. For the HRS comparison bge-m3 (*n*_1_ = 7) vs. all-mpnet (*n*_2_ ≈ 5), the expected overlap is 7 × 5/4413 ≈ 0.008 respondents. The observed intersection of 4 is approximately 500 times the null expectation. The exact hypergeometric probability *P* (*X* ≥ 4) under this null is negligibly small. For the Xinxiang comparison bge-m3 (*n*_1_ = 7) vs. bge-large-en (*n*_2_ ≈ 5), the expected overlap is 7 × 5/24292 ≈ 0.001. The observed intersection of 5 is approximately 5000 times the null expectation. Even the weakest Jaccard in the table (0.200, HRS bge-m3 vs. bge-large-en) represents an intersection far beyond chance. The low absolute Jaccard values reflect the genuine difficulty of rare-event detection at sub-0.2% prevalence rates. The departures from the null demonstrate that the overlaps are not accidental.

### Audit layer

The audit criterion—exclusion of any respondent who endorsed any item at the Likert ceiling on any instrument—is both stringent and principled. It is the most conservative possible single-item screen: it removes anyone who gave the most extreme available response anywhere across all four instruments. This ensures that retained cases are genuinely sub-ceiling on every dimension, and that the tool’s output cannot be reproduced by any combination of single-item extreme-value rules.

### What the tool does not do

The tool detects statistical atypicality. It does not make diagnoses. A retained case may reflect a clinically meaningful response configuration, an idiosyncratic reporting style, cohort-specific measurement structure, or some combination of the three. No outcome data are available here, and outcome validity is not established. The tool should be used as a flag for further clinical assessment, not as a stand-alone clinical decision. The threshold is cohort-specific and empirical, which is a strength for heterogeneous survey environments but means transportability must be demonstrated rather than assumed. The analysis is cross-sectional. The prospective question— whether flagged response configurations predict later disorder onset, help-seeking, functional decline, or other outcomes—is not addressed here and defines the primary direction for future work.

### Limitations of the geometric approach

A larger perspective might question the use of linear PCA when psychological profiles likely inhabit a more complex manifold. One might be tempted to map quantitative responses into complete sentences via the semantic layer and embed those into a nonlinear space. This cannot be done reliably, neither conceptually nor computationally. Conceptually, such a mapping would sacrifice auditability, and the colloquial strength of qualifiers is inherently regional and cultural, making the covariance structure even less transportable. Computationally, characterizing the geometry of such a nonlinear manifold precisely would require inordinate quantities of survey responses. The present approach embeds only the survey prompts to establish semantic neighborhoods and then applies classical linear machinery to respondent data—a deliberate choice of auditability over nonlinear expressiveness.

As dimensionality increases beyond the selected *D*, added principal components capture decreasing fractions of global covariance, increasingly representing item-specific noise. Ledoit-Wolf regularized Mahalanobis distance scales by inverse covariance, so inclusion of low-variance dimensions eventually inflates distances across the entire cohort and washes out structural anomalies. Consequently, the Jaccard analyses (Fig. 2) demonstrate that the 80% variance criterion successfully targets a locally stable macro-structural signal before the distance metric is overwhelmed by high-dimensional noise. They do not claim global invariance.

### Summary

Additive symptom screening remains useful but is incomplete. A semantically aligned covariance subspace does not supersede conventional instruments—it changes the question asked of them. Instead of asking only whether a person is severe on one scale, it asks whether their total configuration is unusually structured relative to their cohort. In both cohorts studied here, that question recovered a small set of respondents who would otherwise be easy to overlook. Whether those respondents are clinically significant is a question this tool alone cannot answer. Establishing that the tool can reliably surface them is the contribution of the present paper.

## Supporting information

Code for tool

## Data Availability

All data used in the present study can be ontained from the DOI URLs :https://doi.org/10.7826/WXYP6118 and https://doi.org/10.1038/s41597-024-03888-8. All code used in the analysis can be obtained from the author upon reasonable request.

https://github.com/nihcompmed/ACIDD

## Acknowledgements

This research was supported by the Intramural Research Program of the National Institute of Diabetes and Digestive and Kidney Diseases (NIDDK) within the National Institutes of Health (NIH). The contributions of the NIH author(s) are considered Works of the United States Government. The findings and conclusions presented in this paper are those of the author(s) and do not necessarily reflect the views of the NIH or the U.S. Department of Health and Human Services.

## Supplementary Appendix 1. Structured analytic summaries of retained multivariate outliers

### Scope and interpretive constraints

The case summaries below are analytic descriptions of response configurations in the finalized semantically aligned subspace. They are not clinical case formulations, diagnostic judgments, or treatment recommendations. Cases were retained only if they exceeded the cohort-specific empirical Mahalanobis threshold after Ledoit-Wolf covariance regularization *and* did not endorse any item at the Likert ceiling on any instrument. Age and sex were regressed out before subspace scoring. Respondent study identifiers have been removed in accordance with data use requirements. Cases are identified by their anonymized case labels only.

Because several questionnaire items were reverse-coded during preprocessing, interpretation is based on the joint configuration of items and component structure, rather than on any single coded response in isolation. For clarity, item labels are reported in their original questionnaire wording.

**Older-adult cohort: HRS**

**Cohort size:** 4,413

**Retained subspace dimensionality:** 10

**Retained cross-instrument outliers:** 7

#### Case HRS-1

**Mahalanobis distance:** 5.2901

**Empirical rank:** 84/4,413 (top 1.903%)

**Non-zero items:** 22/27

| Dominant com-<br>ponent | Z-score | Highest-loading items contributing to separation |
| --- | --- | --- |
| C5 | −3.202 | "I felt that everything I did was an effort."; "I felt hands trembling."; "I enjoyed life."; "I was happy."; "In the last month, how often have you been upset because of something that happened unexpectedly?" |
| C3 | −1.819 | "I had a fear of the worst happening."; "I felt lonely."; "I had a fear of dying."; "I felt sad."; "I felt depressed." |
| C9 | −1.687 | "How often do you feel really rested when you wake up in the morning?"; "In the last month, how often have you felt that things were going your way?"; "In the last month, how often have you found that you could not cope with all the things that you had to do?"; "How often do you have trouble falling asleep?"; "In the last month, how often have you felt nervous and stressed?" |

##### Analytic summary

This respondent’s outlying position was driven by nonzero responses on upset-over-unexpected-events, fear-of-the-worst-happening, fear-of-dying, hands-trembling, trouble-falling-asleep, and nervous/stressed items, together with zero responses on loneliness-, sadness-, depression-, and effort-labeled items. The positive-valence items “I enjoyed life” and “I was happy,” as well as restfulness and coping-related items, also contributed to the separation. The unusual feature is the co-occurrence of these present and absent items within the same response pattern.

#### Case HRS-2

**Mahalanobis distance:** 5.1196

**Empirical rank:** 128/4,413 (top 2.901%)

**Non-zero items:** 21/27

| Dominant component | Z-score | Highest-loading items contributing to separation |
| --- | --- | --- |
| C2 | +2.690 | "I enjoyed life."; "I was happy."; "How often do you feel really rested when you wake up in the morning?"; "I felt that everything I did was an effort."; "In the last month, how often have you felt that you were unable to control the important things in your life?" |
| C9 | -2.220 | "How often do you feel really rested when you wake up in the morning?"; "In the last month, how often have you felt that things were going your way?"; "In the last month, how often have you found that you could not cope with all the things that you had to do?"; "How often do you have trouble falling asleep?"; "In the last month, how often have you felt nervous and stressed?" |
| C5 | -2.195 | "I felt that everything I did was an effort."; "I felt hands trembling."; "I enjoyed life."; "I was happy."; "In the last month, how often have you been upset because of something that happened unexpectedly?" |

##### Analytic summary

This respondent’s outlying position was driven by nonzero responses on the positive-valence items “I enjoyed life” and “I was happy,” restfulness-on-waking, trouble-falling-asleep, things-going-your-way, upset-over-unexpected-events, and could-not-cope items, together with zero responses on the effort item. Hands-trembling also contributed at lower level. The unusual feature is the joint placement of positive-valence, sleep-related, and stress-related items together with a zero response on the effort item.

#### Case HRS-3

**Mahalanobis distance:** 5.2662

**Empirical rank:** 95/4,413 (top 2.153%)

**Non-zero items:** 22/27

| Dominant component | Z-score | Highest-loading items contributing to separation |
| --- | --- | --- |
| C2 | -2.915 | "I enjoyed life."; "I was happy."; "How often do you feel really rested when you wake up in the morning?"; "I felt that everything I did was an effort."; "In the last month, how often have you felt that you were unable to control the important things in your life?" |
| C3 | +2.478 | "I had a fear of the worst happening."; "I felt lonely."; "I had a fear of dying."; "I felt sad."; "I felt depressed." |
| C6 | +1.948 | "In the last month, how often have you been upset because of something that happened unexpectedly?"; "In the last month, how often have you felt confident about your ability to handle your personal problems?"; "I had a fear of dying."; "How often do you have trouble falling asleep?"; "In the last month, how often have you been able to control irritations in your life?" |

##### Analytic summary

This respondent’s outlying position was driven by nonzero responses on fear-of-the-worst-happening, fear-of-dying, loneliness-, sadness-, and depression-labeled items, upset-over-unexpected-events, trouble-falling-asleep, and control-related items, together with zero responses on “I enjoyed life,” “I was happy,” and the effort item. Restfulness-on-waking also contributed. The unusual feature is the co-occurrence of nonzero responses on several fear-, sadness-, and control-related items with zero responses on the positive-valence and effort items.

#### Case HRS-4

**Mahalanobis distance:** 4.9443

**Empirical rank:** 191/4,413 (top 4.328%)

**Non-zero items: 22/27**
| Dominant component | Z-score | Highest-loading items contributing to separation |
| --- | --- | --- |
| C2 | -1.847 | "I enjoyed life."; "I was happy."; "How often do you feel really rested when you wake up in the morning?"; "I felt that everything I did was an effort."; "In the last month, how often have you felt that you were unable to control the important things in your life?" |
| C3 | +1.619 | "I had a fear of the worst happening."; "I felt lonely."; "I had a fear of dying."; "I felt sad."; "I felt depressed." |
| C0 | +1.607 | "I felt sad."; "I felt lonely."; "I felt faint."; "I was happy."; "I felt depressed." |

**Non-zero items:** 22/27

| Dominant component | Z-score | Highest-loading items contributing to separation |
| --- | --- | --- |
| C2 | -2.903 | "I enjoyed life."; "I was happy."; "How often do you feel really rested when you wake up in the morning?"; "I felt that everything I did was an effort."; "In the last month, how often have you felt that you were unable to control the important things in your life?" |
| C3 | +2.893 | "I had a fear of the worst happening."; "I felt lonely."; "I had a fear of dying."; "I felt sad."; "I felt depressed." |
| C6 | +2.581 | "In the last month, how often have you been upset because of something that happened unexpectedly?"; "In the last month, how often have you felt confident about your ability to handle your personal problems?"; "I had a fear of dying."; "How often do you have trouble falling asleep?"; "In the last month, how often have you been able to control irritations in your life?" |

##### Analytic summary

This respondent’s outlying position was driven by nonzero responses on fear-of-the-worst-happening, loneliness-, depression-, and faintness-labeled items, together with zero responses on “I enjoyed life,” “I was happy,” the effort item, and the sadness item in one dominant component. Restfulness-on-waking and inability-to-control-important-things also contributed. The unusual feature is the joint placement of faintness-, fear-, and selected mood-labeled items with zero responses on positive-valence and effort items.

#### Case HRS-5

**Mahalanobis distance:** 5.1654

**Empirical rank:** 120/4,413 (top 2.719%)

**Non-zero items:** 22/27

| Dominant component | Z-score | Highest-loading items contributing to separation |
| --- | --- | --- |
| C6 | -3.325 | "In the last month, how often have you been upset because of something that happened unexpectedly?"; "In the last month, how often have you felt confident about your ability to handle your personal problems?"; "I had a fear of dying."; "How often do you have trouble falling asleep?"; "In the last month, how often have you been able to control irritations in your life?" |
| C4 | +2.877 | "I could not get going."; "In the last month, how often have you found that you could not cope with all the things that you had to do?"; "In the last month, how often have you felt nervous and stressed?"; "I had a fear of the worst happening."; "In the last month, how often have you felt difficulties were piling up so high that you could not overcome them?" |
| C3 | -2.107 | "I had a fear of the worst happening."; "I felt lonely."; "I had a fear of dying."; "I felt sad."; "I felt depressed." |

##### Analytic summary

This respondent’s outlying position was driven by nonzero responses on loneliness-, sadness-, and depression-labeled items, fear-of-dying, upset-over-unexpected-events, confidence/control-related items, and trouble-falling-asleep, together with zero responses on “I enjoyed life,” “I was happy,” and the effort item. Restfulness-on-waking also contributed. The unusual feature is the co-occurrence of nonzero responses on several sadness-, fear-, and control-related items with zero responses on the positive-valence and effort items.

#### Case HRS-6

**Mahalanobis distance:** 5.0286

**Empirical rank:** 156/4,413 (top 3.535%)

**Non-zero items:** 22/27

| Dominant component | Z-score | Highest-loading items contributing to separation |
| --- | --- | --- |
| C5 | -2.521 | "I felt that everything I did was an effort."; "I felt hands trembling."; "I enjoyed life."; "I was happy."; "In the last month, how often have you been upset because of something that happened unexpectedly?" |
| C7 | +2.237 | "I felt hands trembling."; "In the last month, how often have you been able to control irritations in your life?"; "I could not get going."; "In the last month, how often have you been upset because of something that happened unexpectedly?"; "I felt that everything I did was an effort." |
| C1 | +2.064 | "How often do you wake up too early and cannot get back to sleep?"; "How often do you wake up during the night and have trouble getting back to sleep?"; "How often do you have trouble falling asleep?"; "My sleep was restless."; "How often do you feel really rested when you wake up in the morning?" |

##### Analytic summary

This respondent’s outlying position was driven by nonzero responses on upset-over-unexpected-events, inability-to-cope, difficulties-piling-up, nervous/stressed, “I could not get going,” and fear-related items, together with zero responses on loneliness-, sadness-, and depression-labeled items. Trouble-falling-asleep and control-related items also contributed. The unusual feature is the co-occurrence of nonzero responses on several stress-, coping-, and fear-related items with zero responses on several mood-labeled items.

#### Case HRS-7

**Mahalanobis distance:** 5.2535

**Empirical rank:** 99/4,413 (top 2.243%)

**Non-zero items:** 22/27

##### Analytic summary

This respondent’s outlying position was driven by nonzero responses on hands-trembling, upset-over-unexpected-events, early-waking, night-waking, trouble-falling-asleep, restless-sleep, and restfulness-on-waking items, together with zero responses on “I could not get going” and the effort item in two dominant components. The positive-valence items “I enjoyed life” and “I was happy” also contributed. The unusual feature is the joint placement of trembling-, stress-, positive-valence-, and multiple sleep-related items together with zero responses on selected activation-related items.

#### Younger-adult cohort: Xinxiang

**Cohort size:** 24,292

**Retained subspace dimensionality:** 16

**Retained cross-instrument outliers:** 7

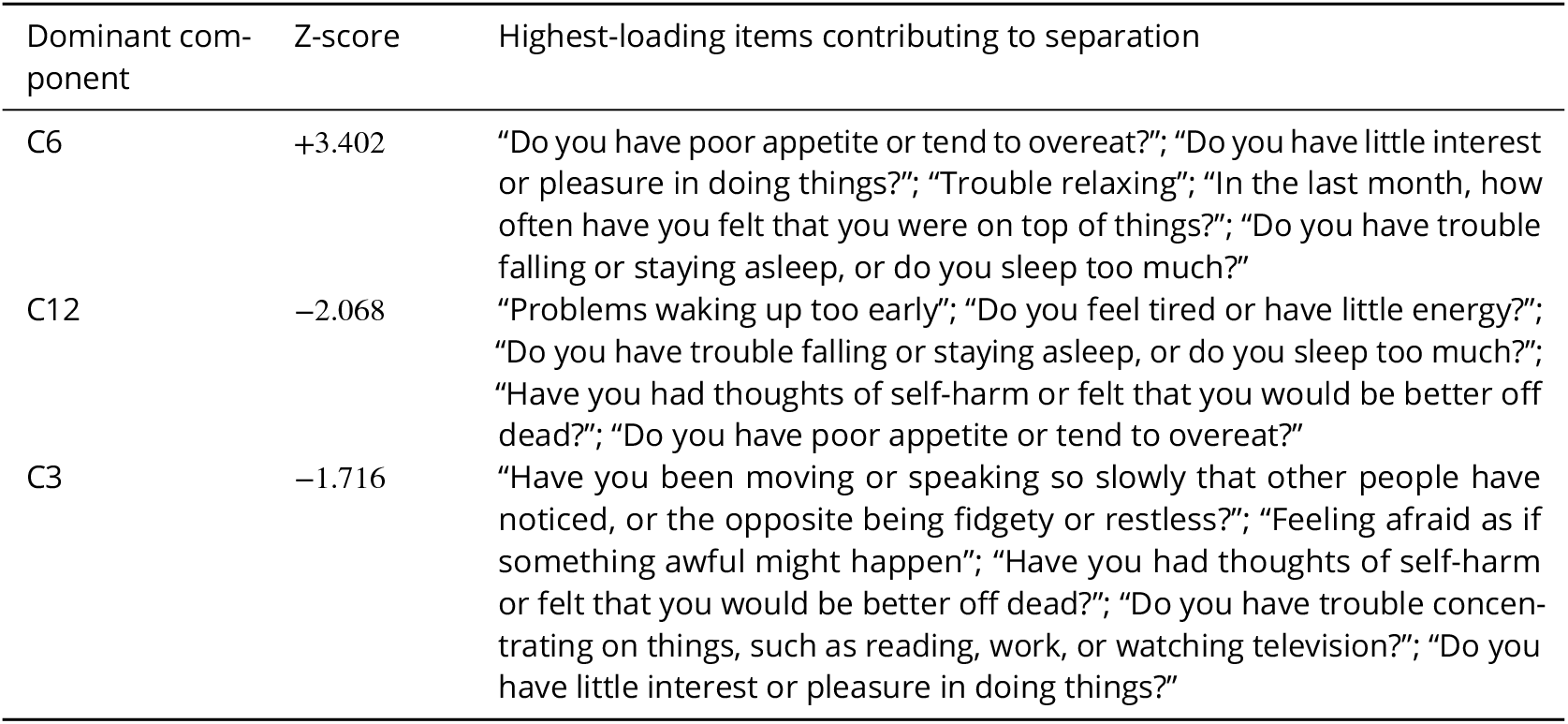

| Dominant component | Z-score | Highest-loading items contributing to separation |
| --- | --- | --- |
| C6 | +3.402 | "Do you have poor appetite or tend to overeat?"; "Do you have little interest or pleasure in doing things?"; "Trouble relaxing"; "In the last month, how often have you felt that you were on top of things?"; "Do you have trouble falling or staying asleep, or do you sleep too much?" |
| C12 | -2.068 | "Problems waking up too early"; "Do you feel tired or have little energy?"; "Do you have trouble falling or staying asleep, or do you sleep too much?"; "Have you had thoughts of self-harm or felt that you would be better off dead?"; "Do you have poor appetite or tend to overeat?" |
| C3 | -1.716 | "Have you been moving or speaking so slowly that other people have noticed, or the opposite being fidgety or restless?"; "Feeling afraid as if something awful might happen"; "Have you had thoughts of self-harm or felt that you would be better off dead?"; "Do you have trouble concentrating on things, such as reading, work, or watching television?"; "Do you have little interest or pleasure in doing things?" |

#### Case XIN-1

**Mahalanobis distance:** 6.5592

**Empirical rank:** 1048/24,292 (top 4.314%)

**Non-zero items:** 35/37

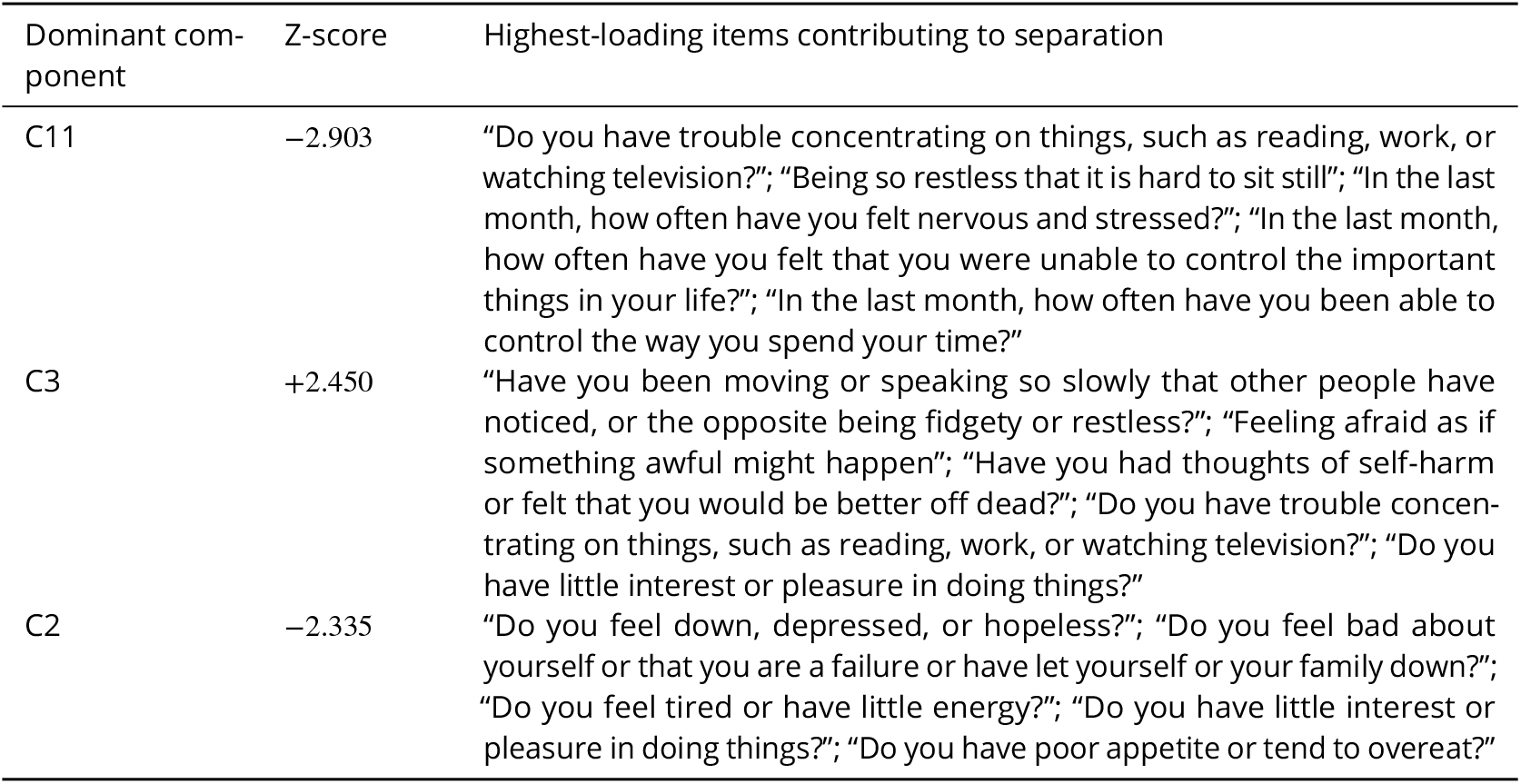

##### Analytic summary

This respondent’s outlying position was driven by nonzero responses on low interest/pleasure, trouble relaxing, trouble falling or staying asleep, tiredness/low energy, and the self-harm/better-off-dead item, together with zero responses on appetite disturbance and early waking. Additional contribution came from fear-related, concentration-related, and moving/speaking-slowly or restless items. The unusual feature is the co-occurrence of these present and absent items within the same response pattern.

#### Case XIN-2

**Mahalanobis distance:** 7.3665

**Empirical rank:** 574/24,292 (top 2.363%)

**Non-zero items:** 28/37

| Dominant component | Z-score | Highest-loading items contributing to separation |
| --- | --- | --- |
| C11 | +2.274 | "Do you have trouble concentrating on things, such as reading, work, or watching television?"; "Being so restless that it is hard to sit still"; "In the last month, how often have you felt nervous and stressed?"; "In the last month, how often have you felt that you were unable to control the important things in your life?"; "In the last month, how often have you been able to control the way you spend your time?" |
| C9 | +2.215 | "Do you feel bad about yourself or that you are a failure or have let yourself or your family down?"; "Do you feel tired or have little energy?"; "Have you been moving or speaking so slowly that other people have noticed, or the opposite being fidgety or restless?"; "Do you have poor appetite or tend to overeat?"; "Feeling nervous, anxious or on edge" |
| C12 | -2.085 | "Problems waking up too early"; "Do you feel tired or have little energy?"; "Do you have trouble falling or staying asleep, or do you sleep too much?"; "Have you had thoughts of self-harm or felt that you would be better off dead?"; "Do you have poor appetite or tend to overeat?" |

##### Analytic summary

This respondent’s outlying position was driven by nonzero responses on the moving/speaking-slowly item, tiredness/low energy, low interest/pleasure, appetite-related items, and mild restlessness, together with zero responses on concentration difficulty, fear that something awful might happen, and the self-harm/better-off-dead item. The unusual feature is the co-occurrence of these present and absent items within the same response pattern.

#### Case XIN-3

**Mahalanobis distance:** 6.6957

**Empirical rank:** 958/24,292 (top 3.944%)

**Non-zero items:** 23/37

**Non-zero items: 29/37**
| Dominant component | Z-score | Highest-loading items contributing to separation |
| --- | --- | --- |
| C3 | -2.911 | "Have you been moving or speaking so slowly that other people have noticed, or the opposite being fidgety or restless?"; "Feeling afraid as if something awful might happen"; "Have you had thoughts of self-harm or felt that you would be better off dead?"; "Do you have trouble concentrating on things, such as reading, work, or watching television?"; "Do you have little interest or pleasure in doing things?" |
| C1 | +1.506 | "How satisfied are you with your current sleep pattern?"; "To what extent do you consider your sleep problem to interfere with your daily functioning?"; "How noticeable to others do you think your sleep problem is in terms of impairing the quality of your life?"; "Feeling afraid as if something awful might happen"; "Becoming easily annoyed or irritable" |
| C13 | -1.460 | "Being so restless that it is hard to sit still"; "Worrying too much about different things"; "In the last month, how often have you dealt successfully with day-to-day problems and annoyances?"; "Do you feel bad about yourself or that you are a failure or have let yourself or your family down?"; "In the last month, how often have you felt that you were unable to control the important things in your life?" |

##### Analytic summary

This respondent’s outlying position was driven by nonzero responses on concentration difficulty, nervous/stressed items, sleep difficulty, mild appetite-related items, and mild anxious/on-edge items, together with weak or zero responses on restlessness, self-reproach, tiredness/low energy, moving/speaking-slowly, the self-harm/better-off-dead item, and early waking. The unusual feature is the co-occurrence of these present and absent items within the same response pattern.

#### Case XIN-4

**Mahalanobis distance:** 6.3804

**Empirical rank:** 1197/24,292 (top 4.928%)

**Non-zero items:** 29/37

##### Analytic summary

This respondent’s outlying position was driven by endorsement of the self-harm/better-off-dead item together with subjective sleep dissatisfaction, sleep interference, and noticeability of the sleep problem, while moving/speaking-slowly, fear-that-something-awful-might-happen, concentration difficulty, restlessness, and worrying-too-much items remained weak or absent. The unusual feature is the co-occurrence of the self-harm/better-off-dead item with selected sleep-related items and weak responses on several other item groups.

#### Case XIN-5

**Mahalanobis distance:** 6.3590

**Empirical rank:** 1214/24,292 (top 4.998%)

**Non-zero items:** 22/37

| Dominant component | Z-score | Highest-loading items contributing to separation |
| --- | --- | --- |
| C5 | -2.071 | "Worrying too much about different things"; "How noticeable to others do you think your sleep problem is in terms of impairing the quality of your life?"; "Do you have trouble concentrating on things, such as reading, work, or watching television?"; "Difficulty falling asleep"; "Difficulty staying asleep" |
| C6 | +1.992 | "Do you have poor appetite or tend to overeat?"; "Do you have little interest or pleasure in doing things?"; "Trouble relaxing"; "In the last month, how often have you felt that you were on top of things?"; "Do you have trouble falling or staying asleep, or do you sleep too much?" |
| C1 | +1.900 | "How satisfied are you with your current sleep pattern?"; "To what extent do you consider your sleep problem to interfere with your daily functioning?"; "How noticeable to others do you think your sleep problem is in terms of impairing the quality of your life?"; "Feeling afraid as if something awful might happen"; "Becoming easily annoyed or irritable" |

##### Analytic summary

This respondent’s outlying position was driven by nonzero responses on worrying-too-much items, sleep-problem noticeability, sleep-related interference, and mild low-interest/pleasure items, to-gether with weak or zero responses on difficulty falling asleep, difficulty staying asleep, trouble relaxing, fear-that-something-awful-might-happen, and irritability. The unusual feature is the co-occurrence of these present and absent items within the same response pattern.

#### Case XIN-6

**Mahalanobis distance:** 7.2424

**Empirical rank:** 629/24,292 (top 2.589%)

**Non-zero items:** 24/37

| Dominant component | Z-score | Highest-loading items contributing to separation |
| --- | --- | --- |
| C2 | -3.837 | "Do you feel down, depressed, or hopeless?"; "Do you feel bad about yourself or that you are a failure or have let yourself or your family down?"; "Do you feel tired or have little energy?"; "Do you have little interest or pleasure in doing things?"; "Do you have poor appetite or tend to overeat?" |
| C7 | -2.676 | "Do you have poor appetite or tend to overeat?"; "Becoming easily annoyed or irritable"; "In the last month, how often have you been upset because of something that happened unexpectedly?"; "Do you have trouble concentrating on things, such as reading, work, or watching television?"; "Have you had thoughts of self-harm or felt that you would be better off dead?" |
| C12 | -2.392 | "Problems waking up too early"; "Do you feel tired or have little energy?"; "Do you have trouble falling or staying asleep, or do you sleep too much?"; "Have you had thoughts of self-harm or felt that you would be better off dead?"; "Do you have poor appetite or tend to overeat?" |

##### Analytic summary

This respondent’s outlying position was driven by nonzero responses on depressed/hopeless, self-reproach, tiredness/low energy, low interest/pleasure, appetite-related items, upset over unexpected events, trouble sleeping, and the self-harm/better-off-dead item, together with weak or zero responses on irritability, concentration difficulty, and early waking. This respondent also exhibited the most extreme single-component deviation among the retained Xinxiang cases (*Z* = −3.837 on Component 2), which is why it is highlighted in the heatmap. The unusual feature is the co-occurrence of these present and absent items within the same response pattern.

#### Case XIN-7

**Mahalanobis distance:** 6.3878

**Empirical rank:** 1192/24,292 (top 4.907%)

**Non-zero items:** 29/37

##### Analytic summary

This respondent’s outlying position was driven by nonzero responses on subjective sleep dissatisfaction, sleep interference, sleep-problem noticeability, and difficulty falling asleep, with smaller contributions from restlessness, worrying-too-much items, and self-reproach. In contrast, fear-that-something-awful-might-happen, difficulty staying asleep, and early waking were weak or absent. The unusual feature is the co-occurrence of these present and absent items within the same response pattern.

#### Cross-case summary

Across the retained cases, the dominant signal was not uniformly high symptom severity but unusual cross-domain response configurations. In HRS, several respondents combined nonzero fear-related, trembling/faintness-related, and sleep-related items with weak or zero responses on sadness-, loneliness-, and depression-labeled items. In Xinxiang, several respondents combined selected mood-, sleep-, stress-, concentration-, movement/restlessness-, appetite-, or self-harm-related items without the broader co-occurrence pattern that would be more strongly prioritized by additive screening. In all cases the anomaly is geometric—a position unusual in the joint covariance space—rather than scalar.

| Dominant component | Z-score | Highest-loading items contributing to separation |
| --- | --- | --- |
| C1 | +3.698 | "How satisfied are you with your current sleep pattern?"; "To what extent do you consider your sleep problem to interfere with your daily functioning?"; "How noticeable to others do you think your sleep problem is in terms of impairing the quality of your life?"; "Feeling afraid as if something awful might happen"; "Becoming easily annoyed or irritable" |
| C13 | +2.200 | "Being so restless that it is hard to sit still"; "Worrying too much about different things"; "In the last month, how often have you dealt successfully with day-to-day problems and annoyances?"; "Do you feel bad about yourself or that you are a failure or have let yourself or your family down?"; "In the last month, how often have you felt that you were unable to control the important things in your life?" |
| C0 | +1.533 | "Difficulty falling asleep"; "Difficulty staying asleep"; "In the last month, how often have you felt that you were effectively coping with important changes that were occurring in your life?"; "In the last month, how often have you felt that things were going your way?"; "Problems waking up too early" |

