## Supplementary material for "Auditable cross-instrument detection of unusual multivariate psychiatric response configurations using a semantically aligned covariance subspace": Xinxiang_cases_bgelarge.txt

Patient ID : 68402

Patient ID : 69862

Patient ID : 70606

Patient ID : 74389

Patient ID : 77053

=================================================================
CASE STUDY EXTRACTION — Xinxiang (N=24,292, D=16)
=================================================================

Patient ID : 68402
  Mahalanobis distance  : 6.8659
  Empirical rank        : 867 (top 3.569% of cohort)
  Raw item responses    : non-zero on 35 of 37 items

  Anomalous semantic axes (|Z| ≥ 1.0):

  ── Component  8  Z = +3.759  (above manifold mean) ──
  Item                W_k   Response  Question
  -------------- --------  ---------  -----------------------------------
  isi_q3          -0.2922          0  ▽ Problems waking up too early
  phq9_q8         -0.2519          1  ▽ Have you been moving or speaking so slowly that other p…
  phq9_q9         -0.2465          1  ▽ Have you had thoughts of self-harm or felt that you wou…
  isi_q6          +0.2393          2  ▲ How worried or distressed are you about your current sl…
  phq9_q7         +0.1821          2  ▲ Do you have trouble concentrating on things, such as re…

  ── Component  9  Z = +3.659  (above manifold mean) ──
  Item                W_k   Response  Question
  -------------- --------  ---------  -----------------------------------
  phq9_q9         +0.2667          1  ▲ Have you had thoughts of self-harm or felt that you wou…
  isi_q3          -0.2483          0  ▽ Problems waking up too early
  phq9_q5         -0.2271          0  ▽ Do you have poor appetite or tend to overeat?
  gad7_q4         +0.2212          2  ▲ Trouble relaxing
  phq9_q1         +0.1856          2  ▲ Do you have little interest or pleasure in doing things…

  ── Component 14  Z = -1.579  (below manifold mean) ──
  Item                W_k   Response  Question
  -------------- --------  ---------  -----------------------------------
  phq9_q6         -0.2849          2  ▲ Do you feel bad about yourself or that you are a failur…
  phq9_q9         +0.1814          1  ▽ Have you had thoughts of self-harm or felt that you wou…
  pss_q13         -0.1721          3  ▲ In the last month, how often have you been able to cont…
  pss_q8          +0.1419          3  ▽ In the last month, how often have you found that you co…
  pss_q4          +0.1284          3  ▽ In the last month, how often have you dealt successfull…

  ────────────────────────────────────────────────────────────

Patient ID : 69862
  Mahalanobis distance  : 7.5994
  Empirical rank        : 497 (top 2.046% of cohort)
  Raw item responses    : non-zero on 28 of 37 items

  Anomalous semantic axes (|Z| ≥ 1.0):

  ── Component  3  Z = -2.181  (below manifold mean) ──
  Item                W_k   Response  Question
  -------------- --------  ---------  -----------------------------------
  phq9_q7         -0.4394          0  ▲ Do you have trouble concentrating on things, such as re…
  phq9_q1         -0.3646          1  ▲ Do you have little interest or pleasure in doing things…
  phq9_q5         -0.2732          1  ▲ Do you have poor appetite or tend to overeat?
  gad7_q7         +0.2677          0  ▽ Feeling afraid as if something awful might happen
  pss_q13         -0.2595          3  ▲ In the last month, how often have you been able to cont…

  ── Component  5  Z = +2.123  (above manifold mean) ──
  Item                W_k   Response  Question
  -------------- --------  ---------  -----------------------------------
  gad7_q6         +0.3081          2  ▲ Becoming easily annoyed or irritable
  pss_q14         -0.2944          1  ▽ In the last month, how often have you felt difficulties…
  phq9_q7         -0.2385          0  ▽ Do you have trouble concentrating on things, such as re…
  isi_q1          -0.2304          0  ▽ Difficulty falling asleep
  phq9_q8         +0.2240          2  ▲ Have you been moving or speaking so slowly that other p…

  ── Component 12  Z = -2.022  (below manifold mean) ──
  Item                W_k   Response  Question
  -------------- --------  ---------  -----------------------------------
  gad7_q5         -0.2207          1  ▲ Being so restless that it is hard to sit still
  phq9_q7         +0.2196          0  ▽ Do you have trouble concentrating on things, such as re…
  phq9_q9         +0.2027          0  ▽ Have you had thoughts of self-harm or felt that you wou…
  pss_q14         -0.1829          1  ▲ In the last month, how often have you felt difficulties…
  pss_q3          +0.1506          1  ▽ In the last month, how often have you felt nervous and …

  ────────────────────────────────────────────────────────────

Patient ID : 70606
  Mahalanobis distance  : 6.5885
  Empirical rank        : 1076 (top 4.429% of cohort)
  Raw item responses    : non-zero on 23 of 37 items

  Anomalous semantic axes (|Z| ≥ 1.0):

  ── Component 11  Z = -2.578  (below manifold mean) ──
  Item                W_k   Response  Question
  -------------- --------  ---------  -----------------------------------
  phq9_q5         -0.2860          1  ▲ Do you have poor appetite or tend to overeat?
  phq9_q3         -0.2215          2  ▲ Do you have trouble falling or staying asleep, or do yo…
  isi_q3          +0.1988          0  ▽ Problems waking up too early
  phq9_q1         +0.1812          0  ▽ Do you have little interest or pleasure in doing things…
  pss_q1          +0.1430          2  ▽ In the last month, how often have you been upset becaus…

  ── Component  9  Z = -2.499  (below manifold mean) ──
  Item                W_k   Response  Question
  -------------- --------  ---------  -----------------------------------
  phq9_q9         +0.2667          0  ▽ Have you had thoughts of self-harm or felt that you wou…
  isi_q3          -0.2483          0  ▲ Problems waking up too early
  phq9_q5         -0.2271          1  ▲ Do you have poor appetite or tend to overeat?
  gad7_q4         +0.2212          0  ▽ Trouble relaxing
  phq9_q1         +0.1856          0  ▽ Do you have little interest or pleasure in doing things…

  ── Component  8  Z = +2.466  (above manifold mean) ──
  Item                W_k   Response  Question
  -------------- --------  ---------  -----------------------------------
  isi_q3          -0.2922          0  ▽ Problems waking up too early
  phq9_q8         -0.2519          0  ▽ Have you been moving or speaking so slowly that other p…
  phq9_q9         -0.2465          0  ▽ Have you had thoughts of self-harm or felt that you wou…
  isi_q6          +0.2393          2  ▲ How worried or distressed are you about your current sl…
  phq9_q7         +0.1821          2  ▲ Do you have trouble concentrating on things, such as re…

  ────────────────────────────────────────────────────────────

Patient ID : 74389
  Mahalanobis distance  : 6.6015
  Empirical rank        : 1066 (top 4.388% of cohort)
  Raw item responses    : non-zero on 29 of 37 items

  Anomalous semantic axes (|Z| ≥ 1.0):

  ── Component  3  Z = +3.234  (above manifold mean) ──
  Item                W_k   Response  Question
  -------------- --------  ---------  -----------------------------------
  phq9_q7         -0.4394          0  ▽ Do you have trouble concentrating on things, such as re…
  phq9_q1         -0.3646          1  ▽ Do you have little interest or pleasure in doing things…
  phq9_q5         -0.2732          0  ▽ Do you have poor appetite or tend to overeat?
  gad7_q7         +0.2677          0  ▲ Feeling afraid as if something awful might happen
  pss_q13         -0.2595          3  ▽ In the last month, how often have you been able to cont…

  ── Component  9  Z = +2.226  (above manifold mean) ──
  Item                W_k   Response  Question
  -------------- --------  ---------  -----------------------------------
  phq9_q9         +0.2667          1  ▲ Have you had thoughts of self-harm or felt that you wou…
  isi_q3          -0.2483          2  ▽ Problems waking up too early
  phq9_q5         -0.2271          0  ▽ Do you have poor appetite or tend to overeat?
  gad7_q4         +0.2212          1  ▲ Trouble relaxing
  phq9_q1         +0.1856          1  ▲ Do you have little interest or pleasure in doing things…

  ── Component  5  Z = -2.127  (below manifold mean) ──
  Item                W_k   Response  Question
  -------------- --------  ---------  -----------------------------------
  gad7_q6         +0.3081          0  ▽ Becoming easily annoyed or irritable
  pss_q14         -0.2944          3  ▲ In the last month, how often have you felt difficulties…
  phq9_q7         -0.2385          0  ▲ Do you have trouble concentrating on things, such as re…
  isi_q1          -0.2304          2  ▲ Difficulty falling asleep
  phq9_q8         +0.2240          0  ▽ Have you been moving or speaking so slowly that other p…

  ────────────────────────────────────────────────────────────

Patient ID : 77053
  Mahalanobis distance  : 7.1749
  Empirical rank        : 694 (top 2.857% of cohort)
  Raw item responses    : non-zero on 24 of 37 items

  Anomalous semantic axes (|Z| ≥ 1.0):

  ── Component  2  Z = +3.662  (above manifold mean) ──
  Item                W_k   Response  Question
  -------------- --------  ---------  -----------------------------------
  phq9_q6         +0.3501          2  ▲ Do you feel bad about yourself or that you are a failur…
  phq9_q2         +0.3394          1  ▲ Do you feel down, depressed, or hopeless?
  phq9_q1         +0.3370          1  ▲ Do you have little interest or pleasure in doing things…
  phq9_q4         +0.3206          2  ▲ Do you feel tired or have little energy?
  gad7_q4         -0.3055          0  ▽ Trouble relaxing

  ── Component  6  Z = +3.000  (above manifold mean) ──
  Item                W_k   Response  Question
  -------------- --------  ---------  -----------------------------------
  phq9_q5         +0.2721          2  ▲ Do you have poor appetite or tend to overeat?
  pss_q12         -0.2640          3  ▽ In the last month, how often have you found yourself th…
  isi_q5          -0.2402          1  ▽ How noticeable to others do you think your sleep proble…
  phq9_q8         -0.2308          1  ▽ Have you been moving or speaking so slowly that other p…
  pss_q1          +0.2289          3  ▲ In the last month, how often have you been upset becaus…

  ── Component  7  Z = -2.006  (below manifold mean) ──
  Item                W_k   Response  Question
  -------------- --------  ---------  -----------------------------------
  phq9_q8         -0.3808          1  ▲ Have you been moving or speaking so slowly that other p…
  isi_q4          +0.2154          1  ▽ How satisfied are you with your current sleep pattern?
  pss_q3          -0.2044          1  ▲ In the last month, how often have you felt nervous and …
  pss_q13         +0.2023          2  ▽ In the last month, how often have you been able to cont…
  gad7_q2         +0.1936          0  ▽ Not being able to stop or control worrying

  ────────────────────────────────────────────────────────────

=================================================================
End of case studies — Xinxiang (N=24,292, D=16)
================================================================
