## Supplementary material for "Auditable cross-instrument detection of unusual multivariate psychiatric response configurations using a semantically aligned covariance subspace": Code for tool: allofus_expert_review.md

# All of Us — Expert Review (2026-06-09)

Three independent expert reviews were run before locking the item set and building
the generate step: **psychometrics / survey methodology** (item curation),
**biostatistics / methods** (fidelity to the manuscript + statistical soundness),
and **code correctness** (converter + reproducibility). This doc records every
finding, what was already fixed, and the decisions still open.

Reviewed artifacts: `allofus_item_inventory.csv` (then 968 items / 356 in-scope),
`convert_allofus.py`, `allofus_preprocessing.md`, `allofus_integration_plan.md`,
the manuscript (`main_final.pdf`), and the source codebook xlsx.

**Outcome:** mechanical fixes (§A) + item-set rulings (§B) applied → **item set
locked at 282 in-scope**. Remaining work is analysis-design (§C), to be resolved
in the Researcher Workbench before/at the generate stage.

Reproducibility was independently confirmed: re-running the converter reproduced
the committed CSV byte-for-byte; all stated invariants held; a 12-item sample
cross-checked against the xlsx matched on every field; dedup keep-first was
verified safe (no collapsed concept differs in category/embed/field_type, and the
COPE-wave wording differences are immaterial).

## A. Fixed (encoded in `convert_allofus.py`, inventory regenerated → 342 in-scope)

1. **[BLOCKER] Free-entry wrapper items are not ordinal (14 in-scope items).**
   `radio` items whose options are "Enter age/number/response" + sentinels carry no
   scale points; order-coding them is meaningless. Now mechanically detected →
   category `free_entry`, always out. Items: `ipaq_7`,
   `mhqukb_25/26/28/48/50/51/52/53`, `smoking_averagedailycigarette`,
   `smoking_currentdailycigarette`, `smoking_dailysmokestartingage`,
   `smoking_numberofyears`, `attemptquitsmoking_completelyquit`. (356 → **342**.)
2. **[BLOCKER] `n_levels` counted sentinel options for 100/342 in-scope items.**
   Sentinels ("Prefer not to answer", "Don't know", PMI codes) sit at the END of
   the REDCap option order, so order-coding would make refusal the scale maximum.
   Added `n_levels_substantive` (sentinel-stripped count; sentinel = option code
   starting `pmi` or label Don't-know / Prefer-not-to-answer / Skip). The generated
   `scales.csv` must use substantive levels for `min/max` and emit the sentinel
   codes per item.
3. **[MAJOR] `section` column was the literal string `"nan"` for 926/968 rows**
   (NaN section headers stringified), which also silently disabled section-text
   keyword matching (verified: zero categorization changes from the fix). Fixed.
4. **[MINOR] Slider `n_levels` counted anchors, not levels** (0–10 slider → 2).
   Now parsed from numeric endpoints (→ 11). No in-scope item affected.
5. **[MINOR] Dead `PREFIX_CATEGORY` entries** (`cssrs`, `du`, `pss`, `sense`)
   removed; the converter now warns if any mapped prefix matches zero items
   (codebook-drift tripwire).
6. **[MINOR] Documentation drift** (stale 964/356 counts, `ss→wellbeing` in two
   docs vs the implemented `ss→psychiatric`, missing categories in the Step 6
   embed=false list) reconciled across `allofus_preprocessing.md` and
   `allofus_integration_plan.md`.

## B. Item-set decisions — RESOLVED (2026-06-09, applied → 282 in-scope)

1. **[MAJOR] Twin instruments — RESOLVED: drop COPE copies (53 items).** GAD-7,
   PHQ-9, CPSS, EDS, MOS-SS, UCLA loneliness each appeared in COPE *and* a home
   survey (EHH/SDOH). Kept the home-survey copy (full scales, no COVID framing, no
   wave repetition); dropped the COPE twin via `DUPLICATE_DROP` keyed `(survey,
   prefix)` → `duplicate_cope`. (Note the EDS scales even differed: COPE 4-level vs
   SDOH 6-level — another reason to keep the standalone SDOH version.)
2. **[MAJOR] Nominal / non-monotone — RESOLVED: drop 3, recode 2.** Dropped
   `mhqukb_11/14/15` (→ `nominal`, no valid scale). Kept `ace_5` and `mhqukb_27`
   as clean binaries by treating the off-axis option ("Parents not married";
   "Not applicable") as a sentinel (`EXTRA_SENTINEL_CODES`; `mhqukb_27`'s already
   carried a PMI code) → `n_levels_substantive = 2`.
3. **[MAJOR] Factual-history items — RESOLVED: drop 8.** The 7 `mhqukb_31_*`
   "Did <drug> help?" items (→ `med_response`) and `nhs_covid_fhc17a` (→ `covid`).
4. **[MAJOR] Inclusion/exclusion inconsistencies — RESOLVED: add 4.** Added the 3
   PROMIS Global mental-health items (`overallhealth_emotionalproblem7days`,
   `_generalmentalhealth` → psychiatric; `_socialsatisfaction` → psychosocial) and
   `livingsituation_stablehouseconcern` (housing-instability worry → psychosocial),
   via `ITEM_OVERRIDE`. (The COPE-smoking vs e-nicotine inconsistency was left
   as-is: the COPE combustible items stay out as part of the twin/COVID-wave drop.)

Still **deferred to generate stage** (not item-set lock):
- **[MINOR] Religiosity anchors:** `bmmrs`/`nhs` items append "I do not believe in
  God / I am not religious" after "Never" — decide sentinel vs recode-to-Never.
- **[MINOR] Set-composition caveats** (document in methods, no change): rare-event
  binaries (`ss_1/2/3`) are high-leverage under [−1,1] min-max; lifetime-history
  items mix with past-2-week state scales; `ies_r_6_*` wording is COVID-anchored;
  the activity domain is now only the 3 `ipaq_1/3/5` screeners.

## C. Open decisions — response-matrix / analysis design (before generate)

1. **[BLOCKER] Cross-survey structural missingness.** The manuscript cohorts (HRS
   4,413×27; Xinxiang 24,292×37) were single co-administered batteries with
   essentially complete matrices; KNN(k=5) imputation filled item-level gaps only.
   The 342 items span 6 surveys taken by overlapping-but-different participants —
   almost no participant will have all items. The pipeline's defaults
   (`min_complete_fraction=0.50` + KNN) would silently impute entire instruments a
   participant never took, fabricating exactly the cross-instrument covariance the
   method measures (a no-silent-fallback violation). **Required:** in-workbench,
   measure the participant completeness distribution over the 342 items, then
   decide the analysis unit (jointly-completed survey subset vs. high-overlap
   complete-case restriction) and set `min_complete_fraction` deliberately.
2. **[MAJOR] COPE wave selection.** COPE has many repeated waves; dedup keep-first
   silently picks a wording wave, and the plan does not say which wave supplies
   responses. **Required:** explicit rule (e.g. first completed wave per
   participant), and pin the embedded prompt wording to the same wave as the
   analyzed responses (auditability: the basis must match what participants saw).
3. **[MAJOR] Branching-logic skips ≠ refusal sentinels.** 124/342 in-scope items
   (pre-exclusions) sit behind branching logic; a gated item is structurally
   not-applicable, not nonresponse, and must not be KNN-imputed. Deep follow-up
   chains (≥2 levels, e.g. cidi5/mhqukb episode modules) have tiny effective N —
   consider excluding. **Required:** decode `Branching Logic` per item; treat
   gated-skip as out-of-scope-for-participant, refusal/don't-know as sentinel→NaN.
4. **[MAJOR] Numeric coding validation.** `scales.csv` min/max must be derived
   from the in-workbench answer-code mapping (not from `n_levels` alone), with a
   hard-error validation that observed values fall in `[min,max]`.
5. **[MAJOR] Dimensionality.** With 342 semantically diverse items, the 80%
   variance threshold will yield D far above the paper's 10–16; combined with
   completeness attrition, N/D may be unfavorable. **Required:** report actual D
   and N at each completeness threshold; run the Jaccard stability analysis; be
   prepared to narrow scope rather than absorb instability.
6. **[MINOR, decided] Weights = all-1s** (volunteer cohort, no probability
   weights; matches manuscript HRS/Xinxiang, contrasts deliberately with NHIS).
7. **[MINOR] Residualization covariates.** Specify the AoU source fields for age
   (from DOB) and sex (sex-at-birth vs gender identity), and the handling of
   refused/missing/non-binary values, before generate.
8. **[MINOR] Pan-mild / ceiling audit.** Populate the `ceiling` allowlist
   deliberately (symptom-ceiling items only); decide how the 33 binary items are
   audited (instrument-level sum, as the manuscript did for CES-D, vs item max).
9. **[MINOR] Valence/direction column.** Response direction varies across kept
   items (Yes-first binaries vs frequency scales, ascending vs descending). Add a
   per-item `reverse`/`direction` column to the inventory and review it as part of
   the lock, not after.
