## Supplementary material for "Auditable cross-instrument detection of unusual multivariate psychiatric response configurations using a semantically aligned covariance subspace": Code for tool: allofus_integration_plan.md

# All of Us Integration Plan

Status: **converter written; categorization reviewed (§6b) + expert panel review +
item-set rulings applied (2026-06-09, see
[allofus_expert_review.md](allofus_expert_review.md)); item set LOCKED at 282.
Remaining opens are analysis-design (review doc §C), not item-set.**

Goal: run All of Us survey data through the `survey-semantics` pipeline. All of Us
is inherently **cross-instrument** (many surveys), which is exactly what the method
targets — but the codebook is large and mixed, so item **curation** is central.

## 1. Source: the codebook

`data/allofus/All_of_Us_Survey_Data_Codebooks.xlsx` (outside the repo, not
committed). It is a **REDCap data dictionary**, very different from the NHIS
replication script.

- **12 survey sheets** (one row per item): `Basics`, `Lifestyle`, `Overall Health`,
  `Family Health History`, `Personal Medical History`, `Personal and Family Health
  Hist`, `Healthcare Access and Utilizati`, `Social Determinants of Health`,
  `COPE`, `Life Functioning`, `Emotional Health History`, `Behavioral Health`.
- **6 scoring sheets**: `COPE Scoring`, `Overall Health Scoring`, `Lifestyle
  Scoring`, `Social Determinants of Health S`, `Emotional Health Scoring`,
  `Behavioral Health Scoring` — columns `Measure | Question and Response Display |
  Source Value | PMI Code`.

### Survey-sheet columns we use
| Codebook column | Use |
|---|---|
| `Item Concept` | the **item key** (e.g. `gad7_1`). Stable, matches workbench data. |
| `Field Label` | the **question wording** → `prompts.csv` (embedded). |
| `Field Type` | embed filter: `radio`/`dropdown`/`slider` = ordinal candidate; `checkbox`/`text`/`descriptive` = exclude. |
| `Choices, Calculations, OR Slider Labels` | ordered response options → number of levels / scale. |
| `Form Name` | the instrument/survey name (namespacing, grouping). |
| `Section Header` | grouping; `descriptive` rows are headers/intros (no data). |
| `Branching Logic` | skip logic (informational). |

### Field Type counts (across the 12 survey sheets, pre-dedup)
`radio` 1434 · `checkbox` 312 · `text` 255 · `descriptive` 49 · `dropdown` 3 · `slider` 1.
→ Only the ordinal `radio`/`dropdown`/`slider` items are embed candidates (1438
pre-dedup), and not even all of those are Likert symptom scales (some are
categorical/yes-no, and some `radio` items are wrappers around free-text entry —
detected and excluded as `free_entry`, see preprocessing doc).

### Wrinkles specific to this codebook
- **Choice codes are *concept codes*, not numbers.** e.g. GAD-7 choices are
  `ehhwb_8, Not at all | ehhwb_9, Several days | ehhwb_10, More than half of the
  days | ehhwb_11, Nearly every day`. The **ordinal value comes from option order**
  (4 options → a 4-level scale), not from the code. The numeric coding the
  *response data* uses (0–3 GAD score vs source value) must be confirmed and
  matched in `scales.csv`.
- **PMI codes** (Skip / Prefer Not To Answer / Don't Know) live in the scoring
  sheets and `Generalized Answer Codes` → these become per-item **sentinels**.
- **Reverse valence** is not labeled per item; it must be derived from instrument
  convention / the scoring sheets (the method reverse-scores positive-valence items).

## 2. The `embed` column (tool change) — DONE

Because the codebook has ~980 ordinal items spanning 12 surveys, we added an
explicit per-item **`embed`** flag so curation is auditable and reversible:

- New optional column in `scales.csv`: `item,min,max,sentinels,reverse,ceiling,embed`.
- **Semantics:** an item is embedded *and* analyzed iff `embed=true`. `embed=false`
  (and blank, when the column is used) items remain documented but excluded.
- **Default:** column absent → all declared items embedded (pre-`embed` behavior).
- **Implemented (2026-06-09):** `ItemScale.embed` (`scales.py`); selection helpers
  `scales_use_embed` / `is_item_embedded`; pipeline item-selection restricts to
  `embed=true` ([`pipeline.py`](../src/survey_semantics/pipeline.py)); the `embed`
  CLI subcommand learns `--scale-file`/`--scale-dir` and embeds only the marked
  items ([`cli.py`](../src/survey_semantics/cli.py)). Parallels the `ceiling`
  allowlist. Tests: `tests/test_scales_weights.py`, `tests/test_pipeline.py`.

## 2b. Categorization & scope (decided)

**Scope = broad across all surveys, restricted to psychological + habit items.**
Each ordinal item gets a `category` assigned by rules (auditable):
1. **Instrument prefix** (primary): `gad→anxiety`, `phq→depression`, `pcl/ies/ace→trauma`,
   `cidi/mhqukb/ukmh/ss→psychiatric`, `bfi→personality`, `asrs→adhd`, `cpss→stress`,
   `brcs/lot→wellbeing`, `smoking/alcohol/tsu/audit→substance`, `ipaq→activity`,
   `eds/mos/nds/bmmrs/scns/hvs/nhs/ucla/sdoh→psychosocial`, `disability→functioning`;
   physical-system / demographic / access / `mentalhealth`(diagnosis) / `ips`
   (built environment; `ips_16` kept by exact override) → out.
2. **Section/label keywords** (fallback). Items whose choices are free-text
   wrappers ("Enter age | Don't know") → `free_entry`, always out.

`embed = category ∈ {anxiety, depression, trauma, psychiatric, personality, adhd,
stress, wellbeing, substance, activity, psychosocial, functioning}`.

**Counts (deduplicated — unique item concepts).** "ordinal" = `radio`/`dropdown`/
`slider` only (1438 candidates). The same Item Concept recurs across sheets/COPE
waves/survey versions; the pipeline needs unique items, so the converter dedups by
concept (keep first occurrence in codebook order) — **470 collapsed → 968 unique →
282 in-scope** (after categorization review §6b, the expert-panel mechanical fixes,
and the four item-set rulings §6c — see
[allofus_preprocessing.md](allofus_preprocessing.md) for the exact, manuscript-grade
procedure). No sheet is skipped by name.

| Survey | in-scope | ordinal | Contributes (by category) |
|---|---:|---:|---|
| Emotional Health History | 79 | 92 | psychiatric, trauma, depression, anxiety, wellbeing |
| Social Determinants of Health | 69 | 78 | psychosocial (discrimination, support, loneliness, cohesion, disorder) |
| COPE | 57 | 158 | substance, psychosocial, trauma, activity (twin GAD-7/PHQ-9/CPSS/EDS/MOS/UCLA dropped) |
| Behavioral Health | 42 | 50 | psychiatric, personality 15, adhd 6 |
| Lifestyle | 25 | 30 | substance 25 |
| Basics | 7 | 25 | functioning 6 (WHODAS) + housing-worry |
| Overall Health | 3 | 20 | PROMIS mental-health (added) |
| Healthcare Access / Personal Medical History / Family + Personal-and-Family Health Hist | 0 | (remainder) | — |
| **Total** | **282** | **968** | |

Note: Life Functioning's 6 disability items are duplicates of Basics' 6 (counted
once, under Basics). Category totals (sum to 282): psychosocial 71, psychiatric 70,
substance 66, trauma 23, personality 15, depression 9, anxiety 8, functioning 6,
adhd 6, wellbeing 5, activity 3.

Rolled into the four domain groups: **Psychological 136** (psychiatric, depression,
anxiety, trauma, stress, wellbeing, personality, adhd) · **Psychosocial 71** ·
**Habit/behavioral 69** (substance 66 + activity 3) · **Functioning 6**.

Inventory artifact: `data/allofus/allofus_item_inventory.csv` (968 rows) —
columns `survey, item, form, section, field_type, prompt, n_levels,
n_levels_substantive, choices, category, embed`. **This is the human-review file:
adjust `embed`/`category` here** (`n_levels_substantive` excludes sentinel options
— PMI codes / Don't-know / Prefer-not-to-answer — which are not scale points).

## 3. Phased plan

1. **Preprocess** — `examples/allofus/convert_allofus.py` parses the xlsx into a
   flat **item inventory** (one row per survey item): `survey, item, prompt,
   field_type, n_levels, choices, sentinels, embed(suggested)`. Suggested
   `embed = field_type in {radio,dropdown,slider}` AND has ordered choices.
2. **Curate** — review the inventory; set `embed` true/false (and pick which
   surveys/instruments are in scope). This is the human decision point.
3. **Generate** — emit `prompts.csv` and `scales.csv` (with `embed`, `reverse`,
   `sentinels`, `min/max`, `ceiling`) for the curated set.
4. **Embed** (tool) → `items.npz` for the `embed=true` items.
5. **Analyze** (tool) — **inside the Researcher Workbench**, where the responses
   live (controlled access; participant data cannot be exported). The converter
   and `items.npz`/`prompts`/`scales` are prepared here; `responses.csv` is built
   in-workbench from the source values.

## 4. Open decisions (need your call)

1. ~~**Scope.**~~ DECIDED: broad cross-instrument, psychological + habit (§2b).
2. **Numeric coding.** What values does the workbench response export use for these
   items (0-based instrument score, source value, or raw concept code)? `scales.csv`
   `min/max` must match it, and observed values must be validated against `[min,max]`
   before normalization (hard error otherwise — no silent fallback).
3. **Reverse valence.** Per-item source of truth — instrument convention, the
   scoring sheets, or a manual reverse list?
4. **Weights.** DECIDED: All of Us is a volunteer cohort with no design-based
   probability weights → all-`1`s (unweighted; WLS=OLS, identical ranking). This
   matches the manuscript's HRS/Xinxiang handling and deliberately contrasts with
   NHIS (probability sample, `WTFA_A`).
5. **`embed` vs `include` naming**, and `embed` in `scales.csv` vs a standalone manifest.
6. **Expert-review decisions** (item set + response-matrix design): see
   [allofus_expert_review.md](allofus_expert_review.md) — notably cross-survey
   structural missingness / COPE wave selection / twin-instrument duplication.

## Out of scope (for now)
- `checkbox` multi-select items, free `text`, and `descriptive` headers.
- The validated-instrument **sum scores** in the scoring sheets (GAD-7 sum, etc.) —
  the tool works on item-level responses, not precomputed sums.
