## Supplementary material for "Auditable cross-instrument detection of unusual multivariate psychiatric response configurations using a semantically aligned covariance subspace": Code for tool: allofus_preprocessing.md

# All of Us — Item Preprocessing (Methods)

Precise, reproducible record of how the All of Us survey **codebook** was processed
into the curated item set used by the pipeline. Written for a manuscript methods
section. The authoritative implementation is
[`examples/allofus/convert_allofus.py`](../examples/allofus/convert_allofus.py);
this document describes exactly what it does and the resulting counts.

## Source & software

- **Input:** `All_of_Us_Survey_Data_Codebooks.xlsx` — the All of Us survey codebook,
  a REDCap data dictionary (one Excel sheet per survey, one row per item).
- **Software:** Python 3.11, `pandas` + `openpyxl` (xlsx reader). The procedure is
  fully deterministic (no randomness, no network, no LLM at this stage).
- **Output:** `allofus_item_inventory.csv` — a flat, per-item inventory (one row per
  unique item) with a rule-assigned `category` and an `embed` flag. This file is the
  reviewable artifact; the analyzed item set is its `embed = true` rows.

## Step 1 — Sheet selection

The workbook has 20 sheets. A sheet is treated as a **survey** iff it contains the
REDCap item columns `Item Concept`, `Field Type`, and `Field Label`. This yields
**12 survey sheets**: Basics, Lifestyle, Overall Health, Family Health History,
Personal Medical History, Personal and Family Health Hist, Healthcare Access and
Utilization, Social Determinants of Health, COPE, Life Functioning, Emotional
Health History, Behavioral Health.

The other **8 sheets are excluded automatically** (they lack those columns): `ReadMe`,
`Minute Survey on COVID-19 Vacci` (a non-REDCap layout), and the 6 scoring sheets
(`COPE Scoring`, `Overall Health Scoring`, `Lifestyle Scoring`, `Social Determinants
of Health S`, `Emotional Health Scoring`, `Behavioral Health Scoring`). No sheet is
excluded by name.

## Step 2 — Item-type filter (ordinal candidates)

Only items whose `Field Type` is **`radio`, `dropdown`, or `slider`** are retained —
these are the single-choice ordinal items that admit a Likert-style numeric scale.
Field-type counts across the 12 survey sheets (pre-dedup):

| radio | checkbox | text | descriptive | dropdown | slider |
|---:|---:|---:|---:|---:|---:|
| 1434 | 312 | 255 | 49 | 3 | 1 |

Excluded by this filter: `checkbox` (multi-select, not ordinal), `text` (free text),
`descriptive` (section headers/instructions, no response). Retained ordinal
candidates: **1438**.

## Step 3 — Field extraction

For each retained item, the following are recorded:

| Inventory column | Codebook source |
|---|---|
| `item` | `Item Concept` (the stable item key, e.g. `gad_7_1`) |
| `prompt` | `Field Label` (question wording; whitespace-normalized) |
| `survey` | sheet name |
| `form` | `Form Name` |
| `section` | `Section Header` (whitespace-normalized; blank when absent — REDCap puts the header only on a section's first item) |
| `field_type` | `Field Type` |
| `n_levels` | number of response options in `Choices, Calculations, OR Slider Labels` (format `code, label \| code, label`); for `slider`, parsed from the numeric endpoint anchors (e.g. 0–10 → 11) |
| `n_levels_substantive` | `n_levels` minus **sentinel options** — options whose code starts with `pmi` or whose label is Don't-know / Prefer-not-to-answer / Skip. Sentinels are not scale points; they must become per-item sentinel codes in `scales.csv`, never the scale max. |
| `choices` | the raw choices string (for review) |

## Step 4 — Deduplication

The same `Item Concept` can appear in multiple sheets or survey waves. The pipeline
requires unique item columns, so items are **deduplicated by `Item Concept`, keeping
the first occurrence in codebook (sheet, then row) order.** **470 duplicate concept
occurrences were collapsed**, leaving **968 unique items**.

Three sources of duplication were identified:
1. **Cross-survey shared items** — e.g. the 6 WHODAS `disability_*` items appear in
   both *Basics* and *Life Functioning* (kept under Basics).
2. **Repeated-wave instruments** — e.g. `gad_7_*` / `phq_9_*` recur across COPE
   COVID survey waves with minor wording differences.
3. **Survey-version overlap** — *Personal and Family Health Hist* (705 concepts) is a
   later combined version that shares 576 concepts with *Personal Medical History*
   (481) + *Family Health History* (117), has 129 unique items, and is missing 22.
   It is **not** a strict duplicate; the dedup rule keeps the first-seen version of
   each shared concept and retains the 129 unique items. (All are physical-condition
   items and out of scope; this overlap does not affect the analyzed set.)

## Step 5 — Categorization

Each unique item is assigned one `category` by a two-stage **rule** (auditable; no
manual per-item labeling, no model):

0. **Free-entry detection (data validity, before any content rule).** Some `radio`
   items are REDCap wrappers around free-text entry: their options are "Enter
   age/number/response" plus sentinels, i.e. they carry **no scale points**, so
   order-coding them is meaningless. Any item with an option label beginning
   "Enter" → category **`free_entry`**, always out of scope (16 items, 14 of which
   would otherwise have been in scope: `ipaq_7`, `mhqukb_25/26/28/48/50/51/52/53`,
   `smoking_averagedailycigarette`, `smoking_currentdailycigarette`,
   `smoking_dailysmokestartingage`, `smoking_numberofyears`,
   `attemptquitsmoking_completelyquit`).
1. **Instrument-prefix map (primary).** The leading alphabetic token of `Item
   Concept` names the source instrument; it maps to a category:
   - psychological: `gad`,`worryanxiety`→anxiety · `phq`→depression ·
     `pcl`,`ies`,`ace`→trauma · `cidi`,`mhqukb`,`ukmh`,`ss`→psychiatric ·
     `bfi`→personality · `asrs`→adhd · `cpss`→stress · `brcs`,`lot`→wellbeing
   - habit/behavioral: `smoking`/`alcohol`/`tsu`/`audit`/… →substance · `ipaq`→activity
   - psychosocial: `eds`,`mos`,`nds`,`bmmrs`,`scns`,`hvs`,`nhs`,`sdoh`,`ucla`→psychosocial
   - functioning: `disability`→functioning
   - out of scope: physical-system prefixes (`cancer`,`circulatory`,…)→physical ·
     `mentalhealth`→mh_diagnosis · `familyhistory`→family_history · demographic /
     `insurance`/access / `overallhealth`→health_perception / covid / admin ·
     `ips`→neighborhood_built (except `ips_16`, kept by exact override)
2. **Keyword fallback.** Items whose prefix is unmapped are categorized by keyword
   match on `prompt`+`section` (e.g. "nervous/anxious/on edge"→anxiety;
   "discriminat/courtesy"→psychosocial; "smok/alcohol/tobacco"→substance).
3. Anything still unmatched → `uncategorized` (excluded; flagged for review).

Of the 968 unique items: 945 prefix-matched, 16 `free_entry`, 1 exact override
(`ips_16`), 2 keyword-matched, 4 `uncategorized` (all verified out of scope:
blood type, prenatal care, household type, hormone medication).

The full mapping is in `convert_allofus.py` (`PREFIX_CATEGORY`, `KEYWORD_CATEGORY`).

## Step 6 — Scope definition (the `embed` flag)

`embed = true` iff `category` is in the **in-scope set** — psychological + habit/
behavioral, operationalized as the 12 categories:
`{anxiety, depression, trauma, psychiatric, personality, adhd, stress, wellbeing,
substance, activity, psychosocial, functioning}`.
All other categories (physical, mh_diagnosis, family_history, demographic, access,
health_perception, covid, reproductive, admin, neighborhood_built, free_entry,
out, uncategorized) → `embed = false`.

## Step 6b — Review of categorization

The rule-assigned `category`/`embed` were reviewed at the instrument level (nearly
all items are prefix-matched, high confidence). The following corrections were
made and encoded back into the rules (so the procedure stays deterministic):

- `ss` (suicidality items) → **psychiatric** (was mislabeled wellbeing); stays in.
- `audit` (AUDIT-C) and an electronic-nicotine recency item → **substance**.
- `ips` (neighborhood **built-environment / walkability**: housing type, sidewalks,
  transit, bike facilities) → **out**, except `ips_16` ("the crime rate makes it
  unsafe to go on walks", a perceived-safety appraisal) → **kept** in (psychosocial),
  via an exact-item override.
- `chis` ("speak a language other than English at home") → **out** (demographic).
- Confirmed **kept in** (perceived environment / psychosocial context affects
  psychology): neighborhood disorder, food insecurity, neighborhood social cohesion,
  religiosity, and the `sdoh_*` items (a survey wrapper whose second token is the
  real instrument: perceived stress / discrimination / social support / loneliness).

## Step 6c — Expert-panel review (2026-06-09)

An expert review pass (psychometrics, statistical methods, code correctness; full
findings + remaining open analysis-design decisions in
[allofus_expert_review.md](allofus_expert_review.md)) led to the corrections now
encoded in the converter. **Mechanical fixes** (data validity, 356 → 342):

- **Free-entry exclusion** (Step 5, stage 0): 14 previously in-scope `radio` items
  were wrappers around free-text entry with no scale points → out.
- **Sentinel-aware level counts**: `n_levels_substantive` added; sentinel options
  inside `choices` would otherwise be miscoded as the scale maximum.
- **Section column fixed** (was the literal string "nan") and slider levels.

**Item-set rulings** (user decisions, 342 → 282):

1. **Twin instruments dropped (53 items).** GAD-7, PHQ-9, perceived stress (CPSS),
   everyday discrimination (EDS), MOS social support, UCLA loneliness each appeared
   in both COPE (COVID waves) and a home survey (EHH/SDOH). The COPE copy is dropped
   (near-duplicate wording → degenerate PCA basis + collinear responses); the
   home-survey copy is kept (full scale, no COVID framing). Encoded as `DUPLICATE_DROP`
   keyed `(survey, prefix)` → category `duplicate_cope`.
2. **Non-ordinal items handled (3 dropped, 2 recoded).** `mhqukb_11/14/15` are
   nominal/non-monotone (mood diurnality, appetite direction, weight change) → out
   (category `nominal`). `ace_5` and `mhqukb_27` kept as clean binaries by treating
   their off-axis option ("Parents not married"; "Not applicable") as a sentinel
   (`EXTRA_SENTINEL_CODES`; `mhqukb_27`'s already carries a PMI code).
3. **Factual-history items dropped (8).** The 7 `mhqukb_31_*` "Did <drug> help?"
   medication-response items (category `med_response`) and `nhs_covid_fhc17a`
   ("know someone who died of COVID-19", category `covid`).
4. **PROMIS + housing items added (4).** `overallhealth_emotionalproblem7days`,
   `overallhealth_generalmentalhealth` (→ psychiatric),
   `overallhealth_socialsatisfaction`, `livingsituation_stablehouseconcern`
   (→ psychosocial), via `ITEM_OVERRIDE` (their parent surveys are otherwise out).

The remaining open decisions are **analysis-design**, not item-set
(cross-survey missingness policy, COPE wave selection, branching-skip vs refusal
sentinels, numeric coding, valence column) — see the review doc §C.

## Results

**282 unique in-scope items** of 968 unique ordinal items (4 uncategorized).

| Survey | in-scope | ordinal |
|---|---:|---:|
| Emotional Health History | 79 | 92 |
| Social Determinants of Health | 69 | 78 |
| COPE | 57 | 158 |
| Behavioral Health | 42 | 50 |
| Lifestyle | 25 | 30 |
| Basics | 7 | 25 |
| Overall Health | 3 | 20 |
| Healthcare Access / Personal Medical History / Family Health History / Personal and Family Health Hist | 0 | (remainder) |
| **Total** | **282** | **968** |

By category (sum to 282): psychosocial 71, psychiatric 70, substance 66, trauma 23,
personality 15, depression 9, anxiety 8, functioning 6, adhd 6, wellbeing 5,
activity 3.

Domain groups: Psychological 136 · Psychosocial 71 · Habit/behavioral 69 · Functioning 6.

## Appendix — Complete shortlisting algorithm (precise, in execution order)

This is the authoritative, ordered statement of every rule. It mirrors
`convert_allofus.py` exactly (counts verified 2026-06-09). An item is **in scope
(`embed = true`)** iff it survives all filters below and its final `category` is in
the in-scope set. 968 unique items → **282 in-scope**.

**0. Sheet selection.** Keep a workbook sheet iff it has columns `Item Concept`,
`Field Type`, `Field Label` (→ 12 survey sheets; the 8 non-item sheets are dropped).

**1. Field-type filter.** Keep rows with `Field Type` ∈ {`radio`, `dropdown`,
`slider`} (1438 candidates). Drop rows with empty/`nan` `Item Concept`.

**2. Free-entry exclusion.** Parse `Choices` as `code, label | …` (split each
segment on the *first* comma; labels may contain commas). If any option label
begins with "Enter" → `category = free_entry` (out). This is checked **before**
content categorization. (16 items, of which 14 were otherwise in scope:
`ipaq_7`, `mhqukb_25`, `mhqukb_26`, `mhqukb_28`, `mhqukb_48`, `mhqukb_50`,
`mhqukb_51`, `mhqukb_52`, `mhqukb_53`, `smoking_averagedailycigarette`,
`smoking_currentdailycigarette`, `smoking_dailysmokestartingage`,
`smoking_numberofyears`, `attemptquitsmoking_completelyquit`.)

**3. Level counts.**
- `n_levels` = number of options (for `slider`, parsed from numeric endpoint
  anchors, e.g. 0–10 → 11).
- `n_levels_substantive` = `n_levels` minus **sentinel options**, where a sentinel
  is any option whose code starts with `pmi` (case-insensitive) **or** whose label
  matches `^(Don't know | Prefer not to answer | Skip)`, **plus** the per-item
  `EXTRA_SENTINEL_CODES` (currently only `ace_5 → {ehhwb_60}`, "Parents not married").

**4. Categorization** (first rule that fires wins):
1. **Exact-item override** (`ITEM_OVERRIDE`): `ips_16`→psychosocial;
   `overallhealth_emotionalproblem7days`→psychiatric;
   `overallhealth_generalmentalhealth`→psychiatric;
   `overallhealth_socialsatisfaction`→psychosocial;
   `livingsituation_stablehouseconcern`→psychosocial;
   `mhqukb_31_{amitriptyline,citalopram,dosulepin,fluoxetine,paroxetine,sertraline,other}`→med_response;
   `nhs_covid_fhc17a`→covid; `mhqukb_11`→nominal; `mhqukb_14`→nominal; `mhqukb_15`→nominal.
2. **Duplicate-drop** (`DUPLICATE_DROP`, keyed `(survey, instrument-prefix)`): in
   survey **COPE**, prefixes `gad`,`phq`,`cpss`,`eds`,`mos`,`ucla` → `duplicate_cope`
   (out). Keeps the home-survey copy (EHH `gad7_*`/`phq9_*`; SDOH `sdoh_cpss_*`/
   `sdoh_eds_*`/`sdoh_mos_ss_*`/`sdoh_ucla_ls8_*`).
3. **Instrument-prefix map** (`PREFIX_CATEGORY`, prefix = leading alphabetic token
   of `Item Concept`): anxiety `gad`,`worryanxiety`; depression `phq`; trauma
   `pcl`,`ies`,`ace`; psychiatric `cidi`,`mhqukb`,`ukmh`,`ss`; personality `bfi`;
   adhd `asrs`; stress `cpss`; wellbeing `brcs`,`lot`; substance `smoking`,
   `cigarsmoking`,`electronicsmoking`,`hookahsmoking`,`smokelesstobacco`,
   `attemptquitsmoking`,`alcohol`,`past`,`tsu`,`audit`; activity `ipaq`;
   psychosocial `eds`,`mos`,`nds`,`bmmrs`,`scns`,`sdoh`,`hvs`,`nhs`,`ucla`;
   functioning `disability`. **Out-of-scope prefixes:** `ips`→neighborhood_built,
   `chis`→demographic, physical-system prefixes→physical, `mentalhealth`→
   mh_diagnosis, `familyhistory`→family_history, demographic/`insurance`-family→
   demographic/access, `overallhealth`,`rand`→health_perception,
   `pregnancy`→reproductive, `yesnone`→out, `copect`,`cdc`,`msds`→covid,
   `section`,`pmi`→admin.
4. **Keyword fallback** on `prompt`+`section` text (only 2 items reach this):
   depress/anxiety/stress/trauma/discrimination/loneliness/religion→psychosocial/
   smoking/activity/personality/adhd/functioning keyword groups.
5. Otherwise → `uncategorized` (4 items, all verified out: blood type, prenatal
   care, household type, hormone medication).

**5. Dedup.** Drop duplicate `Item Concept` keeping the first codebook occurrence
(470 collapsed → 968 unique). Verified safe: no collapsed concept differs in
category/embed/field_type.

**6. In-scope set.** `embed = true` iff `category` ∈ {anxiety, depression, trauma,
psychiatric, personality, adhd, stress, wellbeing, substance, activity,
psychosocial, functioning}. All other categories (physical, mh_diagnosis,
family_history, demographic, access, health_perception, covid, reproductive,
admin, neighborhood_built, free_entry, med_response, nominal, duplicate_cope, out,
uncategorized) → `embed = false`.

**Recode note (generate stage).** `ace_5` and `mhqukb_27` stay in as clean
binaries: their off-axis third option ("Parents not married"; "Not applicable") is
treated as a sentinel (`ace_5` via `EXTRA_SENTINEL_CODES`; `mhqukb_27`'s option
already carries a PMI code), giving `n_levels_substantive = 2`. These sentinel
codes must be written into `scales.csv` at generate time.

## Reproducibility

```bash
pip install pandas openpyxl
python examples/allofus/convert_allofus.py \
  --codebook data/allofus/All_of_Us_Survey_Data_Codebooks.xlsx \
  --out      data/allofus/allofus_item_inventory.csv
```
Deterministic — same inputs reproduce the inventory and counts exactly. The
converter prints a warning if any `PREFIX_CATEGORY` entry matches zero items
(codebook-drift tripwire).

## Decisions deferred to the generate stage (not yet done)

The inventory fixes *which* items are analyzed. Turning the reviewed `embed = true`
rows into `prompts.csv` + `scales.csv` still requires:
1. **Numeric coding** — the source values the All of Us response export uses for
   each item (sets `scales.csv` `min/max`); to be confirmed in the Researcher
   Workbench. Until then only `n_levels`/`n_levels_substantive` are recorded.
   Observed responses must be validated against the declared `[min,max]` before
   normalization (hard error otherwise).
2. **Missing-value codes (sentinels)** — sentinel *options* are now identified
   per-item in `choices` (PMI codes / Don't-know / Prefer-not-to-answer); the
   exact numeric sentinel values in the response export must be confirmed
   in-workbench. Branching-logic skips (question never shown) must be
   distinguished from refusal sentinels — see the expert review doc.
3. **Reverse valence** — per-item, from instrument convention / scoring sheets.
4. **Response-matrix design** — cross-survey structural missingness policy, COPE
   wave selection, and the remaining item-set rulings in
   [allofus_expert_review.md](allofus_expert_review.md).
