## Supplementary material for "Auditable cross-instrument detection of unusual multivariate psychiatric response configurations using a semantically aligned covariance subspace": Code for tool: env_setup.md

# Environment setup (modern / bge-m3)

This documents the conda environment used to run the pipeline with real
`BAAI/bge-m3` embeddings. **Everything lives on the project disk**
(`/data_gpu5/semantic_framework_vipul`), not in `$HOME` — `/home` is small
(~31 GB free) and the env + model + caches are ~6.5 GB.

## Layout (all under `/data_gpu5/semantic_framework_vipul`)

| Path | What |
|------|------|
| `env/` | conda prefix env (Python 3.11, sci stack, torch CPU, sentence-transformers) |
| `models/bge-m3/` | local `BAAI/bge-m3` weights (offline; pass via `--model`) |
| `hf_cache/` | `HF_HOME` for the project (set by the env's activate hook) |
| `.conda_pkgs/` | conda package cache (`CONDA_PKGS_DIRS`) |
| `.pip_cache/` | pip cache (`PIP_CACHE_DIR`) |
| `survey-semantics/environment-bge-m3.yml` | human-readable spec |
| `survey-semantics/environment-bge-m3.lock.yml` | exact pinned export |

## Activate

```bash
source ~/miniconda3/bin/activate
conda activate /data_gpu5/semantic_framework_vipul/env
```

The env's `activate.d` hook automatically:
- prepends `$CONDA_PREFIX/lib` to `LD_LIBRARY_PATH` (see libstdc++ note below), and
- sets `HF_HOME` to the project `hf_cache/`.

## libstdc++ note (why the activate hook exists)

pip-installed manylinux wheels (scipy, torch) need `CXXABI_1.3.15`, but the system
`/lib64/libstdc++.so.6` only provides `1.3.13`. The conda env ships a newer
libstdc++ (`1.3.15`); the activate hook puts `$CONDA_PREFIX/lib` first on the
loader path so the wheels find it. Without it, `import scipy` fails with
`CXXABI_1.3.15 not found`.

## How it was built (reproduce from scratch)

```bash
source ~/miniconda3/bin/activate
PROJECT=/data_gpu5/semantic_framework_vipul
export CONDA_PKGS_DIRS=$PROJECT/.conda_pkgs
export PIP_CACHE_DIR=$PROJECT/.pip_cache

# 1. conda sci stack at a project-local prefix
conda create --yes --prefix "$PROJECT/env" -c conda-forge \
  python=3.11 "numpy>=1.24" "pandas>=2.0" "scipy>=1.10" "scikit-learn>=1.3" \
  "matplotlib>=3.7" numba pip pytest

# 2. activate + (one-time) write the activate.d/deactivate.d hooks
#    (see env/etc/conda/activate.d/zz_survey_semantics.sh)
conda activate "$PROJECT/env"

# 3. CPU torch + embedding stack
pip install torch --index-url https://download.pytorch.org/whl/cpu
pip install "sentence-transformers>=2.2" "transformers>=4.20" \
            "huggingface-hub>=0.20" "umap-learn>=0.5"

# 4. install the package (no-deps: keep the modern stack, ignore the legacy pins)
pip install -e "$PROJECT/survey-semantics" --no-deps

# 5. download the model to the project disk (one-time, needs network)
python - <<'PY'
from huggingface_hub import snapshot_download
snapshot_download(repo_id="BAAI/bge-m3",
                  local_dir="/data_gpu5/semantic_framework_vipul/models/bge-m3",
                  ignore_patterns=["onnx/**", "openvino/**", "imgs/**", "*.onnx"])
PY
```

## Verify

```bash
python -m pytest -q                      # in survey-semantics/  -> 11 passed
python -c "from survey_semantics.embedding import embed_texts_with_metadata as e; \
print(e(['hi'], method='sentence-transformers', \
        model_name='/data_gpu5/semantic_framework_vipul/models/bge-m3').vectors.shape)"
# -> (1, 1024)
```

## Running NHIS (once Part 1 scale/weights support lands)

```bash
survey-semantics analyze-file data/NHIS/2021/nhis2021.csv \
  --prompt-file  data/NHIS/2021/nhis2021_prompts.csv \
  --scale-file   data/NHIS/2021/nhis2021_scales.csv \
  --weights-file data/NHIS/2021/nhis2021_weights.csv \
  --id-col HHX \
  --embedding sentence-transformers \
  --model /data_gpu5/semantic_framework_vipul/models/bge-m3 \
  --d-selection variance --variance-threshold 0.80 \
  --outdir outputs/nhis/2021
```

Note: passing `--model <local path>` makes the embedding slug in output filenames
long (it encodes the path). Cosmetic only; rename outputs if cleaner names matter.

## Reclaiming space (optional)

`.conda_pkgs/` (~1.5 GB) and `.pip_cache/` (~220 MB) are caches; safe to clear:

```bash
conda clean -a -y && pip cache purge
```
