## Supplementary material for "Auditable cross-instrument detection of unusual multivariate psychiatric response configurations using a semantically aligned covariance subspace": Code for tool: HANDOFF.md

# Handoff — continue in a new chat

Last updated: 2026-06-09. This is the single catch-up doc for a new session.
The auto-loaded memory (`MEMORY.md` + memory files) carries the same facts; this
doc ties them together and states the **immediate next action**.

## What this project is

`survey-semantics` is the reusable implementation of **Periwal, "Auditable
cross-instrument detection of unusual multivariate response configurations using a
semantically aligned covariance subspace"** (manuscript PDF: `main_final.pdf`).
Method: embed questionnaire **item wording** → PCA of the prompt embeddings alone
(no responses) → project normalized responses → residualize age/sex → Ledoit-Wolf
Mahalanobis → flag multivariate outliers, auditably traced to items.

Three workstreams:
1. **The tool** (`survey-semantics/`) — done + extended (see below).
2. **NHIS 2021/2024** — done end-to-end (an *extension* adding survey weights).
3. **All of Us** — IN PROGRESS (current focus; at item-curation review).

Repo layout: code in `survey-semantics/` (its own git repo). Data lives OUTSIDE it
under `/data_gpu5/semantic_framework_vipul/data/{NHIS,allofus}/` (not committed;
restricted-data terms). A clean zip is at
`/data_gpu5/semantic_framework_vipul/survey-semantics.zip` (rebuild after changes).

## Environment

```bash
source ~/miniconda3/bin/activate
conda activate /data_gpu5/semantic_framework_vipul/env   # py3.11; bge-m3 at models/bge-m3
```
Embedding: **local sentence-transformers only, NO fallback** (TF-IDF removed per
user; missing model raises). Tests use a fake encoder (`tests/conftest.py`). Run
tests: `cd survey-semantics && python -m pytest -q` (expect ~29 passed).
`openpyxl` was pip-installed for the All of Us xlsx.

## Tool state (done, verified)

- **bge-m3 only**, offline, no fallback. CLI `--embedding sentence-transformers --model <path>`.
- **Scale file** (`--scale-file`, REQUIRED by `analyze-file`): `item,min,max,sentinels,reverse,ceiling,embed` — per-item ranges, per-item missing codes, reverse, ceiling allowlist, and `embed` item-selection allowlist (only `embed=true` items embedded+analyzed; column absent ⇒ all declared items; the `embed` subcommand also takes `--scale-file`). Implemented + tested 2026-06-09.
- **Weights file** (`--weights-file`, REQUIRED by `analyze-file`): WLS residualize + weighted Mahalanobis; matches the NHIS script to 1e-9. Equal weights (all 1s) = unweighted (same ranking).
- **Pan-mild** (`--pan-mild`): `At_Ceiling` + `Is_Pan_Mild_Emp<pct>`; ceiling allowlist via scale-file `ceiling` column.
- **LLM decoupled**: `embed` subcommand → `items.npz`; `analyze-file --embeddings-file` runs with NO model; bit-identical to inline. NHIS `run_nhis.sh` embeds once → both years share basis (W_2021==W_2024 exactly).
- Docs: `docs/pipeline_overview.md` (general), `README.md` (concise, worked example).

## NHIS state (done)

Converter `examples/nhis/convert_nhis.py` + `run_nhis.sh`. Per year: 38 common items.
Real bge-m3 run verified (2021: 29372 rows D=14; 2024: 32497 rows). Weight col `WTFA_A`.

## All of Us state (IN PROGRESS — current thread)

Goal: broad cross-survey, restricted to **psychological + habit/behavioral** items.
- Codebook: `data/allofus/All_of_Us_Survey_Data_Codebooks.xlsx` (REDCap dictionary,
  12 survey sheets + scoring sheets).
- Converter: `examples/allofus/convert_allofus.py` → inventory
  `data/allofus/allofus_item_inventory.csv` (968 unique items, cols
  `survey,item,form,section,field_type,prompt,n_levels,choices,category,embed`).
- Preprocessing methods (manuscript-grade): `docs/allofus_preprocessing.md`.
  Plan + counts: `docs/allofus_integration_plan.md`.
- Procedure: keep `radio/dropdown/slider` items → free-entry detection (radio
  wrappers around text entry → out) → dedup by Item Concept (keep-first,
  470 collapsed) → rule-based `category` (instrument-prefix map + keyword fallback)
  → `embed = category in {anxiety,depression,trauma,psychiatric,personality,adhd,
  stress,wellbeing,substance,activity,psychosocial,functioning}`.
- **ITEM SET LOCKED: 282 in-scope** (categorization §6b + expert panel + user rulings §6c).

### Review status: DONE (item set locked)

1. Categorization review (encoded as rules): KEEP nds/hvs/scns/bmmrs/nhs and all
   `sdoh`; `ss`→psychiatric; `audit`+e-cig→substance; `chis`→out; `ips`→out EXCEPT
   `ips_16`. (§6b)
2. **EXPERT PANEL REVIEW (2026-06-09)** — 3 agents (psychometrics, methods vs
   manuscript, code). Full doc: **`docs/allofus_expert_review.md`**.
   - Mechanical fixes (356→342): free-entry exclusion (14 items),
     `n_levels_substantive` (sentinels stripped), section-"nan" bug, slider levels,
     dead prefixes + drift-warning.
   - **User rulings (342→282)**: drop 53 COPE twin instruments (keep EHH/SDOH copy);
     drop 3 nominal mhqukb_11/14/15 + recode ace_5/mhqukb_27; drop 7 mhqukb_31_*
     med-response + nhs_covid_fhc17a; ADD 3 PROMIS + housing-worry item.
   - Per survey now: EHH 79, SDOH 69, COPE 57, BH 42, Lifestyle 25, Basics 7, OvHealth 3.

### >>> IMMEDIATE NEXT ACTION <<<

Work is split into **embedding** (done) + **downstream** (scales/responses/analysis).

**EMBEDDING MODULE DONE (2026-06-09) — 6 models.**
- `embed` tool column DONE: `ItemScale.embed`, `scales_use_embed`/`is_item_embedded`,
  pipeline restricts to `embed=true`, `embed` subcommand takes `--scale-file`. 32 tests.
- `convert_allofus.py --prompts-out` emits `prompts.csv` (item,prompt) of the 282
  embed=true items → `data/allofus/allofus_prompts.csv` (282 rows, unique keys).
- **Six embedding artifacts** in `data/allofus/` (same 282 items, identical order,
  no NaN; math/pipeline identical — raw prompt → encoder → unit vectors; each model
  gets its own PCA basis downstream, as the manuscript did for its 3 models):
  `allofus_items.npz` (bge-m3, primary, 1024d) ·
  `allofus_items_bge-large-en-v1.5.npz` (1024d, manuscript sensitivity) ·
  `allofus_items_all-mpnet-base-v2.npz` (768d, manuscript sensitivity) ·
  `allofus_items_gte-large-en-v1.5.npz` (1024d) ·
  `allofus_items_e5-large-v2.npz` (1024d) ·
  `allofus_items_qwen3-embedding-0.6b.npz` (1024d; norms 1±0.004, bf16 — harmless).
- Models live in `models/` (downloaded from HF). CLI gained `--trust-remote-code`
  (explicit opt-in; socket blocker stays active) for gte-large-en-v1.5.
- **gte-large-en-v1.5 quirks**: (1) its custom code lives in a SEPARATE HF repo
  `Alibaba-NLP/new-impl` (cached in `hf_cache/hub`, needed offline); (2) that code
  is incompatible with transformers 5.x (garbage position_ids → IndexError), so its
  embedding ran in a side venv `env_gte_tf4/` (torch-cpu + transformers 4.49 +
  sentence-transformers 3.4.1 + pandas). Re-embedding gte requires that venv.
- UMAP views of the bge-m3 item space: `examples/allofus/plot_item_umap.py`
  (`--color-by survey|instrument`, pre-labeled groupings only) →
  `data/allofus/allofus_items_umap_{survey,instrument}.png` + coords CSV.
- **Cross-model comparison**: `examples/allofus/plot_item_umap_models.py` →
  6-panel UMAP grid `data/allofus/allofus_items_umap_models.png` (+CSV). Same
  qualitative structure in all six (substance island / SDOH arm / internalizing
  mass). Quantitative: mean 10-NN Jaccard between item neighborhoods (in each
  model's own space) is 0.49–0.59 for ALL pairs (chance ≈ 0.02) — encoder choice
  is not driving the semantic structure.
- **PCA comparison (manuscript recipe per model)**: `examples/allofus/
  compare_pca_models.py` → `data/allofus/pca_compare/` (D-selection CSV,
  cumulative-variance plot, |corr| heatmaps vs bge-m3, Hungarian-matched
  component CSV, subspace-overlap matrix, bge-m3 top-loading items).
  Results: **D80 = 49–76** (bge-m3 57, bge-large 56, mpnet 49, gte 54, qwen3 56,
  e5 76) — far above the paper's 10–16, as the methods review predicted for 282
  diverse items (N/D in workbench needs care). Leading PCs match nearly 1:1
  across models (PC1 |corr| .93–.97, PC2 .85–.92, PC3 .70–.91), tail components
  rotate (mean matched |corr| over all of D80 ≈ .35–.41). **Retained-subspace
  overlap (mean cos² principal angles) = 0.67–0.79 for all pairs vs chance
  ≈ 0.19–0.23** — the spans largely agree, and Mahalanobis is rotation-invariant
  within the span, so this is the metric that matters downstream.
- **ROSS sparse PCA set up (2026-06-09).** User-supplied method "ROSS" =
  `SmoothSparsePCA` (JAX/optax, smooth-log penalty + Otsu ε + robust weighting),
  extracted to `ross_pca_pkg/ROSS_PCA_Codes/` (lib/sparse_PCA_v10.py). Deps in
  side venv **`env_ross/`** (jax 0.4.30 cpu, optax). Runner
  `examples/allofus/run_ross_spca.py`, analysis `examples/allofus/analyze_ross.py`.
  **Orientation B (user-chosen): items as FEATURES** — X = (m dims × 282 items),
  so sparsity lands on items (mirrors HLCA genes-as-features). NOTE this centers
  per-item-over-dims (HLCA analog), differing from the manuscript's bias-vector
  (per-dim-over-items) centering — a deliberate sparse variant. Sparsity is
  produced in TWO stages: smooth fit concentrates energy, then **Cosine-
  Preserving Pruning (CPP)** hard-zeros the tail at angle θ (keeps ≥cos²θ energy).
  **bge-m3 result (D=39, λs=λo=0.1):** dense-PCA participation ratio 87 → ROSS fit
  PR **22** (4× concentration); CPP θ=15° → 70% zeros / ~91 nnz / 93% energy / dir
  cos 0.97; θ=25° → 86% zeros / 43 nnz. Components map cleanly to instruments:
  PC3=neighborhood-disorder(nds), PC5=discrimination(eds/dms), PC7=smoking/vaping,
  PC8=alcohol(audit/alcohol), PC1=cross-instrument distress. Artifacts in
  `data/allofus/ross_out/bge-m3_ls0.1_lo0.1/` (loadings.npy, top_items*, cpp_sweep.json).
  OPEN: tune λ / pick operating θ; scale to other 5 models; decide if/how the
  sparse basis feeds the downstream Mahalanobis (orientation-B centering differs).
- **D-selection rule DECIDED (user, 2026-06-09): parallel analysis**
  (`--d-selection parallel`, Horn's, 50 perms, 95th pct). D_PA per model:
  bge-m3 39, bge-large 38, mpnet 32, gte 39, e5 37, qwen3 36 — tight across
  models (variance-80 was 49–76; eigengap is DEGENERATE for p<m full-rank
  evaluation: picks the rank cliff at 281; broken-stick 25–30, Kaiser 57–65).
  Variance-80 can be reported as sensitivity; Jaccard stability adjudicates
  in-workbench. **Components are NOT sparse**: median participation ratio
  ≈ 86–99 effective items of 282 per PC (PC1 densest ~155); top-10 items carry
  only ~22–24% of squared loading mass. Dense semantic contrasts, audited via
  top-loading items; varimax rotation of the retained basis would sharpen
  interpretability WITHOUT changing Mahalanobis (span-preserving) if wanted.

**NEXT (downstream module):** build `scales.csv` for the 282 — needs:
- `min`/`max` + sentinel numeric values: from the **scoring sheets** (numeric
  Source Value/Scale per option) for ~103+ scored items; the ~160 non-scored items
  need the workbench export coding confirmed (validate observed ∈ [min,max]).
- `reverse`: DEFERRED (user) — assign from valence later, or empirical auto-reverse.
- `ceiling`: pan-mild allowlist, not yet decided.
Then responses.csv + analysis run INSIDE the Researcher Workbench (reusing items.npz).

Remaining opens are **analysis-design only** (resolve in Researcher Workbench, see
review doc §C): cross-survey missingness policy (BLOCKER — pipeline default 50%+KNN
would impute whole surveys), COPE wave selection, branching-skip vs refusal
sentinels, numeric coding (validate observed ∈ [min,max]), reverse/valence column.

### Generate-phase details

1. **Add the `embed` column to the tool** (parallels `ceiling`): `ItemScale.embed`,
   item-selection restricts to `embed=true`, `embed` subcommand learns `--scale-file`.
2. **Generate step**: turn reviewed `embed=true` rows → `prompts.csv` + `scales.csv`.
   Blocked on: (a) numeric coding the AoU response export uses (→ scales min/max;
   confirm in Researcher Workbench), (b) sentinels from PMI codes (scoring sheets),
   (c) reverse valence per item.
3. **Responses + analysis** run INSIDE the Researcher Workbench (participant data
   can't be exported); converter prepares prompts/scales/items.npz here.

## Open decisions (cross-cutting)

- All of Us: `ips`/`chis` (above); AoU numeric coding, sentinels, reverse, weights (likely all-1s).
- Tool naming not finalized (candidates: from title — ACID / SACS / `cross-instrument-screen`).
- Nothing git-committed yet this session; rebuild `survey-semantics.zip` after changes.
