## Supplementary material for "Auditable cross-instrument detection of unusual multivariate psychiatric response configurations using a semantically aligned covariance subspace": Code for tool: nhis_integration_plan.md

# NHIS Integration Plan

Status: **proposed — awaiting review.** No code written yet.

## Goal

Run the NHIS 2021 and 2024 adult cohorts through the `survey-semantics`
pipeline, faithfully reusing the item definitions in
`data/NHIS/nhis_replication_2021_2024.py`, by:

1. Adding a per-item **scale file** as a first-class tool input (the "third file"
   alongside prompts and responses).
2. Writing a **converter** that turns each raw NHIS adult CSV into a tool-ready
   study folder (data + prompts + scales), derived from the script's dicts.

## Locked decisions

- **Common items only.** Restrict to items present in *both* years with identical
  wording. This is the only way to get a single shared semantic space across two
  independent cohorts (the space *is* the prompt-derived basis W).
- **Enhance the tool** to consume scales (not pre-bake in the converter).
- **Weighted analysis via a separate weights file.** Per-subject survey weights
  live in their own file, **row-aligned to the response file** (not a data column,
  not keyed by id), passed with `--weights-file`. When supplied, the pipeline runs
  weighted Mahalanobis + weighted (WLS) residualization to match the script. When
  omitted, the analysis is unweighted exactly as today.
- **Shared basis is automatic.** W is fit on item *embeddings* (prompts), not on
  responses ([pipeline.py:330](../src/survey_semantics/pipeline.py)). Same common
  items + same prompts + a prompt-only D-rule (`variance`) ⇒ bit-identical W and D
  in each per-year run. No basis save/load feature is needed. We add a sanity
  check that asserts the two years' W are equal.
- Each NHIS year is one subjects×items table ⇒ use **`analyze-file`** per year,
  not combined mode.

## Open decisions (need your call)

1. **Filenames.** Exact names of the raw CSVs to be placed in
   `data/NHIS/2021/` and `data/NHIS/2024/` (script default: `adult21.csv`,
   `adult24.csv`).

## Out of scope (script-only, not reproduced by the tool)

- "Ceiling audit" / "pan-mild" classification and cross-year demographic
  comparison. The tool emits outliers, drivers, stability, and case studies; the
  pan-mild logic stays in the replication script if needed.

---

## Part 1 — Scale file (new tool input)

### File format

Combined file or per-instrument dir, mirroring prompt-file conventions. Bare
`item` keys apply to any instrument; an optional `table` column scopes a row.

```csv
item,min,max,sentinels,reverse
SAD_A,1,5,7;8;9,true
SLPREST_A,1,4,7;8;9,true
LASTDR_A,1,8,0;97;98;99,false
MHTHRPY_A,1,2,7;8;9,false
```

- `sentinels`: `;`-separated codes mapped to missing, **per item**.
- `reverse`: truthy → item negated (authoritative; unifies with reverse-config).
- This is a direct serialization of `ALL_ITEM_SCALES`.

### Code changes

**New module `src/survey_semantics/scales.py`** (parallels `prompts.py`):

```python
def load_scale_sources(scale_file: Optional[Path],
                       scale_dir: Optional[Path]) -> Dict[str, ItemScale]
# ItemScale = {"min": float, "max": float,
#              "sentinels": set[float], "reverse": bool}
```
Same namespacing as prompts: bare `item` → also keyed as `table__item`; resolution
order qualified → bare; case/punctuation-normalized fallback.

**`pipeline.AnalysisConfig`** — add:
```python
item_scales: Optional[Mapping[str, ItemScale]] = None
```

**`pipeline.analyze_survey_table`** — when `config.item_scales` is set:
- **Item set:** analyzed items = declared items present in `table.data`
  (skip `infer_item_columns`). This avoids NHIS `7/8/9` codes inflating unique
  counts and wrongly dropping items like `LASTDR_A`.
- **Sentinels:** clean per item using each item's `sentinels` (replaces the single
  global `config.sentinels`). Implemented by resolving a per-item sentinel set and
  passing it to a per-column-aware coercion (extend `coerce_response_frame` to
  accept a `Mapping[col, set]`, or clean column-by-column in the pipeline).
- **Ranges:** in `_observed_item_ranges`, prefer declared `min/max` when present;
  fall back to observed otherwise.
- **Reverse:** union declared `reverse` items into `config.reverse_items`.

**`cli.py`** — add `--scale-file` / `--scale-dir` to `_add_common_args`; load via
`load_scale_sources`; set `config.item_scales`. (Brings reverse-scoring to
`analyze-file`, which currently lacks it.)

### Backward compatibility

All new behavior is gated on `item_scales` being provided. With no scale input the
pipeline behaves exactly as today (observed ranges, global sentinels, inference).

---

## Part 1b — Weights file (new tool input)

### File format

A standalone file with **one weight per subject, in the same row order as the
response file**. One numeric column, optional header `weight`:

```csv
weight
12345.6
0.0
8901.2
```

- Length **must equal the number of rows in the response CSV** (the raw table,
  before completeness filtering). Mismatch → hard error with both counts.
- Alignment is **positional**, so the file is fragile if rows are reordered by
  hand. Mitigation: the converter emits it together with the data file from the
  same row order, guaranteeing alignment. (Not keyed by id, per the chosen design.)
- Non-finite or `<= 0` weights are replaced with the median of the valid weights
  (same guard as the script).

### Code changes

**`pipeline.AnalysisConfig`** — add:
```python
sample_weights: Optional[np.ndarray] = None   # length == len(table.data)
```

**`pipeline.analyze_survey_table`**:
- Subset weights by the same `keep_rows` completeness mask used for responses, so
  weights stay aligned to the scored subjects.
- Pass weights into residualization and Mahalanobis.

**`pipeline.residualize`** — add optional `weights`: when present, solve weighted
least squares (`sqrt(w)` row-scaling of `X` and `y`) per component, matching the
script's WLS residualization.

**`pipeline.mahalanobis_distances`** — add optional `weights`: when present, use
the weighted mean, weighted scatter `S` scaled by `n_eff/(n_eff-1)` with
`n_eff = 1/sum(w^2)`, and Ledoit-Wolf shrinkage toward `(trace(S)/D)*I` using the
shrinkage coefficient from `LedoitWolf().fit(...)` — i.e. the exact weighted
covariance the script builds. Stability frame uses the same weighted distances.

**`cli.py`** — add `--weights-file` to `_add_common_args`; load the column, length-
check against `len(table.data)`, set `config.sample_weights`.

### Backward compatibility

Gated on `sample_weights`. Absent ⇒ unweighted `LinearRegression` + unweighted
`LedoitWolf`, identical to current behavior.

---

## Part 2 — Converter `data/NHIS/convert_nhis.py`

Single source of truth = the dicts already in `nhis_replication_2021_2024.py`
(imported, not duplicated).

### Inputs
```
--data2021 <path>   default data/NHIS/2021/adult21.csv
--data2024 <path>   default data/NHIS/2024/adult24.csv
--outdir   <path>   default data/NHIS
```

### Steps
1. Load both raw CSVs.
2. `common = find_common_items(df21, df24)` (reuse script's function).
3. Assert prompt text is identical across years for each common item (trivial,
   since prompts come from one `ALL_QUESTION_TEXTS`); fail loudly otherwise.
4. For each year write into `data/NHIS/<year>/`:
   - `nhis<year>.csv`: `HHX` (id) + `age` (from `AGEP_A`) + `sex` (from `SEX_A`)
     + the common item columns, **raw** (sentinels are cleaned by the tool from
     the scales file, keeping the data file auditable).
   - `nhis<year>_prompts.csv`: `item,prompt` from `ALL_QUESTION_TEXTS[common]`.
   - `nhis<year>_scales.csv`: `item,min,max,sentinels,reverse` from
     `ALL_ITEM_SCALES[common]`.
   - `nhis<year>_weights.csv`: single `weight` column from `WTFA_A`, written in the
     **same row order** as `nhis<year>.csv` (guarantees positional alignment).
5. Print the common-item count and the per-year row counts; assert the weights file
   length equals the data row count.

### ID / covariates mapping
- ID: NHIS has no `subjectkey`; pass `--id-col HHX` at run time (one sample adult
  per household ⇒ unique per row).
- Covariates: rename `AGEP_A → age`, `SEX_A → sex` so the tool's
  `default_covariates` picks them up automatically.

---

## Part 3 — Running it (per year)

```bash
survey-semantics analyze-file data/NHIS/2021/nhis2021.csv \
  --prompt-file  data/NHIS/2021/nhis2021_prompts.csv \
  --scale-file   data/NHIS/2021/nhis2021_scales.csv \
  --weights-file data/NHIS/2021/nhis2021_weights.csv \
  --id-col       HHX \
  --embedding sentence-transformers --model /path/to/bge-m3 \
  --d-selection variance --variance-threshold 0.80 \
  --skip-umap \
  --outdir outputs/nhis/2021
```
(and the same for 2024). Use the
single-file `--prompt-file`/`--scale-file` forms (not `--prompt-dir`/`--scale-dir`):
the year folder also holds the raw `adultNN.csv`, which a directory loader would
wrongly try to parse as a prompt/scale file.

## Part 4 — Verification

- Unit tests: scale loader parsing; per-item sentinel cleaning; declared-range
  normalization; reverse from scales; weights length-check; weighted vs unweighted
  Mahalanobis on a tiny fixture.
- Replication check: assert `W_2021 == W_2024` (shared-basis proof).
- Spot-check a few subjects' weighted Mahalanobis distances against the script's
  `process_year` output on the same rows (the strongest end-to-end fidelity test).
- Spot-check a few items' normalized values against the script's normalization.

## Risks / notes

- **Weights alignment is positional** — the weights file must match the response
  file row order and length. The converter emits both together to guarantee this;
  the tool hard-errors on a length mismatch but cannot detect a silent reordering.
- NHIS `AGEP_A`/`SEX_A` own sentinels (e.g. `97/98/99`, `7/8/9`): clean in the
  converter or document that the tool's covariate coercion fills NaN with median.
- If a "common" item was reworded between years but kept its variable name, the
  identical-wording assertion (step 3) will catch it.
