## Supplementary material for "Auditable cross-instrument detection of unusual multivariate psychiatric response configurations using a semantically aligned covariance subspace": Code for tool: notebook_refactor.md

# Notebook Refactor Notes

Source notebook: `DO NOT CHANGE Computational psychometrics6-Copy1.ipynb`.

The notebook combines two validation scripts and repeated plotting/case-study cells. The reusable package extracts the stable computational pattern:

1. Item wording is treated as the semantic dictionary.
2. Item text embeddings are centered to remove global language-model/text-feature bias.
3. PCA on item embeddings yields an instrument-level semantic basis.
4. Normalized item responses are projected through item coordinates.
5. Optional covariates are regressed out of each semantic component.
6. Ledoit-Wolf covariance regularization produces Mahalanobis deviation scores.
7. Theoretical chi-square and empirical percentile thresholds flag outliers.
8. Stability, driver, and standalone case-study text reports replace notebook-only plots/printouts.

Notebook-specific HRS/Xinxiang file paths and publication plotting code were intentionally not hardcoded. Manually defined notebook item dictionaries are now supplied as external prompt files, with NDA-style second header rows and column names available as fallbacks. This keeps the analysis usable on survey data outside the original mental-health examples.
