## Supplementary material for "Auditable cross-instrument detection of unusual multivariate psychiatric response configurations using a semantically aligned covariance subspace": Code for tool: pipeline_overview.md

# survey-semantics — General Pipeline Overview

This document describes the **general** survey-semantics pipeline: what it does,
why, the role of each module, the file formats it consumes, and how to run it.
It is deliberately domain-agnostic — NHIS is just one dataset fed through it (see
[nhis_integration_plan.md](nhis_integration_plan.md) for that specific wiring).

> **Provenance.** This pipeline implements the method in Periwal, *"Auditable
> cross-instrument detection of unusual multivariate response configurations using
> a semantically aligned covariance subspace"* (validated on the **HRS** and
> **Xinxiang** cohorts). It matches the paper: normalize to **[−1,1]**,
> reverse-score, KNN k=5 impute, prompt-only PCA to 80% variance, OLS
> residualization of age/sex, Ledoit-Wolf Mahalanobis, empirical 95th-percentile
> outliers, Jaccard stability, and the **pan-mild ceiling audit** (`--pan-mild`,
> opt-in). One **extension** beyond the paper: **survey weights** (weighted
> residualization + weighted Mahalanobis) for NHIS-style weighted designs, gated
> and off by default.

---

## 1. What the tool does, in one paragraph

Given a table of **subjects × survey items** and the **wording of each item**, the
pipeline builds a latent "semantic space" from the *meaning of the questions*
(not from the response numbers), projects each subject's answers into that space,
removes demographic signal, and then flags subjects who sit unusually far from the
center as **outliers** — reporting which dimensions and items drive each one. The
defining idea: the coordinate system comes from **embedding the item prompts**, so
items that *ask similar things* group together automatically, and the space is
**reproducible across studies that share item wording** (the same prompts always
yield the same basis).

---

## 2. The conceptual pipeline (stage by stage)

```
   prompts.csv      data.csv (subjects × items)      scales.csv*    weights.csv*
       │                    │                            │              │
       ▼                    ▼                            ▼              ▼
  ┌─────────┐        ┌──────────────┐            (per-item ranges,  (per-subject
  │ item    │        │ 2. select    │◄───────────  sentinels,        survey weights,
  │ wording │        │    items     │             reverse)           optional)
  └────┬────┘        └──────┬───────┘                                     │
       │                    ▼                                             │
       │            ┌──────────────┐  3. coerce → clean missing/sentinels │
       │            │ 4. drop      │     (per-item codes when scales given)│
       │            │ incomplete   │                                       │
       │            │ subjects     │                                       │
       │            └──────┬───────┘                                       │
       │                   ▼  5. impute blanks (KNN / median)              │
       │            ┌──────────────┐                                       │
       │            │ 6. normalize │  each item → [-1,1] by its range;     │
       │            │  + reverse   │  reverse-score flagged items          │
       │            └──────┬───────┘                                       │
       ▼                   │                                               │
  ┌─────────┐              │                                               │
  │7. embed │  item text → vectors (local bge-m3 sentence encoder)         │
  │ prompts │              │                                               │
  └────┬────┘              │                                               │
       ▼                   │                                               │
  ┌─────────┐              │                                               │
  │8. PCA → │  basis W = principal axes of *prompt* embeddings             │
  │ basis W │  (the semantic space; independent of responses)             │
  └────┬────┘              │                                               │
       │   9. choose D (variance / eigengap / parallel / stability)        │
       ▼                   ▼                                               │
       └────────►  10. project: scores = response_norm · W[:, :D]          │
                          │                                                │
                          ▼  11. residualize out covariates (WLS if weights)◄┘
                   ┌──────────────┐
                   │ 12. Mahalan- │  weighted covariance if weights given
                   │ obis distance│  → how unusual each subject is
                   └──────┬───────┘
                          ▼
              13. outputs: outlier scores, drivers, loadings, stability, case studies
```
`*` The `analyze-file` CLI **requires** scales and weights (see §4); pan-mild is
optional. The library `analyze_survey_table` can still run without them (item
inference, global sentinels, observed ranges, unweighted statistics) for tests
and flexibility, but the CLI does not expose that path.

### Why embed the *prompts*?
A classic approach treats each item as an independent numeric column and runs
factor analysis on the responses. That makes the latent space depend on *this
sample's* answers, so it differs between cohorts and needs manual factor labeling.
Here the space is the **PCA of the question wording's embeddings** — so:
- Items that mean similar things land near each other with no hand-labeling.
- Two studies with the *same item wording* get a **bit-identical basis W**
  (W is fit on prompts, not responses), which is what makes cross-cohort
  comparison valid. Responses only ever get *projected into* W; they never
  define it.

---

## 3. Modules — what each does and why

| Module | Lines | Responsibility | Why it exists |
|---|---|---|---|
| [`cli.py`](../src/survey_semantics/cli.py) | ~390 | Argument parsing; loads prompts/scales; builds `AnalysisConfig`; dispatches the three modular stages (`embed` / `pca` / `analyze-file`) plus `analyze-package` / combined; writes outputs. | Single entry point; keeps I/O and orchestration out of the math. |
| [`io.py`](../src/survey_semantics/io.py) | 290 | Read survey tables (generic + NDA dictionary-row format); coerce responses to numeric; infer item columns; clean sentinels; build covariate matrix. | All the messy "turn a CSV into clean numeric arrays" logic in one place. |
| [`prompts.py`](../src/survey_semantics/prompts.py) | 286 | Load item **wording** from CSV/JSON/Python-dict/dir; namespaced key resolution (`table__item` → bare → normalized). | Prompt text is the raw material for the semantic space; must match items robustly across naming styles. |
| [`scales.py`](../src/survey_semantics/scales.py) | 412 | Load per-item **scale specs** (`min,max,sentinels,reverse,ceiling,embed`); same namespacing as prompts. *(Part 1, new.)* | Lets the tool know each item's valid range, its own missing-value codes, reverse coding, ceiling-audit eligibility, and whether it's in the analyzed set (`embed`) — things it otherwise has to guess from data. |
| [`embedding.py`](../src/survey_semantics/embedding.py) | ~270 | **Stage 1 (the LLM step)**, fully decoupled: embed item text via a local `sentence-transformers` model (e.g. bge-m3); save/load a reusable `ItemEmbeddings` artifact (`embed` subcommand); enforce the offline policy. No fallback — a missing model raises. | Isolates the only model dependency so embedding can run once and the PCA/downstream analysis can run with no model. |
| [`basis.py`](../src/survey_semantics/basis.py) | ~200 | **Stage 2 (PCA)**: `build_semantic_basis` centers the item embeddings, runs PCA, and computes the embedding-only D diagnostics (variance/eigengap/parallel); save/load a reusable `SemanticBasis` artifact (`pca` subcommand). | The semantic space depends only on the prompts, so it is a cacheable, response-independent artifact — reuse one basis across waves to guarantee a shared space. |
| [`pipeline.py`](../src/survey_semantics/pipeline.py) | ~1120 | **Stage 3 + orchestration**: `analyze_survey_table` (build/accept the basis, then project, residualize, Mahalanobis, stability, drivers, case studies, UMAP). | The heart — everything else feeds it clean inputs and serializes its outputs. |
| [`combined.py`](../src/survey_semantics/combined.py) | 663 | Build a single transdiagnostic matrix across *many* questionnaire files (auto-reverse detection, item merging). | For the "many small instruments → one space" mode; not used for the per-year NHIS runs. |
| [`plotting.py`](../src/survey_semantics/plotting.py) | 431 | Render PDF plots from the output CSVs. | Optional visualization, decoupled from analysis. |

### Key `pipeline.py` functions
- `analyze_survey_table(table, config, item_columns=None, item_embeddings=None, basis=None)`
  — the orchestrator. A precomputed `basis` skips embedding **and** PCA; precomputed
  `item_embeddings` skips only the model; neither embeds + builds the basis inline.
  The basis itself is built by `basis.build_semantic_basis` (centering + PCA + parallel).
- `normalize_responses` — scale each item to `[-1,1]` by its range (matching the manuscript; reverse-scored items then negated).
- `select_component_count` / `eigengap_dimension` / `parallel_analysis` /
  `stability_dimension` / `choose_dimension` — the four **D-selection** rules.
- `residualize(matrix, covariates, weights=None)` — regress covariates out of the scores (OLS, or WLS when weights given).
- `mahalanobis_distances(matrix, weights=None)` — distance from center in the residual semantic space → the outlier score (unweighted Ledoit-Wolf, or weighted scatter when weights given).

---

## 4. Data contract (input file formats)

### Response table — `data.csv` (required)
Subjects in rows, items in columns, plus an id column and any covariates.
```csv
HHX,age,sex,SAD_A,NERVOUS_A,LASTDR_A,...
10001,42,2,1,2,7,...
```
- Covariates named `age`/`sex` (and similar) are auto-detected by `default_covariates`.
- Pass the id column with `--id-col` if it isn't a standard name.

### Prompt file — `*_prompts.csv` (required for meaningful embeddings)
```csv
item,prompt
SAD_A,"How often do you feel sad?"
```
One row per item; supports `--prompt-file` (one file) or `--prompt-dir` (per-instrument).

### Scale file — `*_scales.csv` (**required** by the `analyze-file` CLI) — `--scale-file` / `--scale-dir`
```csv
item,min,max,sentinels,reverse,ceiling,embed
SAD_A,1,5,7;8;9,true,true,true
LASTDR_A,1,8,0;97;98;99,false,false,false
```
- `min,max` — valid range, used for normalization instead of observed range.
- `sentinels` — `;`-separated **per-item** missing codes (mapped to NaN). This is
  why `7` can be "refused" for `SAD_A` but a *valid* answer for `LASTDR_A`.
- `reverse` — truthy → item is negated after normalization.
- `ceiling` *(optional)* — whether the item participates in the `--pan-mild`
  item-level ceiling check. When **any** item declares it, it acts as an explicit
  allowlist (only `ceiling=true` items are checked) — so an item like `LASTDR_A`,
  where "maxed out" is not a symptom ceiling, can be excluded. When the column is
  **absent**, the audit falls back to all polytomous items (≥3 levels).
- `embed` *(optional)* — **item-selection allowlist.** When **any** item declares
  it, only `embed=true` items are embedded *and* analyzed; `embed=false` (and
  blank) items stay in the file as documentation but are excluded from the basis,
  the distances, and the item set. When the column is **absent**, every declared
  item is analyzed (the pre-`embed` behavior). Use it to curate a large
  cross-instrument item pool auditably. The same flag drives the `embed`
  subcommand: `survey-semantics embed --scale-file scales.csv …`
  embeds exactly the `embed=true` items, so `items.npz` matches the analyzed set.
- **When supplied, the declared (and `embed=true`) items also define the analyzed
  set** (inference is skipped), so valid items aren't dropped for having "too many"
  unique codes.

### Weights file — `*_weights.csv` (**required** by the `analyze-file` CLI) — `--weights-file`
```csv
weight
5423.324
3832.196
```
One weight per subject, **row-aligned to `data.csv`** (positional, not keyed; the
length must equal the raw response-row count or the run hard-errors). Header is
optional. When present, residualization uses WLS (sqrt-weight row scaling) and
Mahalanobis uses a weighted mean + weighted scatter with Ledoit-Wolf shrinkage —
matching the NHIS replication script exactly (verified to 1e-9). Non-finite or
`<= 0` weights are replaced with the median of the valid weights.

> **Required vs optional.** The `analyze-file` CLI **requires** a scale file (per-item
> `reverse` flag — part of the method) **and** a weights file. For an unweighted
> analysis (as in the paper), pass a column of **equal weights** (e.g. all `1`s):
> equal weights make WLS = OLS, and the weighted Mahalanobis becomes the unweighted
> distances times a constant `sqrt((n-1)/n)` — so the **outlier ranking and empirical
> outlier set are identical** (only absolute distances differ, negligibly). Pan-mild stays
> optional. The library `analyze_survey_table` still accepts no `item_scales` /
> no `sample_weights` (for flexibility and tests); only the CLI enforces them.

---

## 5. How to run

### Environment
See [env_setup.md](env_setup.md). In short:
```bash
source ~/miniconda3/bin/activate
conda activate /data_gpu5/semantic_framework_vipul/env
```

### Generic single-file run
```bash
python -m survey_semantics.cli analyze-file path/to/data.csv \
  --prompt-file  path/to/data_prompts.csv \
  --scale-file   path/to/data_scales.csv \    # required
  --weights-file path/to/data_weights.csv \   # required (all-1s for unweighted)
  --id-col       <ID_COLUMN> \
  --embedding    sentence-transformers --model /path/to/bge-m3 \
  --d-selection  variance --variance-threshold 0.80 \
  --outdir outputs/<study>
```

> **Embedding backend.** The only supported backend is a local
> `sentence-transformers` model (e.g. bge-m3); there is no TF-IDF/auto fallback.
> The model must already be on local disk (the pipeline blocks network egress).
> Tests use a deterministic fake encoder so they run without the model.

### The three modular stages (embed → pca → score)

Embedding (stage 7) depends only on the prompts, and the PCA basis only on the
embeddings — neither touches responses. So both are separable, cacheable stages.
Run them once and reuse the artifacts:

```bash
# Stage 1 — LLM step → reusable embeddings (prompts only):
python -m survey_semantics.cli embed --prompt-file prompts.csv --model /path/to/bge-m3 --out items.npz
# Stage 2 — PCA → reusable semantic basis (embeddings only, no responses):
python -m survey_semantics.cli pca --embeddings-file items.npz --out basis.npz
# Stage 3 — score with NO model (project + Mahalanobis + …):
python -m survey_semantics.cli analyze-file responses.csv \
  --prompt-file prompts.csv --scale-file scales.csv --weights-file weights.csv \
  --basis-file basis.npz --outdir outputs/run
```

The result is **bit-identical** to embedding inline (verified on NHIS 2021: max
abs distance diff `0.0`). You can also stop after stage 1 and pass
`--embeddings-file items.npz` to `analyze-file` (it builds the basis inline but
still needs no model). Reusing one `basis.npz` across waves/cohorts guarantees a
shared basis `W`. Both artifacts validate item alignment and **raise** on any
missing/extra item — `ItemEmbeddings.matrix_for` and `SemanticBasis.coordinates_for`
never silently substitute or drop.

Any local sentence-transformers model works the same way (the manuscript used
bge-m3 as primary and bge-large-en-v1.5 / all-mpnet-base-v2 for sensitivity;
each model gets its own `items.npz` and its own PCA basis). For models whose
repo ships custom architecture code (e.g. `gte-large-en-v1.5`), pass the
explicit `--trust-remote-code` opt-in — the code runs from the already-local
files and the outbound-socket blocker stays active either way.

### Worked example — NHIS 2021
```bash
python -m survey_semantics.cli analyze-file data/NHIS/2021/nhis2021.csv \
  --prompt-file  data/NHIS/2021/nhis2021_prompts.csv \
  --scale-file   data/NHIS/2021/nhis2021_scales.csv \
  --weights-file data/NHIS/2021/nhis2021_weights.csv \
  --id-col HHX --embedding sentence-transformers --model /path/to/bge-m3 \
  --d-selection variance --variance-threshold 0.80 \
  --skip-umap --outdir outputs/nhis/2021
# → Analyzed nhis2021: ... rows, 38 items, D=...
```
With the scale file, all 38 declared items are kept; without it, item inference
drops one (a valid item whose missing codes inflated its unique-value count).

> **Use `--scale-file` not `--scale-dir`** when the data folder also holds the raw
> source CSV — a directory loader would try to parse that raw file as a scale file.

---

## 6. Outputs

Written to `--outdir`, prefixed by `<table>__<embedding-slug>`:

| File | Contents |
|---|---|
| `*_scores.csv` | Per-subject Mahalanobis distance + semantic PC scores + id/covariates + outlier flags. The outlier ranking. With `--pan-mild`: also `At_Ceiling` and `Is_Pan_Mild_Emp<pct>` (outlier and nowhere at the Likert ceiling). |
| `*_drivers.csv` | Which dimensions/items drive each top outlier. |
| `*_prompt_loadings.csv` | How each item prompt loads on each semantic dimension (interpret the axes). |
| `*_item_weights.csv` | Item contribution weights. |
| `*_stability.csv` | Jaccard stability of the outlier set vs. dimensionality. |
| `*_dimension_selection.csv`, `*_dimension_methods.csv` | What D each rule picked and why. |
| `case_studies/` | Narrative per-subject reports for the top outliers. |
| `summary.csv` | One-row run summary (n_rows, n_items, D, embedding, ...). |

---

## 7. Extension points (keeping it general)

The design rule is: **new survey-specific needs become generic tool inputs, not
hard-coded branches.** Examples:
- Per-item measurement metadata → the **scale file** (`scales.py`), not NHIS code.
- Survey weights → a generic **weights file**, row-aligned (Part 1b).
- New dataset → write a *converter* that emits `data/prompts/scales/weights` in the
  formats above (e.g. `data/NHIS/convert_nhis.py`); the pipeline is untouched.

This keeps one pipeline that any subjects×items survey can run through, with
dataset quirks isolated in small converters at the edge.

---

## 8. Status

- **Done:** core pipeline; prompt loading; embeddings (local bge-m3, no fallback); scale file
  end-to-end (`scales.py` + per-item sentinels + declared ranges + reverse), verified
  on NHIS 2021; weights file end-to-end (`--weights-file`, WLS residualization,
  weighted Mahalanobis), numerically matched to the NHIS script to 1e-9.
- **Next:** run both NHIS years with bge-m3 and the shared-basis check
  (`W_2021 == W_2024`); see [nhis_integration_plan.md](nhis_integration_plan.md) §Part 4.
