## Supplementary material for "Auditable cross-instrument detection of unusual multivariate psychiatric response configurations using a semantically aligned covariance subspace": Code for tool: environment-bge-m3.lock.yml

```
name: /data_gpu5/semantic_framework_vipul/env
channels:
  - conda-forge
  - defaults
dependencies:
  - _openmp_mutex=4.5=20_gnu
  - alsa-lib=1.2.16=hb03c661_0
  - brotli=1.2.0=hed03a55_1
  - brotli-bin=1.2.0=hb03c661_1
  - bzip2=1.0.8=hda65f42_9
  - ca-certificates=2026.5.20=hbd8a1cb_0
  - cairo=1.18.4=he90730b_1
  - colorama=0.4.6=pyhd8ed1ab_1
  - contourpy=1.3.3=py311h724c32c_4
  - cycler=0.12.1=pyhcf101f3_2
  - cyrus-sasl=2.1.28=hac629b4_1
  - dbus=1.16.2=h24cb091_1
  - double-conversion=3.4.0=hecca717_0
  - exceptiongroup=1.3.1=pyhd8ed1ab_0
  - font-ttf-dejavu-sans-mono=2.37=hab24e00_0
  - font-ttf-inconsolata=3.000=h77eed37_0
  - font-ttf-source-code-pro=2.038=h77eed37_0
  - font-ttf-ubuntu=0.83=h77eed37_3
  - fontconfig=2.18.1=h27c8c51_0
  - fonts-conda-ecosystem=1=0
  - fonts-conda-forge=1=hc364b38_1
  - fonttools=4.63.0=py311h3778330_0
  - freetype=2.14.3=ha770c72_0
  - graphite2=1.3.15=hecca717_0
  - harfbuzz=14.2.1=h6083320_0
  - icu=78.3=h33c6efd_0
  - iniconfig=2.3.0=pyhd8ed1ab_0
  - joblib=1.5.3=pyhd8ed1ab_0
  - keyutils=1.6.3=hb9d3cd8_0
  - kiwisolver=1.5.0=py311h724c32c_0
  - krb5=1.22.2=ha1258a1_0
  - lcms2=2.19.1=h0c24ade_1
  - ld_impl_linux-64=2.45.1=default_hbd61a6d_102
  - lerc=4.1.0=hdb68285_0
  - libblas=3.11.0=8_h4a7cf45_openblas
  - libbrotlicommon=1.2.0=hb03c661_1
  - libbrotlidec=1.2.0=hb03c661_1
  - libbrotlienc=1.2.0=hb03c661_1
  - libcblas=3.11.0=8_h0358290_openblas
  - libclang13=22.1.7=default_h746c552_1
  - libcups=2.3.3=h7a8fb5f_6
  - libdeflate=1.25=h17f619e_0
  - libdrm=2.4.127=hb03c661_0
  - libedit=3.1.20250104=pl5321h7949ede_0
  - libegl=1.7.0=ha4b6fd6_3
  - libegl-devel=1.7.0=ha4b6fd6_3
  - libexpat=2.8.1=hecca717_0
  - libffi=3.5.2=h3435931_0
  - libfreetype=2.14.3=ha770c72_0
  - libfreetype6=2.14.3=h73754d4_0
  - libgcc=15.2.0=he0feb66_19
  - libgcc-ng=15.2.0=h69a702a_19
  - libgfortran=15.2.0=h69a702a_19
  - libgfortran5=15.2.0=h68bc16d_19
  - libgl=1.7.0=ha4b6fd6_3
  - libgl-devel=1.7.0=ha4b6fd6_3
  - libglib=2.88.1=h0d30a3d_2
  - libglvnd=1.7.0=ha4b6fd6_3
  - libglx=1.7.0=ha4b6fd6_3
  - libglx-devel=1.7.0=ha4b6fd6_3
  - libgomp=15.2.0=he0feb66_19
  - libiconv=1.18=h3b78370_2
  - libjpeg-turbo=3.1.4.1=hb03c661_0
  - liblapack=3.11.0=8_h47877c9_openblas
  - libllvm22=22.1.7=hf7376ad_0
  - liblzma=5.8.3=hb03c661_0
  - libnsl=2.0.1=hb9d3cd8_1
  - libntlm=1.8=hb9d3cd8_0
  - libopenblas=0.3.33=pthreads_h94d23a6_0
  - libopengl=1.7.0=ha4b6fd6_3
  - libpciaccess=0.19=hb03c661_0
  - libpng=1.6.58=h421ea60_0
  - libpq=18.4=hd5a49e9_0
  - libsqlite=3.53.2=h0c1763c_0
  - libstdcxx=15.2.0=h934c35e_19
  - libstdcxx-ng=15.2.0=hdf11a46_19
  - libtiff=4.7.1=h9d88235_1
  - libuuid=2.42.1=h5347b49_0
  - libvulkan-loader=1.4.341.0=h5279c79_0
  - libwebp-base=1.6.0=hd42ef1d_0
  - libxcb=1.17.0=h8a09558_0
  - libxcrypt=4.4.36=hd590300_1
  - libxkbcommon=1.13.2=hca5e8e5_0
  - libxml2=2.15.3=h49c6c72_0
  - libxml2-16=2.15.3=hca6bf5a_0
  - libxslt=1.1.43=h711ed8c_1
  - libzlib=1.3.2=h25fd6f3_2
  - llvmlite=0.47.0=py311h41a00d4_1
  - matplotlib=3.10.9=py311h38be061_0
  - matplotlib-base=3.10.9=py311h0f3be63_0
  - munkres=1.1.4=pyhd8ed1ab_1
  - narwhals=2.22.1=pyhcf101f3_0
  - ncurses=6.6=hdb14827_0
  - numba=0.65.1=py311h3c884d5_1
  - numpy=2.4.6=py311h2e04523_0
  - openjpeg=2.5.4=h55fea9a_0
  - openldap=2.6.13=hbde042b_0
  - openssl=3.6.2=h35e630c_0
  - packaging=26.2=pyhc364b38_0
  - pandas=3.0.3=py311h8032f78_0
  - pcre2=10.47=haa7fec5_0
  - pillow=12.2.0=py311hf88fc01_0
  - pip=26.1.2=pyh8b19718_0
  - pixman=0.46.4=h54a6638_1
  - pluggy=1.6.0=pyhf9edf01_1
  - pthread-stubs=0.4=hb9d3cd8_1002
  - pygments=2.20.0=pyhd8ed1ab_0
  - pyparsing=3.3.2=pyhcf101f3_0
  - pyside6=6.11.1=py311hf27b23e_1
  - pytest=9.0.3=pyhc364b38_1
  - python=3.11.15=hd63d673_0_cpython
  - python-dateutil=2.9.0.post0=pyhe01879c_2
  - python_abi=3.11=8_cp311
  - qhull=2020.2=h434a139_5
  - qt6-main=6.11.1=pl5321h16c4a6b_1
  - readline=8.3=h853b02a_0
  - scikit-learn=1.9.0=np2py311ha15b03d_0
  - scipy=1.17.1=py311hbe70eeb_1
  - six=1.17.0=pyhe01879c_1
  - threadpoolctl=3.6.0=pyhecae5ae_0
  - tk=8.6.13=noxft_h366c992_103
  - tomli=2.4.1=pyhcf101f3_0
  - tornado=6.5.6=py311h49ec1c0_0
  - typing_extensions=4.15.0=pyhcf101f3_0
  - tzdata=2025c=hc9c84f9_1
  - unicodedata2=17.0.1=py311h49ec1c0_0
  - wayland=1.25.0=hd6090a7_0
  - wheel=0.47.0=pyhd8ed1ab_0
  - xcb-util=0.4.1=h4f16b4b_2
  - xcb-util-cursor=0.1.6=hb03c661_0
  - xcb-util-image=0.4.0=hb711507_2
  - xcb-util-keysyms=0.4.1=hb711507_0
  - xcb-util-renderutil=0.3.10=hb711507_0
  - xcb-util-wm=0.4.2=hb711507_0
  - xkeyboard-config=2.47=hb03c661_0
  - xorg-libice=1.1.2=hb9d3cd8_0
  - xorg-libsm=1.2.6=he73a12e_0
  - xorg-libx11=1.8.13=he1eb515_0
  - xorg-libxau=1.0.12=hb03c661_1
  - xorg-libxcomposite=0.4.7=hb03c661_0
  - xorg-libxcursor=1.2.3=hb9d3cd8_0
  - xorg-libxdamage=1.1.6=hb9d3cd8_0
  - xorg-libxdmcp=1.1.5=hb03c661_1
  - xorg-libxext=1.3.7=hb03c661_0
  - xorg-libxfixes=6.0.2=hb03c661_0
  - xorg-libxi=1.8.3=hb03c661_0
  - xorg-libxrandr=1.5.5=hb03c661_0
  - xorg-libxrender=0.9.12=hb9d3cd8_0
  - xorg-libxtst=1.2.5=hb9d3cd8_3
  - xorg-libxxf86vm=1.1.7=hb03c661_0
  - xorg-xorgproto=2025.1=hb03c661_0
  - zlib-ng=2.3.3=hceb46e0_1
  - zstd=1.5.7=hb78ec9c_6
  - pip:
      - annotated-doc==0.0.4
      - anyio==4.13.0
      - certifi==2026.5.20
      - click==8.4.1
      - filelock==3.29.0
      - fsspec==2026.4.0
      - h11==0.16.0
      - hf-xet==1.5.0
      - httpcore==1.0.9
      - httpx==0.28.1
      - huggingface-hub==1.18.0
      - idna==3.18
      - jinja2==3.1.6
      - markdown-it-py==4.2.0
      - markupsafe==3.0.3
      - mdurl==0.1.2
      - mpmath==1.3.0
      - networkx==3.6.1
      - pynndescent==0.6.0
      - pyyaml==6.0.3
      - regex==2026.5.9
      - rich==15.0.0
      - safetensors==0.7.0
      - sentence-transformers==5.5.1
      - setuptools==70.2.0
      - shellingham==1.5.4
      - survey-semantics==0.1.0
      - sympy==1.14.0
      - tokenizers==0.22.2
      - torch==2.12.0+cpu
      - tqdm==4.68.1
      - transformers==5.10.2
      - typer==0.25.1
      - umap-learn==0.5.12
prefix: /data_gpu5/semantic_framework_vipul/env
```
