## Supplementary material for "Auditable cross-instrument detection of unusual multivariate psychiatric response configurations using a semantically aligned covariance subspace": Code for tool: environment-bge-m3.yml

```
# Modern survey-semantics environment with the bge-m3 (sentence-transformers)
# embedding backend (Python 3.11, modern sci stack, CPU torch).
#
# This project installs to a PREFIX env on the project disk (not a named env in
# $HOME). Recreate with:
#
#   source ~/miniconda3/bin/activate
#   export CONDA_PKGS_DIRS=/data_gpu5/semantic_framework_vipul/.conda_pkgs
#   conda env create -f environment-bge-m3.yml -p /data_gpu5/semantic_framework_vipul/env
#
# Then apply the libstdc++ activation fix and download the model as described in
# docs/env_setup.md. The exact resolved versions are pinned in
# environment-bge-m3.lock.yml.
name: survey-semantics-bge-m3
channels:
  - conda-forge
dependencies:
  - python=3.11
  - numpy>=1.24
  - pandas>=2.0
  - scipy>=1.10
  - scikit-learn>=1.3
  - matplotlib>=3.7
  - numba
  - pip
  - pytest
  - pip:
      # CPU torch is sufficient: we embed only a few dozen short prompts.
      - --extra-index-url https://download.pytorch.org/whl/cpu
      - torch
      - sentence-transformers>=2.2
      - transformers>=4.20
      - huggingface-hub>=0.20
      - umap-learn>=0.5
```
