## Supplementary material for "Auditable cross-instrument detection of unusual multivariate psychiatric response configurations using a semantically aligned covariance subspace": Code for tool: analyze_ross.py

```
#!/usr/bin/env python3
"""Compare ROSS sparse components vs dense PCA on the same (orientation-B) matrix,
and sweep Cosine-Preserving Pruning (CPP) to show the sparsity/fidelity tradeoff.

Runs in the main conda env (numpy/sklearn only); CPP is reimplemented (pure numpy).
"""
import argparse
import json
import os

import numpy as np
from sklearn.decomposition import PCA


def cpp_prune_at_theta(W, theta_deg):
    """Cosine-Preserving Pruning: zero smallest entries of each row of W (k,p)
    while retaining >= cos^2(theta) of its energy (direction within theta deg)."""
    cos_t = np.cos(np.radians(theta_deg))
    out = W.copy()
    for j in range(W.shape[0]):
        e_tot = np.sum(W[j] ** 2)
        if e_tot == 0:
            continue
        order = np.argsort(np.abs(W[j]))
        e_rem = e_tot - np.cumsum(W[j, order] ** 2)
        can_zero = np.searchsorted(-e_rem, -cos_t ** 2 * e_tot)
        out[j, order[:can_zero]] = 0.0
    return out


def pr(v):
    s2 = v ** 2
    t = s2.sum()
    return float(t ** 2 / (s2 ** 2).sum()) if t > 0 else 0.0


def median_nnz_pr(W):
    nnz = (np.abs(W) > 1e-10).sum(axis=1)
    prs = np.array([pr(W[j]) for j in range(W.shape[0])])
    return float(np.median(nnz)), float(np.median(prs))


def main():
    ap = argparse.ArgumentParser()
    ap.add_argument("--embeddings", default="data/allofus/allofus_items.npz")
    ap.add_argument("--ross-dir", default="data/allofus/ross_out/bge-m3_ls0.1_lo0.1")
    ap.add_argument("--n-components", type=int, default=39)
    ap.add_argument("--thetas", type=float, nargs="+", default=[5, 10, 15, 20, 25])
    args = ap.parse_args()

    data = np.load(args.embeddings, allow_pickle=True)
    items = [str(x) for x in data["items"]]
    Q = np.asarray(data["vectors"], dtype=np.float64)          # (p items, m dims)
    X = Q.T                                                     # orientation B (m dims, p items)
    Xc = X - X.mean(axis=0, keepdims=True)

    # dense PCA in the SAME orientation/centering -> loadings (k, p items)
    pca = PCA(n_components=args.n_components, svd_solver="full").fit(Xc)
    W_pca = pca.components_                                     # (k, p)
    nnz_pca, pr_pca = median_nnz_pr(W_pca)

    W_ross = np.load(os.path.join(args.ross_dir, "loadings.npy"))   # (k, p), continuous
    nnz_ross, pr_ross = median_nnz_pr(W_ross)

    print("DENSE PCA (orientation B):   median nnz {:.0f}/{}  PR {:.0f}".format(nnz_pca, len(items), pr_pca))
    print("ROSS fit (pre-prune):        median nnz {:.0f}/{}  PR {:.0f}".format(nnz_ross, len(items), pr_ross))
    print("\nCPP pruning sweep (ROSS fit): theta -> sparsity / median nnz / energy kept / dir cos")
    rows = []
    for th in args.thetas:
        Wp = cpp_prune_at_theta(W_ross, th)
        zero_frac = float((Wp == 0).mean())
        nnz = float(np.median((np.abs(Wp) > 1e-10).sum(axis=1)))
        # energy kept + direction cosine per component
        ek, cs = [], []
        for j in range(Wp.shape[0]):
            num = np.dot(Wp[j], W_ross[j])
            den = np.linalg.norm(Wp[j]) * np.linalg.norm(W_ross[j])
            cs.append(num / den if den > 0 else 0.0)
            ek.append(np.sum(Wp[j] ** 2) / np.sum(W_ross[j] ** 2))
        print("  theta={:>4.0f}  zeros {:5.1%}  nnz {:5.0f}/{}  energy {:.2f}  cos {:.3f}".format(
            th, zero_frac, nnz, len(items), float(np.mean(ek)), float(np.mean(cs))))
        rows.append({"theta_deg": th, "zero_fraction": zero_frac, "median_nnz": nnz,
                     "mean_energy_kept": float(np.mean(ek)), "mean_dir_cosine": float(np.mean(cs))})

    out = os.path.join(args.ross_dir, "cpp_sweep.json")
    json.dump({"dense_pca": {"median_nnz": nnz_pca, "median_pr": pr_pca},
               "ross_fit": {"median_nnz": nnz_ross, "median_pr": pr_ross},
               "cpp_sweep": rows}, open(out, "w"), indent=2)

    # top items per component at theta=15 (a moderate operating point)
    Wp = cpp_prune_at_theta(W_ross, 15.0)
    lines = ["component,nnz,top_items"]
    for j in range(min(8, Wp.shape[0])):
        idx = np.argsort(-np.abs(Wp[j]))
        idx = [i for i in idx if abs(Wp[j][i]) > 1e-10][:10]
        lines.append("PC{},{},{}".format(j + 1, int((np.abs(Wp[j]) > 1e-10).sum()),
                     "; ".join("{}({:+.2f})".format(items[i], Wp[j][i]) for i in idx)))
    open(os.path.join(args.ross_dir, "top_items_theta15.csv"), "w").write("\n".join(lines) + "\n")
    print("\nwrote", out, "and top_items_theta15.csv")


if __name__ == "__main__":
    main()
```
