## Supplementary material for "Auditable cross-instrument detection of unusual multivariate psychiatric response configurations using a semantically aligned covariance subspace": Code for tool: compare_pca_models.py

```
#!/usr/bin/env python3
"""PCA the All of Us item embeddings per model and compare the components.

Mirrors the tool/manuscript exactly per model: center the (282 x m) item
embedding matrix over items (the "language-model bias vector"), full-solver
PCA, D = first crossing of the 80% cumulative-variance threshold
(``select_component_count``, same function the pipeline uses).

Components from different models live in different m-dimensional spaces, so
they are compared through their **item-projection vectors**: each PC induces a
282-vector of item scores, which is model-agnostic. Comparisons (all
sign-invariant, PCA sign is arbitrary):

1. D selection + cumulative-variance curves per model.
2. Component matching: |corr| between item-score vectors for every PC pair of
   two models; one-to-one assignment via the Hungarian algorithm.
3. Subspace overlap: principal angles between the D-dimensional column spaces
   of the item-score matrices; reported as mean cos^2 (1 = identical span;
   chance for random D-dim subspaces in R^282 is ~D/282).

Outputs to data/allofus/pca_compare/.
"""

import os
import sys

import numpy as np
import pandas as pd
from scipy.linalg import subspace_angles
from scipy.optimize import linear_sum_assignment
from sklearn.decomposition import PCA

SCRIPT_DIR = os.path.dirname(os.path.abspath(__file__))
DATA = os.path.join(SCRIPT_DIR, "..", "..", "..", "data", "allofus")
OUT = os.path.join(DATA, "pca_compare")

sys.path.insert(0, os.path.join(SCRIPT_DIR, "..", "..", "src"))
from survey_semantics.embedding import load_item_embeddings  # noqa: E402
from survey_semantics.pipeline import select_component_count  # noqa: E402

MODELS = [
    ("bge-m3", "allofus_items.npz"),
    ("bge-large-en-v1.5", "allofus_items_bge-large-en-v1.5.npz"),
    ("all-mpnet-base-v2", "allofus_items_all-mpnet-base-v2.npz"),
    ("gte-large-en-v1.5", "allofus_items_gte-large-en-v1.5.npz"),
    ("e5-large-v2", "allofus_items_e5-large-v2.npz"),
    ("qwen3-embedding-0.6b", "allofus_items_qwen3-embedding-0.6b.npz"),
]
VARIANCE_THRESHOLD = 0.80


def fit_model_pca(npz_name):
    emb = load_item_embeddings(os.path.join(DATA, npz_name))
    items = list(emb.items)
    X = emb.matrix_for(items)
    X = X - X.mean(axis=0, keepdims=True)          # bias-vector subtraction
    max_components = min(X.shape)                   # evaluate all PCs
    pca = PCA(n_components=max_components, svd_solver="full")
    scores = pca.fit_transform(X)                   # (282, max_components) item coordinates
    cumulative = np.cumsum(pca.explained_variance_ratio_)
    d80, reached = select_component_count(cumulative, VARIANCE_THRESHOLD, max_components)
    assert reached, "80% variance not reached — unexpected for a full PCA"
    d90 = int(np.argmax(cumulative >= 0.90)) + 1
    return items, scores, cumulative, d80, d90


def standardized(scores_d):
    """Unit-normalize each component's item-score vector (they are mean-zero
    by construction, so |cosine| == |Pearson correlation|)."""
    return scores_d / np.linalg.norm(scores_d, axis=0, keepdims=True)


def main():
    os.makedirs(OUT, exist_ok=True)
    results = {}
    ref_items = None
    rows = []
    for name, fname in MODELS:
        items, scores, cumulative, d80, d90 = fit_model_pca(fname)
        if ref_items is None:
            ref_items = items
        assert items == ref_items, "item order mismatch across artifacts"
        results[name] = {"scores": scores, "cum": cumulative, "d80": d80, "d90": d90}
        rows.append({"model": name, "embed_dim": scores.shape[1] if scores.shape[1] < 282 else "(282 cap)",
                     "D_80pct": d80, "cumvar_at_D": round(float(cumulative[d80 - 1]), 4),
                     "D_90pct": d90})
        print("{:24s} D80={:3d}  D90={:3d}  cumvar@D80={:.3f}".format(name, d80, d90, cumulative[d80 - 1]))
    d_table = pd.DataFrame(rows)
    d_table.to_csv(os.path.join(OUT, "allofus_pca_d_selection.csv"), index=False)

    names = [n for n, _ in MODELS]

    # --- pairwise component matching + subspace overlap ---
    match_rows, overlap = [], pd.DataFrame(np.eye(len(names)), index=names, columns=names)
    for i, a in enumerate(names):
        for j, b in enumerate(names):
            if j <= i:
                continue
            Da, Db = results[a]["d80"], results[b]["d80"]
            A = standardized(results[a]["scores"][:, :Da])
            B = standardized(results[b]["scores"][:, :Db])
            C = np.abs(A.T @ B)                       # (Da, Db) |corr| matrix
            ra, cb = linear_sum_assignment(-C)        # maximize total |corr|
            for pa, pb in zip(ra, cb):
                match_rows.append({"model_a": a, "model_b": b,
                                   "pc_a": pa + 1, "pc_b": pb + 1,
                                   "abs_corr": round(float(C[pa, pb]), 4)})
            angles = subspace_angles(A, B)            # principal angles
            ov = float(np.mean(np.cos(angles) ** 2))
            overlap.loc[a, b] = overlap.loc[b, a] = round(ov, 4)
    matches = pd.DataFrame(match_rows)
    matches.to_csv(os.path.join(OUT, "allofus_pca_matched_components.csv"), index=False)
    overlap.to_csv(os.path.join(OUT, "allofus_pca_subspace_overlap.csv"))

    # --- top-loading items per bge-m3 PC, for interpretation ---
    d_ref = results["bge-m3"]["d80"]
    S = standardized(results["bge-m3"]["scores"][:, :d_ref])
    top = []
    for k in range(d_ref):
        idx = np.argsort(-np.abs(S[:, k]))[:5]
        top.append({"pc": k + 1,
                    "top_items": "; ".join("{}({:+.2f})".format(ref_items[i], S[i, k]) for i in idx)})
    pd.DataFrame(top).to_csv(os.path.join(OUT, "allofus_pca_bge-m3_top_items.csv"), index=False)

    _plots(results, names, matches, overlap)
    print("Outputs in", OUT)
    return 0


def _plots(results, names, matches, overlap):
    import matplotlib
    matplotlib.use("Agg")
    import matplotlib.pyplot as plt

    # 1) cumulative variance curves with D80 marks
    fig, ax = plt.subplots(figsize=(9, 6))
    cmap = plt.get_cmap("tab10")
    for i, n in enumerate(names):
        cum, d80 = results[n]["cum"], results[n]["d80"]
        x = np.arange(1, len(cum) + 1)
        ax.plot(x[:120], cum[:120], color=cmap(i), label="{} (D={})".format(n, d80))
        ax.scatter([d80], [cum[d80 - 1]], color=cmap(i), zorder=5, s=30)
    ax.axhline(VARIANCE_THRESHOLD, color="gray", ls="--", lw=1, label="80% threshold")
    ax.set_xlabel("PCA components"); ax.set_ylabel("Cumulative explained variance")
    ax.set_title("Prompt-embedding PCA: cumulative variance per model (282 items)")
    ax.legend(fontsize=9); ax.spines[["top", "right"]].set_visible(False)
    fig.tight_layout(); fig.savefig(os.path.join(OUT, "allofus_pca_variance.png"), dpi=150)
    plt.close(fig)

    # 2) |corr| heatmaps: bge-m3 PCs vs each other model's PCs
    others = [n for n in names if n != "bge-m3"]
    Dref = results["bge-m3"]["d80"]
    A = standardized(results["bge-m3"]["scores"][:, :Dref])
    fig, axes = plt.subplots(1, len(others), figsize=(4.2 * len(others), 4.6))
    for ax, n in zip(np.atleast_1d(axes), others):
        Dn = results[n]["d80"]
        B = standardized(results[n]["scores"][:, :Dn])
        C = np.abs(A.T @ B)
        im = ax.imshow(C, vmin=0, vmax=1, cmap="viridis", aspect="auto")
        ax.set_title("bge-m3 vs {}".format(n), fontsize=9)
        ax.set_xlabel("{} PC".format(n), fontsize=8); ax.set_ylabel("bge-m3 PC", fontsize=8)
    fig.colorbar(im, ax=axes, shrink=0.8, label="|corr| of item-score vectors")
    fig.suptitle("Component correspondence to the primary model (item-projection |corr|)", fontsize=11)
    fig.savefig(os.path.join(OUT, "allofus_pca_match_bge-m3.png"), dpi=150, bbox_inches="tight")
    plt.close(fig)

    # 3) subspace-overlap matrix
    fig, ax = plt.subplots(figsize=(7, 5.5))
    im = ax.imshow(overlap.to_numpy(dtype=float), vmin=0, vmax=1, cmap="viridis")
    ax.set_xticks(range(len(names)), names, rotation=45, ha="right", fontsize=8)
    ax.set_yticks(range(len(names)), names, fontsize=8)
    for i in range(len(names)):
        for j in range(len(names)):
            v = float(overlap.iloc[i, j])
            ax.text(j, i, "{:.2f}".format(v), ha="center", va="center",
                    color="white" if v < 0.6 else "black", fontsize=8)
    ax.set_title("Retained-subspace overlap (mean cos² of principal angles)\n"
                 "1 = identical span; chance ≈ D/282 ≈ 0.1–0.2", fontsize=10)
    fig.colorbar(im, ax=ax, shrink=0.85)
    fig.tight_layout(); fig.savefig(os.path.join(OUT, "allofus_pca_subspace_overlap.png"), dpi=150)
    plt.close(fig)


if __name__ == "__main__":
    raise SystemExit(main())
```
