## Supplementary material for "Auditable cross-instrument detection of unusual multivariate psychiatric response configurations using a semantically aligned covariance subspace": Code for tool: convert_allofus.py

```
#!/usr/bin/env python3
"""All of Us survey codebook -> reviewable item inventory.

The All of Us codebook is a REDCap data dictionary: one Excel sheet per survey,
one row per item. This script flattens the *ordinal* survey items (radio /
dropdown / slider field types) into a single CSV, tagging each with a suggested
``category`` and an ``embed`` flag.

That CSV is the **human-review artifact**: eyeball it, adjust ``embed`` (and
``category``) as needed, then a later step turns the ``embed=true`` rows into the
pipeline's ``prompts.csv`` / ``scales.csv``.

Categorization is rule-based and auditable: an instrument-prefix map (the All of
Us ``Item Concept`` prefixes are validated-instrument names, e.g. ``gad7_*``,
``phq*``) plus a keyword fallback on the question text / section header.

Usage:
    python convert_allofus.py \
        --codebook ../../../data/allofus/All_of_Us_Survey_Data_Codebooks.xlsx \
        --out      ../../../data/allofus/allofus_item_inventory.csv
"""

import argparse
import os
import re
import sys

import pandas as pd

SCRIPT_DIR = os.path.dirname(os.path.abspath(__file__))

# Survey sheets to scan (scoring sheets, ReadMe, and the odd-format COVID minute
# survey are skipped automatically because they lack the REDCap columns).
ORDINAL_FIELD_TYPES = {"radio", "dropdown", "slider"}

# No sheets are skipped by name. Non-item sheets (ReadMe, the 6 scoring sheets,
# and the odd-format "Minute Survey on COVID-19 Vacci") are excluded automatically
# because they lack the REDCap item columns. Survey-version overlaps — e.g.
# "Personal and Family Health Hist" shares 576/705 concepts with the two split
# physical-history sheets — are resolved deterministically by the dedup-by-concept
# rule below (keep first occurrence in codebook order), not by an arbitrary skip.
SKIP_SHEETS = set()

# ── Categorization rules (edit here) ─────────────────────────────────────────
# Instrument prefix (token before the first digit/underscore of Item Concept).
PREFIX_CATEGORY = {
    # psychological — symptom / trait
    "gad": "anxiety", "worryanxiety": "anxiety",
    "phq": "depression",
    "pcl": "trauma", "ies": "trauma", "ace": "trauma",
    "cidi": "psychiatric", "mhqukb": "psychiatric", "ukmh": "psychiatric",
    "ss": "psychiatric",  # suicidality items (thoughts of self-harm / suicide)
    "bfi": "personality",
    "asrs": "adhd",
    "cpss": "stress",
    "brcs": "wellbeing", "lot": "wellbeing",
    # habit / behavioral
    "smoking": "substance", "cigarsmoking": "substance", "electronicsmoking": "substance",
    "hookahsmoking": "substance", "smokelesstobacco": "substance", "attemptquitsmoking": "substance",
    "alcohol": "substance", "past": "substance", "tsu": "substance", "audit": "substance",
    "ipaq": "activity",
    # psychosocial
    "eds": "psychosocial", "mos": "psychosocial", "nds": "psychosocial",
    "bmmrs": "psychosocial", "scns": "psychosocial", "sdoh": "psychosocial", "hvs": "psychosocial",
    "nhs": "psychosocial", "ucla": "psychosocial",
    # neighborhood built environment / language -> out of scope (only ips_16,
    # perceived safety, is kept, via ITEM_OVERRIDE below)
    "ips": "neighborhood_built", "chis": "demographic",
    # functioning
    "disability": "functioning",
    # out of scope
    "cancer": "physical", "nervoussystem": "physical", "infectiousdiseases": "physical",
    "circulatory": "physical", "digestive": "physical", "skeletalmuscular": "physical",
    "endocrine": "physical", "respiratory": "physical", "other": "physical",
    "hearingvision": "physical", "kidney": "physical", "dmfs": "physical",
    "mentalhealth": "mh_diagnosis", "familyhistory": "family_history",
    "livingsituation": "demographic", "biologicalsexatbirth": "demographic",
    "genderidentity": "demographic", "educationlevel": "demographic", "activeduty": "demographic",
    "thebasics": "demographic", "maritalstatus": "demographic", "employment": "demographic",
    "employmentworkaddress": "demographic", "income": "demographic", "homeown": "demographic",
    "socialsecurity": "demographic", "secondarycontactinfo": "demographic",
    "persononeaddress": "demographic", "secondcontactsaddress": "demographic", "basics": "demographic",
    "insurance": "access", "healthadvice": "access", "cantaffordcare": "access",
    "delayedmedicalcare": "access", "healthproviderracereligion": "access",
    "overallhealth": "health_perception", "rand": "health_perception",
    "pregnancy": "reproductive", "yesnone": "out", "copect": "covid", "cdc": "covid",
    "msds": "covid", "section": "admin", "pmi": "admin",
}

# Fallback: (keywords, category) tried in order against label+section text.
KEYWORD_CATEGORY = [
    (("depress", "little interest", "feeling down", "hopeless"), "depression"),
    (("nervous", "anxious", "on edge", "worry", "panic", "afraid"), "anxiety"),
    (("stress", "overwhelm", "unable to cope"), "stress"),
    (("nightmare", "flashback", "frightening"), "trauma"),
    (("discriminat", "courtesy", "poorer service", "treated with less"), "psychosocial"),
    (("lonely", "companionship", "isolat", "left out"), "psychosocial"),
    (("religi", "spiritual", "faith", "prayer"), "psychosocial"),
    (("smok", "cigarett", "tobacco", "nicotine", "vap", "alcohol", "drink", "marijuana", "cannabis", "substance"), "substance"),
    (("physical activity", "exercise", "walk", "vigorous", "moderate activity"), "activity"),
    (("see myself as", "am someone who", "tend to"), "personality"),
    (("concentrat", "fidget", "restless", "hyperactiv"), "adhd"),
    (("difficulty", "functioning", "because of a health problem"), "functioning"),
]

# Exact-item overrides (checked before the prefix map). Use for single items whose
# category differs from their instrument's default. Source: expert review (see
# docs/allofus_expert_review.md).
ITEM_OVERRIDE = {
    # ips_16 = perceived neighborhood safety (a psychological appraisal) kept in,
    # while the rest of the ips walkability scale (sidewalks/transit/housing) is out.
    "ips_16": "psychosocial",
    # --- Decision 4: PROMIS Global mental-health items + housing-instability worry,
    # brought in (their parent surveys are otherwise out of scope). ---
    "overallhealth_emotionalproblem7days": "psychiatric",
    "overallhealth_generalmentalhealth": "psychiatric",
    "overallhealth_socialsatisfaction": "psychosocial",
    "livingsituation_stablehouseconcern": "psychosocial",
    # --- Decision 3: factual medication-response history / COVID exposure -> out. ---
    "mhqukb_31_amitriptyline": "med_response", "mhqukb_31_citalopram": "med_response",
    "mhqukb_31_dosulepin": "med_response", "mhqukb_31_fluoxetine": "med_response",
    "mhqukb_31_paroxetine": "med_response", "mhqukb_31_sertraline": "med_response",
    "mhqukb_31_other": "med_response",
    "nhs_covid_fhc17a": "covid",
    # --- Decision 2: genuinely nominal / non-monotone response options -> out
    # (no valid ordinal scale; cannot be salvaged by recoding a single option). ---
    "mhqukb_11": "nominal",  # mood worse: morning / evening / did not vary
    "mhqukb_14": "nominal",  # appetite: no change / increased / decreased
    "mhqukb_15": "nominal",  # weight: gained / lost / both / same
}

# --- Decision 1: twin instruments. These validated scales appear twice — a COPE
# (COVID-wave) copy and a home-survey copy (EHH / SDOH). Near-identical wording
# yields near-duplicate prompt embeddings (degenerate PCA basis) and collinear
# responses. Keep the home-survey copy; drop the COPE copy. Keyed (survey, prefix)
# so only the COPE administration is removed. ---
DUPLICATE_DROP = {
    ("COPE", "gad"),   # gad_7_*  -> keep EHH gad7_*
    ("COPE", "phq"),   # phq_9_*  -> keep EHH phq9_*
    ("COPE", "cpss"),  # cpss_*   -> keep SDOH sdoh_cpss_*
    ("COPE", "eds"),   # eds_*    -> keep SDOH sdoh_eds_*
    ("COPE", "mos"),   # mos_ss_* -> keep SDOH sdoh_mos_ss_*
    ("COPE", "ucla"),  # ucla_ls8_* -> keep SDOH sdoh_ucla_ls8_*
}

# --- Decision 2 (recode): items kept as clean binaries by treating one off-axis
# option as a sentinel (not a scale point). These extra codes are excluded from
# n_levels_substantive and must become sentinels in the generated scales.csv. ---
EXTRA_SENTINEL_CODES = {
    "ace_5": {"ehhwb_60"},  # "Parents not married" — off the Yes/No axis
}

# Categories that count as "psychological or habit" -> embed.
IN_SCOPE = {
    "anxiety", "depression", "trauma", "psychiatric", "personality", "adhd",
    "stress", "wellbeing", "substance", "activity", "psychosocial", "functioning",
}

# Sentinel options (skip / don't know / prefer not to answer) are not scale
# points: they are excluded from n_levels_substantive and must become per-item
# sentinel codes, not the scale max, in the generated scales.csv.
SENTINEL_LABEL_RE = re.compile(
    r"(?i)^(don'?t know|prefer not to answer|skip)\b")


def _parse_choices(choices):
    """REDCap choices string -> list of (code, label).

    Format: ``code, label | code, label``; labels may contain commas, so split
    each segment on the first comma only.
    """
    text = str(choices)
    if not text or text.lower() == "nan":
        return []
    parsed = []
    for segment in text.split("|"):
        segment = segment.strip()
        if not segment:
            continue
        code, _, label = segment.partition(",")
        parsed.append((code.strip(), label.strip()))
    return parsed


def _is_sentinel(code, label):
    return code.lower().startswith("pmi") or bool(SENTINEL_LABEL_RE.match(label))


def _is_free_entry(parsed):
    """Radio wrapper around free-text entry ("Enter age | Don't know"): the
    choices carry no scale points, so order-coding is meaningless -> out."""
    return any(label.lower().startswith("enter") for _, label in parsed)


def _prefix(concept):
    match = re.match(r"^([a-zA-Z]+)", str(concept))
    return match.group(1).lower() if match else ""


def categorize(concept, label, section, survey):
    if str(concept) in ITEM_OVERRIDE:
        return ITEM_OVERRIDE[str(concept)]
    prefix = _prefix(concept)
    if (survey, prefix) in DUPLICATE_DROP:
        return "duplicate_cope"
    if prefix in PREFIX_CATEGORY:
        return PREFIX_CATEGORY[prefix]
    text = "{} {}".format(label, section).lower()
    for keywords, category in KEYWORD_CATEGORY:
        if any(k in text for k in keywords):
            return category
    return "uncategorized"


def _clean(value):
    return re.sub(r"\s+", " ", str(value)).strip()


def _n_levels(parsed, field_type):
    if field_type == "slider":
        # Slider choices are endpoint *anchors* ("0 (No pain) | | 10 (Worst
        # pain imaginable)"), not levels: parse the numeric endpoints.
        endpoints = [m for code, label in parsed
                     for m in [re.match(r"^(-?\d+)", code or label)] if m]
        if len(endpoints) >= 2:
            lo, hi = int(endpoints[0].group(1)), int(endpoints[-1].group(1))
            return abs(hi - lo) + 1
    return len(parsed)


def _n_levels_substantive(parsed, extra_codes=frozenset()):
    return len([1 for code, label in parsed
                if not _is_sentinel(code, label) and code not in extra_codes])


def build_inventory(codebook_path):
    xl = pd.ExcelFile(codebook_path)
    records = []
    for sheet in xl.sheet_names:
        if sheet.strip() in SKIP_SHEETS:
            continue
        df = xl.parse(sheet)
        cols = set(df.columns)
        if not {"Item Concept", "Field Type", "Field Label"}.issubset(cols):
            continue  # scoring sheets / ReadMe / odd-format sheets
        for _, row in df.iterrows():
            field_type = str(row["Field Type"]).strip().lower()
            if field_type not in ORDINAL_FIELD_TYPES:
                continue
            concept = _clean(row["Item Concept"])
            if not concept or concept.lower() == "nan":
                continue
            label = _clean(row["Field Label"])
            raw_section = row.get("Section Header")
            section = "" if pd.isna(raw_section) else _clean(raw_section)
            choices = row.get("Choices, Calculations, OR Slider Labels", "")
            parsed = _parse_choices(choices)
            n_levels = _n_levels(parsed, field_type)
            # Slider anchors are endpoint labels, not options: no sentinels.
            n_substantive = (n_levels if field_type == "slider"
                             else _n_levels_substantive(
                                 parsed, EXTRA_SENTINEL_CODES.get(concept, frozenset())))
            if _is_free_entry(parsed):
                category = "free_entry"
            else:
                category = categorize(concept, label, section, sheet.strip())
            records.append({
                "survey": sheet.strip(),
                "item": concept,
                "form": _clean(row.get("Form Name", "")),
                "section": section,
                "field_type": field_type,
                "prompt": label,
                "n_levels": n_levels,
                "n_levels_substantive": n_substantive,
                "choices": _clean(choices),
                "category": category,
                "embed": str(category in IN_SCOPE).lower(),
            })
    df = pd.DataFrame(records)
    dead = [p for p in PREFIX_CATEGORY
            if not any(_prefix(c) == p for c in df["item"])]
    if dead:
        print("WARNING: PREFIX_CATEGORY entries matching zero items "
              "(codebook drift?): {}".format(", ".join(sorted(dead))),
              file=sys.stderr)
    return df


def print_summary(inv):
    inv_in = inv[inv["embed"] == "true"]
    print("\n==== in-scope per survey ====")
    g = inv.assign(ins=inv["embed"] == "true").groupby("survey").agg(
        ins=("ins", "sum"), ordinal=("ins", "size"))
    for survey, r in g.sort_values("ins", ascending=False).iterrows():
        print("  {:34s} {:4d} / {:4d}".format(survey, int(r["ins"]), int(r["ordinal"])))
    print("  {:34s} {:4d} / {:4d}".format("TOTAL", len(inv_in), len(inv)))
    print("\n==== in-scope by category ====")
    for cat, n in inv_in["category"].value_counts().items():
        print("  {:14s} {}".format(cat, n))
    uncat = int((inv["category"] == "uncategorized").sum())
    if uncat:
        print("\n  uncategorized (review): {}".format(uncat))


def main(argv=None):
    parser = argparse.ArgumentParser(description="All of Us codebook -> item inventory")
    parser.add_argument(
        "--codebook",
        default=os.path.join(SCRIPT_DIR, "..", "..", "..", "data", "allofus",
                             "All_of_Us_Survey_Data_Codebooks.xlsx"),
    )
    parser.add_argument(
        "--out",
        default=os.path.join(SCRIPT_DIR, "..", "..", "..", "data", "allofus",
                             "allofus_item_inventory.csv"),
    )
    parser.add_argument(
        "--prompts-out", default=None,
        help="Also write a prompts.csv (item,prompt) of the embed=true items only — "
             "the sole input the embedding step needs. Downstream scale/reverse "
             "metadata is produced separately.",
    )
    args = parser.parse_args(argv)

    inv = build_inventory(args.codebook)
    os.makedirs(os.path.dirname(os.path.abspath(args.out)), exist_ok=True)

    # The same Item Concept can appear in multiple sheets/survey waves (e.g.
    # disability items in both Basics and Life Functioning; gad/phq across COPE
    # versions). The pipeline needs unique item columns, so keep one row per
    # Item Concept (first occurrence in codebook order).
    n_before = len(inv)
    inv = inv.drop_duplicates(subset="item", keep="first")
    n_dups = n_before - len(inv)

    # Stable, reviewable order: in-scope first, then by survey + item.
    inv = inv.sort_values(
        ["embed", "survey", "item"], ascending=[False, True, True]
    ).reset_index(drop=True)
    inv.to_csv(args.out, index=False)
    print("Wrote {} unique items ({} embed=true; {} duplicate concepts collapsed) -> {}".format(
        len(inv), int((inv["embed"] == "true").sum()), n_dups, args.out))
    print_summary(inv)

    if args.prompts_out:
        _write_prompts(inv, args.prompts_out)
    return 0


def _write_prompts(inv, path):
    """Emit prompts.csv (item,prompt) for the embed=true items — the embedding
    step's only input. Item Concept is the stable, unique key (dedup guarantees
    uniqueness); the prompt is the verbatim Field Label."""
    prompts = inv[inv["embed"] == "true"][["item", "prompt"]].copy()
    assert not prompts["item"].duplicated().any(), "duplicate item keys in prompts"
    assert (prompts["prompt"].str.strip() != "").all(), "empty prompt text"
    os.makedirs(os.path.dirname(os.path.abspath(path)), exist_ok=True)
    prompts.to_csv(path, index=False)
    print("Wrote {} embed=true prompts -> {}".format(len(prompts), path))


if __name__ == "__main__":
    sys.exit(main())
```
