## Supplementary material for "Auditable cross-instrument detection of unusual multivariate psychiatric response configurations using a semantically aligned covariance subspace": Code for tool: plot_item_umap.py

```
#!/usr/bin/env python3
"""2D UMAP of the All of Us item embeddings, colored by a chosen grouping.

Reads the embedding artifact (``allofus_items.npz``) and the inventory
(``allofus_item_inventory.csv``), runs UMAP on the 1024-d bge-m3 vectors, and
writes a scatter plot + the 2D coordinates. Color by any of three grouping
levels (``--color-by``):

- ``survey``     — the broad AoU survey *module* (Emotional Health History, …).
- ``category``   — the rule-assigned *construct* bucket (anxiety, depression, …).
- ``instrument`` — the actual *psychometric form* (GAD-7, PHQ-9, IES-6, …),
  derived from the item-concept prefix (with the ``sdoh_`` wrapper unwrapped to
  its 2nd token, e.g. ``sdoh_cpss_1`` -> ``sdoh/cpss``).

The embedding is the *only* input the semantic basis uses, so this view shows
how the item wording clusters — and whether a grouping lines up with semantic
neighborhoods. To keep the layout fixed across colorings, the UMAP coordinates
are computed once and cached in the CSV; pass ``--reuse-coords`` to recolor.
"""

import argparse
import os
import re
import sys

import pandas as pd

SCRIPT_DIR = os.path.dirname(os.path.abspath(__file__))
DATA = os.path.join(SCRIPT_DIR, "..", "..", "..", "data", "allofus")

sys.path.insert(0, os.path.join(SCRIPT_DIR, "..", "..", "src"))
from survey_semantics.embedding import load_item_embeddings  # noqa: E402


def instrument_of(item):
    """The psychometric instrument for an item concept. ``sdoh_`` is an AoU
    wrapper whose 2nd token names the real instrument (cpss/eds/mos/ucla/dms)."""
    it = str(item)
    if it.startswith("sdoh_"):
        m = re.match(r"([a-zA-Z]+)", it[len("sdoh_"):])
        return "sdoh/" + (m.group(1) if m else it[len("sdoh_"):])
    m = re.match(r"([a-zA-Z]+)", it)
    return m.group(1) if m else it


def main(argv=None):
    p = argparse.ArgumentParser(description="2D UMAP of AoU item embeddings")
    p.add_argument("--color-by", choices=["survey", "instrument"],
                   default="survey",
                   help="Pre-labeled groupings only: 'survey' (AoU module / REDCap "
                        "Form Name) or 'instrument' (AoU Item Concept variable name).")
    p.add_argument("--annotate", action="store_true",
                   help="Label each group's cluster centroid on the plot (clearer "
                        "than a long legend for the ~39 instruments).")
    p.add_argument("--embeddings", default=os.path.join(DATA, "allofus_items.npz"))
    p.add_argument("--inventory", default=os.path.join(DATA, "allofus_item_inventory.csv"))
    p.add_argument("--out-png", default=None, help="Defaults to allofus_items_umap_<color-by>.png")
    p.add_argument("--coords-csv", default=os.path.join(DATA, "allofus_items_umap.csv"))
    p.add_argument("--reuse-coords", action="store_true",
                   help="Reuse umap_x/umap_y from --coords-csv instead of recomputing.")
    p.add_argument("--n-neighbors", type=int, default=15)
    p.add_argument("--min-dist", type=float, default=0.10)
    p.add_argument("--metric", default="cosine", help="bge-m3 vectors compare by cosine.")
    p.add_argument("--random-state", type=int, default=42)
    args = p.parse_args(argv)

    emb = load_item_embeddings(args.embeddings)
    items = list(emb.items)

    inv = pd.read_csv(args.inventory).set_index("item")
    survey_of = inv["survey"].to_dict()
    missing = [it for it in items if it not in survey_of]
    if missing:
        raise SystemExit("No inventory row for {} items, e.g. {}".format(len(missing), missing[:5]))

    if args.reuse_coords:
        cached = pd.read_csv(args.coords_csv).set_index("item")
        coords = cached.loc[items, ["umap_x", "umap_y"]].to_numpy()
    else:
        import umap  # imported late so --help works without the dep
        coords = umap.UMAP(
            n_neighbors=args.n_neighbors, min_dist=args.min_dist,
            metric=args.metric, random_state=args.random_state,
        ).fit_transform(emb.matrix_for(items))

    # Only pre-labeled groupings: 'survey' (AoU module) and 'instrument' (AoU
    # Item Concept variable name, parsed deterministically). No model-assigned labels.
    frame = pd.DataFrame({"item": items, "survey": [survey_of[i] for i in items],
                          "instrument": [instrument_of(i) for i in items],
                          "umap_x": coords[:, 0], "umap_y": coords[:, 1]})
    if not args.reuse_coords:
        frame[["item", "survey", "instrument", "umap_x", "umap_y"]].to_csv(
            args.coords_csv, index=False)

    out_png = args.out_png or os.path.join(
        DATA, "allofus_items_umap_{}.png".format(args.color_by))
    _plot(frame, args.color_by, out_png, args.metric, args.n_neighbors,
          args.min_dist, args.annotate)
    print("Wrote {} ({} items, {} {} groups)".format(
        out_png, len(items), frame[args.color_by].nunique(), args.color_by))
    return 0


def _palette(n):
    """A qualitative palette big enough for up to ~40 groups."""
    import matplotlib.pyplot as plt
    if n <= 10:
        return [plt.get_cmap("tab10")(i) for i in range(n)]
    if n <= 20:
        return [plt.get_cmap("tab20")(i) for i in range(n)]
    cols = ([plt.get_cmap("tab20")(i) for i in range(20)]
            + [plt.get_cmap("tab20b")(i) for i in range(20)]
            + [plt.get_cmap("tab20c")(i) for i in range(20)])
    return cols[:n]


def _plot(frame, color_by, out_png, metric, n_neighbors, min_dist, annotate=False):
    import matplotlib
    matplotlib.use("Agg")
    import matplotlib.pyplot as plt

    # Largest group first so its color reads as the "background".
    order = frame[color_by].value_counts().index.tolist()
    palette = _palette(len(order))
    colors = {g: palette[i] for i, g in enumerate(order)}

    fig, ax = plt.subplots(figsize=(13, 8.5) if len(order) > 12 else (11, 8))
    for g in order:
        sub = frame[frame[color_by] == g]
        ax.scatter(sub["umap_x"], sub["umap_y"], s=42, alpha=0.85,
                   color=colors[g], edgecolor="white", linewidth=0.4,
                   label="{} ({})".format(g, len(sub)))

    if annotate:
        # Label each group's centroid directly — clearer than a long legend.
        for g in order:
            sub = frame[frame[color_by] == g]
            cx, cy = sub["umap_x"].median(), sub["umap_y"].median()
            ax.text(cx, cy, g, fontsize=8, fontweight="bold", ha="center",
                    va="center", color="black",
                    bbox=dict(boxstyle="round,pad=0.15", fc="white", ec="none", alpha=0.7))

    ax.set_title("All of Us item embeddings (bge-m3) — 2D UMAP, colored by {}\n"
                 "{} items · metric={} · n_neighbors={} · min_dist={}".format(
                     color_by, len(frame), metric, n_neighbors, min_dist), fontsize=11)
    ax.set_xlabel("UMAP-1")
    ax.set_ylabel("UMAP-2")
    if not annotate:
        ncol = 2 if len(order) > 16 else 1
        ax.legend(title=color_by.capitalize(), loc="center left",
                  bbox_to_anchor=(1.0, 0.5), fontsize=8, ncol=ncol, framealpha=0.9)
    ax.spines[["top", "right"]].set_visible(False)
    fig.tight_layout()
    fig.savefig(out_png, dpi=150, bbox_inches="tight")
    plt.close(fig)


if __name__ == "__main__":
    raise SystemExit(main())
```
