## Supplementary material for "Auditable cross-instrument detection of unusual multivariate psychiatric response configurations using a semantically aligned covariance subspace": Code for tool: plot_item_umap_models.py

```
#!/usr/bin/env python3
"""6-panel UMAP comparison of the All of Us item embeddings across models.

One panel per embedding artifact, identical UMAP parameters (cosine, seed 42),
colored by the pre-labeled survey form. Shows whether the semantic neighborhood
structure of the 282 item prompts is stable across encoders — the qualitative
counterpart of the manuscript's cross-embedder sensitivity analysis.
"""

import argparse
import os
import sys

import pandas as pd

SCRIPT_DIR = os.path.dirname(os.path.abspath(__file__))
DATA = os.path.join(SCRIPT_DIR, "..", "..", "..", "data", "allofus")

sys.path.insert(0, os.path.join(SCRIPT_DIR, "..", "..", "src"))
from survey_semantics.embedding import load_item_embeddings  # noqa: E402

MODELS = [
    ("bge-m3 (primary)", "allofus_items.npz"),
    ("bge-large-en-v1.5", "allofus_items_bge-large-en-v1.5.npz"),
    ("all-mpnet-base-v2", "allofus_items_all-mpnet-base-v2.npz"),
    ("gte-large-en-v1.5", "allofus_items_gte-large-en-v1.5.npz"),
    ("e5-large-v2", "allofus_items_e5-large-v2.npz"),
    ("qwen3-embedding-0.6b", "allofus_items_qwen3-embedding-0.6b.npz"),
]


def main(argv=None):
    p = argparse.ArgumentParser(description="UMAP grid across embedding models")
    p.add_argument("--inventory", default=os.path.join(DATA, "allofus_item_inventory.csv"))
    p.add_argument("--out-png", default=os.path.join(DATA, "allofus_items_umap_models.png"))
    p.add_argument("--out-csv", default=os.path.join(DATA, "allofus_items_umap_models.csv"))
    p.add_argument("--n-neighbors", type=int, default=15)
    p.add_argument("--min-dist", type=float, default=0.10)
    p.add_argument("--metric", default="cosine")
    p.add_argument("--random-state", type=int, default=42)
    args = p.parse_args(argv)

    inv = pd.read_csv(args.inventory).set_index("item")
    survey_of = inv["survey"].to_dict()

    import umap
    frames = []
    for label, fname in MODELS:
        emb = load_item_embeddings(os.path.join(DATA, fname))
        items = list(emb.items)
        coords = umap.UMAP(
            n_neighbors=args.n_neighbors, min_dist=args.min_dist,
            metric=args.metric, random_state=args.random_state,
        ).fit_transform(emb.matrix_for(items))
        frames.append(pd.DataFrame({
            "model": label, "item": items,
            "survey": [survey_of[i] for i in items],
            "umap_x": coords[:, 0], "umap_y": coords[:, 1],
        }))
        print("UMAP done:", label, flush=True)
    allf = pd.concat(frames, ignore_index=True)
    allf.to_csv(args.out_csv, index=False)

    _plot(allf, args.out_png, args.metric, args.n_neighbors, args.min_dist)
    print("Wrote {} and {}".format(args.out_png, args.out_csv))
    return 0


def _plot(allf, out_png, metric, n_neighbors, min_dist):
    import matplotlib
    matplotlib.use("Agg")
    import matplotlib.pyplot as plt

    order = allf[allf["model"] == MODELS[0][0]]["survey"].value_counts().index.tolist()
    cmap = plt.get_cmap("tab10")
    colors = {s: cmap(i % 10) for i, s in enumerate(order)}

    fig, axes = plt.subplots(2, 3, figsize=(16, 10))
    for ax, (label, _) in zip(axes.ravel(), MODELS):
        sub = allf[allf["model"] == label]
        for s in order:
            g = sub[sub["survey"] == s]
            ax.scatter(g["umap_x"], g["umap_y"], s=14, alpha=0.85,
                       color=colors[s], edgecolor="none",
                       label=s if label == MODELS[0][0] else None)
        ax.set_title(label, fontsize=11)
        ax.set_xticks([]); ax.set_yticks([])
        for side in ("top", "right"):
            ax.spines[side].set_visible(False)
    handles = [plt.Line2D([], [], marker="o", linestyle="", color=colors[s],
                          label="{} ({})".format(s, (allf["survey"] == s).sum() // len(MODELS)))
               for s in order]
    fig.legend(handles=handles, title="Survey form", loc="lower center",
               ncol=4, fontsize=9, frameon=False, bbox_to_anchor=(0.5, -0.02))
    fig.suptitle("All of Us item prompts (282) — 2D UMAP per embedding model, colored by survey form\n"
                 "metric={} · n_neighbors={} · min_dist={} · seed=42".format(
                     metric, n_neighbors, min_dist), fontsize=12)
    fig.tight_layout(rect=(0, 0.03, 1, 1))
    fig.savefig(out_png, dpi=150, bbox_inches="tight")
    plt.close(fig)


if __name__ == "__main__":
    raise SystemExit(main())
```
