## Supplementary material for "Auditable cross-instrument detection of unusual multivariate psychiatric response configurations using a semantically aligned covariance subspace": Code for tool: ross_operating_point.py

```
#!/usr/bin/env python3
"""Find the ROSS/CPP operating point on the package's intended range (0, 10 deg].

Replicates the package's Cosine-Preserving Pruning sweep (cpp.py default
theta-max=10, 100 steps) and its two operating-point selectors
(select_elbow, select_dtheta_ds from extract_pruned_W.py) on the bge-m3
sparse fit, to locate the knee. Pure numpy/scipy (main conda env).
"""
import argparse
import json
import os

import numpy as np
from scipy.signal import savgol_filter


def cpp_prune_at_theta(W, theta_deg):
    cos_t = np.cos(np.radians(theta_deg))
    out = W.copy()
    for j in range(W.shape[0]):
        e_tot = np.sum(W[j] ** 2)
        if e_tot == 0:
            continue
        order = np.argsort(np.abs(W[j]))
        e_rem = e_tot - np.cumsum(W[j, order] ** 2)
        can_zero = np.searchsorted(-e_rem, -cos_t ** 2 * e_tot)
        out[j, order[:can_zero]] = 0.0
    return out


def recon_metrics(Xc, W_kp):
    """Package convention: loadings (p,k)=W_kp.T; recon = (Xc@L)@L.T."""
    L = W_kp.T
    recon = (Xc @ L) @ L.T
    rel = float(np.linalg.norm(Xc - recon) / np.linalg.norm(Xc))
    return rel, 1.0 - rel ** 2


def select_elbow(pareto):
    if len(pareto) <= 2:
        return 0
    s = np.array([p["sparsity"] for p in pareto])
    r = np.array([p["relative_recon_loss"] for p in pareto])
    if s.max() - s.min() < 1e-12 or r.max() - r.min() < 1e-12:
        return 0
    sn = (s - s.min()) / (s.max() - s.min())
    rn = (r - r.min()) / (r.max() - r.min())
    x0, y0, x1, y1 = sn[0], rn[0], sn[-1], rn[-1]
    dx, dy = x1 - x0, y1 - y0
    L = np.hypot(dx, dy)
    dist = (dy * sn - dx * rn + x1 * y0 - y1 * x0) / L
    return int(np.argmax(dist))


def select_dtheta_ds(pareto):
    if len(pareto) < 7:
        return select_elbow(pareto)
    s = np.array([p["sparsity"] for p in pareto])
    ve = np.array([p["var_explained"] for p in pareto])
    sn = (s - s.min()) / (s.max() - s.min() + 1e-15)
    vn = (ve - ve.min()) / (ve.max() - ve.min() + 1e-15)
    win = min(len(sn) // 3, 21)
    if win % 2 == 0:
        win -= 1
    win = max(win, 5)
    vs = savgol_filter(vn, window_length=win, polyorder=3)
    d1 = np.gradient(vs, sn)
    d2 = np.gradient(d1, sn)
    curv = d2 / (1.0 + d1 ** 2)
    m = max(2, len(curv) // 20)
    curv[:m] = 0
    curv[-m:] = 0
    return int(np.argmin(curv))


def main():
    ap = argparse.ArgumentParser()
    ap.add_argument("--embeddings", default="data/allofus/allofus_items.npz")
    ap.add_argument("--ross-dir", default="data/allofus/ross_out/bge-m3_ls0.1_lo0.1")
    ap.add_argument("--theta-max", type=float, default=10.0)
    ap.add_argument("--theta-steps", type=int, default=100)
    args = ap.parse_args()

    data = np.load(args.embeddings, allow_pickle=True)
    items = [str(x) for x in data["items"]]
    Q = np.asarray(data["vectors"], dtype=np.float64)
    Xc = Q.T - Q.T.mean(axis=0, keepdims=True)            # orientation B
    W = np.load(os.path.join(args.ross_dir, "loadings.npy"))   # (k, p)

    thetas = np.linspace(0, args.theta_max, args.theta_steps + 1)
    pareto = []
    for th in thetas:
        Wp = W.copy() if th == 0 else cpp_prune_at_theta(W, th)
        rel, ve = recon_metrics(Xc, Wp)
        spar = float((Wp == 0).mean())
        nnz = float(np.median((np.abs(Wp) > 1e-12).sum(axis=1)))
        pareto.append({"theta_deg": float(th), "sparsity": spar,
                       "relative_recon_loss": rel, "var_explained": ve, "median_nnz": nnz})

    i_elbow = select_elbow(pareto)
    i_curv = select_dtheta_ds(pareto)
    json.dump({"pareto": pareto,
               "elbow": pareto[i_elbow], "dtheta_ds": pareto[i_curv]},
              open(os.path.join(args.ross_dir, "operating_point.json"), "w"), indent=2)

    print("theta  sparsity  med_nnz  var_expl   (range 0-10 deg)")
    for p in pareto[::10]:
        print("  {:4.1f}   {:6.1%}   {:5.0f}    {:.3f}".format(
            p["theta_deg"], p["sparsity"], p["median_nnz"], p["var_explained"]))
    for name, i in [("select_elbow", i_elbow), ("select_dtheta_ds", i_curv)]:
        p = pareto[i]
        print(">>> {:16s}: theta={:.2f}deg  sparsity={:.1%}  med_nnz={:.0f}  var_expl={:.3f}".format(
            name, p["theta_deg"], p["sparsity"], p["median_nnz"], p["var_explained"]))

    _plot(pareto, i_elbow, i_curv, os.path.join(args.ross_dir, "operating_point.png"))


def _plot(pareto, i_elbow, i_curv, out):
    import matplotlib
    matplotlib.use("Agg")
    import matplotlib.pyplot as plt
    s = [p["sparsity"] for p in pareto]
    ve = [p["var_explained"] for p in pareto]
    th = [p["theta_deg"] for p in pareto]
    fig, ax = plt.subplots(figsize=(8, 6))
    sc = ax.scatter(s, ve, c=th, cmap="viridis", s=20)
    ax.plot(s, ve, color="gray", lw=0.6, alpha=0.6)
    for i, name, col in [(i_elbow, "elbow", "red"), (i_curv, "dθ/ds knee", "black")]:
        ax.scatter([s[i]], [ve[i]], s=140, facecolors="none", edgecolors=col, linewidths=2,
                   label="{} (θ={:.1f}°, {:.0%} zeros)".format(name, th[i], s[i]))
    fig.colorbar(sc, label="θ (degrees)")
    ax.set_xlabel("Sparsity (zero fraction)"); ax.set_ylabel("Variance explained")
    ax.set_title("ROSS bge-m3: CPP sparsity–fidelity Pareto (θ = {:.0f}–{:.0f}°)".format(
        min(th), max(th)))
    ax.legend(loc="lower left"); ax.spines[["top", "right"]].set_visible(False)
    fig.tight_layout(); fig.savefig(out, dpi=150); plt.close(fig)
    print("wrote", out)


if __name__ == "__main__":
    main()
```
