## Supplementary material for "Auditable cross-instrument detection of unusual multivariate psychiatric response configurations using a semantically aligned covariance subspace": Code for tool: run_ross_spca.py

```
#!/usr/bin/env python3
"""Run ROSS SmoothSparsePCA on an All of Us item-embedding matrix (orientation B).

Run with the ROSS side-venv (jax/optax): env_ross/bin/python.

Orientation B — items are the FEATURES that get sparsified (mirrors the HLCA
setup where genes are features). The input embedding artifact is (p items x m
dims); we transpose to X = (m dims x p items) so ROSS's per-feature sparsity
lands on items. The resulting sparse loadings (k x p) are the sparse analog of
the manuscript's item-coordinate matrix — each component defined by few items.

ROSS centers X over axis 0 (per item, across the m dims) — the exact analog of
HLCA's per-gene centering. This differs from the manuscript's bias-vector
(per-dim, over items) centering; orientation B is a deliberate sparse variant.

Outputs to --out-dir: loadings.npy (k x p), scores.npy (m x k),
top_items.csv, sparsity.json.
"""

import argparse
import json
import os
import sys

import numpy as np

ROSS_LIB = os.path.join(os.path.dirname(os.path.abspath(__file__)),
                        "..", "..", "..", "ross_pca_pkg", "ROSS_PCA_Codes", "lib")


def participation_ratio(v):
    s2 = v ** 2
    tot = s2.sum()
    return float((tot ** 2) / (s2 ** 2).sum()) if tot > 0 else 0.0


def main():
    ap = argparse.ArgumentParser(description="ROSS SmoothSparsePCA, orientation B (items as features)")
    ap.add_argument("--embeddings", required=True, help="items-embedding .npz (keys: items, vectors)")
    ap.add_argument("--out-dir", required=True)
    ap.add_argument("--n-components", type=int, required=True)
    ap.add_argument("--lambda-sparsity", type=float, default=0.1)
    ap.add_argument("--lambda-ortho", type=float, default=0.1)
    ap.add_argument("--init", default="pca", choices=["pca", "random_normalized"])
    ap.add_argument("--seed", type=int, default=0)
    ap.add_argument("--total-steps", type=int, default=5000)
    ap.add_argument("--top-n", type=int, default=12)
    ap.add_argument("--ross-lib", default=ROSS_LIB)
    args = ap.parse_args()

    sys.path.insert(0, os.path.abspath(args.ross_lib))
    from sparse_PCA_v10 import SmoothSparsePCA

    data = np.load(args.embeddings, allow_pickle=True)
    items = [str(x) for x in data["items"]]
    Q = np.asarray(data["vectors"], dtype=np.float64)        # (p items, m dims)
    X = np.ascontiguousarray(Q.T)                            # (m dims, p items): items are features
    p = len(items)
    print("X (n_samples=dims, n_features=items):", X.shape, "| k =", args.n_components)

    model = SmoothSparsePCA(
        n_components=args.n_components,
        init=args.init,
        lambda_sparsity=args.lambda_sparsity,
        lambda_ortho=args.lambda_ortho,
        total_steps=args.total_steps,
        random_state=args.seed,
        verbose=False,
    )
    model.fit(X)
    loadings = np.asarray(model.components_, dtype=np.float64)   # (k, p items)
    Xc = X - X.mean(axis=0, keepdims=True)
    scores = Xc @ loadings.T                                     # (m dims, k)

    os.makedirs(args.out_dir, exist_ok=True)
    np.save(os.path.join(args.out_dir, "loadings.npy"), loadings)
    np.save(os.path.join(args.out_dir, "scores.npy"), scores)

    # sparsity metrics (per component, over the p items)
    nnz = (np.abs(loadings) > 1e-8).sum(axis=1)
    pr = np.array([participation_ratio(loadings[k]) for k in range(loadings.shape[0])])
    overall_sparsity = float((np.abs(loadings) <= 1e-8).mean())
    sparsity = {
        "n_components": int(loadings.shape[0]), "n_items": p,
        "overall_zero_fraction": overall_sparsity,
        "nnz_per_component_median": float(np.median(nnz)),
        "nnz_per_component_min": int(nnz.min()), "nnz_per_component_max": int(nnz.max()),
        "participation_ratio_median": float(np.median(pr)),
        "lambda_sparsity": args.lambda_sparsity, "lambda_ortho": args.lambda_ortho,
    }
    with open(os.path.join(args.out_dir, "sparsity.json"), "w") as f:
        json.dump(sparsity, f, indent=2)

    # top items per component
    rows = []
    for k in range(loadings.shape[0]):
        idx = np.argsort(-np.abs(loadings[k]))[:args.top_n]
        idx = [i for i in idx if abs(loadings[k][i]) > 1e-8]
        rows.append("PC{},{},{}".format(
            k + 1, int(nnz[k]),
            "; ".join("{}({:+.2f})".format(items[i], loadings[k][i]) for i in idx)))
    with open(os.path.join(args.out_dir, "top_items.csv"), "w") as f:
        f.write("component,nnz_items,top_items\n" + "\n".join(rows) + "\n")

    print("nnz/comp: median {:.0f} (min {}, max {}) of {} items | overall zeros {:.1%}".format(
        sparsity["nnz_per_component_median"], sparsity["nnz_per_component_min"],
        sparsity["nnz_per_component_max"], p, overall_sparsity))
    print("wrote ->", args.out_dir)


if __name__ == "__main__":
    main()
```
