## Supplementary material for "Auditable cross-instrument detection of unusual multivariate psychiatric response configurations using a semantically aligned covariance subspace": Code for tool: convert_nhis.py

```
#!/usr/bin/env python3
"""
NHIS -> survey-semantics converter
==================================

Turns a raw NHIS adult CSV into the four files the *general* survey-semantics
pipeline consumes, one set per year:

    <outdir>/<year>/nhis<year>.csv          responses (id + age + sex + items, raw)
    <outdir>/<year>/nhis<year>_prompts.csv  item,prompt
    <outdir>/<year>/nhis<year>_scales.csv   item,min,max,sentinels,reverse
    <outdir>/<year>/nhis<year>_weights.csv  weight   (row-aligned to the responses)

Design intent: ALL NHIS-specific knowledge lives here (which columns are items,
their scales/sentinels/reverse flags, the survey-weight column, covariate names).
The pipeline stays general -- it just reads the generic prompt/scale/weight files
produced here. The item definitions are sourced from the replication script's
ALL_QUESTION_TEXTS / ALL_ITEM_SCALES dicts (parsed, not executed) so they remain
the single source of truth.

Responses are written RAW: sentinel codes are NOT stripped here. The pipeline
removes them per item using the scales file, which keeps the response file
auditable against the original NHIS extract.

Usage:
    python convert_nhis.py
    python convert_nhis.py --data2021 2021/adult21.csv --data2024 2024/adult24.csv
    python convert_nhis.py --all-items     # keep every pipeline item per year
"""

import argparse
import ast
import csv
import os
import sys

SCRIPT_DIR = os.path.dirname(os.path.abspath(__file__))

# NHIS-specific column roles (kept here, not in the pipeline).
ID_COL = "HHX"            # one sample adult per household -> unique per row
AGE_SRC = "AGEP_A"        # -> "age"   (tool auto-detects this covariate name)
SEX_SRC = "SEX_A"         # -> "sex"
WEIGHT_SRC = "WTFA_A"     # final adult sample weight

# csv field-size: NHIS rows are wide but values are short; default limit is fine.


def load_item_defs(script_path):
    """Extract ALL_QUESTION_TEXTS and ALL_ITEM_SCALES via AST (no execution)."""
    with open(script_path, "r", encoding="utf-8") as handle:
        tree = ast.parse(handle.read())
    found = {}
    for node in tree.body:
        if isinstance(node, ast.Assign) and len(node.targets) == 1 \
                and isinstance(node.targets[0], ast.Name):
            name = node.targets[0].id
            if name in ("ALL_QUESTION_TEXTS", "ALL_ITEM_SCALES"):
                found[name] = ast.literal_eval(node.value)
    missing = {"ALL_QUESTION_TEXTS", "ALL_ITEM_SCALES"} - set(found)
    if missing:
        raise SystemExit("Could not find {} in {}".format(missing, script_path))
    texts, scales = found["ALL_QUESTION_TEXTS"], found["ALL_ITEM_SCALES"]
    if set(texts) != set(scales):
        raise SystemExit("ALL_QUESTION_TEXTS and ALL_ITEM_SCALES keys differ.")
    return texts, scales


def read_header(path):
    with open(path, "r", newline="", encoding="utf-8-sig") as handle:
        return next(csv.reader(handle))


def common_items(item_order, header21, header24):
    h21, h24 = set(header21), set(header24)
    return [it for it in item_order if it in h21 and it in h24]


def items_in(item_order, header):
    present = set(header)
    return [it for it in item_order if it in present]


def write_prompts(path, items, texts):
    with open(path, "w", newline="", encoding="utf-8") as handle:
        writer = csv.writer(handle)
        writer.writerow(["item", "prompt"])
        for it in items:
            writer.writerow([it, texts[it]])


def write_scales(path, items, scales):
    # Emit an optional `ceiling` column only when the item defs declare it (i.e.
    # ALL_ITEM_SCALES carries a "ceiling" key). Absent, the pan-mild audit falls
    # back to all polytomous items; present, it is an explicit allowlist of the
    # items where "maxed out" is a genuine symptom ceiling.
    has_ceiling = any("ceiling" in scales[it] for it in items)
    header = ["item", "min", "max", "sentinels", "reverse"]
    if has_ceiling:
        header.append("ceiling")
    with open(path, "w", newline="", encoding="utf-8") as handle:
        writer = csv.writer(handle)
        writer.writerow(header)
        for it in items:
            s = scales[it]
            sentinels = ";".join(str(int(x)) for x in sorted(s["sentinels"]))
            row = [
                it, int(s["min"]), int(s["max"]),
                sentinels, "true" if s["reverse"] else "false",
            ]
            if has_ceiling:
                row.append("true" if s.get("ceiling") else "false")
            writer.writerow(row)


def convert_year(year, data_path, out_dir, items, texts, scales):
    """Stream the raw CSV once, emitting the response and weight files together."""
    header = read_header(data_path)
    col_idx = {name: i for i, name in enumerate(header)}

    for required in (ID_COL, AGE_SRC, SEX_SRC, WEIGHT_SRC):
        if required not in col_idx:
            raise SystemExit(
                "{}: required column {!r} not found.".format(data_path, required)
            )
    year_items = [it for it in items if it in col_idx]

    os.makedirs(out_dir, exist_ok=True)
    data_path_out = os.path.join(out_dir, "nhis{}.csv".format(year))
    weights_path = os.path.join(out_dir, "nhis{}_weights.csv".format(year))
    prompts_path = os.path.join(out_dir, "nhis{}_prompts.csv".format(year))
    scales_path = os.path.join(out_dir, "nhis{}_scales.csv".format(year))

    out_header = [ID_COL, "age", "sex"] + year_items
    item_positions = [col_idx[it] for it in year_items]
    id_i, age_i, sex_i, w_i = (
        col_idx[ID_COL], col_idx[AGE_SRC], col_idx[SEX_SRC], col_idx[WEIGHT_SRC],
    )

    n_rows = 0
    with open(data_path, "r", newline="", encoding="utf-8-sig") as src, \
            open(data_path_out, "w", newline="", encoding="utf-8") as data_out, \
            open(weights_path, "w", newline="", encoding="utf-8") as w_out:
        reader = csv.reader(src)
        data_writer = csv.writer(data_out)
        w_writer = csv.writer(w_out)
        next(reader)  # skip header
        data_writer.writerow(out_header)
        w_writer.writerow(["weight"])
        for row in reader:
            data_writer.writerow(
                [row[id_i], row[age_i], row[sex_i]] + [row[p] for p in item_positions]
            )
            w_writer.writerow([row[w_i]])
            n_rows += 1

    write_prompts(prompts_path, year_items, texts)
    write_scales(scales_path, year_items, scales)

    print("[{}] {} subjects, {} items".format(year, n_rows, len(year_items)))
    print("       responses: {}".format(data_path_out))
    print("       weights:   {} ({} rows + header)".format(weights_path, n_rows))
    print("       prompts:   {}".format(prompts_path))
    print("       scales:    {}".format(scales_path))
    return n_rows, year_items


def main(argv=None):
    parser = argparse.ArgumentParser(description="NHIS -> survey-semantics converter")
    parser.add_argument("--data2021", default=os.path.join(SCRIPT_DIR, "2021", "adult21.csv"))
    parser.add_argument("--data2024", default=os.path.join(SCRIPT_DIR, "2024", "adult24.csv"))
    parser.add_argument("--outdir", default=SCRIPT_DIR,
                        help="Per-year folders <outdir>/2021 and <outdir>/2024.")
    parser.add_argument("--script", default=os.path.join(SCRIPT_DIR, "nhis_replication_2021_2024.py"),
                        help="Source of ALL_QUESTION_TEXTS / ALL_ITEM_SCALES.")
    parser.add_argument("--all-items", action="store_true",
                        help="Keep every pipeline item present per year instead of "
                             "the common-across-years intersection (default).")
    args = parser.parse_args(argv)

    texts, scales = load_item_defs(args.script)
    item_order = list(texts.keys())
    h21, h24 = read_header(args.data2021), read_header(args.data2024)

    if args.all_items:
        items21 = items_in(item_order, h21)
        items24 = items_in(item_order, h24)
        print("Mode: all available items per year (bases may differ).")
    else:
        shared = common_items(item_order, h21, h24)
        items21 = items24 = shared
        dropped = [it for it in item_order if it not in shared]
        print("Mode: common items only -> {} shared items.".format(len(shared)))
        if dropped:
            print("Dropped (not in both years): {}".format(", ".join(dropped)))

    # Identical-wording guarantee: both years draw text from one dict, so any
    # shared item is byte-identical by construction. (Assert defensively.)
    for it in set(items21) & set(items24):
        assert texts[it] == texts[it]  # single source -> always true; documents intent

    convert_year("2021", args.data2021, os.path.join(args.outdir, "2021"),
                 items21, texts, scales)
    convert_year("2024", args.data2024, os.path.join(args.outdir, "2024"),
                 items24, texts, scales)
    print("\nDone.")
    return 0


if __name__ == "__main__":
    sys.exit(main())
```
