## Supplementary material for "Auditable cross-instrument detection of unusual multivariate psychiatric response configurations using a semantically aligned covariance subspace": Code for tool: README.md

# Worked Example — NHIS 2021 & 2024

This example runs the **NHIS adult cohorts** (National Health Interview Survey,
2021 and 2024) through the general `survey-semantics` pipeline: it builds a
shared semantic space from the survey item wording, projects each respondent's
answers into it, removes age/sex signal with **survey-weighted** regression, and
ranks respondents by how unusual they are in that space.

It demonstrates every general tool feature in one place:
- a **prompt file** (item wording),
- a **scale file** (per-item ranges, missing-value codes, reverse coding),
- a **weights file** (NHIS survey weights → weighted statistics),
- the **common-items** design that makes the two years directly comparable.

> **Relation to the paper.** The published method (Periwal, *"Auditable
> cross-instrument detection…"*) was validated on the **HRS** and **Xinxiang**
> cohorts. NHIS here is an **extension**: it is the first application to use the
> tool's **survey-weighting** path (weighted residualization + weighted
> Mahalanobis), which is beyond the original two-cohort study. The paper's
> downstream **"pan-mild" / ceiling-exclusion** step is *not* run by the tool (see
> §6).

> The pipeline itself is **not** NHIS-aware. All NHIS-specific knowledge lives in
> [`convert_nhis.py`](convert_nhis.py); the tool only reads the generic
> prompt/scale/weight files it emits. See
> [`docs/pipeline_overview.md`](../../docs/pipeline_overview.md) for the general
> pipeline.

---

## 1. What you provide

NHIS microdata and the replication script are **not committed** (the `data/`
directory is gitignored). You supply:

| File | What it is | Where to get it |
|---|---|---|
| `adult21.csv`, `adult24.csv` | Raw NHIS Sample Adult public-use CSVs | [CDC NHIS data releases](https://www.cdc.gov/nhis/data-questionnaires-documentation.htm) |
| `nhis_replication_2021_2024.py` | The replication script holding `ALL_QUESTION_TEXTS` and `ALL_ITEM_SCALES` | Your study repo (the single source of truth for item definitions) |

Put them under a gitignored working dir, e.g.:

```text
data/NHIS/
  nhis_replication_2021_2024.py
  2021/adult21.csv
  2024/adult24.csv
```

---

## 2. Convert raw extracts → tool-ready files

```bash
python examples/nhis/convert_nhis.py \
  --script   data/NHIS/nhis_replication_2021_2024.py \
  --data2021 data/NHIS/2021/adult21.csv \
  --data2024 data/NHIS/2024/adult24.csv \
  --outdir   data/NHIS
```

For each year this writes four generic files into `data/NHIS/<year>/`:

| File | Columns | Role |
|---|---|---|
| `nhis<year>.csv` | `HHX, age, sex, <items…>` | Responses, **raw** (sentinels left in for auditability; the tool cleans them) |
| `nhis<year>_prompts.csv` | `item, prompt` | The question wording — raw material for the semantic space |
| `nhis<year>_scales.csv` | `item, min, max, sentinels, reverse` (optional `ceiling`) | Per-item range, missing codes, reverse flag, and optionally which items count for the pan-mild ceiling audit |
| `nhis<year>_weights.csv` | `weight` | One NHIS survey weight (`WTFA_A`) per row, aligned to `nhis<year>.csv` |

**Common-items design (default).** The converter keeps only items present in
*both* years with identical wording (38 of 47). This is what makes the two years
comparable: the semantic basis **W** is fit on the item *prompts*, so identical
prompts ⇒ an identical basis in each year — no basis needs to be saved or frozen.
Pass `--all-items` to keep every item per year instead (bases may then differ).

Why these four files and not hard-coded NHIS logic: each is a generic tool input,
so any other survey can be run the same way by writing its own converter. See
[the converter's header](convert_nhis.py) for the NHIS column roles
(`HHX`/`AGEP_A`/`SEX_A`/`WTFA_A`).

---

## 3. Analyze each year

The convenience script converts, then runs the three modular stages — **embed the
common prompts once** (the only LLM step) and **build the semantic basis once**
(PCA, no responses), then analyzes both years from that one shared basis — so the
years provably share one semantic space and the per-year analyses need no model:

```bash
# Requires a local bge-m3 (sentence-transformers) model; there is no fallback.
MODEL=/path/to/bge-m3 examples/nhis/run_nhis.sh
```

Equivalently, by hand — embed once, build the basis once, then score each year
with `--basis-file`:

```bash
python -m survey_semantics.cli embed \
  --prompt-file data/NHIS/2021/nhis2021_prompts.csv \
  --model /path/to/bge-m3 --out outputs/nhis/nhis_items.npz

python -m survey_semantics.cli pca \
  --embeddings-file outputs/nhis/nhis_items.npz \
  --out outputs/nhis/nhis_basis.npz

python -m survey_semantics.cli analyze-file data/NHIS/2021/nhis2021.csv \
  --prompt-file   data/NHIS/2021/nhis2021_prompts.csv \
  --scale-file    data/NHIS/2021/nhis2021_scales.csv \
  --weights-file  data/NHIS/2021/nhis2021_weights.csv \
  --basis-file    outputs/nhis/nhis_basis.npz \
  --id-col HHX \
  --d-selection variance --variance-threshold 0.80 \
  --skip-umap \
  --outdir outputs/nhis/2021
# → Analyzed nhis2021: 29372 rows, 38 items, D=14 (bge-m3).
```

What each NHIS-specific flag does:
- `--id-col HHX` — NHIS has no `subjectkey`; the household id is unique per sample adult.
- `--scale-file …_scales.csv` — **declares the 38 analyzed items**, cleans each
  item's own missing codes (e.g. `7;8;9` is "refused" for `SAD_A` but `0;97;98;99`
  for `LASTDR_A`), applies declared ranges, and reverse-scores flagged items.
  Without it, item inference drops valid items whose `7/8/9` codes inflate their
  unique-value count.
- `--weights-file …_weights.csv` — turns on **WLS residualization** and
  **weighted Mahalanobis** so age/sex adjustment and the outlier metric respect
  NHIS survey weights. (`age`/`sex` are auto-detected covariates.)

> Use `--scale-file` (single file), **not** `--scale-dir`: the year folder also
> holds the raw `adultNN.csv`, which a directory loader would try to parse.

---

## 4. Read the results

Per year, under `outputs/nhis/<year>/` (filenames prefixed by
`nhis<year>__<embedding-slug>`):

| File | Use |
|---|---|
| `…_scores.csv` | **The ranking** — per-respondent Mahalanobis distance (`Mahalanobis_Dist`), semantic PC scores, id, covariates. Larger = more semantically unusual. |
| `…_prompt_loadings.csv` | How each item loads on each semantic dimension — interpret what the axes *mean*. |
| `…_drivers.csv` | For top outliers, which dimensions/items drive them. |
| `…_stability.csv`, `…_dimension_selection.csv` | How the dimensionality `D` was chosen and how stable the outlier set is. |
| `case_studies/` | Per-outlier narrative reports. |
| `summary.csv` | One-row run summary (n_rows, n_items, D, embedding). |

---

## 5. Cross-year verification (recommended)

Because the basis is fit on identical prompts, the two years should share it:

- **Shared-basis check** — assert the 2021 and 2024 prompt loadings describe the
  same space (`W_2021 == W_2024` up to PCA sign flips). This is the proof that
  cross-cohort comparison is valid.
- **Fidelity spot-check** — compare a few respondents' weighted Mahalanobis
  distances against the replication script's `process_year` output on the same
  rows. The pipeline's weighted math is matched to that script to `1e-9`.

---

## 6. Notes & gotchas

- **Weights are positional.** The weights file must match the response file's row
  order and length; the tool hard-errors on a length mismatch. The converter
  writes both from the same pass, guaranteeing alignment.
- **PCA sign is arbitrary.** Treat `|z|` / distance as the stable severity; signed
  PC scores are only meaningful within one run's orientation.
- **Pan-mild is an opt-in audit (`--pan-mild`).** The paper isolates *below-threshold*
  unusual cases by excluding any flagged respondent who maxed out a Likert item.
  Add `--pan-mild` to emit an `At_Ceiling` column and, per empirical percentile, an
  `Is_Pan_Mild_Emp<pct>` column (outlier **and** nowhere at the ceiling).
  **Which items count** is set by an optional `ceiling` column in the scale file —
  an explicit allowlist, important for NHIS where non-symptom items (e.g.
  `LASTDR_A`, time since last doctor visit) should *not* be treated as a symptom
  ceiling. Without that column it falls back to all polytomous items (≥
  `--ceiling-min-levels`, default 3); binary items are excluded. The paper's
  *instrument-sum* ceiling variant for binary scales (e.g. `sum(CES-D) < 4`) is
  study-specific and still left to a downstream script.
