## Supplementary material for "Auditable cross-instrument detection of unusual multivariate psychiatric response configurations using a semantically aligned covariance subspace": Code for tool: run_nhis.sh

```
#!/usr/bin/env bash
#
# End-to-end NHIS 2021 + 2024 run: convert raw extracts, then analyze each year
# through the general survey-semantics pipeline (scales + survey weights).
#
# Prerequisites (NOT committed — NHIS data and the replication script are yours):
#   - The replication script with ALL_QUESTION_TEXTS / ALL_ITEM_SCALES.
#   - Raw adult CSVs: adult21.csv (2021) and adult24.csv (2024).
#   - A LOCAL bge-m3 (sentence-transformers) model directory.
#
# Override any path via environment variables; defaults assume a gitignored
# ./data/NHIS working dir at the repo root.
#
# Usage:
#   MODEL=/path/to/bge-m3 examples/nhis/run_nhis.sh
set -euo pipefail

HERE="$(cd "$(dirname "${BASH_SOURCE[0]}")" && pwd)"
REPO="$(cd "$HERE/../.." && pwd)"

DATA_DIR="${DATA_DIR:-$REPO/data/NHIS}"
SCRIPT="${SCRIPT:-$DATA_DIR/nhis_replication_2021_2024.py}"
ADULT21="${ADULT21:-$DATA_DIR/2021/adult21.csv}"
ADULT24="${ADULT24:-$DATA_DIR/2024/adult24.csv}"
OUTDIR="${OUTDIR:-$REPO/outputs/nhis}"

# The only embedding backend is a local sentence-transformers model (e.g. bge-m3).
# There is NO TF-IDF/offline fallback — the model must already be on local disk.
MODEL="${MODEL:-}"
if [[ -z "$MODEL" ]]; then
  echo "ERROR: set MODEL=/path/to/bge-m3 (a local sentence-transformers model)." >&2
  echo "       There is no TF-IDF or auto fallback; the model must be on local disk." >&2
  exit 1
fi
echo "==> 1/4  Convert raw NHIS extracts -> tool-ready per-year files"
python "$HERE/convert_nhis.py" \
  --script   "$SCRIPT" \
  --data2021 "$ADULT21" \
  --data2024 "$ADULT24" \
  --outdir   "$DATA_DIR"

# Stages 1 (embed) and 2 (pca) are done ONCE and shared by both years. Both years
# use the same common items with identical wording, so one embeddings file and one
# basis serve both — which *guarantees* they share the same semantic space. The
# per-year analyses below need no model.
EMB="$OUTDIR/nhis_items.npz"
BASIS="$OUTDIR/nhis_basis.npz"
mkdir -p "$OUTDIR"
echo "==> 2/4  Embed common item prompts once (LLM step)"
python -m survey_semantics.cli embed \
  --prompt-file "$DATA_DIR/2021/nhis2021_prompts.csv" \
  --model "$MODEL" --out "$EMB"

echo "==> 3/4  Build the shared semantic basis once (PCA, no responses)"
python -m survey_semantics.cli pca \
  --embeddings-file "$EMB" --out "$BASIS"

for YEAR in 2021 2024; do
  echo "==> 4/4  Analyze NHIS $YEAR (no model — from the shared basis)"
  python -m survey_semantics.cli analyze-file "$DATA_DIR/$YEAR/nhis$YEAR.csv" \
    --prompt-file   "$DATA_DIR/$YEAR/nhis${YEAR}_prompts.csv" \
    --scale-file    "$DATA_DIR/$YEAR/nhis${YEAR}_scales.csv" \
    --weights-file  "$DATA_DIR/$YEAR/nhis${YEAR}_weights.csv" \
    --basis-file    "$BASIS" \
    --id-col HHX \
    --d-selection variance --variance-threshold 0.80 \
    --pan-mild --empirical-percentiles 95 99 \
    --skip-umap \
    --outdir "$OUTDIR/$YEAR"
done

echo "Done. Outputs under: $OUTDIR/{2021,2024}"
echo "  Ranked outliers:  $OUTDIR/<year>/nhis<year>__*_scores.csv"
```
