## Supplementary material for "Auditable cross-instrument detection of unusual multivariate psychiatric response configurations using a semantically aligned covariance subspace": Code for tool: pyproject.toml

```
[build-system]
requires = ["setuptools>=61"]
build-backend = "setuptools.build_meta"

[project]
name = "survey-semantics"
version = "0.1.0"
description = "Semantic manifold analysis tools for survey item response data."
readme = "README.md"
requires-python = ">=3.7"
dependencies = [
  "numpy>=1.17,<1.22",
  "pandas>=1.0,<1.4",
  "scikit-learn>=0.22,<1.1",
  "scipy>=1.3.1,<1.8",
  "matplotlib>=3.5,<3.6",
]

[project.optional-dependencies]
sentence-transformers = [
  "sentence-transformers>=2.2,<6",
  "transformers>=4.20,<5",
  "huggingface-hub>=0.10,<1",
]
umap = ["umap-learn==0.4.6"]
dev = ["pytest"]

[project.scripts]
survey-semantics = "survey_semantics.cli:main"
survey-semantics-plot = "survey_semantics.plotting:main"

[tool.setuptools.packages.find]
where = ["src"]
```
