## Supplementary material for "Auditable cross-instrument detection of unusual multivariate psychiatric response configurations using a semantically aligned covariance subspace": Code for tool: README.md

# ACIDD: Auditable Cross-Instrument Distress Detection

Reusable implementation of **Periwal, *"Auditable cross-instrument detection of
unusual multivariate response configurations using a semantically aligned
covariance subspace."*** It embeds questionnaire **item wording** into a shared
semantic space, projects responses into it, and flags respondents whose
cross-instrument response pattern is multivariate-unusual — auditably, by tracing
each flag back to the original items.

> Validated in the paper on the **HRS** and **Xinxiang** cohorts. This package
> generalizes the method to any subjects×items survey; the worked example applies
> it to **NHIS 2021/2024** and adds survey weighting.

**Full docs:** [docs/pipeline_overview.md](docs/pipeline_overview.md) — how it
works, file formats, every option. · [examples/nhis](examples/nhis/README.md) —
end-to-end worked example.

## Install

```bash
conda env create -f environment-bge-m3.yml
conda activate survey-semantics-bge-m3
pip install -e . --no-deps     # --no-deps keeps the env's modern stack
python -m pytest -q            # verify
```

## Embedding model (required)

The only backend is a local `sentence-transformers` model (e.g. bge-m3) — **no
fallback**, a missing model raises. It is offline-only, so fetch it once:

```bash
python -c "from huggingface_hub import snapshot_download; \
snapshot_download(repo_id='BAAI/bge-m3', local_dir='models/bge-m3', \
ignore_patterns=['onnx/**','openvino/**','*.onnx'])"
```

## How it runs — three modular stages

You give the tool the **wording of each question** and a table of **responses**;
it returns a ranked list of people whose answers form an unusually different
overall pattern. The work splits into three stages, and **each stage needs only
its own inputs** — so the expensive model step is decoupled from the analysis:

```text
Stage 1 · embed    prompts.csv ──────────────► items.npz    (question wording only)
Stage 2 · pca      items.npz ───────────────► basis.npz     (embeddings only → PCA + dimension diagnostics)
Stage 3 · score    basis.npz + responses.csv + scales.csv + weights.csv ──► select D, then rank outliers
```

The coordinate system is built from the **meaning of the questions**, so the
embedding (stage 1) and the PCA basis (stage 2) depend only on the prompts — never
on anyone's answers. Stage 2 produces the **full** PCA decomposition plus the
*dimension diagnostics*; it does **not** yet commit to a working dimension `D`. The
responses, scales, reverse-scoring — and the **choice of `D`** — all enter at
stage 3. Stages 1–2 are deterministic, so the same prompts always yield the same
basis: build it once and reuse it across waves/cohorts to guarantee a shared space.

### Stage 1 — embed (needs only the prompts)

**`prompts.csv`** — the wording of each item. This is what builds the space:

```csv
item,prompt
sad,How often do you feel sad?
sleep,How often do you have trouble sleeping?
worry,How often do you feel worried?
```

```bash
survey-semantics embed --prompt-file prompts.csv --model models/bge-m3 --out items.npz
```

`items.npz` is a reusable embedding artifact (one vector per item). It depends
only on the wording — reuse it for every cohort that shares these questions.

### Stage 2 — pca: decomposition + dimension diagnostics (needs only the embeddings)

```bash
survey-semantics pca --embeddings-file items.npz --out basis.npz
```

`basis.npz` holds the **full** PCA of the item embeddings (every component) **plus
the dimension diagnostics** — the cumulative-variance curve, the eigengaps, and the
parallel-analysis null. It deliberately does **not** pick a working dimension `D`;
that is a stage-3 choice (below). No responses are involved, so the basis is fixed
by the prompts alone — reuse one `basis.npz` across waves and they share it.
(`--d-null-permutations` / `--d-null-percentile` configure the parallel-analysis
null computed here.)

### Stage 3 — score: select D, then rank outliers (responses, scales, weights enter here)

This stage first **chooses the working dimension `D`**, then projects the
responses into those `D` semantic axes, removes covariates, and ranks outliers.
`--d-selection` picks the rule: `variance` (default), `eigengap`, `parallel`, and
`max` read `D` straight from the basis's diagnostics; `stability` derives it from
the flagged outlier sets, so it needs the responses — which is why `D` selection
lives here, not in stage 2. One basis serves every rule.

**`responses.csv`** — one row per person: an id, optional `age`/`sex`, then one
column per item (names match the `item` keys above):

```csv
id,age,sex,sad,sleep,worry
P001,42,F,1,3,2
P002,67,M,5,1,4
```

**`scales.csv`** *(required)* — per-item valid range, missing-value codes, and the
`reverse` flag. Reverse-scoring is part of the method, so this file is mandatory
(set `reverse` to `false` for items that don't need it):

```csv
item,min,max,sentinels,reverse
sad,1,5,7;8;9,false
sleep,1,5,7;8;9,false
worry,1,5,7;8;9,false
```

Two optional columns can be added. **`embed`** is an item-selection allowlist:
when present, only `embed=true` rows are embedded and analyzed, and `embed=false`
rows stay in the file as documentation but are excluded — pass the same
`--scale-file` to `embed` so `items.npz` matches the analyzed item set.
**`ceiling`** marks which items count toward the `--pan-mild` ceiling check.

**`weights.csv`** *(required)* — one survey weight per row, in the same order as
`responses.csv` (one number per line; the `weight` header is optional):

```csv
weight
1842.6
903.1
```

**Unweighted analysis (as in the paper): make every weight `1`.** Equal weights
turn the weighting into a no-op — WLS residualization reduces to ordinary least
squares and the weighted Mahalanobis matches the unweighted distances (up to a
negligible constant), so the outlier ranking and empirical outlier set are
identical to an unweighted run:

```csv
weight
1
1
```

The file stays required either way, so a run always states its weighting choice
explicitly rather than defaulting silently.

Stage 3 needs the **responses**, the **basis**, the **scale file**, and the
**weights file**. `--prompt-file` is *optional* here — it only carries the wording
into the loadings/case-study outputs (the basis already fixes the items); the
inline shortcut below does need it, since that path re-embeds.

```bash
survey-semantics analyze-file responses.csv \
  --basis-file    basis.npz \
  --scale-file    scales.csv \
  --weights-file  weights.csv \
  --d-selection   variance \   # choose D here: variance|eigengap|parallel|stability|max
  --prompt-file   prompts.csv \  # optional: labels the loadings/case studies
  --outdir outputs/run         # no --model needed — the basis holds the decomposition
```

The ranking lands in `outputs/run/*_scores.csv` — the larger a person's
`Mahalanobis_Dist`, the more unusual their overall response pattern.

### One-shot shortcut

If you don't need the intermediate `items.npz` / `basis.npz`, `analyze-file` can
do all three stages at once — pass `--model` instead of `--basis-file` and it
embeds + builds the basis inline (a bit-for-bit identical result):

```bash
survey-semantics analyze-file responses.csv \
  --prompt-file prompts.csv --scale-file scales.csv --weights-file weights.csv \
  --model models/bge-m3 --outdir outputs/run
```

### Optional refinements

- **`--pan-mild`** — also flag below-ceiling outliers (adds `At_Ceiling`, `Is_Pan_Mild_Emp<pct>`).

(The dimension rule, `--d-selection`, is part of stage 3 above — `variance`
default, plus `eigengap` / `parallel` / `stability` / `max`.)

Full file formats and options: [docs/pipeline_overview.md](docs/pipeline_overview.md).

### Variants

- **Build the basis once, analyze many cohorts.** Stages 1–2 don't depend on
  responses, so reuse one `basis.npz` across waves/cohorts — they're guaranteed to
  share the same semantic space. (`--embeddings-file items.npz` is also accepted by
  `analyze-file` if you'd rather skip the explicit `pca` step but still avoid the model.)
- **Questionnaires in separate files** (not one merged table): `run-study --data-dir <dir> --prompt-dir <dir>` merges them for you.
- **Longitudinal / multiple waves** with differing item sets (e.g. NHIS 2021 vs 2024): restrict to common items so the waves share one space, run each, and compare — see the [NHIS example](examples/nhis/README.md).

## Outputs (`--outdir`)

- `*_scores.csv` — per-subject Mahalanobis distance, semantic PCs, outlier flags (the ranking).
- `*_prompt_loadings.csv` — how items load on each semantic dimension.
- `*_drivers.csv` — which items/dimensions drive each top outlier.
- `*_stability.csv`, `*_dimension_selection.csv` — how `D` was chosen and how stable it is.
- `case_studies/`, `summary.csv` — anonymized per-outlier reports and a run summary.

Add `--skip-umap` to skip UMAP files. Render plots with
`survey-semantics-plot outputs/run`.

## Layout

```text
src/survey_semantics/   package (io, prompts, scales, embedding, basis, pipeline, combined, cli, plotting)
tests/                  test suite
examples/nhis/          NHIS worked example (converter + run script + README)
docs/                   pipeline_overview.md, env_setup.md
```
