## Supplementary material for "Auditable cross-instrument detection of unusual multivariate psychiatric response configurations using a semantically aligned covariance subspace": Code for tool: requirements.txt

# Python 3.7-compatible local analysis environment.
# These pins are chosen to work with the default local Anaconda stack in this
# workspace and with umap-learn 0.4.6.
numpy>=1.17,<1.22
pandas>=1.0,<1.4
scipy>=1.3.1,<1.8
scikit-learn>=0.22,<1.1
umap-learn==0.4.6
matplotlib>=3.5,<3.6
