## Supplementary material for "Auditable cross-instrument detection of unusual multivariate psychiatric response configurations using a semantically aligned covariance subspace": Code for tool: plot_semantic_outputs.py

```
#!/usr/bin/env python
"""Create PDF plots from survey-semantics CSV outputs."""

import argparse
import os
import re
from pathlib import Path
from typing import Dict, Iterable, List, Optional

import numpy as np
import pandas as pd

os.environ.setdefault("MPLCONFIGDIR", "/private/tmp/survey_semantics_mplconfig")
Path(os.environ["MPLCONFIGDIR"]).mkdir(parents=True, exist_ok=True)

import matplotlib

matplotlib.use("Agg")
import matplotlib.pyplot as plt


ID_CANDIDATES = [
    "subjectkey",
    "src_subject_id",
    "export_id",
    "participant_id",
    "id",
    "Source_Row",
]


def main() -> int:
    parser = argparse.ArgumentParser(description=__doc__)
    parser.add_argument("input_dir", type=Path, help="Directory containing survey-semantics CSV outputs.")
    parser.add_argument("--outdir", type=Path, default=None, help="PDF output directory. Defaults to INPUT_DIR/plots.")
    parser.add_argument("--prefix", action="append", default=None, help="Only plot runs whose filename prefix contains this string.")
    parser.add_argument("--outlier-column", default="Is_Outlier_Emp95")
    parser.add_argument("--max-heatmap-rows", type=int, default=60)
    parser.add_argument("--label-column", default=None)
    parser.add_argument("--absolute-heatmap", action="store_true", help="Plot absolute PC z-scores instead of signed z.")
    args = parser.parse_args()

    input_dir = args.input_dir
    outdir = args.outdir or input_dir / "plots"
    outdir.mkdir(parents=True, exist_ok=True)

    runs = discover_runs(input_dir, args.prefix)
    if not runs:
        raise SystemExit("No *_scores.csv files found in {}".format(input_dir))

    for prefix, files in runs.items():
        scores = pd.read_csv(files["scores"])
        label_col = args.label_column or infer_label_column(scores)
        outlier_label_map = build_outlier_label_map(
            scores=scores,
            outlier_col=args.outlier_column,
            label_col=label_col,
            fallback_count=args.max_heatmap_rows,
        )
        outlier_label_map.to_csv(outdir / "{}_outlier_label_map.csv".format(prefix), index=False)

        if "stability" in files:
            stability = pd.read_csv(files["stability"])
            plot_variance_jaccard(stability, prefix, outdir / "{}_variance_jaccard.pdf".format(prefix))

        if "raw_response_umap" in files:
            raw_umap = pd.read_csv(files["raw_response_umap"])
            plot_umap(
                raw_umap,
                scores,
                prefix,
                "Raw Response UMAP",
                args.outlier_column,
                outlier_label_map,
                outdir / "{}_raw_response_umap.pdf".format(prefix),
            )

        if "semantic_pc_umap" in files:
            semantic_umap = pd.read_csv(files["semantic_pc_umap"])
            plot_umap(
                semantic_umap,
                scores,
                prefix,
                "Semantic PC UMAP",
                args.outlier_column,
                outlier_label_map,
                outdir / "{}_semantic_pc_umap.pdf".format(prefix),
            )

        plot_pc_z_heatmap(
            scores,
            prefix,
            args.outlier_column,
            outlier_label_map,
            args.max_heatmap_rows,
            outdir / "{}_pc_z_heatmap.pdf".format(prefix),
            absolute=args.absolute_heatmap,
        )

    print("Wrote plots for {} run(s) to {}".format(len(runs), outdir))
    return 0


def discover_runs(input_dir: Path, prefix_filters: Optional[Iterable[str]]) -> Dict[str, Dict[str, Path]]:
    suffixes = {
        "scores": "_scores.csv",
        "stability": "_stability.csv",
        "raw_response_umap": "_raw_response_umap.csv",
        "semantic_pc_umap": "_semantic_pc_umap.csv",
    }
    runs: Dict[str, Dict[str, Path]] = {}
    for scores_path in sorted(input_dir.glob("*_scores.csv")):
        prefix = scores_path.name[: -len("_scores.csv")]
        if prefix_filters and not any(token in prefix for token in prefix_filters):
            continue
        files = {"scores": scores_path}
        for key, suffix in suffixes.items():
            if key == "scores":
                continue
            candidate = input_dir / "{}{}".format(prefix, suffix)
            if candidate.exists():
                files[key] = candidate
        runs[prefix] = files
    return runs


def plot_variance_jaccard(stability: pd.DataFrame, prefix: str, outpath: Path) -> None:
    x = stability["components"].astype(float).values
    jaccard_next = neighboring_jaccard(stability)

    fig, axes = plt.subplots(2, 1, figsize=(9, 7), sharex=True)
    fig.suptitle("{}: Variance And Neighboring-D Outlier Stability".format(prefix), fontsize=12)

    if "cumulative_explained_variance" in stability.columns:
        axes[0].plot(
            x,
            stability["cumulative_explained_variance"].values,
            color="black",
            marker="o",
            linewidth=1.8,
            label="Cumulative explained variance",
        )
        if "explained_variance_ratio" in stability.columns:
            axes[0].bar(
                x,
                stability["explained_variance_ratio"].values,
                color="0.75",
                label="Per-PC variance",
            )
        axes[0].set_ylim(0, min(1.05, max(1.0, stability["cumulative_explained_variance"].max() * 1.08)))
        axes[0].set_ylabel("Semantic variance")
    else:
        axes[0].text(
            0.5,
            0.5,
            "Variance columns unavailable.\nRegenerate analysis outputs to include variance.",
            transform=axes[0].transAxes,
            ha="center",
            va="center",
        )
        axes[0].set_ylabel("Semantic variance")
    axes[0].grid(alpha=0.25)
    handles, labels = axes[0].get_legend_handles_labels()
    if handles:
        axes[0].legend(handles, labels, loc="best", fontsize=8)

    axes[1].plot(
        x,
        jaccard_next,
        color="tab:blue",
        marker="s",
        linewidth=1.8,
        label="Jaccard(D, D+1)",
    )
    axes_count = axes[1].twinx()
    axes_count.plot(
        x,
        stability["outlier_count"].values,
        color="tab:red",
        marker="o",
        linestyle="--",
        linewidth=1.4,
        label="Outlier count",
    )
    axes[1].set_ylim(0, 1.05)
    axes[1].set_xlabel("Semantic PCA components (D)")
    axes[1].set_ylabel("Neighboring-D Jaccard")
    axes_count.set_ylabel("Theoretical outlier count")
    axes[1].grid(alpha=0.25)
    lines, labels = axes[1].get_legend_handles_labels()
    lines2, labels2 = axes_count.get_legend_handles_labels()
    axes[1].legend(lines + lines2, labels + labels2, loc="best", fontsize=8)

    fig.tight_layout(rect=[0, 0, 1, 0.96])
    fig.savefig(outpath, bbox_inches="tight")
    plt.close(fig)


def neighboring_jaccard(stability: pd.DataFrame) -> np.ndarray:
    if "jaccard_vs_next" in stability.columns:
        return stability["jaccard_vs_next"].astype(float).values
    if "jaccard_vs_previous" in stability.columns:
        values = stability["jaccard_vs_previous"].astype(float).values
        shifted = np.empty_like(values)
        shifted[:-1] = values[1:]
        shifted[-1] = np.nan
        return shifted
    return np.full(len(stability), np.nan)


def plot_umap(
    umap_df: pd.DataFrame,
    scores: pd.DataFrame,
    prefix: str,
    title: str,
    outlier_col: str,
    outlier_label_map: pd.DataFrame,
    outpath: Path,
) -> None:
    merged = merge_on_source_row(umap_df, scores)
    outlier_mask = outlier_series(merged, outlier_col)

    fig, ax = plt.subplots(figsize=(8, 7))
    ax.scatter(
        merged.loc[~outlier_mask, "UMAP_Dim1"],
        merged.loc[~outlier_mask, "UMAP_Dim2"],
        s=18,
        alpha=0.45,
        c="0.65",
        edgecolors="none",
        label="Not {}".format(outlier_col),
    )
    ax.scatter(
        merged.loc[outlier_mask, "UMAP_Dim1"],
        merged.loc[outlier_mask, "UMAP_Dim2"],
        s=48,
        alpha=0.95,
        c="tab:red",
        edgecolors="black",
        linewidths=0.4,
        label=outlier_col,
    )

    annotate_top_outliers(ax, merged.loc[outlier_mask], outlier_label_map, max_labels=10)
    ax.set_title("{}: {}".format(prefix, title), fontsize=12)
    ax.set_xlabel("UMAP dimension 1")
    ax.set_ylabel("UMAP dimension 2")
    ax.grid(alpha=0.18)
    ax.legend(loc="best", fontsize=8)
    fig.tight_layout()
    fig.savefig(outpath, bbox_inches="tight")
    plt.close(fig)


def plot_pc_z_heatmap(
    scores: pd.DataFrame,
    prefix: str,
    outlier_col: str,
    outlier_label_map: pd.DataFrame,
    max_rows: int,
    outpath: Path,
    absolute: bool = False,
) -> None:
    pc_cols = semantic_pc_columns(scores)
    if not pc_cols:
        raise ValueError("No Semantic_PC* columns found in scores for {}".format(prefix))

    values = scores[pc_cols].astype(float)
    z = (values - values.mean(axis=0)) / values.std(axis=0).replace(0, 1)
    outliers = scores.loc[outlier_series(scores, outlier_col)].copy()
    if outliers.empty:
        outliers = scores.sort_values("Mahalanobis_Dist", ascending=False).head(max_rows).copy()
    else:
        outliers = outliers.sort_values("Mahal_Rank").head(max_rows).copy()

    heat = z.loc[outliers.index, pc_cols]
    if absolute:
        heat = heat.abs()
    labels = anonymized_labels_for_rows(outliers, outlier_label_map)
    vmax = np.nanpercentile(np.abs(heat.values), 98)
    vmax = max(float(vmax), 1.0)

    fig_height = min(14, max(4, 0.28 * len(heat) + 1.8))
    fig_width = min(16, max(6, 0.45 * len(pc_cols) + 3.0))
    fig, ax = plt.subplots(figsize=(fig_width, fig_height))
    if absolute:
        image = ax.imshow(heat.values, aspect="auto", cmap="magma", vmin=0, vmax=vmax)
        title = "{}: Outlier Semantic PC Absolute Z-Scores".format(prefix)
        colorbar_label = "|z-score|"
    else:
        image = ax.imshow(heat.values, aspect="auto", cmap="coolwarm", vmin=-vmax, vmax=vmax)
        title = "{}: Outlier Semantic PC Z-Scores".format(prefix)
        colorbar_label = "Signed z-score"
    ax.set_title(title, fontsize=12)
    ax.set_xlabel("Semantic PC")
    ax.set_ylabel("Outlier")
    ax.set_xticks(np.arange(len(pc_cols)))
    ax.set_xticklabels([col.replace("Semantic_", "") for col in pc_cols], rotation=45, ha="right")
    ax.set_yticks(np.arange(len(labels)))
    ax.set_yticklabels(labels, fontsize=7 if len(labels) > 35 else 8)
    colorbar = fig.colorbar(image, ax=ax, fraction=0.035, pad=0.02)
    colorbar.set_label(colorbar_label)
    fig.tight_layout()
    fig.savefig(outpath, bbox_inches="tight")
    plt.close(fig)


def merge_on_source_row(left: pd.DataFrame, scores: pd.DataFrame) -> pd.DataFrame:
    score_cols = [
        col
        for col in scores.columns
        if col in {"Source_Row", "Mahalanobis_Dist", "Mahal_Rank", "Is_Outlier_Theo", "Is_Outlier_Emp95", "Is_Outlier_Emp99"}
        or col in ID_CANDIDATES
    ]
    score_cols = list(dict.fromkeys(score_cols))
    return left.merge(scores[score_cols], on="Source_Row", how="left", suffixes=("", "_score"))


def outlier_series(df: pd.DataFrame, outlier_col: str) -> pd.Series:
    if outlier_col in df.columns:
        return df[outlier_col].fillna(False).astype(bool)
    if "Is_Outlier_Emp95" in df.columns:
        return df["Is_Outlier_Emp95"].fillna(False).astype(bool)
    if "Mahal_Rank" in df.columns:
        return df["Mahal_Rank"] <= max(1, int(np.ceil(0.05 * len(df))))
    return pd.Series(False, index=df.index)


def semantic_pc_columns(df: pd.DataFrame) -> List[str]:
    cols = [col for col in df.columns if re.match(r"^Semantic_PC\d+$", str(col))]
    return sorted(cols, key=lambda col: int(re.search(r"\d+", col).group(0)))


def infer_label_column(df: pd.DataFrame) -> str:
    for col in ID_CANDIDATES:
        if col in df.columns:
            return col
    return "Source_Row"


def build_outlier_label_map(
    scores: pd.DataFrame,
    outlier_col: str,
    label_col: str,
    fallback_count: int,
) -> pd.DataFrame:
    outliers = scores.loc[outlier_series(scores, outlier_col)].copy()
    included_by = outlier_col
    if outliers.empty:
        outliers = scores.sort_values("Mahalanobis_Dist", ascending=False).head(fallback_count).copy()
        included_by = "top_mahalanobis_fallback"
    else:
        outliers = outliers.sort_values("Mahal_Rank").copy()

    records = []
    for anon_idx, (_, row) in enumerate(outliers.iterrows(), start=1):
        records.append(
            {
                "outlier_label": "Outlier #{}".format(anon_idx),
                "subject_id": row[label_col] if label_col in row.index else pd.NA,
                "label_column": label_col if label_col in row.index else "",
                "source_row": row["Source_Row"] if "Source_Row" in row.index else pd.NA,
                "mahal_rank": row["Mahal_Rank"] if "Mahal_Rank" in row.index else pd.NA,
                "mahalanobis_dist": row["Mahalanobis_Dist"] if "Mahalanobis_Dist" in row.index else pd.NA,
                "outlier_column": outlier_col,
                "included_by": included_by,
            }
        )
    return pd.DataFrame(records)


def anonymized_labels_for_rows(rows: pd.DataFrame, outlier_label_map: pd.DataFrame) -> np.ndarray:
    lookup = {
        row["source_row"]: row["outlier_label"]
        for _, row in outlier_label_map.iterrows()
        if pd.notna(row.get("source_row"))
    }
    labels = []
    for _, row in rows.iterrows():
        source_row = row["Source_Row"] if "Source_Row" in row.index else pd.NA
        labels.append(lookup.get(source_row, "Outlier"))
    return np.asarray(labels, dtype=object)


def annotate_top_outliers(ax, outliers: pd.DataFrame, outlier_label_map: pd.DataFrame, max_labels: int) -> None:
    if outliers.empty:
        return
    label_lookup = {
        row["source_row"]: row["outlier_label"]
        for _, row in outlier_label_map.iterrows()
        if pd.notna(row.get("source_row"))
    }
    ordered = outliers.sort_values("Mahal_Rank") if "Mahal_Rank" in outliers.columns else outliers
    for _, row in ordered.head(max_labels).iterrows():
        source_row = row["Source_Row"] if "Source_Row" in row.index else pd.NA
        label = str(label_lookup.get(source_row, "Outlier"))
        ax.annotate(
            label,
            (row["UMAP_Dim1"], row["UMAP_Dim2"]),
            xytext=(3, 3),
            textcoords="offset points",
            fontsize=6,
            alpha=0.85,
        )


if __name__ == "__main__":
    raise SystemExit(main())
```
