## Supplementary material for "Auditable cross-instrument detection of unusual multivariate psychiatric response configurations using a semantically aligned covariance subspace": Code for tool: __init__.py

```
"""Semantic survey analysis tools."""

from survey_semantics.io import SurveyTable, infer_item_columns, read_survey_table
from survey_semantics.pipeline import AnalysisConfig, AnalysisResult, analyze_survey_table
from survey_semantics.combined import CombinedPackage, build_combined_package_table

__all__ = [
    "AnalysisConfig",
    "AnalysisResult",
    "CombinedPackage",
    "SurveyTable",
    "analyze_survey_table",
    "build_combined_package_table",
    "infer_item_columns",
    "read_survey_table",
]
```
