## Supplementary material for "Auditable cross-instrument detection of unusual multivariate psychiatric response configurations using a semantically aligned covariance subspace": Code for tool: basis.py

```
"""Semantic PCA basis — the response-independent middle stage of the pipeline.

The pipeline has three modular stages:

1. **embed**  : item prompts -> :class:`~survey_semantics.embedding.ItemEmbeddings`
   (depends only on the wording).
2. **pca**    : item embeddings -> :class:`SemanticBasis` (this module): the PCA
   decomposition of the centered item embeddings plus the embedding-only
   dimension diagnostics (variance/eigengap/parallel). Depends only on the
   embeddings, never on responses.
3. **score**  : basis + responses + scales (+ weights) -> outliers.

Because the basis depends only on the prompts, the same embeddings always yield
the same basis, and one basis can be reused across cohorts/waves to guarantee a
shared semantic space. The stability dimension rule is the only D diagnostic that
needs responses, so it is computed downstream at score time — not here.
"""

from dataclasses import dataclass
from pathlib import Path
from typing import Dict, List, Sequence

import numpy as np
from sklearn.decomposition import PCA


@dataclass
class SemanticBasis:
    """A reusable PCA basis fit on centered item embeddings.

    ``components`` are the item coordinates (one row per item, columns = PCs);
    the eigen/variance arrays and ``parallel`` diagnostics are global (per-PC),
    and the embedding provenance flows through for output naming.
    """

    items: List[str]
    components: np.ndarray              # (len(items), n_components) item coordinates — full decomposition
    eigenvalues: np.ndarray
    explained_variance_ratio: np.ndarray
    cumulative_variance: np.ndarray
    parallel: Dict[str, object]         # selected_d, null_mean, null_percentile, significant
    n_components: int                   # number of PCs in the basis (always the full decomposition)
    embedding_backend: str
    embedding_model: str
    embedding_slug: str
    d_null_permutations: int
    d_null_percentile: float
    random_state: int

    def coordinates_for(self, item_columns: Sequence[str]) -> np.ndarray:
        """Return the item coordinates reordered to ``item_columns``.

        Requires the basis to have been fit on **exactly** the analyzed item set
        (same items, any order). Raises on any missing or extra item rather than
        silently doing a partial projection through a basis fit on a different
        item set.
        """
        index = {item: i for i, item in enumerate(self.items)}
        requested = list(item_columns)
        missing = [c for c in requested if c not in index]
        if missing:
            raise ValueError(
                "Semantic basis is missing {} analyzed item(s): {}. Rebuild the "
                "basis (pca step) over exactly the analyzed items.".format(
                    len(missing), ", ".join(map(str, missing[:10]))
                )
            )
        extra = [it for it in self.items if it not in set(requested)]
        if extra:
            raise ValueError(
                "Semantic basis was fit on {} item(s) not in the analyzed set: {}. "
                "A basis must be fit on exactly the analyzed items, else the "
                "projection would be partial. Rebuild the basis over this item "
                "set.".format(len(extra), ", ".join(map(str, extra[:10])))
            )
        return self.components[[index[c] for c in requested]]


def build_semantic_basis(
    items: Sequence[str],
    embedding_vectors: np.ndarray,
    d_null_permutations: int = 50,
    d_null_percentile: float = 95.0,
    random_state: int = 42,
    embedding_backend: str = "",
    embedding_model: str = "",
    embedding_slug: str = "",
) -> SemanticBasis:
    """Center the item embeddings, run the **full** PCA, and compute the
    embedding-only D diagnostics. The basis always keeps every component — the
    dimension is chosen later, at score time, via ``--d-selection``."""

    # Lazy import avoids a circular import: pipeline imports this module.
    from survey_semantics.pipeline import parallel_analysis

    vectors = np.asarray(embedding_vectors, dtype=float)
    centered = vectors - vectors.mean(axis=0, keepdims=True)

    n_components = min(centered.shape[0], centered.shape[1])
    if n_components < 1:
        raise ValueError("Item text embedding did not produce usable features.")

    pca = PCA(n_components=n_components, svd_solver="full")
    components = pca.fit_transform(centered)
    eigenvalues = np.nan_to_num(pca.explained_variance_)
    explained = np.nan_to_num(pca.explained_variance_ratio_)
    cumulative = np.cumsum(explained)
    parallel = parallel_analysis(
        matrix=centered,
        observed_eigenvalues=eigenvalues,
        n_components=n_components,
        n_permutations=d_null_permutations,
        percentile=d_null_percentile,
        random_state=random_state,
    )
    return SemanticBasis(
        items=[str(it) for it in items],
        components=components,
        eigenvalues=eigenvalues,
        explained_variance_ratio=explained,
        cumulative_variance=cumulative,
        parallel=parallel,
        n_components=n_components,
        embedding_backend=embedding_backend,
        embedding_model=embedding_model,
        embedding_slug=embedding_slug,
        d_null_permutations=int(d_null_permutations),
        d_null_percentile=float(d_null_percentile),
        random_state=int(random_state),
    )


def save_semantic_basis(path: Path, basis: SemanticBasis) -> None:
    """Persist a basis to a `.npz` file (no pickle; portable)."""

    import pandas as pd

    selected_d = basis.parallel.get("selected_d")
    selected_d_value = float("nan") if pd.isna(selected_d) else float(selected_d)
    np.savez(
        Path(path),
        items=np.array(basis.items),
        components=np.asarray(basis.components, dtype=float),
        eigenvalues=np.asarray(basis.eigenvalues, dtype=float),
        explained_variance_ratio=np.asarray(basis.explained_variance_ratio, dtype=float),
        cumulative_variance=np.asarray(basis.cumulative_variance, dtype=float),
        parallel_null_mean=np.asarray(basis.parallel["null_mean"], dtype=float),
        parallel_null_percentile=np.asarray(basis.parallel["null_percentile"], dtype=float),
        parallel_significant=np.asarray(basis.parallel["significant"], dtype=bool),
        parallel_selected_d=np.array([selected_d_value], dtype=float),
        meta=np.array([basis.embedding_backend, basis.embedding_model, basis.embedding_slug]),
        params=np.array(
            [basis.n_components, basis.d_null_permutations,
             basis.d_null_percentile, basis.random_state],
            dtype=float,
        ),
    )


def load_semantic_basis(path: Path) -> SemanticBasis:
    """Load a basis written by :func:`save_semantic_basis`."""

    import pandas as pd

    data = np.load(Path(path))
    backend, model_name, slug = (str(x) for x in data["meta"])
    _n_components, d_null_perm, d_null_pct, random_state = data["params"]
    selected_d_raw = float(data["parallel_selected_d"][0])
    selected_d = pd.NA if np.isnan(selected_d_raw) else int(selected_d_raw)
    parallel = {
        "selected_d": selected_d,
        "null_mean": np.asarray(data["parallel_null_mean"], dtype=float),
        "null_percentile": np.asarray(data["parallel_null_percentile"], dtype=float),
        "significant": np.asarray(data["parallel_significant"], dtype=bool),
    }
    return SemanticBasis(
        items=[str(x) for x in data["items"]],
        components=np.asarray(data["components"], dtype=float),
        eigenvalues=np.asarray(data["eigenvalues"], dtype=float),
        explained_variance_ratio=np.asarray(data["explained_variance_ratio"], dtype=float),
        cumulative_variance=np.asarray(data["cumulative_variance"], dtype=float),
        parallel=parallel,
        n_components=int(data["components"].shape[1]),
        embedding_backend=backend,
        embedding_model=model_name,
        embedding_slug=slug,
        d_null_permutations=int(d_null_perm),
        d_null_percentile=float(d_null_pct),
        random_state=int(random_state),
    )
```
