## Supplementary material for "Auditable cross-instrument detection of unusual multivariate psychiatric response configurations using a semantically aligned covariance subspace": Code for tool: cli.py

```
"""Command line interface for survey semantic analysis."""

import argparse
import re
from pathlib import Path
from typing import List, Optional

import pandas as pd

from survey_semantics.basis import (
    build_semantic_basis,
    load_semantic_basis,
    save_semantic_basis,
)
from survey_semantics.combined import build_combined_package_table
from survey_semantics.embedding import (
    embed_item_prompts,
    embedding_slug,
    load_item_embeddings,
    save_item_embeddings,
)
from survey_semantics.io import load_weights_file, read_survey_table
from survey_semantics.pipeline import AnalysisConfig, analyze_survey_table
from survey_semantics.prompts import load_prompt_sources
from survey_semantics.scales import (
    is_item_embedded,
    load_scale_sources,
    resolve_scale,
    scales_use_embed,
)


def main(argv: Optional[List[str]] = None) -> int:
    parser = argparse.ArgumentParser(prog="survey-semantics")
    subparsers = parser.add_subparsers(dest="command", required=True)

    embed_parser = subparsers.add_parser(
        "embed",
        help="LLM step only: embed item prompts into a reusable embeddings file (no responses).",
    )
    embed_parser.add_argument("--prompt-file", type=Path, default=None, help="item,prompt CSV/TSV.")
    embed_parser.add_argument("--prompt-dir", type=Path, default=None)
    embed_parser.add_argument("--model", default=None, help="Local sentence-transformers model path (e.g. bge-m3).")
    embed_parser.add_argument("--out", type=Path, required=True, help="Output .npz embeddings file.")
    embed_parser.add_argument(
        "--scale-file", type=Path, default=None,
        help="Optional per-item scale file. When it declares an `embed` column, only "
             "embed=true items are embedded (matches the analysis item set exactly).",
    )
    embed_parser.add_argument("--scale-dir", type=Path, default=None, help="Directory of per-instrument scale files.")
    embed_parser.add_argument(
        "--trust-remote-code", action="store_true",
        help="Explicit opt-in for models whose repo ships custom architecture code "
             "(e.g. gte-large-en-v1.5). Code runs from local files only; the "
             "outbound-socket blocker stays active regardless.",
    )

    pca_parser = subparsers.add_parser(
        "pca",
        help="PCA step only: turn an embeddings file into a reusable semantic basis (no responses).",
    )
    pca_parser.add_argument(
        "--embeddings-file", type=Path, required=True,
        help="Item embeddings produced by `embed`.",
    )
    pca_parser.add_argument("--out", type=Path, required=True, help="Output .npz basis file.")
    pca_parser.add_argument("--d-null-permutations", type=int, default=50)
    pca_parser.add_argument("--d-null-percentile", type=float, default=95.0)
    pca_parser.add_argument("--random-state", type=int, default=42)

    file_parser = subparsers.add_parser("analyze-file", help="Analyze one survey table.")
    _add_common_args(file_parser)
    file_parser.add_argument("path", type=Path)
    file_parser.add_argument("--name", default=None)
    file_parser.add_argument(
        "--embeddings-file", type=Path, default=None,
        help="Use precomputed item embeddings from `embed` (skips the LLM; --model not needed).",
    )
    file_parser.add_argument(
        "--basis-file", type=Path, default=None,
        help="Use a precomputed semantic basis from `pca` (skips the LLM and PCA; "
             "--model/--embeddings-file not needed). Must be fit on exactly the analyzed items.",
    )

    package_parser = subparsers.add_parser("analyze-package", help="Analyze all viable survey tables in a folder.")
    _add_common_args(package_parser)
    package_parser.add_argument("path", type=Path)
    package_parser.add_argument("--include-regex", default=None)
    package_parser.add_argument("--exclude-regex", default=None)
    package_parser.add_argument("--max-tables", type=int, default=0, help="0 means no limit.")

    combined_parser = subparsers.add_parser(
        "analyze-package-combined",
        help="Analyze all viable questionnaires in one package-level common semantic space.",
    )
    _add_common_args(combined_parser)
    combined_parser.add_argument("path", type=Path)
    combined_parser.add_argument("--include-regex", default=None)
    combined_parser.add_argument("--exclude-regex", default=None)
    combined_parser.add_argument("--reverse-config", type=Path, default=None)
    combined_parser.add_argument("--combined-min-complete-fraction", type=float, default=0.05)
    combined_parser.add_argument("--no-auto-reverse", action="store_true")
    combined_parser.add_argument("--auto-reverse-corr-threshold", type=float, default=0.70)
    combined_parser.add_argument("--auto-reverse-min-pairwise-subjects", type=int, default=10)
    combined_parser.add_argument("--auto-reverse-min-pairwise-fraction", type=float, default=0.50)

    study_parser = subparsers.add_parser(
        "run-study",
        help="Run the notebook-equivalent combined workflow from a prompt dictionary and data directory.",
    )
    _add_common_args(study_parser)
    study_parser.add_argument("--embedding-model", required=True, help="Local sentence-transformers model name/path (e.g. a bge-m3 path). No TF-IDF/auto fallback.")
    study_parser.add_argument("--data-dir", type=Path, required=True, help="Directory containing survey data files.")
    study_parser.add_argument("--include-regex", default=None)
    study_parser.add_argument("--exclude-regex", default=None)
    study_parser.add_argument("--reverse-config", type=Path, default=None)
    study_parser.add_argument("--combined-min-complete-fraction", type=float, default=0.05)
    study_parser.add_argument("--no-auto-reverse", action="store_true")
    study_parser.add_argument("--auto-reverse-corr-threshold", type=float, default=0.70)
    study_parser.add_argument("--auto-reverse-min-pairwise-subjects", type=int, default=10)
    study_parser.add_argument("--auto-reverse-min-pairwise-fraction", type=float, default=0.50)
    study_parser.add_argument("--skip-plots", action="store_true")
    study_parser.add_argument("--plots-outdir", type=Path, default=None)
    study_parser.add_argument("--plot-outlier-column", default="Is_Outlier_Emp95")
    study_parser.add_argument("--max-heatmap-rows", type=int, default=60)
    study_parser.add_argument("--absolute-heatmap", action="store_true")

    args = parser.parse_args(argv)

    if args.command == "embed":
        return _embed(args)
    if args.command == "pca":
        return _pca(args)
    if args.command == "analyze-file":
        return _analyze_file(args)
    if args.command == "analyze-package":
        return _analyze_package(args)
    if args.command == "analyze-package-combined":
        return _analyze_package_combined(args)
    if args.command == "run-study":
        return _run_study(args, parser)
    parser.error("Unknown command.")
    return 2


def _add_common_args(parser: argparse.ArgumentParser) -> None:
    parser.add_argument("--outdir", type=Path, required=True)
    parser.add_argument(
        "--embedding", default="sentence-transformers", choices=["sentence-transformers"],
        help="Embedding backend. Only a local sentence-transformers model is supported; "
             "there is no fallback.",
    )
    parser.add_argument(
        "--model", default=None,
        help="Local path or cached name of the sentence-transformers model (e.g. a bge-m3 path). "
             "Required in practice — the model must already be on local disk (offline-only).",
    )
    parser.add_argument("--prompt-file", type=Path, default=None)
    parser.add_argument("--prompt-dir", type=Path, default=None)
    parser.add_argument(
        "--scale-file", type=Path, default=None,
        help="Per-item scale file (item,min,max,sentinels,reverse[,ceiling,embed]). "
             "Declares the analyzed item set, per-item missing codes, valid ranges, "
             "and reverse scoring. An optional `embed` column is an allowlist: only "
             "embed=true items are analyzed (embed=false items stay documented).",
    )
    parser.add_argument("--scale-dir", type=Path, default=None, help="Directory of per-instrument scale files.")
    parser.add_argument(
        "--weights-file", type=Path, default=None,
        help="One survey weight per subject, row-aligned to the response file. Enables "
             "weighted (WLS) residualization and weighted Mahalanobis distances.",
    )
    parser.add_argument(
        "--pan-mild", action="store_true",
        help="Flag pan-mild outliers: empirical-percentile outliers whose item profile "
             "is nowhere at the Likert ceiling (adds At_Ceiling + Is_Pan_Mild_Emp<pct> columns).",
    )
    parser.add_argument(
        "--ceiling-min-levels", type=int, default=3,
        help="Minimum response levels for an item to count toward the ceiling audit (default 3; "
             "binary items are excluded, matching the manuscript).",
    )
    parser.add_argument(
        "--empirical-percentiles", type=int, nargs="+", default=[95, 99],
        help="Empirical Mahalanobis percentiles for outlier/pan-mild flagging (default: 95 99).",
    )
    parser.add_argument("--skip-umap", action="store_true", help="Do not generate raw/semantic UMAP coordinate files.")
    parser.add_argument("--umap-neighbors", type=int, default=15)
    parser.add_argument("--umap-min-dist", type=float, default=0.10)
    parser.add_argument("--umap-metric", default="euclidean")
    parser.add_argument("--random-state", type=int, default=42)
    parser.add_argument("--min-rows", type=int, default=10)
    parser.add_argument("--min-items", type=int, default=5)
    parser.add_argument("--max-unique", type=int, default=8)
    parser.add_argument("--variance-threshold", type=float, default=0.80)
    parser.add_argument(
        "--d-selection",
        default="variance",
        choices=["variance", "eigengap", "parallel", "stability", "max", "all"],
        help="D selection rule used for scoring; 'max'/'all' uses every evaluated prompt PC.",
    )
    parser.add_argument("--d-null-permutations", type=int, default=50)
    parser.add_argument("--d-null-percentile", type=float, default=95.0)
    parser.add_argument("--stability-jaccard-threshold", type=float, default=0.90)
    parser.add_argument("--stability-consecutive", type=int, default=2)
    parser.add_argument("--alpha", type=float, default=0.01)
    parser.add_argument("--id-col", default=None)
    parser.add_argument("--covariates", nargs="*", default=None)
    parser.add_argument("--top-outliers", type=int, default=10)
    parser.add_argument("--top-components", type=int, default=3)
    parser.add_argument("--top-items", type=int, default=5)


def _config_from_args(args: argparse.Namespace) -> AnalysisConfig:
    item_scales = load_scale_sources(
        getattr(args, "scale_file", None),
        getattr(args, "scale_dir", None),
    ) or None
    weights_file = getattr(args, "weights_file", None)
    sample_weights = load_weights_file(weights_file) if weights_file else None
    return AnalysisConfig(
        item_scales=item_scales,
        sample_weights=sample_weights,
        pan_mild=getattr(args, "pan_mild", False),
        ceiling_min_levels=getattr(args, "ceiling_min_levels", 3),
        empirical_percentiles=tuple(getattr(args, "empirical_percentiles", None) or (95, 99)),
        embedding=args.embedding,
        model_name=args.model,
        disable_network=True,
        compute_umap=not args.skip_umap,
        umap_n_neighbors=args.umap_neighbors,
        umap_min_dist=args.umap_min_dist,
        umap_metric=args.umap_metric,
        random_state=args.random_state,
        min_rows=args.min_rows,
        min_items=args.min_items,
        max_unique=args.max_unique,
        variance_threshold=args.variance_threshold,
        d_selection_method=args.d_selection,
        d_null_permutations=args.d_null_permutations,
        d_null_percentile=args.d_null_percentile,
        stability_jaccard_threshold=args.stability_jaccard_threshold,
        stability_consecutive=args.stability_consecutive,
        alpha=args.alpha,
        id_col=args.id_col,
        covariates=args.covariates,
        top_outliers=args.top_outliers,
        top_components=args.top_components,
        top_items=args.top_items,
    )


def _flat_prompts(args: argparse.Namespace) -> dict:
    """Read a flat item->wording mapping from --prompt-file / --prompt-dir."""
    files = []
    if args.prompt_dir:
        files += sorted(
            p for p in Path(args.prompt_dir).iterdir()
            if p.suffix.lower() in {".csv", ".tsv", ".tab", ".txt"}
        )
    if args.prompt_file:
        files.append(Path(args.prompt_file))
    if not files:
        raise SystemExit("embed: provide --prompt-file or --prompt-dir.")

    mapping = {}
    for path in files:
        sep = "\t" if path.suffix.lower() in {".tsv", ".tab"} else ","
        frame = pd.read_csv(path, sep=sep)
        cols = {str(c).strip().lower(): c for c in frame.columns}
        item_col = next((cols[k] for k in ("item", "feature", "column", "variable") if k in cols), None)
        prompt_col = cols.get("prompt")
        if item_col is None or prompt_col is None:
            raise SystemExit("embed: {} needs 'item' and 'prompt' columns.".format(path))
        table_col = cols.get("table") or cols.get("instrument")
        for _, row in frame.iterrows():
            item = str(row[item_col]).strip()
            text = str(row[prompt_col]).strip()
            if not item or not text:
                continue
            if table_col and str(row[table_col]).strip():
                item = "{}__{}".format(str(row[table_col]).strip(), item)
            mapping[item] = text
    return mapping


def _embed(args: argparse.Namespace) -> int:
    prompts = _flat_prompts(args)
    if not prompts:
        raise SystemExit("embed: no item,prompt rows found.")
    scales = load_scale_sources(
        getattr(args, "scale_file", None), getattr(args, "scale_dir", None)
    )
    if scales:
        prompts = _filter_prompts_by_embed(prompts, scales)
        if not prompts:
            raise SystemExit(
                "embed: the scale file marks no items embed=true (nothing to embed)."
            )
    embeddings = embed_item_prompts(
        prompts, method="sentence-transformers", model_name=args.model,
        trust_remote_code=getattr(args, "trust_remote_code", False),
    )
    args.out.parent.mkdir(parents=True, exist_ok=True)
    save_item_embeddings(args.out, embeddings)
    print("Embedded {} items -> {} ({}).".format(len(embeddings.items), args.out, embeddings.slug))
    return 0


def _pca(args: argparse.Namespace) -> int:
    embeddings = load_item_embeddings(args.embeddings_file)
    basis = build_semantic_basis(
        items=embeddings.items,
        embedding_vectors=embeddings.vectors,
        d_null_permutations=args.d_null_permutations,
        d_null_percentile=args.d_null_percentile,
        random_state=args.random_state,
        embedding_backend=embeddings.backend,
        embedding_model=embeddings.model_name,
        embedding_slug=embeddings.slug,
    )
    args.out.parent.mkdir(parents=True, exist_ok=True)
    save_semantic_basis(args.out, basis)
    print(
        "Built semantic basis: {} items, {} PCs -> {} ({}).".format(
            len(basis.items), basis.n_components, args.out, basis.embedding_slug
        )
    )
    return 0


def _filter_prompts_by_embed(prompts: dict, scales: dict) -> dict:
    """Keep only prompt items selected by the scale file's `embed` allowlist.

    Each prompt key may be bare (``gad7_1``) or qualified (``COPE__gad7_1``);
    it is resolved against the scales with the same key-matching as the pipeline.
    When no item declares `embed`, all prompt items with a declared scale are
    kept (the pre-`embed` behavior).
    """
    uses_embed = scales_use_embed(scales)
    kept = {}
    for key, text in prompts.items():
        table, _, item = str(key).rpartition("__")
        scale = resolve_scale(scales, table, item or key)
        if is_item_embedded(scale, uses_embed):
            kept[key] = text
    return kept


def _analyze_file(args: argparse.Namespace) -> int:
    if not (args.scale_file or args.scale_dir):
        raise SystemExit(
            "A scale file is required: pass --scale-file (or --scale-dir). It carries "
            "the per-item reverse flag, valid range, and missing-value codes that the "
            "method needs (reverse-scoring is part of the method). Running without it "
            "would silently skip reverse-scoring and guess ranges from the data."
        )
    if not args.weights_file:
        raise SystemExit(
            "A weights file is required: pass --weights-file (one weight per response "
            "row). For an unweighted analysis, supply a column of equal weights (e.g. all "
            "1s) — equal weights give the same outlier ranking and empirical outlier set "
            "as the unweighted method (distances differ only by a negligible constant)."
        )
    args.outdir.mkdir(parents=True, exist_ok=True)
    prompts = load_prompt_sources(args.prompt_file, args.prompt_dir)
    table = read_survey_table(args.path, prompt_dictionary=prompts)
    if args.name:
        table.name = args.name
    item_embeddings = None
    basis = None
    if getattr(args, "basis_file", None):
        basis = load_semantic_basis(args.basis_file)
    elif getattr(args, "embeddings_file", None):
        item_embeddings = load_item_embeddings(args.embeddings_file)
    result = analyze_survey_table(
        table, _config_from_args(args), item_embeddings=item_embeddings, basis=basis,
    )
    _write_result(result, args.outdir, _safe_name(result.table_name))
    _write_summary(pd.DataFrame([result.summary]), args.outdir, result.summary["embedding_slug"])
    print("Analyzed {}: {} rows, {} items, D={}.".format(
        result.table_name,
        result.summary["n_rows"],
        result.summary["n_items"],
        result.summary["optimal_d"],
    ))
    return 0


def _analyze_package(args: argparse.Namespace) -> int:
    args.outdir.mkdir(parents=True, exist_ok=True)
    include = re.compile(args.include_regex) if args.include_regex else None
    exclude = re.compile(args.exclude_regex) if args.exclude_regex else None
    config = _config_from_args(args)
    prompts = load_prompt_sources(args.prompt_file, args.prompt_dir)

    summaries = []
    analyzed = 0
    files = sorted(list(args.path.glob("*.txt")) + list(args.path.glob("*.tsv")) + list(args.path.glob("*.csv")))
    for path in files:
        if include and not include.search(path.name):
            continue
        if exclude and exclude.search(path.name):
            continue
        if args.max_tables and analyzed >= args.max_tables:
            break

        try:
            table = read_survey_table(path, prompt_dictionary=prompts)
            result = analyze_survey_table(table, config)
        except Exception as exc:
            reason = str(exc)
            summaries.append({"table": path.stem, "path": str(path), "status": "skipped", "reason": reason})
            print("Skipped {}: {}".format(path.stem, reason))
            if "UMAP outputs require" in reason:
                _write_summary(pd.DataFrame(summaries), args.outdir, _requested_embedding_slug(config))
                return 1
            continue

        prefix = _safe_name(result.table_name)
        _write_result(result, args.outdir, prefix)
        row = dict(result.summary)
        row["status"] = "analyzed"
        row["reason"] = ""
        summaries.append(row)
        analyzed += 1
        print("Analyzed {}: {} rows, {} items, D={}.".format(
            result.table_name,
            result.summary["n_rows"],
            result.summary["n_items"],
            result.summary["optimal_d"],
        ))

    summary = pd.DataFrame(summaries)
    summary_slug = _summary_slug(summary, config)
    _write_summary(summary, args.outdir, summary_slug)
    print("Package complete: {} analyzed, {} scanned.".format(analyzed, len(summaries)))
    return 0


def _analyze_package_combined(args: argparse.Namespace) -> int:
    args.outdir.mkdir(parents=True, exist_ok=True)
    config = _config_from_args(args)
    config.min_complete_fraction = args.combined_min_complete_fraction
    prompts = load_prompt_sources(args.prompt_file, args.prompt_dir)

    combined = build_combined_package_table(
        package_dir=args.path,
        reverse_config=args.reverse_config,
        prompt_dictionary=prompts,
        min_nonmissing=args.min_rows,
        max_unique=args.max_unique,
        id_col=args.id_col,
        include_regex=args.include_regex,
        exclude_regex=args.exclude_regex,
        auto_reverse=not args.no_auto_reverse,
        auto_reverse_corr_threshold=args.auto_reverse_corr_threshold,
        auto_reverse_min_pairwise_subjects=args.auto_reverse_min_pairwise_subjects,
        auto_reverse_min_pairwise_fraction=args.auto_reverse_min_pairwise_fraction,
    )
    config.reverse_items = combined.reverse_items
    result = analyze_survey_table(combined.table, config, item_columns=combined.item_columns)
    _write_result(result, args.outdir, _safe_name(result.table_name))
    _write_summary(pd.DataFrame([result.summary]), args.outdir, result.summary["embedding_slug"])
    combined.prompt_inventory.to_csv(args.outdir / "combined_prompt_inventory.csv", index=False)
    combined.source_summary.to_csv(args.outdir / "combined_source_summary.csv", index=False)
    combined.auto_reverse_warnings.to_csv(args.outdir / "combined_auto_reverse_warnings.csv", index=False)
    (args.outdir / "combined_auto_reverse_warnings.txt").write_text(
        combined.auto_reverse_warning_text,
        encoding="utf-8",
    )
    print(
        "Analyzed combined package {}: {} subjects, {} prompts, D={}.".format(
            args.path.name,
            result.summary["n_rows"],
            result.summary["n_items"],
            result.summary["optimal_d"],
        )
    )
    return 0


def _run_study(args: argparse.Namespace, parser: argparse.ArgumentParser) -> int:
    if args.prompt_file is None and args.prompt_dir is None:
        parser.error("run-study requires --prompt-file or --prompt-dir.")
    args.path = args.data_dir
    _apply_embedding_model_arg(args)
    status = _analyze_package_combined(args)
    if status != 0 or args.skip_plots:
        return status
    from survey_semantics.plotting import plot_output_directory

    plot_output_directory(
        input_dir=args.outdir,
        outdir=args.plots_outdir,
        outlier_column=args.plot_outlier_column,
        max_heatmap_rows=args.max_heatmap_rows,
        absolute_heatmap=args.absolute_heatmap,
    )
    return status


def _apply_embedding_model_arg(args: argparse.Namespace) -> None:
    model = str(args.embedding_model).strip()
    if model.lower() in {"tfidf", "tf-idf", "word-1-2gram-max1024", "auto"}:
        raise SystemExit(
            "Embedding backend {!r} is not supported. Pass a local sentence-transformers "
            "model path or cached name (e.g. a bge-m3 path); there is no TF-IDF or auto "
            "fallback.".format(model)
        )
    args.embedding = "sentence-transformers"
    args.model = model


def _write_result(result, outdir: Path, prefix: str) -> None:
    run_prefix = _safe_name("{}__{}".format(prefix, result.summary["embedding_slug"]))
    result.scores.to_csv(outdir / "{}_scores.csv".format(run_prefix), index=False)
    result.prompt_loadings.to_csv(outdir / "{}_prompt_loadings.csv".format(run_prefix), index=False)
    result.item_weights.to_csv(outdir / "{}_item_weights.csv".format(run_prefix), index=False)
    result.drivers.to_csv(outdir / "{}_drivers.csv".format(run_prefix), index=False)
    result.stability.to_csv(outdir / "{}_stability.csv".format(run_prefix), index=False)
    result.dimension_selection.to_csv(outdir / "{}_dimension_selection.csv".format(run_prefix), index=False)
    result.dimension_methods.to_csv(outdir / "{}_dimension_methods.csv".format(run_prefix), index=False)
    result.case_study_label_map.to_csv(outdir / "{}_case_study_label_map.csv".format(run_prefix), index=False)
    if not result.raw_response_umap.empty:
        result.raw_response_umap.to_csv(outdir / "{}_raw_response_umap.csv".format(run_prefix), index=False)
    if not result.semantic_pc_umap.empty:
        result.semantic_pc_umap.to_csv(outdir / "{}_semantic_pc_umap.csv".format(run_prefix), index=False)
    case_dir = outdir / "case_studies"
    case_dir.mkdir(parents=True, exist_ok=True)
    for stale in case_dir.glob("{}_*.txt".format(run_prefix)):
        stale.unlink()
    for filename, text in result.case_studies.items():
        (case_dir / "{}_{}".format(run_prefix, filename)).write_text(text, encoding="utf-8")


def _write_summary(summary: pd.DataFrame, outdir: Path, slug: str) -> None:
    summary.to_csv(outdir / "summary.csv", index=False)
    summary.to_csv(outdir / "{}_summary.csv".format(_safe_name(slug)), index=False)


def _summary_slug(summary: pd.DataFrame, config: AnalysisConfig) -> str:
    if "embedding_slug" in summary.columns:
        slugs = [slug for slug in summary["embedding_slug"].dropna().unique() if str(slug)]
        if len(slugs) == 1:
            return str(slugs[0])
    return _requested_embedding_slug(config)


def _requested_embedding_slug(config: AnalysisConfig) -> str:
    model_name = config.model_name or ("BAAI/bge-m3" if config.embedding == "sentence-transformers" else None)
    return embedding_slug(config.embedding, model_name)


def _safe_name(name: str) -> str:
    safe = re.sub(r"[^A-Za-z0-9_.-]+", "_", str(name)).strip("_")
    return safe or "survey"


if __name__ == "__main__":
    raise SystemExit(main())
```
