## Supplementary material for "Auditable cross-instrument detection of unusual multivariate psychiatric response configurations using a semantically aligned covariance subspace": Code for tool: combined.py

```
"""Build combined, transdiagnostic survey matrices across questionnaire files."""

import csv
from dataclasses import dataclass
import math
from pathlib import Path
from typing import Dict, Iterable, List, Mapping, Optional, Sequence, Set, Tuple

import pandas as pd

from survey_semantics.io import (
    SurveyTable,
    coerce_response_frame,
    default_covariates,
    default_id_column,
    infer_item_columns,
    read_survey_table,
)


@dataclass
class CombinedPackage:
    table: SurveyTable
    item_columns: List[str]
    reverse_items: Set[str]
    prompt_inventory: pd.DataFrame
    source_summary: pd.DataFrame
    manual_reverse_items: Set[str]
    auto_reverse_items: Set[str]
    auto_reverse_warnings: pd.DataFrame
    auto_reverse_warning_text: str


@dataclass
class AutoReverseResult:
    reverse_items: Set[str]
    warnings: pd.DataFrame


AUTO_REVERSE_WARNING_COLUMNS = [
    "table",
    "action",
    "orientation_rule",
    "item_kept",
    "feature_kept",
    "prompt_kept",
    "item_negated",
    "feature_negated",
    "prompt_negated",
    "correlation",
    "pairwise_subjects",
    "pairwise_subject_fraction",
    "required_pairwise_subjects",
    "threshold",
    "notes",
]


def build_combined_package_table(
    package_dir: Path,
    reverse_config: Optional[Path] = None,
    prompt_dictionary: Optional[Mapping[str, str]] = None,
    min_nonmissing: int = 10,
    max_unique: int = 8,
    id_col: Optional[str] = None,
    include_regex: Optional[str] = None,
    exclude_regex: Optional[str] = None,
    auto_reverse: bool = True,
    auto_reverse_corr_threshold: float = 0.70,
    auto_reverse_min_pairwise_subjects: int = 10,
    auto_reverse_min_pairwise_fraction: float = 0.50,
) -> CombinedPackage:
    """Create one subject-by-all-items table for a package directory."""

    import re

    package_dir = Path(package_dir)
    include = re.compile(include_regex) if include_regex else None
    exclude = re.compile(exclude_regex) if exclude_regex else None
    manual_reverse_items = read_reverse_config(reverse_config)
    auto_reverse_items: Set[str] = set()

    response_frames = []
    covariate_frames = []
    inventory_rows = []
    summary_rows = []
    warning_frames = []

    files = sorted(list(package_dir.glob("*.txt")) + list(package_dir.glob("*.tsv")) + list(package_dir.glob("*.csv")))
    for path in files:
        if include and not include.search(path.name):
            continue
        if exclude and exclude.search(path.name):
            continue

        try:
            table = read_survey_table(path, prompt_dictionary=prompt_dictionary)
            subject_col = id_col or default_id_column(table.data)
            if not subject_col or subject_col not in table.data.columns:
                raise ValueError("No subject identifier column found.")
            item_cols = infer_item_columns(
                table,
                min_nonmissing=min_nonmissing,
                max_unique=max_unique,
            )
            if not item_cols:
                raise ValueError("No usable item columns found.")

            responses = coerce_response_frame(table.data, item_cols)
            responses[subject_col] = table.data[subject_col].values
            grouped = responses.dropna(subset=[subject_col]).groupby(subject_col)[item_cols].mean()
            feature_names = {col: feature_name(table.name, col) for col in item_cols}

            if auto_reverse:
                inferred = infer_auto_reverse_items(
                    table_name=table.name,
                    responses=grouped,
                    item_columns=item_cols,
                    dictionary=table.dictionary,
                    feature_names=feature_names,
                    manual_reverse_items=manual_reverse_items,
                    corr_threshold=auto_reverse_corr_threshold,
                    min_pairwise_subjects=auto_reverse_min_pairwise_subjects,
                    min_pairwise_fraction=auto_reverse_min_pairwise_fraction,
                )
                auto_reverse_items.update(inferred.reverse_items)
                if not inferred.warnings.empty:
                    warning_frames.append(inferred.warnings)

            grouped = grouped.rename(columns=feature_names)
            response_frames.append(grouped)

            covariates = standardize_covariates(table.data, subject_col)
            if len(covariates.columns) > 1:
                covariate_frames.append(covariates)

            for col in item_cols:
                feature = feature_names[col]
                reverse_source = reverse_source_label(
                    feature in manual_reverse_items,
                    feature in auto_reverse_items,
                )
                inventory_rows.append(
                    {
                        "table": table.name,
                        "item": col,
                        "feature": feature,
                        "prompt": table.dictionary.get(col, col),
                        "reverse_scored": bool(reverse_source),
                        "reverse_source": reverse_source,
                    }
                )
            summary_rows.append(
                {
                    "table": table.name,
                    "path": str(path),
                    "status": "included",
                    "n_subjects": int(len(grouped)),
                    "n_items": int(len(item_cols)),
                    "reason": "",
                }
            )
        except Exception as exc:
            summary_rows.append(
                {
                    "table": path.stem,
                    "path": str(path),
                    "status": "skipped",
                    "n_subjects": 0,
                    "n_items": 0,
                    "reason": str(exc),
                }
            )

    if not response_frames:
        raise ValueError("No questionnaire files with usable item responses were found.")

    combined = pd.concat(response_frames, axis=1, join="outer")
    combined.index.name = "subjectkey"
    combined = combined.reset_index()

    if covariate_frames:
        covariates = pd.concat(covariate_frames, axis=0, ignore_index=True)
        aggregations = {}
        if "interview_age" in covariates.columns:
            aggregations["interview_age"] = mean_numeric
        if "sex" in covariates.columns:
            aggregations["sex"] = first_nonmissing
        covariates = covariates.groupby("subjectkey").agg(aggregations).reset_index()
        combined = combined.merge(covariates, on="subjectkey", how="left")

    inventory = pd.DataFrame(inventory_rows)
    reverse_items = set(manual_reverse_items) | set(auto_reverse_items)
    auto_warnings = combine_warning_frames(warning_frames)
    dictionary = dict(zip(inventory["feature"], inventory["prompt"]))
    table = SurveyTable(
        name="{}_combined".format(package_dir.name),
        path=package_dir,
        data=combined,
        dictionary=dictionary,
    )
    return CombinedPackage(
        table=table,
        item_columns=list(inventory["feature"]),
        reverse_items=reverse_items,
        prompt_inventory=inventory,
        source_summary=pd.DataFrame(summary_rows),
        manual_reverse_items=manual_reverse_items,
        auto_reverse_items=auto_reverse_items,
        auto_reverse_warnings=auto_warnings,
        auto_reverse_warning_text=format_auto_reverse_warning_text(auto_warnings),
    )


def infer_auto_reverse_items(
    table_name: str,
    responses: pd.DataFrame,
    item_columns: Sequence[str],
    dictionary: Dict[str, str],
    feature_names: Dict[str, str],
    manual_reverse_items: Set[str],
    corr_threshold: float = 0.70,
    min_pairwise_subjects: int = 10,
    min_pairwise_fraction: float = 0.50,
) -> AutoReverseResult:
    """Infer likely reverse-scored items from strong within-instrument anticorrelation.

    Anticorrelation identifies opposite polarity, not absolute construct
    direction. The orientation heuristic is intentionally auditable:
    manual reverse config wins when present; otherwise the smaller polarity
    group is negated, with one-vs-one ties keeping the earlier item.
    """

    threshold = abs(float(corr_threshold))
    if threshold <= 0 or threshold >= 1:
        raise ValueError("auto_reverse_corr_threshold must be between 0 and 1.")

    n_subjects = int(len(responses))
    required = max(
        int(min_pairwise_subjects),
        int(math.ceil(n_subjects * float(min_pairwise_fraction))),
    )
    if len(item_columns) < 2 or n_subjects < required:
        return AutoReverseResult(set(), empty_warning_frame())

    pair_records = strong_correlation_pairs(
        responses=responses,
        item_columns=item_columns,
        threshold=threshold,
        required_pairwise_subjects=required,
        n_subjects=n_subjects,
    )
    negative_pairs = [record for record in pair_records if record["sign"] < 0]
    if not negative_pairs:
        return AutoReverseResult(set(), empty_warning_frame())

    graph = build_signed_graph(pair_records)
    components = signed_components(item_columns, graph)

    reverse_raw: Set[str] = set()
    warning_rows = []
    for component in components:
        component_items = component["items"]
        component_negative_pairs = [
            record for record in negative_pairs
            if record["left"] in component_items and record["right"] in component_items
        ]
        if not component_negative_pairs:
            continue

        signs = component["signs"]
        conflicts = component["conflicts"]
        if conflicts:
            warning_rows.extend(
                conflict_warning_rows(
                    table_name,
                    component_negative_pairs,
                    dictionary,
                    feature_names,
                    threshold,
                    required,
                    "inconsistent strong-correlation graph; no automatic negation applied for this component",
                )
            )
            continue

        reverse_sign, orientation_rule, notes = choose_reverse_sign(
            component_items=component_items,
            signs=signs,
            item_order=item_columns,
            feature_names=feature_names,
            manual_reverse_items=manual_reverse_items,
        )
        component_reversed = {item for item in component_items if signs[item] == reverse_sign}
        reverse_raw.update(component_reversed)

        for record in component_negative_pairs:
            left = record["left"]
            right = record["right"]
            if left in component_reversed and right not in component_reversed:
                negated = left
                kept = right
            elif right in component_reversed and left not in component_reversed:
                negated = right
                kept = left
            else:
                warning_rows.append(
                    auto_reverse_warning_row(
                        table_name=table_name,
                        action="ambiguous_component_pair",
                        orientation_rule=orientation_rule,
                        kept=left,
                        negated=right,
                        dictionary=dictionary,
                        feature_names=feature_names,
                        correlation=record["correlation"],
                        pairwise_subjects=record["pairwise_subjects"],
                        pairwise_fraction=record["pairwise_subject_fraction"],
                        required_pairwise_subjects=required,
                        threshold=threshold,
                        notes="negative pair did not split cleanly across inferred polarity groups; review manually",
                    )
                )
                continue

            warning_rows.append(
                auto_reverse_warning_row(
                    table_name=table_name,
                    action="negated",
                    orientation_rule=orientation_rule,
                    kept=kept,
                    negated=negated,
                    dictionary=dictionary,
                    feature_names=feature_names,
                    correlation=record["correlation"],
                    pairwise_subjects=record["pairwise_subjects"],
                    pairwise_fraction=record["pairwise_subject_fraction"],
                    required_pairwise_subjects=required,
                    threshold=threshold,
                    notes=notes,
                )
            )

    reverse_features = {feature_names[item] for item in reverse_raw}
    return AutoReverseResult(reverse_features, pd.DataFrame(warning_rows, columns=AUTO_REVERSE_WARNING_COLUMNS))


def strong_correlation_pairs(
    responses: pd.DataFrame,
    item_columns: Sequence[str],
    threshold: float,
    required_pairwise_subjects: int,
    n_subjects: int,
) -> List[Dict[str, object]]:
    records: List[Dict[str, object]] = []
    for left_idx, left in enumerate(item_columns):
        for right in item_columns[left_idx + 1:]:
            pair = responses[[left, right]].dropna()
            pairwise_subjects = int(len(pair))
            if pairwise_subjects < required_pairwise_subjects:
                continue
            if pair[left].nunique(dropna=True) < 2 or pair[right].nunique(dropna=True) < 2:
                continue
            correlation = pair[left].astype(float).corr(pair[right].astype(float))
            if pd.isna(correlation):
                continue
            if abs(float(correlation)) < threshold:
                continue
            records.append(
                {
                    "left": left,
                    "right": right,
                    "correlation": float(correlation),
                    "sign": 1 if correlation >= 0 else -1,
                    "pairwise_subjects": pairwise_subjects,
                    "pairwise_subject_fraction": (
                        float(pairwise_subjects) / float(n_subjects) if n_subjects else 0.0
                    ),
                }
            )
    return records


def build_signed_graph(pair_records: Sequence[Dict[str, object]]) -> Dict[str, List[Tuple[str, int]]]:
    graph: Dict[str, List[Tuple[str, int]]] = {}
    for record in pair_records:
        left = str(record["left"])
        right = str(record["right"])
        sign = int(record["sign"])
        graph.setdefault(left, []).append((right, sign))
        graph.setdefault(right, []).append((left, sign))
    return graph


def signed_components(
    item_columns: Sequence[str],
    graph: Dict[str, List[Tuple[str, int]]],
) -> List[Dict[str, object]]:
    components = []
    visited: Set[str] = set()
    for start in item_columns:
        if start in visited or start not in graph:
            continue
        queue = [start]
        signs = {start: 1}
        items: Set[str] = set()
        conflicts: List[Tuple[str, str]] = []
        visited.add(start)

        while queue:
            current = queue.pop(0)
            items.add(current)
            current_sign = signs[current]
            for neighbor, edge_sign in graph.get(current, []):
                expected_sign = current_sign * edge_sign
                if neighbor not in signs:
                    signs[neighbor] = expected_sign
                elif signs[neighbor] != expected_sign:
                    conflicts.append((current, neighbor))
                if neighbor not in visited:
                    visited.add(neighbor)
                    queue.append(neighbor)

        components.append({"items": items, "signs": signs, "conflicts": conflicts})
    return components


def choose_reverse_sign(
    component_items: Set[str],
    signs: Dict[str, int],
    item_order: Sequence[str],
    feature_names: Dict[str, str],
    manual_reverse_items: Set[str],
) -> Tuple[int, str, str]:
    positive_items = [item for item in item_order if item in component_items and signs[item] > 0]
    negative_items = [item for item in item_order if item in component_items and signs[item] < 0]
    manual_positive = sum(1 for item in positive_items if feature_names[item] in manual_reverse_items)
    manual_negative = sum(1 for item in negative_items if feature_names[item] in manual_reverse_items)

    if manual_positive != manual_negative:
        if manual_positive > manual_negative:
            return 1, "manual_reverse_config", "manual reverse config oriented this polarity component"
        return -1, "manual_reverse_config", "manual reverse config oriented this polarity component"

    if len(positive_items) != len(negative_items):
        if len(positive_items) < len(negative_items):
            return 1, "smaller_polarity_group", "the smaller inferred polarity group was negated"
        return -1, "smaller_polarity_group", "the smaller inferred polarity group was negated"

    first_item = next(item for item in item_order if item in component_items)
    reverse_sign = -signs[first_item]
    return (
        reverse_sign,
        "tie_keep_first_item",
        "equal-size polarity groups; kept the earlier item as the anchor and negated the other side",
    )


def auto_reverse_warning_row(
    table_name: str,
    action: str,
    orientation_rule: str,
    kept: str,
    negated: str,
    dictionary: Dict[str, str],
    feature_names: Dict[str, str],
    correlation: float,
    pairwise_subjects: int,
    pairwise_fraction: float,
    required_pairwise_subjects: int,
    threshold: float,
    notes: str,
) -> Dict[str, object]:
    return {
        "table": table_name,
        "action": action,
        "orientation_rule": orientation_rule,
        "item_kept": kept,
        "feature_kept": feature_names.get(kept, feature_name(table_name, kept)),
        "prompt_kept": dictionary.get(kept, kept),
        "item_negated": negated,
        "feature_negated": feature_names.get(negated, feature_name(table_name, negated)),
        "prompt_negated": dictionary.get(negated, negated),
        "correlation": float(correlation),
        "pairwise_subjects": int(pairwise_subjects),
        "pairwise_subject_fraction": float(pairwise_fraction),
        "required_pairwise_subjects": int(required_pairwise_subjects),
        "threshold": float(threshold),
        "notes": notes,
    }


def conflict_warning_rows(
    table_name: str,
    negative_pairs: Sequence[Dict[str, object]],
    dictionary: Dict[str, str],
    feature_names: Dict[str, str],
    threshold: float,
    required_pairwise_subjects: int,
    notes: str,
) -> List[Dict[str, object]]:
    rows = []
    for record in negative_pairs:
        rows.append(
            auto_reverse_warning_row(
                table_name=table_name,
                action="not_negated_conflict",
                orientation_rule="conflict",
                kept=str(record["left"]),
                negated=str(record["right"]),
                dictionary=dictionary,
                feature_names=feature_names,
                correlation=float(record["correlation"]),
                pairwise_subjects=int(record["pairwise_subjects"]),
                pairwise_fraction=float(record["pairwise_subject_fraction"]),
                required_pairwise_subjects=required_pairwise_subjects,
                threshold=threshold,
                notes=notes,
            )
        )
    return rows


def reverse_source_label(manual: bool, auto: bool) -> str:
    if manual and auto:
        return "manual+auto_anticorrelation"
    if manual:
        return "manual"
    if auto:
        return "auto_anticorrelation"
    return ""


def empty_warning_frame() -> pd.DataFrame:
    return pd.DataFrame(columns=AUTO_REVERSE_WARNING_COLUMNS)


def combine_warning_frames(frames: Sequence[pd.DataFrame]) -> pd.DataFrame:
    if not frames:
        return empty_warning_frame()
    combined = pd.concat(frames, axis=0, ignore_index=True)
    for col in AUTO_REVERSE_WARNING_COLUMNS:
        if col not in combined.columns:
            combined[col] = pd.NA
    return combined[AUTO_REVERSE_WARNING_COLUMNS]


def format_auto_reverse_warning_text(warnings: pd.DataFrame) -> str:
    lines = [
        "Automatic reverse-scoring warning report",
        "",
        "Strong within-instrument anticorrelation was interpreted as evidence that one prompt is oppositely scored.",
        "This is a deterministic response-pattern heuristic, not semantic validation; review these items manually.",
        "",
    ]
    if warnings.empty:
        lines.append("No automatic reverse-scoring warnings were triggered.")
        return "\n".join(lines) + "\n"

    for table_name, table_rows in warnings.groupby("table", sort=True):
        lines.append("Instrument: {}".format(table_name))
        for _, row in table_rows.iterrows():
            action = str(row.get("action", ""))
            correlation = float(row.get("correlation", float("nan")))
            n_pairwise = int(row.get("pairwise_subjects", 0))
            fraction = float(row.get("pairwise_subject_fraction", 0.0))
            lines.append(
                "  - Action: {}; corr={:.3f}; paired subjects={} ({:.1%}); rule={}".format(
                    action,
                    correlation,
                    n_pairwise,
                    fraction,
                    row.get("orientation_rule", ""),
                )
            )
            lines.append(
                "    Kept: {} | {}".format(
                    row.get("feature_kept", ""),
                    row.get("prompt_kept", ""),
                )
            )
            lines.append(
                "    Negated: {} | {}".format(
                    row.get("feature_negated", ""),
                    row.get("prompt_negated", ""),
                )
            )
            notes = str(row.get("notes", "") or "")
            if notes:
                lines.append("    Notes: {}".format(notes))
        lines.append("")

    return "\n".join(lines).rstrip() + "\n"


def read_reverse_config(path: Optional[Path]) -> Set[str]:
    """Read manual reverse-scoring config.

    Accepted CSV columns:
      - feature, reverse
      - table, item, reverse
      - item, reverse, where item is already a combined feature name
    """

    if not path:
        return set()
    path = Path(path)
    reverse_items: Set[str] = set()
    with open(str(path), newline="", encoding="utf-8-sig") as handle:
        reader = csv.DictReader(handle)
        for row in reader:
            if not truthy(row.get("reverse") or row.get("reverse_scored") or row.get("reversed")):
                continue
            feature = (row.get("feature") or "").strip()
            table = (row.get("table") or "").strip()
            item = (row.get("item") or "").strip()
            if not feature and table and item:
                feature = feature_name(table, item)
            elif not feature and item:
                feature = item
            if feature:
                reverse_items.add(feature)
    return reverse_items


def feature_name(table: str, item: str) -> str:
    return "{}__{}".format(table, item)


def first_nonmissing(values: pd.Series):
    observed = values.dropna()
    if observed.empty:
        return pd.NA
    return observed.iloc[0]


def mean_numeric(values: pd.Series):
    numeric = pd.to_numeric(values, errors="coerce")
    if numeric.dropna().empty:
        return pd.NA
    return numeric.mean()


def standardize_covariates(df: pd.DataFrame, subject_col: str) -> pd.DataFrame:
    cols = [subject_col]
    output = pd.DataFrame({subject_col: df[subject_col].values})
    if "interview_age" in df.columns:
        output["interview_age"] = df["interview_age"].values
    elif "age" in df.columns:
        output["interview_age"] = df["age"].values
    if "sex" in df.columns:
        output["sex"] = df["sex"].values
    elif "gender" in df.columns:
        output["sex"] = df["gender"].values

    if len(output.columns) == 1:
        return output
    output = output.dropna(subset=[subject_col]).rename(columns={subject_col: "subjectkey"})
    return output


def truthy(value: object) -> bool:
    return str(value).strip().lower() in {"1", "true", "t", "yes", "y", "reverse", "reversed"}
```
