## Supplementary material for "Auditable cross-instrument detection of unusual multivariate psychiatric response configurations using a semantically aligned covariance subspace": Code for tool: embedding.py

```
"""Text embedding backends for item wording."""

import os
import re
import socket
from contextlib import contextmanager
from dataclasses import dataclass
from pathlib import Path
from typing import List, Mapping, Optional, Sequence

import numpy as np


@dataclass
class EmbeddingResult:
    vectors: np.ndarray
    backend: str
    model_name: str
    requested_backend: str
    requested_model_name: str
    slug: str


@dataclass
class ItemEmbeddings:
    """A reusable, response-independent embedding of survey items.

    This is the artifact produced by the LLM step and consumed by the downstream
    PCA/analysis. It carries the item names alongside the vectors so the
    analysis can align them to its own item columns, and the embedding
    provenance (backend/model/slug) for output naming.
    """

    items: List[str]
    vectors: np.ndarray            # shape (len(items), dim)
    backend: str
    model_name: str
    slug: str

    def matrix_for(self, item_columns: Sequence[str]) -> np.ndarray:
        """Return the embedding rows for `item_columns`, in that order.

        Raises if any requested item has no precomputed embedding — never
        silently drops or substitutes items.
        """
        index = {item: i for i, item in enumerate(self.items)}
        missing = [c for c in item_columns if c not in index]
        if missing:
            raise ValueError(
                "Precomputed embeddings are missing {} item(s): {}".format(
                    len(missing), ", ".join(map(str, missing[:10]))
                )
            )
        return self.vectors[[index[c] for c in item_columns]]


def embed_item_prompts(
    prompts: Mapping[str, str],
    method: str = "sentence-transformers",
    model_name: Optional[str] = None,
    trust_remote_code: bool = False,
) -> ItemEmbeddings:
    """Embed an item→wording mapping into a reusable :class:`ItemEmbeddings`."""

    items = [str(item) for item in prompts.keys()]
    texts = [str(prompts[item]) for item in prompts.keys()]
    result = embed_texts_with_metadata(
        texts, method=method, model_name=model_name,
        trust_remote_code=trust_remote_code,
    )
    return ItemEmbeddings(
        items=items,
        vectors=result.vectors,
        backend=result.backend,
        model_name=result.model_name,
        slug=result.slug,
    )


def save_item_embeddings(path: Path, embeddings: ItemEmbeddings) -> None:
    """Persist item embeddings to a `.npz` file (no pickle; portable)."""

    np.savez(
        Path(path),
        items=np.array(embeddings.items),
        vectors=np.asarray(embeddings.vectors, dtype=float),
        meta=np.array([embeddings.backend, embeddings.model_name, embeddings.slug]),
    )


def load_item_embeddings(path: Path) -> ItemEmbeddings:
    """Load item embeddings written by :func:`save_item_embeddings`."""

    data = np.load(Path(path))
    backend, model_name, slug = (str(x) for x in data["meta"])
    return ItemEmbeddings(
        items=[str(x) for x in data["items"]],
        vectors=np.asarray(data["vectors"], dtype=float),
        backend=backend,
        model_name=model_name,
        slug=slug,
    )


OFFLINE_ENV = {
    "HF_HUB_OFFLINE": "1",
    "TRANSFORMERS_OFFLINE": "1",
    "HF_DATASETS_OFFLINE": "1",
    "HF_HUB_DISABLE_TELEMETRY": "1",
    "DISABLE_TELEMETRY": "1",
    "TOKENIZERS_PARALLELISM": "false",
}

_SOCKET_BLOCK_INSTALLED = False


def embed_texts(
    texts: Sequence[str],
    method: str = "sentence-transformers",
    model_name: Optional[str] = None,
) -> np.ndarray:
    """Embed item text with a local sentence-transformers model (e.g. bge-m3)."""

    return embed_texts_with_metadata(texts, method=method, model_name=model_name).vectors


def embed_texts_with_metadata(
    texts: Sequence[str],
    method: str = "sentence-transformers",
    model_name: Optional[str] = None,
    trust_remote_code: bool = False,
) -> EmbeddingResult:
    """Embed item text and return provenance for output naming.

    The only supported backend is a local ``sentence-transformers`` model (e.g.
    ``BAAI/bge-m3``). There is **no** automatic fallback: if the requested backend
    is unknown, or the model cannot be loaded locally, this raises rather than
    silently substituting a different (unvalidated) embedding method.
    """

    method = (method or "sentence-transformers").lower()
    if method not in {"sentence-transformers", "sentence_transformers"}:
        raise ValueError(
            "Unsupported embedding backend {!r}. The only supported backend is "
            "'sentence-transformers' (a local sentence encoder such as BAAI/bge-m3). "
            "There is no automatic fallback.".format(method)
        )

    requested_model = model_name or ""
    resolved_model = model_name or "BAAI/bge-m3"
    vectors = _sentence_transformer_embeddings(
        texts, resolved_model, trust_remote_code=trust_remote_code
    )
    backend = "sentence-transformers"
    return EmbeddingResult(
        vectors=vectors,
        backend=backend,
        model_name=resolved_model,
        requested_backend="sentence-transformers",
        requested_model_name=requested_model,
        slug=embedding_slug(backend, resolved_model),
    )


def embedding_slug(backend: str, model_name: Optional[str] = None) -> str:
    label = backend if not model_name else "{}__{}".format(backend, model_name)
    label = label.replace("/", "_").replace("\\", "_")
    label = re.sub(r"[^A-Za-z0-9_.-]+", "_", label).strip("_")
    return label or "embedding"


def _sentence_transformer_embeddings(
    texts: Sequence[str],
    model_name: Optional[str] = None,
    trust_remote_code: bool = False,
) -> np.ndarray:
    """Encode texts with a local sentence-transformers model.

    ``trust_remote_code`` is an explicit opt-in for models that ship custom
    architecture code in their repo (e.g. ``gte-large-en-v1.5``). The code runs
    from the already-downloaded local files and the outbound-socket blocker
    stays active, so nothing can be fetched at load/encode time either way.
    """
    enforce_local_ai_offline_policy()
    with block_outbound_sockets():
        from sentence_transformers import SentenceTransformer

        try:
            model = SentenceTransformer(
                model_name or "BAAI/bge-m3",
                local_files_only=True,
                trust_remote_code=trust_remote_code,
            )
        except TypeError as exc:
            raise RuntimeError(
                "This sentence-transformers version does not expose local_files_only=True. "
                "Refusing to load the model because the pipeline is configured to prohibit "
                "all Hugging Face/network callbacks."
            ) from exc

        vectors = model.encode(list(texts), normalize_embeddings=True)
    return np.asarray(vectors, dtype=float)


def enforce_local_ai_offline_policy() -> None:
    """Set offline/telemetry-disable environment variables before HF imports."""

    for key, value in OFFLINE_ENV.items():
        os.environ[key] = value


def install_outbound_socket_blocker() -> None:
    """Install a process-wide outbound network blocker."""

    global _SOCKET_BLOCK_INSTALLED
    if _SOCKET_BLOCK_INSTALLED:
        return

    def blocked_connect(self, address):
        raise RuntimeError(
            "Outbound network access is disabled for local survey analysis. "
            "Use local files and locally cached/local embedding models only."
        )

    def blocked_connect_ex(self, address):
        raise RuntimeError(
            "Outbound network access is disabled for local survey analysis. "
            "Use local files and locally cached/local embedding models only."
        )

    def blocked_create_connection(*args, **kwargs):
        raise RuntimeError(
            "Outbound network access is disabled for local survey analysis. "
            "Use local files and locally cached/local embedding models only."
        )

    socket.socket.connect = blocked_connect
    socket.socket.connect_ex = blocked_connect_ex
    socket.create_connection = blocked_create_connection
    _SOCKET_BLOCK_INSTALLED = True


@contextmanager
def block_outbound_sockets():
    """Block outbound socket connections while local embedding code runs."""

    original_connect = socket.socket.connect
    original_connect_ex = socket.socket.connect_ex
    original_create_connection = socket.create_connection

    def blocked_connect(self, address):
        raise RuntimeError(
            "Outbound network access is disabled for local survey embedding. "
            "Use a model that is already present on local disk or in the local HF cache."
        )

    def blocked_create_connection(*args, **kwargs):
        raise RuntimeError(
            "Outbound network access is disabled for local survey embedding. "
            "Use a model that is already present on local disk or in the local HF cache."
        )

    socket.socket.connect = blocked_connect
    socket.socket.connect_ex = blocked_connect
    socket.create_connection = blocked_create_connection
    try:
        yield
    finally:
        socket.socket.connect = original_connect
        socket.socket.connect_ex = original_connect_ex
        socket.create_connection = original_create_connection
```
