## Supplementary material for "Auditable cross-instrument detection of unusual multivariate psychiatric response configurations using a semantically aligned covariance subspace": Code for tool: io.py

```
"""Input helpers for generic and NDA-style survey tables."""

import csv
import re
from dataclasses import dataclass
from pathlib import Path
from typing import Dict, Iterable, List, Mapping, Optional, Sequence, Tuple

import numpy as np
import pandas as pd


MISSING_STRINGS = {
    "",
    ".",
    "na",
    "n/a",
    "nan",
    "none",
    "null",
    "missing",
    "not applicable",
    "not available",
}

DEFAULT_SENTINELS = {-9999, -999, -99, -9, -4, -3, -2, -1, 777, 888, 999, 9999}

YES_NO_MAP = {
    "yes": 1.0,
    "y": 1.0,
    "true": 1.0,
    "t": 1.0,
    "no": 0.0,
    "n": 0.0,
    "false": 0.0,
    "f": 0.0,
}

METADATA_NAME_RE = re.compile(
    r"(^|_)(id|guid|subject|date|time|age|sex|gender|visit|site|group|comments?|"
    r"respond(ent|ent_detail)?|informant|relationship|collection|dataset|"
    r"phenotype|diagnosis|race|ethnicity|version|validity|missing|calc_status)($|_)",
    re.IGNORECASE,
)

AGGREGATE_NAME_RE = re.compile(
    r"(^|_)(total|subtotal|score|rawscore|raw_score|tscore|t_score|mean|"
    r"subscale|num_endr|endorsed|result|status|adjusted|adj|standard)($|_)",
    re.IGNORECASE,
)

AGGREGATE_DESC_RE = re.compile(
    r"\b(total|subtotal|score|t-score|raw score|standard score|subscale|"
    r"number of .*endorsed|calculated|calculation status|result)\b",
    re.IGNORECASE,
)

METADATA_DESC_RE = re.compile(
    r"\b(global unique identifier|subject id|date on which|age in months|"
    r"sex of subject|comments?|respondent|informant|collection title|visit name|"
    r"phenotype|diagnosis|race|validity|missing values?)\b",
    re.IGNORECASE,
)


@dataclass
class SurveyTable:
    """A loaded survey table plus item wording metadata."""

    name: str
    path: Path
    data: pd.DataFrame
    dictionary: Dict[str, str]


def guess_delimiter(path: Path) -> str:
    if path.suffix.lower() == ".csv":
        return ","
    return "\t"


def read_survey_table(
    path: Path,
    prompt_dictionary: Optional[Mapping[str, str]] = None,
) -> SurveyTable:
    """Read a generic CSV/TSV or NDA-style two-header-row table.

    NDA package files usually store variable names in row 1 and human-readable
    variable descriptions in row 2. Generic files without a dictionary row are
    also supported; column names become item text fallback.
    """

    path = Path(path)
    sep = guess_delimiter(path)
    header, description = _peek_header_rows(path, sep)
    has_dictionary = _looks_like_dictionary_row(header, description)

    skiprows = [1] if has_dictionary else None
    data = pd.read_csv(
        str(path),
        sep=sep,
        skiprows=skiprows,
        dtype=str,
        low_memory=False,
    )
    data.columns = [str(c).strip().strip('"') for c in data.columns]

    if has_dictionary:
        dictionary = {
            str(col).strip().strip('"'): str(desc).strip().strip('"')
            for col, desc in zip(header, description)
        }
    else:
        dictionary = {str(col): str(col) for col in data.columns}

    if prompt_dictionary:
        from survey_semantics.prompts import apply_prompt_dictionary

        dictionary = apply_prompt_dictionary(dictionary, path.stem, prompt_dictionary)

    return SurveyTable(name=path.stem, path=path, data=data, dictionary=dictionary)


def _peek_header_rows(path: Path, sep: str) -> Tuple[List[str], List[str]]:
    with open(str(path), "r", newline="", encoding="utf-8-sig") as handle:
        reader = csv.reader(handle, delimiter=sep)
        try:
            header = next(reader)
        except StopIteration:
            return [], []
        try:
            description = next(reader)
        except StopIteration:
            description = []
    return header, description


def _looks_like_dictionary_row(header: Sequence[str], row: Sequence[str]) -> bool:
    if not header or not row or len(header) != len(row):
        return False
    informative = 0
    for col, value in zip(header, row):
        value = str(value).strip()
        col = str(col).strip()
        if not value or value == col:
            continue
        if " " in value or len(value) > len(col) + 8:
            informative += 1
    return informative >= max(2, int(0.10 * len(header)))


def coerce_response_series(
    series: pd.Series,
    sentinels: Optional[Iterable[int]] = None,
) -> pd.Series:
    """Coerce a response column to numeric values, preserving Likert structure."""

    sentinels = DEFAULT_SENTINELS if sentinels is None else set(sentinels)
    clean = series.astype(str).str.strip().str.strip('"')
    lower = clean.str.lower()
    lower = lower.where(~lower.isin(MISSING_STRINGS), np.nan)

    observed = set(lower.dropna().unique())
    if observed and observed.issubset(set(YES_NO_MAP.keys())):
        numeric = lower.map(YES_NO_MAP)
    else:
        numeric = pd.to_numeric(lower, errors="coerce")

    numeric = numeric.astype(float)
    numeric = numeric.where(~numeric.isin(list(sentinels)), np.nan)
    return numeric


def coerce_response_frame(
    df: pd.DataFrame,
    columns: Sequence[str],
    sentinels: Optional[Iterable[int]] = None,
) -> pd.DataFrame:
    """Coerce response columns to numeric.

    ``sentinels`` may be a single iterable applied to every column (global), or
    a ``Mapping[column -> set]`` for per-item sentinel codes (each item's own
    missing-value codes). Columns absent from the mapping fall back to the
    default sentinels.
    """

    per_column = isinstance(sentinels, Mapping)
    coerced = {}
    for col in columns:
        col_sentinels = sentinels.get(col) if per_column else sentinels
        coerced[col] = coerce_response_series(df[col], sentinels=col_sentinels)
    return pd.DataFrame(coerced, index=df.index)


def load_weights_file(path: Path) -> np.ndarray:
    """Load a one-weight-per-subject file, row-aligned to the response table.

    A single numeric column with an optional ``weight`` header. The values are
    returned in file order; alignment to the response rows is positional, so the
    caller must verify the length matches the raw response table.
    """

    path = Path(path)
    raw = pd.read_csv(path, header=None)
    column = raw.iloc[:, 0]
    if raw.shape[1] != 1:
        # Multi-column: require a labeled 'weight' column (header row present).
        header = {str(v).strip().lower(): i for i, v in enumerate(raw.iloc[0])}
        if "weight" not in header:
            raise ValueError(
                "Weights file {} must have one column or a labeled 'weight' column.".format(path)
            )
        column = raw.iloc[1:, header["weight"]]
    else:
        # Drop a leading non-numeric header cell (e.g. 'weight') if present.
        if pd.to_numeric(pd.Series([column.iloc[0]]), errors="coerce").isna().iloc[0]:
            column = column.iloc[1:]
    return pd.to_numeric(column, errors="coerce").to_numpy(dtype=float)


def infer_item_columns(
    table: SurveyTable,
    min_nonmissing: int = 10,
    max_unique: int = 8,
    min_variance: float = 1e-9,
    sentinels: Optional[Iterable[int]] = None,
) -> List[str]:
    """Infer item-response columns from a survey table.

    The heuristic is intentionally conservative: it keeps low-cardinality
    numeric or yes/no response columns and excludes identifiers, demographics,
    scores, totals, subscales, and other derived fields.
    """

    items = []
    for col in table.data.columns:
        desc = table.dictionary.get(col, "")
        if _is_metadata_or_aggregate(col, desc):
            continue

        numeric = coerce_response_series(table.data[col], sentinels=sentinels)
        observed = numeric.dropna()
        if len(observed) < min_nonmissing:
            continue
        if observed.nunique() < 2:
            continue
        if observed.nunique() > max_unique:
            continue
        if float(observed.var()) <= min_variance:
            continue
        items.append(col)
    return items


def _is_metadata_or_aggregate(name: str, description: str) -> bool:
    normalized_name = str(name).strip().lower()
    normalized_desc = str(description).strip().lower()

    if METADATA_NAME_RE.search(normalized_name):
        return True
    if AGGREGATE_NAME_RE.search(normalized_name):
        return True
    if METADATA_DESC_RE.search(normalized_desc):
        return True
    if AGGREGATE_DESC_RE.search(normalized_desc):
        return True
    return False


def default_id_column(df: pd.DataFrame) -> Optional[str]:
    for col in ["subjectkey", "src_subject_id", "export_id", "participant_id", "id"]:
        if col in df.columns:
            return col
    return None


def default_covariates(df: pd.DataFrame) -> List[str]:
    candidates = ["interview_age", "age", "gender_num", "gender", "sex"]
    return [col for col in candidates if col in df.columns]


def build_covariate_matrix(df: pd.DataFrame, columns: Sequence[str]) -> Tuple[np.ndarray, List[str]]:
    """Build a numeric covariate matrix, dropping unusable covariates."""

    arrays = []
    kept = []
    for col in columns:
        if col not in df.columns:
            continue
        values = _coerce_covariate(df[col])
        if values.notna().sum() < 2 or values.nunique(dropna=True) < 2:
            continue
        values = values.fillna(values.median())
        arrays.append(values.astype(float).values)
        kept.append(col)

    if not arrays:
        return np.empty((len(df), 0)), []

    return np.vstack(arrays).T, kept


def _coerce_covariate(series: pd.Series) -> pd.Series:
    lower = series.astype(str).str.strip().str.lower()
    mapped = lower.map(
        {
            "male": 1.0,
            "m": 1.0,
            "female": 0.0,
            "f": 0.0,
            "man": 1.0,
            "woman": 0.0,
        }
    )
    numeric = pd.to_numeric(lower, errors="coerce")
    return numeric.where(numeric.notna(), mapped)
```
