## Supplementary material for "Auditable cross-instrument detection of unusual multivariate psychiatric response configurations using a semantically aligned covariance subspace": Code for tool: pipeline.py

```
"""Core semantic survey analysis pipeline."""

from dataclasses import dataclass
from typing import Dict, Iterable, List, Mapping, Optional, Sequence, Tuple

import numpy as np
import pandas as pd
from scipy.stats import chi2
from sklearn.covariance import LedoitWolf
from sklearn.decomposition import PCA
from sklearn.impute import KNNImputer
from sklearn.linear_model import LinearRegression

from survey_semantics.basis import SemanticBasis, build_semantic_basis
from survey_semantics.embedding import (
    ItemEmbeddings,
    embed_texts_with_metadata,
    enforce_local_ai_offline_policy,
    install_outbound_socket_blocker,
)
from survey_semantics.io import (
    SurveyTable,
    build_covariate_matrix,
    coerce_response_frame,
    default_covariates,
    default_id_column,
    infer_item_columns,
)
from survey_semantics.scales import (
    ItemScale,
    is_item_embedded,
    resolve_scale,
    scales_use_embed,
)


@dataclass
class AnalysisConfig:
    """Configuration for semantic survey analysis."""

    embedding: str = "sentence-transformers"
    model_name: Optional[str] = None
    disable_network: bool = True
    compute_umap: bool = True
    umap_n_components: int = 2
    umap_n_neighbors: int = 15
    umap_min_dist: float = 0.10
    umap_metric: str = "euclidean"
    random_state: int = 42
    variance_threshold: float = 0.80
    d_selection_method: str = "variance"
    d_null_permutations: int = 50
    d_null_percentile: float = 95.0
    stability_jaccard_threshold: float = 0.90
    stability_consecutive: int = 2
    alpha: float = 0.01
    empirical_percentiles: Tuple[int, int] = (95, 99)
    min_complete_fraction: float = 0.50
    min_rows: int = 10
    min_items: int = 5
    max_unique: int = 8
    impute_neighbors: int = 5
    id_col: Optional[str] = None
    covariates: Optional[Sequence[str]] = None
    top_outliers: int = 10
    top_components: int = 3
    top_items: int = 5
    reverse_items: Optional[Iterable[str]] = None
    sentinels: Optional[Iterable[int]] = None
    item_scales: Optional[Mapping[str, ItemScale]] = None
    sample_weights: Optional[np.ndarray] = None  # length == len(table.data), row-aligned
    pan_mild: bool = False           # flag outliers that are nowhere at the Likert ceiling
    ceiling_min_levels: int = 3      # only items with >= this many response levels count for ceiling


@dataclass
class AnalysisResult:
    table_name: str
    scores: pd.DataFrame
    prompt_loadings: pd.DataFrame
    item_weights: pd.DataFrame
    drivers: pd.DataFrame
    stability: pd.DataFrame
    dimension_selection: pd.DataFrame
    dimension_methods: pd.DataFrame
    raw_response_umap: pd.DataFrame
    semantic_pc_umap: pd.DataFrame
    case_studies: Dict[str, str]
    case_study_label_map: pd.DataFrame
    summary: Dict[str, object]


def normalize_responses(
    response_matrix: np.ndarray,
    item_ranges: Sequence[Tuple[float, float]],
) -> np.ndarray:
    mins = np.asarray([item_range[0] for item_range in item_ranges], dtype=float)
    maxima = np.asarray([item_range[1] for item_range in item_ranges], dtype=float)
    ranges = maxima - mins
    if np.any(ranges == 0):
        raise ValueError("Cannot normalize items with zero response range.")
    normalized = 2.0 * ((response_matrix - mins) / ranges) - 1.0
    return np.clip(normalized, -1.0, 1.0)


def select_component_count(
    cumulative_variance: np.ndarray,
    variance_threshold: float,
    n_components: int,
) -> Tuple[int, bool]:
    """Choose the variance-selected PC count.

    If the threshold is not reached within the full decomposition, fall back to
    the full set of components instead of one PC.
    """

    if n_components < 1 or cumulative_variance.size == 0 or cumulative_variance[-1] <= 0:
        return 1, False

    reached = cumulative_variance >= variance_threshold
    if reached.any():
        selected = int(np.argmax(reached)) + 1
        return max(1, min(selected, n_components)), True
    return n_components, False


def leading_true_count(values: Sequence[bool]) -> int:
    count = 0
    for value in values:
        if not bool(value):
            break
        count += 1
    return count


def eigengap_dimension(eigenvalues: np.ndarray) -> Tuple[int, float]:
    if len(eigenvalues) < 2:
        return 1, float("nan")
    current = np.asarray(eigenvalues[:-1], dtype=float)
    following = np.asarray(eigenvalues[1:], dtype=float)
    ratios = np.divide(
        current,
        following,
        out=np.full_like(current, np.nan, dtype=float),
        where=following > 0,
    )
    if np.all(np.isnan(ratios)):
        return 1, float("nan")
    idx = int(np.nanargmax(ratios))
    return idx + 1, float(ratios[idx])


def parallel_analysis(
    matrix: np.ndarray,
    observed_eigenvalues: np.ndarray,
    n_components: int,
    n_permutations: int,
    percentile: float,
    random_state: int,
) -> Dict[str, object]:
    if n_permutations <= 0:
        return {
            "selected_d": pd.NA,
            "null_mean": np.full(n_components, np.nan),
            "null_percentile": np.full(n_components, np.nan),
            "significant": np.full(n_components, False),
        }

    rng = np.random.RandomState(random_state)
    null_eigenvalues = np.zeros((n_permutations, n_components), dtype=float)
    for perm_idx in range(n_permutations):
        permuted = permute_columns_independently(matrix, rng)
        pca = PCA(
            n_components=n_components,
            svd_solver="randomized",
            random_state=int(rng.randint(0, np.iinfo(np.int32).max)),
        )
        pca.fit(permuted)
        null_eigenvalues[perm_idx, :] = pca.explained_variance_

    cutoff = np.percentile(null_eigenvalues, percentile, axis=0)
    significant = np.asarray(observed_eigenvalues[:n_components] > cutoff, dtype=bool)
    return {
        "selected_d": leading_true_count(significant),
        "null_mean": null_eigenvalues.mean(axis=0),
        "null_percentile": cutoff,
        "significant": significant,
    }


def permute_columns_independently(matrix: np.ndarray, rng: np.random.RandomState) -> np.ndarray:
    permuted = np.empty_like(matrix)
    for col in range(matrix.shape[1]):
        permuted[:, col] = matrix[rng.permutation(matrix.shape[0]), col]
    return permuted


def stability_dimension(
    stability: pd.DataFrame,
    threshold: float,
    consecutive: int,
) -> Tuple[int, bool]:
    if stability.empty or "jaccard_vs_previous" not in stability.columns:
        return 1, False
    consecutive = max(1, int(consecutive))
    values = stability["jaccard_vs_previous"].astype(float).values
    dims = stability["components"].astype(int).values
    for idx in range(1, len(values)):
        window = values[idx: idx + consecutive]
        if len(window) == consecutive and np.all(window >= threshold):
            return int(dims[idx]), True
    return int(dims[-1]), False


def normalize_d_selection_method(method: str) -> str:
    aliases = {
        "variance": "variance",
        "variance_threshold": "variance",
        "variance_threshold_with_max_fallback": "variance",
        "eigengap": "eigengap",
        "eigengap_ratio": "eigengap",
        "parallel": "parallel",
        "parallel_analysis": "parallel",
        "stability": "stability",
        "outlier_stability": "stability",
        "max": "max",
        "all": "max",
        "all_pcs": "max",
        "all_components": "max",
    }
    normalized = aliases.get(str(method or "variance").strip().lower())
    if normalized is None:
        raise ValueError(
            "Unsupported D selection method {!r}; choose variance, eigengap, parallel, stability, or max.".format(
                method
            )
        )
    return normalized


def choose_dimension(
    method: str,
    d_variance: int,
    d_eigengap: int,
    parallel: Dict[str, object],
    d_stability: int,
    n_components: int,
) -> int:
    """Choose the semantic PC subspace used for scoring."""

    normalized = normalize_d_selection_method(method)
    if normalized == "variance":
        selected = d_variance
    elif normalized == "eigengap":
        selected = d_eigengap
    elif normalized == "parallel":
        parallel_d = parallel.get("selected_d", pd.NA)
        if pd.isna(parallel_d):
            raise ValueError(
                "D selection method 'parallel' requires --d-null-permutations greater than 0."
            )
        selected = int(parallel_d)
    elif normalized == "stability":
        selected = d_stability
    else:
        selected = n_components
    return max(1, min(int(selected), int(n_components)))


def analyze_survey_table(
    table: SurveyTable,
    config: Optional[AnalysisConfig] = None,
    item_columns: Optional[Sequence[str]] = None,
    item_embeddings: Optional[ItemEmbeddings] = None,
    basis: Optional[SemanticBasis] = None,
) -> AnalysisResult:
    """Run semantic manifold analysis for one survey table.

    The three pipeline stages are decoupled. You may pass, in order of precedence:

    - ``basis``: a precomputed :class:`~survey_semantics.basis.SemanticBasis`
      (from a prior `pca` step) — skips both the model and PCA.
    - ``item_embeddings``: precomputed item embeddings (from a prior `embed`
      step) — skips the model; PCA runs here.
    - neither: items are embedded inline (loads the model), then PCA runs here.
    """

    config = config or AnalysisConfig()
    # Network/offline guards are only needed when we actually load the model.
    if config.disable_network and item_embeddings is None and basis is None:
        enforce_local_ai_offline_policy()
        install_outbound_socket_blocker()

    # Resolve a per-item scale spec (min/max/sentinels/reverse) for each column,
    # when a scale file was supplied. Built once and reused below.
    resolved_scales: Dict[str, ItemScale] = {}

    if item_columns is not None:
        item_columns = list(item_columns)
    elif config.item_scales:
        # Declared items take the place of inference: an item is analyzed iff a
        # scale was declared for it and the column exists in the data. This
        # avoids NHIS-style sentinel codes (7/8/9) inflating the unique-value
        # count and wrongly dropping valid items. When the scale file declares an
        # `embed` column, it is an explicit allowlist: only embed=true items are
        # analyzed (embed=false items stay documented but excluded).
        uses_embed = scales_use_embed(config.item_scales)
        item_columns = [
            col for col in table.data.columns
            if is_item_embedded(
                resolve_scale(config.item_scales, table.name, col), uses_embed
            )
        ]
    else:
        item_columns = list(infer_item_columns(
            table,
            min_nonmissing=config.min_rows,
            max_unique=config.max_unique,
            sentinels=config.sentinels,
        ))

    if config.item_scales:
        for col in item_columns:
            scale = resolve_scale(config.item_scales, table.name, col)
            if scale is not None:
                resolved_scales[col] = scale

    if len(item_columns) < config.min_items:
        raise ValueError(
            "Only {} usable item columns found; need at least {}.".format(
                len(item_columns), config.min_items
            )
        )

    # Per-item sentinels when scales are present; otherwise the global set.
    sentinels: object = config.sentinels
    if resolved_scales:
        sentinels = {
            col: scale["sentinels"] for col, scale in resolved_scales.items()
        }

    responses_raw = coerce_response_frame(
        table.data,
        item_columns,
        sentinels=sentinels,
    )
    min_complete = max(1, int(np.ceil(len(item_columns) * config.min_complete_fraction)))
    keep_rows = responses_raw.notna().sum(axis=1) >= min_complete
    responses_raw = responses_raw.loc[keep_rows]
    metadata = table.data.loc[keep_rows].copy()

    if len(responses_raw) < config.min_rows:
        raise ValueError(
            "Only {} usable rows after completeness filtering; need at least {}.".format(
                len(responses_raw), config.min_rows
            )
        )

    # Survey weights (optional): row-aligned to the raw table, subset to the kept
    # subjects, then guarded like the NHIS script (non-finite/<=0 -> median).
    weights = None
    if config.sample_weights is not None:
        weights = np.asarray(config.sample_weights, dtype=float)
        if weights.shape[0] != table.data.shape[0]:
            raise ValueError(
                "Weights length ({}) must equal the number of response rows ({}).".format(
                    weights.shape[0], table.data.shape[0]
                )
            )
        weights = weights[keep_rows.to_numpy()]
        valid = np.isfinite(weights) & (weights > 0)
        if not valid.all():
            if not valid.any():
                raise ValueError("Weights file has no finite positive values.")
            weights = weights.copy()
            weights[~valid] = float(np.median(weights[valid]))

    responses = _impute_response_frame(responses_raw, config.impute_neighbors)
    # Prefer declared min/max from scales; fall back to observed ranges per item.
    declared_ranges = None
    if resolved_scales:
        declared_ranges = [
            _declared_range(resolved_scales.get(col)) for col in item_columns
        ]
    item_ranges = _observed_item_ranges(responses_raw, responses, declared_ranges)
    response_norm = normalize_responses(responses.values, item_ranges)

    # Reverse-scored items: explicit config plus any declared by the scales.
    reverse_items = set(config.reverse_items or [])
    for col, scale in resolved_scales.items():
        if scale.get("reverse"):
            reverse_items.add(col)
    for idx, col in enumerate(item_columns):
        if col in reverse_items:
            response_norm[:, idx] *= -1.0

    # Ceiling audit (pan-mild): a subject is "at ceiling" if any polytomous item
    # (>= ceiling_min_levels response levels) sits at its maximum-severity end of
    # the normalized [-1, 1] scale. response_norm is already severity-oriented
    # (reverse-scored), so the ceiling is +1 for every item — matching the
    # manuscript's reverse-then-normalize convention.
    at_ceiling = None
    if config.pan_mild:
        ceiling_mask = ceiling_item_mask(
            item_columns, item_ranges, config.item_scales, config.ceiling_min_levels
        )
        at_ceiling = ceiling_flags(response_norm, ceiling_mask)

    # Item wording, used for prompt-loading labels regardless of embedding source.
    item_texts = [
        _item_text(col, table.dictionary.get(col, col))
        for col in item_columns
    ]

    # Stages 1-2 (embed -> PCA). A precomputed basis skips both; precomputed
    # embeddings skip only the model; otherwise items are embedded inline. Either
    # way this yields the item coordinates, eigen/variance arrays, the parallel
    # diagnostic, and the embedding provenance for output naming.
    if basis is not None:
        item_coordinates_full = basis.coordinates_for(item_columns)
        eigenvalues = basis.eigenvalues
        explained = basis.explained_variance_ratio
        cumulative = basis.cumulative_variance
        parallel = basis.parallel
        n_components = basis.n_components
        emb_backend = basis.embedding_backend
        emb_model = basis.embedding_model
        emb_slug = basis.embedding_slug
        emb_requested_backend = basis.embedding_backend
        emb_requested_model = basis.embedding_model
        basis_source = "precomputed"
    else:
        if item_embeddings is not None:
            embedding_vectors = item_embeddings.matrix_for(item_columns)
            emb_backend = item_embeddings.backend
            emb_model = item_embeddings.model_name
            emb_slug = item_embeddings.slug
            emb_requested_backend = item_embeddings.backend
            emb_requested_model = item_embeddings.model_name
            basis_source = "computed_from_embeddings"
        else:
            embedding_result = embed_texts_with_metadata(
                item_texts,
                method=config.embedding,
                model_name=config.model_name,
            )
            embedding_vectors = embedding_result.vectors
            emb_backend = embedding_result.backend
            emb_model = embedding_result.model_name
            emb_slug = embedding_result.slug
            emb_requested_backend = embedding_result.requested_backend
            emb_requested_model = embedding_result.requested_model_name
            basis_source = "computed_inline"

        computed_basis = build_semantic_basis(
            items=item_columns,
            embedding_vectors=embedding_vectors,
            d_null_permutations=config.d_null_permutations,
            d_null_percentile=config.d_null_percentile,
            random_state=config.random_state,
            embedding_backend=emb_backend,
            embedding_model=emb_model,
            embedding_slug=emb_slug,
        )
        item_coordinates_full = computed_basis.components
        eigenvalues = computed_basis.eigenvalues
        explained = computed_basis.explained_variance_ratio
        cumulative = computed_basis.cumulative_variance
        parallel = computed_basis.parallel
        n_components = computed_basis.n_components

    d_variance, variance_threshold_reached = select_component_count(
        cumulative_variance=cumulative,
        variance_threshold=config.variance_threshold,
        n_components=n_components,
    )
    d_eigengap, eigengap_ratio = eigengap_dimension(eigenvalues)

    covariate_names = list(config.covariates) if config.covariates is not None else default_covariates(metadata)
    covariates, kept_covariates = build_covariate_matrix(metadata, covariate_names)
    stability = _stability_frame(
        response_norm=response_norm,
        item_coordinates_full=item_coordinates_full,
        covariates=covariates,
        alpha=config.alpha,
        n_components=n_components,
        explained_variance=explained,
        weights=weights,
    )
    d_stability, stability_reached = stability_dimension(
        stability=stability,
        threshold=config.stability_jaccard_threshold,
        consecutive=config.stability_consecutive,
    )
    optimal_d = choose_dimension(
        method=config.d_selection_method,
        d_variance=d_variance,
        d_eigengap=d_eigengap,
        parallel=parallel,
        d_stability=d_stability,
        n_components=n_components,
    )

    item_coordinates = item_coordinates_full[:, :optimal_d]
    semantic_scores = np.dot(response_norm, item_coordinates)
    residual_scores = residualize(semantic_scores, covariates, weights=weights)

    if config.compute_umap:
        raw_response_umap = umap_embedding_frame(
            features=response_norm,
            metadata=metadata,
            covariates=covariates,
            id_col=config.id_col or default_id_column(metadata),
            source="raw_response",
            config=config,
        )
        semantic_pc_umap = umap_embedding_frame(
            features=semantic_scores,
            metadata=metadata,
            covariates=covariates,
            id_col=config.id_col or default_id_column(metadata),
            source="semantic_pc",
            config=config,
        )
    else:
        raw_response_umap = pd.DataFrame()
        semantic_pc_umap = pd.DataFrame()

    distances = mahalanobis_distances(residual_scores, weights=weights)
    scores = _score_frame(
        table=table,
        metadata=metadata,
        responses=responses,
        item_columns=item_columns,
        residual_scores=residual_scores,
        distances=distances,
        config=config,
        optimal_d=optimal_d,
        at_ceiling=at_ceiling,
    )

    prompt_loadings = _prompt_loadings_frame(
        item_columns=item_columns,
        item_texts=item_texts,
        item_ranges=item_ranges,
        item_coordinates=item_coordinates,
        explained_variance=explained[:optimal_d],
        reverse_items=reverse_items,
    )
    item_weights = prompt_loadings.copy()
    dimension_selection = _dimension_selection_frame(
        eigenvalues=eigenvalues,
        explained_variance=explained,
        cumulative_variance=cumulative,
        parallel=parallel,
        stability=stability,
    )
    dimension_methods = _dimension_methods_frame(
        optimal_d=optimal_d,
        d_selection_method=config.d_selection_method,
        d_variance=d_variance,
        cumulative_variance=cumulative,
        variance_threshold=config.variance_threshold,
        variance_threshold_reached=variance_threshold_reached,
        n_components=n_components,
        d_eigengap=d_eigengap,
        eigengap_ratio=eigengap_ratio,
        parallel=parallel,
        config=config,
        d_stability=d_stability,
        stability_reached=stability_reached,
    )
    drivers = _driver_frame(
        table_name=table.name,
        scores=scores,
        responses=responses,
        residual_scores=residual_scores,
        item_coordinates=item_coordinates,
        item_columns=item_columns,
        item_texts=item_texts,
        id_col=config.id_col or default_id_column(metadata),
        top_outliers=config.top_outliers,
        top_components=config.top_components,
        top_items=config.top_items,
    )
    case_studies, case_study_label_map = _case_study_reports(
        table.name,
        scores,
        drivers,
        config.id_col or default_id_column(metadata),
    )

    summary = {
        "table": table.name,
        "path": str(table.path),
        "n_rows": int(len(scores)),
        "n_items": int(len(item_columns)),
        "optimal_d": int(optimal_d),
        "embedding": emb_backend,
        "embedding_model": emb_model,
        "embedding_slug": emb_slug,
        "requested_embedding": emb_requested_backend,
        "requested_embedding_model": emb_requested_model,
        "semantic_basis_source": basis_source,
        "explained_variance": float(cumulative[optimal_d - 1]) if cumulative.size else 0.0,
        "variance_threshold": float(config.variance_threshold),
        "variance_threshold_reached": bool(variance_threshold_reached),
        "components_evaluated": int(n_components),
        "d_selection_method": normalize_d_selection_method(config.d_selection_method),
        "d_variance_selected": int(d_variance),
        "d_eigengap": int(d_eigengap),
        "eigengap_ratio": float(eigengap_ratio) if not pd.isna(eigengap_ratio) else pd.NA,
        "d_parallel_analysis": parallel["selected_d"],
        "parallel_analysis_stop_reached": bool(
            (not pd.isna(parallel["selected_d"])) and int(parallel["selected_d"]) < n_components
        ),
        "parallel_null_permutations": int(config.d_null_permutations),
        "parallel_null_percentile": float(config.d_null_percentile),
        "d_outlier_stability": int(d_stability),
        "outlier_stability_reached": bool(stability_reached),
        "outlier_stability_jaccard_threshold": float(config.stability_jaccard_threshold),
        "covariates": ",".join(kept_covariates),
        "theoretical_threshold": float(np.sqrt(chi2.ppf(1 - config.alpha, df=optimal_d))),
        "outliers_theoretical": int(scores["Is_Outlier_Theo"].sum()),
        "outliers_empirical_95": int(scores.get("Is_Outlier_Emp95", pd.Series(dtype=bool)).sum()),
        "outliers_empirical_99": int(scores.get("Is_Outlier_Emp99", pd.Series(dtype=bool)).sum()),
        "pan_mild_enabled": bool(config.pan_mild),
        "at_ceiling_count": int(scores.get("At_Ceiling", pd.Series(dtype=bool)).sum()),
        "pan_mild_empirical_95": int(scores.get("Is_Pan_Mild_Emp95", pd.Series(dtype=bool)).sum()),
        "pan_mild_empirical_99": int(scores.get("Is_Pan_Mild_Emp99", pd.Series(dtype=bool)).sum()),
        "max_mahalanobis": float(np.nanmax(distances)),
        "umap_outputs": bool(config.compute_umap),
    }
    return AnalysisResult(
        table_name=table.name,
        scores=scores,
        prompt_loadings=prompt_loadings,
        item_weights=item_weights,
        drivers=drivers,
        stability=stability,
        dimension_selection=dimension_selection,
        dimension_methods=dimension_methods,
        raw_response_umap=raw_response_umap,
        semantic_pc_umap=semantic_pc_umap,
        case_studies=case_studies,
        case_study_label_map=case_study_label_map,
        summary=summary,
    )


def residualize(
    matrix: np.ndarray,
    covariates: np.ndarray,
    weights: Optional[np.ndarray] = None,
) -> np.ndarray:
    if covariates.size == 0 or covariates.shape[1] == 0:
        if weights is not None and weights.sum() > 0:
            w = weights / weights.sum()
            return matrix - (w[:, np.newaxis] * matrix).sum(axis=0)
        return matrix - matrix.mean(axis=0, keepdims=True)

    if weights is None:
        residual = np.zeros_like(matrix, dtype=float)
        for idx in range(matrix.shape[1]):
            model = LinearRegression()
            model.fit(covariates, matrix[:, idx])
            residual[:, idx] = matrix[:, idx] - model.predict(covariates)
        return residual

    # Weighted least squares: row-scale design and targets by sqrt(w), matching
    # the NHIS replication script's WLS residualization.
    w = weights / weights.sum()
    design = np.column_stack([np.ones(len(matrix)), covariates])
    sqrt_w = np.sqrt(w)[:, np.newaxis]
    beta, _, _, _ = np.linalg.lstsq(design * sqrt_w, matrix * sqrt_w, rcond=None)
    return matrix - design @ beta


def mahalanobis_distances(
    matrix: np.ndarray,
    weights: Optional[np.ndarray] = None,
) -> np.ndarray:
    if matrix.shape[0] < 2:
        return np.zeros(matrix.shape[0], dtype=float)

    if weights is None:
        covariance = LedoitWolf().fit(matrix)
        squared = covariance.mahalanobis(matrix)
        return np.sqrt(np.maximum(squared, 0.0))

    # Weighted mean + weighted scatter with Ledoit-Wolf shrinkage, matching the
    # NHIS script: the shrinkage coefficient comes from an unweighted LedoitWolf
    # fit, but the covariance shrunk is the weighted scatter S.
    n_components = matrix.shape[1]
    w = weights / weights.sum()
    mu = (w[:, np.newaxis] * matrix).sum(axis=0)
    diff = matrix - mu
    n_eff = 1.0 / float((w ** 2).sum())
    scatter = (diff * w[:, np.newaxis]).T @ diff * (n_eff / (n_eff - 1.0))
    alpha = float(LedoitWolf().fit(matrix).shrinkage_)
    target = (np.trace(scatter) / n_components) * np.eye(n_components)
    cov_shrunk = (1.0 - alpha) * scatter + alpha * target
    cov_inv = np.linalg.inv(cov_shrunk)
    squared = np.sum((diff @ cov_inv) * diff, axis=1)
    return np.sqrt(np.maximum(squared, 0.0))


def umap_embedding_frame(
    features: np.ndarray,
    metadata: pd.DataFrame,
    covariates: np.ndarray,
    id_col: Optional[str],
    source: str,
    config: AnalysisConfig,
) -> pd.DataFrame:
    """Fit UMAP and return covariate-residualized 2D coordinates.

    This mirrors the notebook's UMAP contrast: UMAP is fit on the chosen feature
    representation first, then age/sex/etc. are regressed out of the UMAP axes
    when covariates are available.
    """

    if features.shape[0] < 3:
        raise ValueError("UMAP requires at least 3 usable rows.")
    try:
        umap = _import_umap()
    except ImportError as exc:
        raise ImportError(
            "UMAP outputs require the optional dependency 'umap-learn'. "
            "Install it with: pip install -e '.[umap]' or pip install umap-learn. "
            "Use --skip-umap to run only the semantic PCA/Mahalanobis outputs."
        ) from exc

    n_neighbors = max(2, min(config.umap_n_neighbors, features.shape[0] - 1))
    reducer = umap.UMAP(
        n_components=config.umap_n_components,
        n_neighbors=n_neighbors,
        min_dist=config.umap_min_dist,
        metric=config.umap_metric,
        random_state=config.random_state,
    )
    coordinates = reducer.fit_transform(features)
    coordinates = residualize(np.asarray(coordinates, dtype=float), covariates)

    frame = pd.DataFrame(index=metadata.index)
    frame["Source_Row"] = metadata.index.values
    if id_col and id_col in metadata.columns:
        frame[id_col] = metadata[id_col].values
    frame["UMAP_Source"] = source
    frame["UMAP_Neighbors"] = n_neighbors
    frame["UMAP_Min_Dist"] = config.umap_min_dist
    frame["UMAP_Metric"] = config.umap_metric
    for idx in range(coordinates.shape[1]):
        frame["UMAP_Dim{}".format(idx + 1)] = coordinates[:, idx]
    return frame.reset_index(drop=True)


def _import_umap():
    """Import UMAP with a numba-cache workaround for older conda stacks."""

    try:
        import numba
    except ImportError:
        pass
    else:
        if not getattr(numba, "_survey_semantics_no_cache_patch", False):
            original_jit = numba.jit
            original_njit = numba.njit
            original_vectorize = getattr(numba, "vectorize", None)
            original_guvectorize = getattr(numba, "guvectorize", None)

            def no_cache_jit(*args, **kwargs):
                kwargs.pop("cache", None)
                return original_jit(*args, **kwargs)

            def no_cache_njit(*args, **kwargs):
                kwargs.pop("cache", None)
                return original_njit(*args, **kwargs)

            def no_cache_vectorize(*args, **kwargs):
                kwargs.pop("cache", None)
                return original_vectorize(*args, **kwargs)

            def no_cache_guvectorize(*args, **kwargs):
                kwargs.pop("cache", None)
                return original_guvectorize(*args, **kwargs)

            numba.jit = no_cache_jit
            numba.njit = no_cache_njit
            if original_vectorize is not None:
                numba.vectorize = no_cache_vectorize
            if original_guvectorize is not None:
                numba.guvectorize = no_cache_guvectorize
            numba._survey_semantics_no_cache_patch = True

    import umap

    return umap


def _impute_response_frame(frame: pd.DataFrame, n_neighbors: int) -> pd.DataFrame:
    if not frame.isna().any().any():
        return frame.astype(float)

    medians = frame.median(axis=0)
    safe = frame.fillna(medians)
    if len(frame) <= 2:
        return safe.astype(float)

    neighbors = max(1, min(n_neighbors, len(frame) - 1))
    try:
        imputer = KNNImputer(n_neighbors=neighbors, weights="distance")
        values = imputer.fit_transform(frame)
        return pd.DataFrame(values, index=frame.index, columns=frame.columns)
    except Exception:
        return safe.astype(float)


def _observed_item_ranges(
    raw: pd.DataFrame,
    imputed: pd.DataFrame,
    declared: Optional[Sequence[Optional[Tuple[float, float]]]] = None,
) -> List[Tuple[float, float]]:
    ranges = []
    for idx, col in enumerate(imputed.columns):
        if declared is not None and declared[idx] is not None:
            ranges.append(declared[idx])
            continue
        observed = raw[col].dropna()
        if observed.empty:
            observed = imputed[col].dropna()
        low = float(observed.min())
        high = float(observed.max())
        if low == high:
            low = float(imputed[col].min())
            high = float(imputed[col].max())
        if low == high:
            high = low + 1.0
        ranges.append((low, high))
    return ranges


def ceiling_item_mask(
    item_columns: Sequence[str],
    item_ranges: Sequence[Tuple[float, float]],
    item_scales: Optional[Mapping[str, ItemScale]] = None,
    min_levels: int = 3,
) -> np.ndarray:
    """Which items participate in the item-level ceiling check.

    If any item declares a ``ceiling`` flag in its scale, those flags are an
    explicit allowlist (only ``ceiling=True`` items are checked) — use this to
    exclude items where "maxed out" is not a symptom ceiling (e.g. time-since-last-
    doctor-visit). If no item declares ``ceiling``, fall back to all polytomous
    items (>= ``min_levels`` response levels), matching the manuscript.
    """

    declared = [
        (item_scales.get(col, {}).get("ceiling") if item_scales else None)
        for col in item_columns
    ]
    if any(flag is not None for flag in declared):
        return np.array([bool(flag) for flag in declared])

    levels = np.array([int(round(high - low)) + 1 for (low, high) in item_ranges])
    return levels >= min_levels


def ceiling_flags(response_norm: np.ndarray, item_mask: np.ndarray) -> np.ndarray:
    """Per-subject boolean: is any selected item at its severity ceiling?

    `response_norm` is the severity-oriented matrix normalized to [-1, 1] (already
    reverse-scored), so the ceiling is +1 for every item. A subject is "at ceiling"
    if any item in `item_mask` is >= 0.999.
    """

    item_mask = np.asarray(item_mask, dtype=bool)
    if not item_mask.any():
        return np.zeros(response_norm.shape[0], dtype=bool)
    return (response_norm[:, item_mask] >= 0.999).any(axis=1)


def _declared_range(scale: Optional[ItemScale]) -> Optional[Tuple[float, float]]:
    """Return a usable (min, max) from a scale spec, or None to fall back to
    observed ranges (when min/max are absent or non-increasing)."""

    if scale is None:
        return None
    low = scale.get("min")
    high = scale.get("max")
    if low is None or high is None or high <= low:
        return None
    return (float(low), float(high))


def _item_text(col: str, description: str) -> str:
    description = str(description).strip()
    if description and description.lower() != str(col).lower():
        return description
    return str(col).replace("_", " ")


def _score_frame(
    table: SurveyTable,
    metadata: pd.DataFrame,
    responses: pd.DataFrame,
    item_columns: Sequence[str],
    residual_scores: np.ndarray,
    distances: np.ndarray,
    config: AnalysisConfig,
    optimal_d: int,
    at_ceiling: Optional[np.ndarray] = None,
) -> pd.DataFrame:
    id_col = config.id_col or default_id_column(metadata)
    keep_cols = []
    if id_col and id_col in metadata.columns:
        keep_cols.append(id_col)
    for col in default_covariates(metadata):
        if col not in keep_cols:
            keep_cols.append(col)

    scores = metadata[keep_cols].copy() if keep_cols else pd.DataFrame(index=metadata.index)
    scores.insert(0, "Source_Row", metadata.index.values)
    scores["Mahalanobis_Dist"] = distances
    scores["Mahal_Rank"] = scores["Mahalanobis_Dist"].rank(ascending=False, method="min").astype(int)
    scores["Mahal_Empirical_Pctile"] = (
        (len(scores) - scores["Mahal_Rank"]) / float(len(scores)) * 100.0
    )

    theoretical = np.sqrt(chi2.ppf(1 - config.alpha, df=optimal_d))
    scores["Is_Outlier_Theo"] = distances > theoretical
    for percentile in config.empirical_percentiles:
        threshold = np.percentile(distances, percentile)
        scores["Is_Outlier_Emp{}".format(percentile)] = distances > threshold

    # Pan-mild audit: outliers whose item-level profile is nowhere at the Likert
    # ceiling. Computed per empirical percentile so each outlier threshold has a
    # matching pan-mild column (e.g. Is_Pan_Mild_Emp95).
    if at_ceiling is not None:
        scores["At_Ceiling"] = at_ceiling
        for percentile in config.empirical_percentiles:
            outlier_col = "Is_Outlier_Emp{}".format(percentile)
            scores["Is_Pan_Mild_Emp{}".format(percentile)] = (
                scores[outlier_col].to_numpy() & (~at_ceiling)
            )

    for idx in range(optimal_d):
        scores["Semantic_PC{}".format(idx + 1)] = residual_scores[:, idx]

    response_columns = responses.loc[:, item_columns]
    return pd.concat([scores, response_columns], axis=1)


def _prompt_loadings_frame(
    item_columns: Sequence[str],
    item_texts: Sequence[str],
    item_ranges: Sequence[Tuple[float, float]],
    item_coordinates: np.ndarray,
    explained_variance: np.ndarray,
    reverse_items: Iterable[str],
) -> pd.DataFrame:
    reverse_items = set(reverse_items)
    records = []
    for idx, col in enumerate(item_columns):
        record = {
            "item": col,
            "prompt": item_texts[idx],
            "reverse_scored": col in reverse_items,
            "observed_min": item_ranges[idx][0],
            "observed_max": item_ranges[idx][1],
        }
        for dim in range(item_coordinates.shape[1]):
            record["PC{}".format(dim + 1)] = item_coordinates[idx, dim]
            record["PC{}_explained_variance_ratio".format(dim + 1)] = explained_variance[dim]
        records.append(record)
    return pd.DataFrame(records)


def _stability_frame(
    response_norm: np.ndarray,
    item_coordinates_full: np.ndarray,
    covariates: np.ndarray,
    alpha: float,
    n_components: int,
    explained_variance: np.ndarray,
    weights: Optional[np.ndarray] = None,
) -> pd.DataFrame:
    records = []
    previous = set()
    cumulative = np.cumsum(explained_variance)
    for dim in range(1, n_components + 1):
        scores = np.dot(response_norm, item_coordinates_full[:, :dim])
        residual = residualize(scores, covariates, weights=weights)
        distances = mahalanobis_distances(residual, weights=weights)
        threshold = np.sqrt(chi2.ppf(1 - alpha, df=dim))
        outliers = set(np.where(distances > threshold)[0])
        if not previous:
            jaccard = 1.0
        else:
            union = outliers | previous
            jaccard = float(len(outliers & previous)) / len(union) if union else 1.0
        records.append(
            {
                "components": dim,
                "theoretical_threshold": threshold,
                "outlier_count": len(outliers),
                "jaccard_vs_previous": jaccard,
                "explained_variance_ratio": explained_variance[dim - 1],
                "cumulative_explained_variance": cumulative[dim - 1],
            }
        )
        previous = outliers
    return pd.DataFrame(records)


def _dimension_selection_frame(
    eigenvalues: np.ndarray,
    explained_variance: np.ndarray,
    cumulative_variance: np.ndarray,
    parallel: Dict[str, object],
    stability: pd.DataFrame,
) -> pd.DataFrame:
    components = np.arange(1, len(explained_variance) + 1)
    eigengap = np.full(len(explained_variance), np.nan, dtype=float)
    if len(eigenvalues) > 1:
        eigengap[:-1] = np.divide(
            eigenvalues[:-1],
            eigenvalues[1:],
            out=np.full(len(eigenvalues) - 1, np.nan, dtype=float),
            where=eigenvalues[1:] > 0,
        )

    frame = pd.DataFrame(
        {
            "components": components,
            "eigenvalue": eigenvalues,
            "explained_variance_ratio": explained_variance,
            "cumulative_explained_variance": cumulative_variance,
            "eigengap_ratio_to_next": eigengap,
            "parallel_null_mean_eigenvalue": parallel["null_mean"],
            "parallel_null_percentile_eigenvalue": parallel["null_percentile"],
            "parallel_significant": parallel["significant"],
        }
    )
    if not stability.empty:
        keep = [
            col
            for col in ["components", "jaccard_vs_previous", "outlier_count", "theoretical_threshold"]
            if col in stability.columns
        ]
        frame = frame.merge(stability[keep], on="components", how="left")
    return frame


def _dimension_methods_frame(
    optimal_d: int,
    d_selection_method: str,
    d_variance: int,
    cumulative_variance: np.ndarray,
    variance_threshold: float,
    variance_threshold_reached: bool,
    n_components: int,
    d_eigengap: int,
    eigengap_ratio: float,
    parallel: Dict[str, object],
    config: AnalysisConfig,
    d_stability: int,
    stability_reached: bool,
) -> pd.DataFrame:
    selection_method = normalize_d_selection_method(d_selection_method)
    variance_at_selected = float(cumulative_variance[optimal_d - 1]) if cumulative_variance.size else 0.0
    variance_at_variance_d = float(cumulative_variance[d_variance - 1]) if cumulative_variance.size else 0.0
    parallel_d = parallel["selected_d"]
    parallel_stop_reached = (not pd.isna(parallel_d)) and int(parallel_d) < n_components
    parallel_detail = (
        "leading PCs above null percentile"
        if not pd.isna(parallel_d)
        else "parallel analysis disabled"
    )
    records = [
        {
            "method": "variance_threshold_with_max_fallback",
            "selected_d": int(d_variance),
            "criterion_value": variance_at_variance_d,
            "criterion_threshold": float(variance_threshold),
            "criterion_reached": bool(variance_threshold_reached),
            "used_for_scores": selection_method == "variance",
            "details": "first cumulative variance crossing; all components if not reached",
        },
        {
            "method": "eigengap_ratio",
            "selected_d": int(d_eigengap),
            "criterion_value": eigengap_ratio,
            "criterion_threshold": pd.NA,
            "criterion_reached": True,
            "used_for_scores": selection_method == "eigengap",
            "details": "largest adjacent eigenvalue ratio lambda_D/lambda_D+1",
        },
        {
            "method": "parallel_analysis",
            "selected_d": parallel_d,
            "criterion_value": pd.NA,
            "criterion_threshold": float(config.d_null_percentile),
            "criterion_reached": bool(parallel_stop_reached),
            "used_for_scores": selection_method == "parallel",
            "details": "{}; permutations={}; stop reached before all components={}".format(
                parallel_detail,
                config.d_null_permutations,
                bool(parallel_stop_reached),
            ),
        },
        {
            "method": "outlier_stability",
            "selected_d": int(d_stability),
            "criterion_value": float(config.stability_jaccard_threshold),
            "criterion_threshold": float(config.stability_jaccard_threshold),
            "criterion_reached": bool(stability_reached),
            "used_for_scores": selection_method == "stability",
            "details": "first D with neighboring-D Jaccard stable for {} step(s); all components if not reached".format(
                config.stability_consecutive
            ),
        },
        {
            "method": "all_components",
            "selected_d": int(n_components),
            "criterion_value": pd.NA,
            "criterion_threshold": pd.NA,
            "criterion_reached": True,
            "used_for_scores": selection_method == "max",
            "details": "the full semantic subspace (all components)",
        },
        {
            "method": "selected_for_scores",
            "selected_d": int(optimal_d),
            "criterion_value": variance_at_selected,
            "criterion_threshold": pd.NA,
            "criterion_reached": True,
            "used_for_scores": True,
            "details": "actual semantic PC count used for scores; requested method={}".format(
                selection_method
            ),
        },
    ]
    return pd.DataFrame(records)


def _driver_frame(
    table_name: str,
    scores: pd.DataFrame,
    responses: pd.DataFrame,
    residual_scores: np.ndarray,
    item_coordinates: np.ndarray,
    item_columns: Sequence[str],
    item_texts: Sequence[str],
    id_col: Optional[str],
    top_outliers: int,
    top_components: int,
    top_items: int,
) -> pd.DataFrame:
    stds = residual_scores.std(axis=0)
    stds[stds == 0] = 1.0

    ordered = scores.sort_values("Mahalanobis_Dist", ascending=False).head(top_outliers)
    records = []
    for score_pos, (idx, row) in enumerate(ordered.iterrows()):
        matrix_pos = scores.index.get_loc(idx)
        z_scores = residual_scores[matrix_pos] / stds
        component_order = np.argsort(np.abs(z_scores))[::-1][:top_components]
        response_row = responses.iloc[matrix_pos]
        subject_id = row[id_col] if id_col and id_col in row else row["Source_Row"]

        for component in component_order:
            weights = item_coordinates[:, component]
            item_order = np.argsort(np.abs(weights))[::-1][:top_items]
            for item_idx in item_order:
                alignment = "aligned" if weights[item_idx] * z_scores[component] > 0 else "opposed"
                records.append(
                    {
                        "table": table_name,
                        "subject_id": subject_id,
                        "source_row": row["Source_Row"],
                        "mahalanobis": row["Mahalanobis_Dist"],
                        "rank": row["Mahal_Rank"],
                        "component": component + 1,
                        "component_z": z_scores[component],
                        "component_abs_z": abs(z_scores[component]),
                        "item": item_columns[item_idx],
                        "item_text": item_texts[item_idx],
                        "item_weight": weights[item_idx],
                        "response": response_row[item_columns[item_idx]],
                        "alignment": alignment,
                    }
                )
    return pd.DataFrame(records)


def _case_study_reports(
    table_name: str,
    scores: pd.DataFrame,
    drivers: pd.DataFrame,
    id_col: Optional[str],
) -> Tuple[Dict[str, str], pd.DataFrame]:
    if drivers.empty:
        return {}, pd.DataFrame(columns=case_study_label_map_columns())

    reports = {}
    label_rows = []
    grouped = drivers.groupby(["source_row", "subject_id"], sort=False)
    ordered_groups = sorted(
        grouped,
        key=lambda item: int(item[1].iloc[0]["rank"]),
    )
    for (source_row, subject_id), group in ordered_groups:
        first = group.iloc[0]
        score_rows = scores[scores["Source_Row"] == source_row]
        score = score_rows.iloc[0] if not score_rows.empty else None
        rank = int(first["rank"])
        outlier_label = "Outlier #{}".format(rank)
        filename = "rank{:03d}_outlier_{:03d}.txt".format(rank, rank)
        label_rows.append(
            {
                "outlier_label": outlier_label,
                "subject_id": subject_id,
                "source_row": source_row,
                "mahal_rank": rank,
                "mahalanobis_dist": float(first["mahalanobis"]),
            }
        )

        lines = [
            "Semantic Survey Case Study",
            "==========================",
            "",
            "Table: {}".format(table_name),
            "Outlier label: {}".format(outlier_label),
            "Mahalanobis distance: {:.4f}".format(float(first["mahalanobis"])),
            "Empirical rank: {}".format(rank),
        ]
        if score is not None:
            lines.append("Empirical percentile: {:.3f}".format(float(score["Mahal_Empirical_Pctile"])))
            flag_cols = [col for col in score.index if str(col).startswith("Is_Outlier_")]
            if flag_cols:
                flags = ["{}={}".format(col, bool(score[col])) for col in flag_cols]
                lines.append("Outlier flags: {}".format(", ".join(flags)))

        lines.extend(["", "Semantic Drivers", "----------------"])
        for component, component_group in group.groupby("component", sort=False):
            z_value = float(component_group.iloc[0]["component_z"])
            abs_z_value = abs(z_value)
            lines.extend(
                [
                    "",
                    "Component {}: |z|={:.3f} (signed z={:+.3f}; PC sign is arbitrary)".format(
                        int(component), abs_z_value, z_value
                    ),
                    "Top item drivers:",
                ]
            )
            for _, item in component_group.iterrows():
                lines.append(
                    "- {item}: weight={weight:+.4f}, response={response}, {alignment}".format(
                        item=item["item"],
                        weight=float(item["item_weight"]),
                        response=_format_response(item["response"]),
                        alignment=item["alignment"],
                    )
                )
                lines.append("  {}".format(item["item_text"]))

        reports[filename] = "\n".join(lines).rstrip() + "\n"
    return reports, pd.DataFrame(label_rows, columns=case_study_label_map_columns())


def case_study_label_map_columns() -> List[str]:
    return ["outlier_label", "subject_id", "source_row", "mahal_rank", "mahalanobis_dist"]


def _format_response(value: object) -> str:
    try:
        number = float(value)
    except Exception:
        return str(value)
    if number.is_integer():
        return str(int(number))
    return "{:.3f}".format(number)


def _safe_fragment(value: object) -> str:
    import re

    safe = re.sub(r"[^A-Za-z0-9_.-]+", "_", str(value)).strip("_")
    return safe[:80] or "case"
```
