## Supplementary material for "Auditable cross-instrument detection of unusual multivariate psychiatric response configurations using a semantically aligned covariance subspace": Code for tool: prompts.py

```
"""Prompt dictionary loading and matching helpers."""

import ast
import csv
import json
import re
from pathlib import Path
from typing import Dict, Mapping, Optional


PROMPT_FILE_SUFFIXES = {".csv", ".tsv", ".tab", ".txt", ".json", ".py"}


def load_prompt_sources(
    prompt_file: Optional[Path] = None,
    prompt_dir: Optional[Path] = None,
) -> Dict[str, str]:
    """Load prompt text from one combined file, a prompt directory, or both."""

    prompts: Dict[str, str] = {}
    if prompt_dir is not None:
        prompts.update(load_prompt_directory(prompt_dir))
    if prompt_file is not None:
        prompts.update(load_prompt_dictionary(prompt_file))
    return prompts


def load_prompt_directory(path: Path) -> Dict[str, str]:
    """Load per-instrument prompt files from a directory.

    Bare keys inside each file are namespaced by the file's instrument stem.
    For example, `prompts/srs02_prompts.csv` containing `item,prompt` rows
    creates keys like `srs02__parentreport_18`.
    """

    path = Path(path)
    if not path.exists() or not path.is_dir():
        raise ValueError("Prompt directory does not exist or is not a directory: {}".format(path))

    prompts: Dict[str, str] = {}
    loaded = 0
    for prompt_file in sorted(path.iterdir()):
        if prompt_file.name.startswith(".") or not prompt_file.is_file():
            continue
        if prompt_file.suffix.lower() not in PROMPT_FILE_SUFFIXES:
            continue
        table_name = prompt_table_name_from_path(prompt_file)
        prompts.update(
            load_prompt_dictionary(
                prompt_file,
                default_table=table_name,
                include_bare_keys=False,
            )
        )
        loaded += 1

    if loaded == 0:
        raise ValueError("No prompt files found in {}".format(path))
    return prompts


def load_prompt_dictionary(
    path: Optional[Path],
    default_table: Optional[str] = None,
    include_bare_keys: bool = True,
) -> Dict[str, str]:
    """Load prompt text overrides from JSON, CSV/TSV, or a Python literal dict.

    Supported text formats are intentionally permissive because notebook prompt
    dictionaries are often saved as copied Python code:

    - JSON object: {"item": "Prompt text"}
    - Python literal dict, optionally assigned to a variable.
    - CSV/TSV with `prompt` plus `item`, `feature`, or `table`+`item`.
    - line-oriented `key: prompt` or `key<TAB>prompt` text.
    """

    if path is None:
        return {}
    path = Path(path)
    text = path.read_text(encoding="utf-8-sig").strip()
    if not text:
        return {}

    mapping = _try_json_or_python_mapping(text)
    if mapping is not None:
        return _clean_mapping(mapping, default_table, include_bare_keys)

    delimited = _try_delimited_mapping(path, text, default_table, include_bare_keys)
    if delimited is not None:
        return delimited

    line_mapping = _try_line_mapping(text, default_table, include_bare_keys)
    if line_mapping:
        return line_mapping

    raise ValueError("Could not parse prompt dictionary file: {}".format(path))


def resolve_prompt(
    prompts: Mapping[str, str],
    table_name: str,
    item_name: str,
    default: str,
) -> str:
    """Return the best prompt override for a table/item pair."""

    if not prompts:
        return default
    candidates = [
        "{}__{}".format(table_name, item_name),
        "{}.{}".format(table_name, item_name),
        "{}/{}".format(table_name, item_name),
        item_name,
        _normalize_key("{}__{}".format(table_name, item_name)),
        _normalize_key(item_name),
    ]
    for key in candidates:
        if key in prompts and str(prompts[key]).strip():
            return str(prompts[key]).strip()
    return default


def apply_prompt_dictionary(
    dictionary: Mapping[str, str],
    table_name: str,
    prompts: Mapping[str, str],
) -> Dict[str, str]:
    """Overlay prompt text overrides onto an existing item dictionary."""

    if not prompts:
        return dict(dictionary)
    out = {}
    for item, default in dictionary.items():
        out[item] = resolve_prompt(prompts, table_name, item, default)
    return out


def _try_json_or_python_mapping(text: str) -> Optional[Mapping[str, object]]:
    try:
        value = json.loads(text)
    except Exception:
        value = None
    if isinstance(value, dict):
        return value

    candidates = [text]
    match = re.search(r"=\s*(\{.*\})\s*$", text, flags=re.DOTALL)
    if match:
        candidates.append(match.group(1))
    brace_start = text.find("{")
    brace_end = text.rfind("}")
    if brace_start >= 0 and brace_end > brace_start:
        candidates.append(text[brace_start : brace_end + 1])

    for candidate in candidates:
        try:
            value = ast.literal_eval(candidate)
        except Exception:
            continue
        if isinstance(value, dict):
            return value
    return None


def _try_delimited_mapping(
    path: Path,
    text: str,
    default_table: Optional[str],
    include_bare_keys: bool,
) -> Optional[Dict[str, str]]:
    suffix = path.suffix.lower()
    delimiters = []
    if suffix == ".csv":
        delimiters = [","]
    elif suffix in {".tsv", ".tab"}:
        delimiters = ["\t"]
    else:
        delimiters = ["\t", ","]

    for delimiter in delimiters:
        try:
            rows = list(csv.DictReader(text.splitlines(), delimiter=delimiter))
        except Exception:
            continue
        if not rows or not rows[0]:
            continue
        fieldnames = {str(name).strip().lower() for name in rows[0].keys() if name}
        if "prompt" not in fieldnames:
            continue
        mapping: Dict[str, str] = {}
        for row in rows:
            lower = {str(key).strip().lower(): value for key, value in row.items() if key}
            prompt = str(lower.get("prompt", "")).strip()
            if not prompt:
                continue
            feature = str(lower.get("feature", "")).strip()
            item = str(lower.get("item", lower.get("column", lower.get("variable", "")))).strip()
            table = str(lower.get("table", lower.get("instrument", ""))).strip()
            key = feature or ("{}__{}".format(table, item) if table and item else item)
            if key:
                _add_prompt_mapping(mapping, key, prompt, default_table, include_bare_keys)
        if mapping:
            return mapping
    return None


def _try_line_mapping(
    text: str,
    default_table: Optional[str],
    include_bare_keys: bool,
) -> Dict[str, str]:
    mapping: Dict[str, str] = {}
    for raw_line in text.splitlines():
        line = raw_line.strip()
        if not line or line.startswith("#"):
            continue
        if "\t" in line:
            key, prompt = line.split("\t", 1)
        elif ":" in line:
            key, prompt = line.split(":", 1)
        else:
            continue
        key = key.strip().strip("\"'")
        prompt = prompt.strip().strip("\"'")
        if key and prompt:
            _add_prompt_mapping(mapping, key, prompt, default_table, include_bare_keys)
    return mapping


def _clean_mapping(
    mapping: Mapping[str, object],
    default_table: Optional[str],
    include_bare_keys: bool,
) -> Dict[str, str]:
    clean: Dict[str, str] = {}
    for key, value in mapping.items():
        if value is None:
            continue
        key_text = str(key).strip()
        value_text = str(value).strip()
        if key_text and value_text:
            _add_prompt_mapping(clean, key_text, value_text, default_table, include_bare_keys)
    return clean


def _add_prompt_mapping(
    mapping: Dict[str, str],
    key: str,
    prompt: str,
    default_table: Optional[str],
    include_bare_keys: bool,
) -> None:
    key = str(key).strip()
    prompt = str(prompt).strip()
    if not key or not prompt:
        return

    if default_table and not _is_qualified_key(key):
        qualified = "{}__{}".format(default_table, key)
        mapping[qualified] = prompt
        mapping[_normalize_key(qualified)] = prompt
        if not include_bare_keys:
            return

    mapping[key] = prompt
    mapping[_normalize_key(key)] = prompt


def _is_qualified_key(key: str) -> bool:
    return "__" in key or "." in key or "/" in key


def prompt_table_name_from_path(path: Path) -> str:
    stem = Path(path).stem.strip()
    stem = re.sub(
        r"([_-]?(prompt_dictionary|prompts|prompt|item_prompts|dictionary|items))$",
        "",
        stem,
        flags=re.IGNORECASE,
    ).strip("_-")
    return stem or Path(path).stem


def _normalize_key(value: str) -> str:
    return re.sub(r"[^a-z0-9]+", "_", str(value).strip().lower()).strip("_")
```
