## Supplementary material for "Auditable cross-instrument detection of unusual multivariate psychiatric response configurations using a semantically aligned covariance subspace": Code for tool: scales.py

```
"""Per-item scale loading and matching helpers.

A *scale file* is the third tool input alongside prompts and responses. It
declares, for each survey item, the measurement metadata the pipeline cannot
reliably infer from the raw response numbers:

- ``min`` / ``max`` — the item's valid response range.
- ``sentinels`` — reserved codes that mean "non-answer" (refused, don't know,
  not ascertained). These are mapped to missing *per item* before analysis.
- ``reverse`` — whether the item is reverse-coded.
- ``ceiling`` *(optional)* — whether the item participates in the pan-mild
  item-level ceiling audit (an explicit allowlist when present).
- ``embed`` *(optional)* — whether the item is embedded and analyzed at all (an
  explicit item-selection allowlist when present; see :func:`scales_use_embed`).

The canonical serialization is a CSV mirroring the prompt-file conventions::

    item,min,max,sentinels,reverse,ceiling,embed
    SAD_A,1,5,7;8;9,true,true,true
    LASTDR_A,1,8,0;97;98;99,false,false,false

This module only *loads and normalizes* scale files into a lookup; acting on
the values (cleaning sentinels, applying ranges, reversing) is the pipeline's
job. It parallels :mod:`survey_semantics.prompts` and reuses its generic key
helpers so scopes/namespacing resolve identically for prompts and scales.
"""

import csv
import re
from pathlib import Path
from typing import Dict, Iterable, Mapping, Optional, Set

try:  # TypedDict lives in typing on 3.8+, this codebase targets 3.11
    from typing import TypedDict
except ImportError:  # pragma: no cover - fallback for very old interpreters
    from typing_extensions import TypedDict  # type: ignore

from survey_semantics.prompts import (
    _is_qualified_key,
    _normalize_key,
    _try_json_or_python_mapping,
)


SCALE_FILE_SUFFIXES = {".csv", ".tsv", ".tab", ".txt", ".json", ".py"}

_ITEM_KEYS = ("item", "column", "variable", "feature")
_TABLE_KEYS = ("table", "instrument")
_MIN_KEYS = ("min", "minimum", "low", "lower")
_MAX_KEYS = ("max", "maximum", "high", "upper")
_SENTINEL_KEYS = ("sentinels", "sentinel", "missing", "missing_codes", "na_codes")
_REVERSE_KEYS = ("reverse", "reversed", "reverse_scored", "reverse_score")
_CEILING_KEYS = ("ceiling", "ceiling_check", "ceiling_eligible", "audit_ceiling")
_EMBED_KEYS = ("embed", "embedded", "include", "in_scope", "analyze")

_TRUTHY = {"true", "t", "yes", "y", "1"}
_FALSY = {"false", "f", "no", "n", "0", ""}


class ItemScale(TypedDict):
    """Resolved measurement spec for a single item.

    ``min``/``max`` are ``None`` when undeclared (the pipeline then falls back
    to observed ranges). ``sentinels`` is always a set (possibly empty) and
    ``reverse`` is always a bool. ``ceiling`` is ``None`` when undeclared
    (the pan-mild audit then falls back to the polytomous heuristic); ``True``
    means the item participates in the item-level ceiling check, ``False``
    excludes it. ``embed`` is ``None`` when undeclared (the item is analyzed,
    today's behavior); when *any* item declares it, ``embed`` becomes an explicit
    item-selection allowlist — only ``embed=True`` items are embedded and
    analyzed, ``embed=False`` (and undeclared) items are documented but excluded.
    """

    min: Optional[float]
    max: Optional[float]
    sentinels: Set[float]
    reverse: bool
    ceiling: Optional[bool]
    embed: Optional[bool]


def load_scale_sources(
    scale_file: Optional[Path] = None,
    scale_dir: Optional[Path] = None,
) -> Dict[str, ItemScale]:
    """Load item scales from one combined file, a scale directory, or both."""

    scales: Dict[str, ItemScale] = {}
    if scale_dir is not None:
        scales.update(load_scale_directory(scale_dir))
    if scale_file is not None:
        scales.update(load_scale_dictionary(scale_file))
    return scales


def load_scale_directory(path: Path) -> Dict[str, ItemScale]:
    """Load per-instrument scale files from a directory.

    Bare item keys inside each file are namespaced by the file's instrument
    stem, e.g. ``scales/srs02_scales.csv`` yields keys like ``srs02__parentreport_18``.
    """

    path = Path(path)
    if not path.exists() or not path.is_dir():
        raise ValueError("Scale directory does not exist or is not a directory: {}".format(path))

    scales: Dict[str, ItemScale] = {}
    loaded = 0
    for scale_file in sorted(path.iterdir()):
        if scale_file.name.startswith(".") or not scale_file.is_file():
            continue
        if scale_file.suffix.lower() not in SCALE_FILE_SUFFIXES:
            continue
        table_name = scale_table_name_from_path(scale_file)
        scales.update(
            load_scale_dictionary(
                scale_file,
                default_table=table_name,
                include_bare_keys=False,
            )
        )
        loaded += 1

    if loaded == 0:
        raise ValueError("No scale files found in {}".format(path))
    return scales


def load_scale_dictionary(
    path: Optional[Path],
    default_table: Optional[str] = None,
    include_bare_keys: bool = True,
) -> Dict[str, ItemScale]:
    """Load item scales from CSV/TSV, JSON, or a Python literal mapping.

    Supported formats:

    - CSV/TSV with an ``item`` column plus any of ``min``, ``max``,
      ``sentinels``, ``reverse`` (and optional ``table``).
    - JSON object / Python literal dict mapping ``item -> {min, max,
      sentinels, reverse}``.
    """

    if path is None:
        return {}
    path = Path(path)
    text = path.read_text(encoding="utf-8-sig").strip()
    if not text:
        return {}

    mapping = _try_json_or_python_mapping(text)
    if mapping is not None:
        cleaned = _clean_scale_mapping(mapping, default_table, include_bare_keys)
        if cleaned:
            return cleaned

    delimited = _try_delimited_scales(path, text, default_table, include_bare_keys)
    if delimited is not None:
        return delimited

    raise ValueError("Could not parse scale dictionary file: {}".format(path))


def resolve_scale(
    scales: Mapping[str, ItemScale],
    table_name: str,
    item_name: str,
) -> Optional[ItemScale]:
    """Return the best scale override for a table/item pair, or ``None``.

    Resolution order matches :func:`survey_semantics.prompts.resolve_prompt`:
    qualified keys first, then the bare item, then normalized fallbacks.
    """

    if not scales:
        return None
    candidates = [
        "{}__{}".format(table_name, item_name),
        "{}.{}".format(table_name, item_name),
        "{}/{}".format(table_name, item_name),
        item_name,
        _normalize_key("{}__{}".format(table_name, item_name)),
        _normalize_key(item_name),
    ]
    for key in candidates:
        if key in scales:
            return scales[key]
    return None


def scales_use_embed(scales: Optional[Mapping[str, ItemScale]]) -> bool:
    """Whether the scale file declares the ``embed`` column for any item.

    When no item declares ``embed`` (the column is absent), every declared item
    is embedded/analyzed — the pre-``embed`` behavior. When at least one item
    declares it, ``embed`` is an explicit allowlist (see :func:`is_item_embedded`).
    """

    if not scales:
        return False
    return any(scale.get("embed") is not None for scale in scales.values())


def is_item_embedded(
    scale: Optional[ItemScale],
    uses_embed: bool,
) -> bool:
    """Whether an item is embedded/analyzed under the ``embed`` allowlist rule.

    - No scale declared for the item → ``False`` (it is not in the declared set).
    - ``embed`` column absent from the file (``uses_embed=False``) → ``True``
      (all declared items are analyzed, the pre-``embed`` behavior).
    - ``embed`` column present → ``True`` only when this item's ``embed`` is
      ``True``; ``embed=False`` and undeclared (blank) items are excluded.
    """

    if scale is None:
        return False
    if not uses_embed:
        return True
    return scale.get("embed") is True


def _try_delimited_scales(
    path: Path,
    text: str,
    default_table: Optional[str],
    include_bare_keys: bool,
) -> Optional[Dict[str, ItemScale]]:
    suffix = path.suffix.lower()
    if suffix == ".csv":
        delimiters = [","]
    elif suffix in {".tsv", ".tab"}:
        delimiters = ["\t"]
    else:
        delimiters = ["\t", ","]

    for delimiter in delimiters:
        try:
            rows = list(csv.DictReader(text.splitlines(), delimiter=delimiter))
        except Exception:
            continue
        if not rows or not rows[0]:
            continue
        fieldnames = {str(name).strip().lower() for name in rows[0].keys() if name}
        if not (set(_ITEM_KEYS) & fieldnames):
            continue
        mapping: Dict[str, ItemScale] = {}
        for row in rows:
            lower = {str(key).strip().lower(): value for key, value in row.items() if key}
            item = _first(lower, _ITEM_KEYS)
            if not item:
                continue
            table = _first(lower, _TABLE_KEYS)
            scale: ItemScale = {
                "min": _parse_float(_first(lower, _MIN_KEYS)),
                "max": _parse_float(_first(lower, _MAX_KEYS)),
                "sentinels": _parse_sentinels(_first_value(lower, _SENTINEL_KEYS)),
                "reverse": _parse_bool(_first(lower, _REVERSE_KEYS)),
                "ceiling": _parse_optional_bool(_first_value(lower, _CEILING_KEYS)),
                "embed": _parse_optional_bool(_first_value(lower, _EMBED_KEYS)),
            }
            key = "{}__{}".format(table, item) if table and item else item
            _add_scale_mapping(mapping, key, scale, default_table, include_bare_keys)
        if mapping:
            return mapping
    return None


def _clean_scale_mapping(
    mapping: Mapping[str, object],
    default_table: Optional[str],
    include_bare_keys: bool,
) -> Dict[str, ItemScale]:
    clean: Dict[str, ItemScale] = {}
    for key, value in mapping.items():
        scale = _coerce_scale_value(value)
        if scale is None:
            continue
        key_text = str(key).strip()
        if key_text:
            _add_scale_mapping(clean, key_text, scale, default_table, include_bare_keys)
    return clean


def _coerce_scale_value(value: object) -> Optional[ItemScale]:
    if not isinstance(value, Mapping):
        return None
    lower = {str(key).strip().lower(): val for key, val in value.items()}
    return {
        "min": _parse_float(_first(lower, _MIN_KEYS)),
        "max": _parse_float(_first(lower, _MAX_KEYS)),
        "sentinels": _parse_sentinels(_first_value(lower, _SENTINEL_KEYS)),
        "reverse": _parse_bool(_first_value(lower, _REVERSE_KEYS)),
        "ceiling": _parse_optional_bool(_first_value(lower, _CEILING_KEYS)),
        "embed": _parse_optional_bool(_first_value(lower, _EMBED_KEYS)),
    }


def _add_scale_mapping(
    mapping: Dict[str, ItemScale],
    key: str,
    scale: Optional[ItemScale],
    default_table: Optional[str],
    include_bare_keys: bool,
) -> None:
    key = str(key).strip()
    if not key or scale is None:
        return

    if default_table and not _is_qualified_key(key):
        qualified = "{}__{}".format(default_table, key)
        mapping[qualified] = scale
        mapping[_normalize_key(qualified)] = scale
        if not include_bare_keys:
            return

    mapping[key] = scale
    mapping[_normalize_key(key)] = scale


def scale_table_name_from_path(path: Path) -> str:
    stem = Path(path).stem.strip()
    stem = re.sub(
        r"([_-]?(item_scales|scales|scale|ranges|scaling))$",
        "",
        stem,
        flags=re.IGNORECASE,
    ).strip("_-")
    return stem or Path(path).stem


def _first(lower: Mapping[str, object], keys: Iterable[str]) -> str:
    for key in keys:
        if key in lower and lower[key] is not None:
            text = str(lower[key]).strip()
            if text:
                return text
    return ""


def _first_value(lower: Mapping[str, object], keys: Iterable[str]) -> object:
    """Like :func:`_first` but returns the raw value (e.g. a list of sentinels
    from a JSON mapping) instead of stringifying it."""

    for key in keys:
        if key in lower and lower[key] is not None:
            value = lower[key]
            if isinstance(value, str) and not value.strip():
                continue
            return value
    return None


def _parse_float(value: object) -> Optional[float]:
    if value is None:
        return None
    text = str(value).strip()
    if not text:
        return None
    try:
        return float(text)
    except ValueError:
        return None


def _parse_bool(value: object, default: bool = False) -> bool:
    if isinstance(value, bool):
        return value
    text = str(value).strip().lower()
    if text in _TRUTHY:
        return True
    if text in _FALSY:
        return False
    return default


def _parse_optional_bool(value: object) -> Optional[bool]:
    """Like ``_parse_bool`` but returns ``None`` when the value is absent/blank,
    so callers can distinguish "not declared" from an explicit ``false``."""

    if isinstance(value, bool):
        return value
    if value is None:
        return None
    text = str(value).strip().lower()
    if not text:
        return None
    if text in _TRUTHY:
        return True
    if text in _FALSY:
        return False
    return None


def _parse_sentinels(value: object) -> Set[float]:
    out: Set[float] = set()
    if value is None:
        return out
    if isinstance(value, (list, tuple, set)):
        tokens: Iterable[object] = value
    else:
        tokens = re.split(r"[;,|\s]+", str(value))
    for token in tokens:
        text = str(token).strip()
        if not text:
            continue
        try:
            out.add(float(text))
        except ValueError:
            continue
    return out
```
