## Supplementary material for "Auditable cross-instrument detection of unusual multivariate psychiatric response configurations using a semantically aligned covariance subspace": Code for tool: conftest.py

```
"""Shared test fixtures.

The pipeline's only embedding backend is a local sentence-transformers model
(e.g. bge-m3), which is not available in CI and is offline-only by design — and
there is intentionally no TF-IDF/auto fallback. So tests patch the model call
with a deterministic, dependency-free fake encoder. This exercises all the
pipeline logic (PCA, projection, residualization, Mahalanobis, pan-mild, …)
without needing the real model, while keeping the real backend the *only*
supported one in production.
"""

import hashlib

import numpy as np
import pytest
from unittest import mock


def _fake_sentence_transformer_embeddings(texts, model_name=None, trust_remote_code=False):
    """Deterministic per-text vectors (seeded by a stable hash of the text)."""
    dim = 64
    out = np.zeros((len(texts), dim), dtype=float)
    for i, text in enumerate(texts):
        seed = int(hashlib.md5(str(text).encode("utf-8")).hexdigest()[:8], 16)
        out[i] = np.random.RandomState(seed).normal(size=dim)
    return out


@pytest.fixture(autouse=True)
def _patch_embedder():
    """Replace the real sentence-transformers call everywhere for the test run."""
    with mock.patch(
        "survey_semantics.embedding._sentence_transformer_embeddings",
        _fake_sentence_transformer_embeddings,
    ):
        yield
```
