## Supplementary material for "Auditable cross-instrument detection of unusual multivariate psychiatric response configurations using a semantically aligned covariance subspace": Code for tool: test_basis.py

```
"""Tests for the decoupled PCA basis stage (embed -> pca -> score)."""

from pathlib import Path

import numpy as np
import pandas as pd
import pytest

from survey_semantics.basis import (
    build_semantic_basis,
    load_semantic_basis,
    save_semantic_basis,
)
from survey_semantics.embedding import embed_item_prompts
from survey_semantics.io import SurveyTable
from survey_semantics.pipeline import AnalysisConfig, analyze_survey_table
from survey_semantics.scales import load_scale_sources


ITEMS = ["A", "B", "C", "D", "E"]


def _basis(items, **kw):
    emb = embed_item_prompts({it: "wording about item {}".format(it) for it in items})
    return build_semantic_basis(
        items=emb.items, embedding_vectors=emb.vectors,
        embedding_backend=emb.backend, embedding_model=emb.model_name,
        embedding_slug=emb.slug, **kw,
    )


def test_basis_npz_roundtrip(tmp_path):
    basis = _basis(["a", "b", "c"], d_null_permutations=5)
    path = tmp_path / "b.npz"
    save_semantic_basis(path, basis)
    back = load_semantic_basis(path)

    assert back.items == basis.items
    assert back.n_components == basis.n_components
    assert back.embedding_slug == basis.embedding_slug
    assert np.allclose(back.components, basis.components)
    assert np.allclose(back.eigenvalues, basis.eigenvalues)
    assert np.allclose(back.cumulative_variance, basis.cumulative_variance)
    assert np.allclose(back.parallel["null_mean"], basis.parallel["null_mean"])
    assert back.parallel["selected_d"] == basis.parallel["selected_d"]


def test_coordinates_for_reorders_and_validates():
    basis = _basis(["a", "b", "c"], d_null_permutations=0)
    out = basis.coordinates_for(["c", "a", "b"])
    assert np.allclose(out, basis.components[[2, 0, 1]])

    with pytest.raises(ValueError):       # missing item
        basis.coordinates_for(["a", "b", "c", "d"])
    with pytest.raises(ValueError):       # extra basis item -> partial projection
        basis.coordinates_for(["a", "b"])


def _table():
    rng = np.random.RandomState(0)
    data = {"id": ["S{}".format(i) for i in range(14)]}
    for it in ITEMS:
        data[it] = rng.randint(1, 6, size=14)
    df = pd.DataFrame(data)
    dictionary = {it: "wording about item {}".format(it) for it in ITEMS}
    dictionary["id"] = "id"
    return SurveyTable(name="t", path=Path("t.csv"), data=df, dictionary=dictionary)


def _config(tmp_path):
    scales = load_scale_sources(scale_file=_scales(tmp_path))
    return AnalysisConfig(
        compute_umap=False, item_scales=scales, d_selection_method="variance",
        d_null_permutations=0, id_col="id",
    )


def _scales(tmp_path):
    path = tmp_path / "t_scales.csv"
    path.write_text(
        "item,min,max,sentinels,reverse\n"
        + "".join("{},1,5,9,false\n".format(it) for it in ITEMS)
    )
    return path


def test_precomputed_basis_matches_inline(tmp_path):
    """embed -> pca -> score gives the same result as the one-shot analysis."""
    table = _table()
    config = _config(tmp_path)

    inline = analyze_survey_table(table, config)
    basis = _basis(ITEMS, d_null_permutations=0)
    viabasis = analyze_survey_table(table, config, basis=basis)

    assert inline.summary["semantic_basis_source"] == "computed_inline"
    assert viabasis.summary["semantic_basis_source"] == "precomputed"
    assert inline.summary["optimal_d"] == viabasis.summary["optimal_d"]
    assert np.allclose(
        inline.scores["Mahalanobis_Dist"].values,
        viabasis.scores["Mahalanobis_Dist"].values,
    )
    pc_cols = [c for c in inline.scores.columns if c.startswith("Semantic_PC")]
    assert pc_cols
    assert np.allclose(inline.scores[pc_cols].values, viabasis.scores[pc_cols].values)


def test_basis_item_mismatch_raises(tmp_path):
    """A basis fit on a different item set is rejected, not silently misused."""
    table = _table()
    config = _config(tmp_path)
    wrong_basis = _basis(ITEMS[:-1] + ["Z"], d_null_permutations=0)
    with pytest.raises(ValueError):
        analyze_survey_table(table, config, basis=wrong_basis)
```
