## Supplementary material for "Auditable cross-instrument detection of unusual multivariate psychiatric response configurations using a semantically aligned covariance subspace": Code for tool: test_combined.py

```
import tempfile
import unittest
from pathlib import Path

from survey_semantics.combined import build_combined_package_table
from survey_semantics.pipeline import AnalysisConfig, analyze_survey_table


class CombinedPackageTests(unittest.TestCase):
    def test_combines_questionnaires_and_applies_manual_reverse_config(self):
        with tempfile.TemporaryDirectory() as tmpdir:
            root = Path(tmpdir)
            (root / "a.tsv").write_text(
                "\n".join(
                    [
                        "\t".join(["subjectkey", "interview_age", "sex", "a1", "a2", "a3", "a4", "a5"]),
                        "\t".join(["GUID", "Age in months", "Sex", "Feels sad", "Feels calm", "Worries", "Sleeps poorly", "Avoids people"]),
                        "\t".join(["S1", "100", "M", "0", "3", "1", "0", "1"]),
                        "\t".join(["S2", "110", "F", "1", "2", "1", "1", "1"]),
                        "\t".join(["S3", "120", "M", "2", "1", "2", "1", "2"]),
                        "\t".join(["S4", "130", "F", "3", "0", "3", "2", "2"]),
                        "\t".join(["S5", "140", "M", "1", "1", "0", "1", "0"]),
                    ]
                ),
                encoding="utf-8",
            )
            (root / "b.tsv").write_text(
                "\n".join(
                    [
                        "\t".join(["subjectkey", "interview_age", "sex", "b1", "b2", "b3", "b4", "b5"]),
                        "\t".join(["GUID", "Age in months", "Sex", "Rigid routines", "Flexible with changes", "Sensory interests", "Social difficulty", "Repeats phrases"]),
                        "\t".join(["S1", "101", "M", "0", "3", "1", "1", "0"]),
                        "\t".join(["S2", "111", "F", "1", "2", "1", "1", "1"]),
                        "\t".join(["S3", "121", "M", "2", "1", "2", "2", "2"]),
                        "\t".join(["S4", "131", "F", "3", "0", "3", "3", "2"]),
                        "\t".join(["S5", "141", "M", "0", "1", "0", "0", "0"]),
                    ]
                ),
                encoding="utf-8",
            )
            reverse = root / "reverse.csv"
            reverse.write_text("table,item,reverse\na,a2,true\nb,b2,true\n", encoding="utf-8")

            combined = build_combined_package_table(
                root,
                reverse_config=reverse,
                min_nonmissing=2,
                auto_reverse=False,
            )
            self.assertEqual(len(combined.item_columns), 10)
            self.assertIn("a__a2", combined.reverse_items)
            self.assertIn("b__b2", combined.reverse_items)

            result = analyze_survey_table(
                combined.table,
                AnalysisConfig(
                    compute_umap=False,
                    min_rows=5,
                    min_items=5,
                    min_complete_fraction=0.2,
                    reverse_items=combined.reverse_items,
                ),
                item_columns=combined.item_columns,
            )
            self.assertEqual(result.summary["n_rows"], 5)
            self.assertEqual(result.summary["n_items"], 10)
            reversed_loadings = result.prompt_loadings[result.prompt_loadings["reverse_scored"]]
            self.assertEqual(set(reversed_loadings["item"]), {"a__a2", "b__b2"})

    def test_auto_reverse_flags_strong_within_instrument_anticorrelation(self):
        with tempfile.TemporaryDirectory() as tmpdir:
            root = Path(tmpdir)
            (root / "scale.tsv").write_text(
                "\n".join(
                    [
                        "\t".join(["subjectkey", "p1", "p2", "p3"]),
                        "\t".join(["GUID", "Feels tense", "Feels calm", "Worries often"]),
                        "\t".join(["S1", "0", "3", "0"]),
                        "\t".join(["S2", "1", "2", "1"]),
                        "\t".join(["S3", "2", "1", "2"]),
                        "\t".join(["S4", "3", "0", "3"]),
                        "\t".join(["S5", "1", "2", "1"]),
                    ]
                ),
                encoding="utf-8",
            )

            combined = build_combined_package_table(
                root,
                min_nonmissing=2,
                auto_reverse_corr_threshold=0.95,
                auto_reverse_min_pairwise_subjects=4,
                auto_reverse_min_pairwise_fraction=0.8,
            )

            self.assertIn("scale__p2", combined.reverse_items)
            inventory = combined.prompt_inventory.set_index("feature")
            self.assertEqual(inventory.loc["scale__p2", "reverse_source"], "auto_anticorrelation")
            self.assertFalse(combined.auto_reverse_warnings.empty)
            warnings = combined.auto_reverse_warnings
            negated = set(warnings["feature_negated"])
            self.assertIn("scale__p2", negated)
            self.assertIn("Feels calm", combined.auto_reverse_warning_text)
            self.assertIn("Feels tense", combined.auto_reverse_warning_text)


if __name__ == "__main__":
    unittest.main()
```
