## Supplementary material for "Auditable cross-instrument detection of unusual multivariate psychiatric response configurations using a semantically aligned covariance subspace": Code for tool: test_embedding.py

```
"""Tests for the decoupled item-embedding artifact (LLM step separated)."""

import numpy as np
import pytest

from survey_semantics.embedding import (
    ItemEmbeddings,
    embed_item_prompts,
    load_item_embeddings,
    save_item_embeddings,
)


def test_embed_item_prompts_and_npz_roundtrip(tmp_path):
    # Uses the deterministic fake encoder patched in conftest.
    emb = embed_item_prompts({"sad": "feel sad", "sleep": "sleep poorly"})
    assert emb.items == ["sad", "sleep"]
    assert emb.vectors.shape[0] == 2
    assert emb.backend == "sentence-transformers"

    path = tmp_path / "items.npz"
    save_item_embeddings(path, emb)
    back = load_item_embeddings(path)

    assert back.items == emb.items
    assert back.backend == emb.backend and back.slug == emb.slug
    assert np.allclose(back.vectors, emb.vectors)


def test_matrix_for_aligns_to_requested_order():
    emb = ItemEmbeddings(
        items=["a", "b", "c"],
        vectors=np.arange(9).reshape(3, 3).astype(float),
        backend="x", model_name="m", slug="s",
    )
    out = emb.matrix_for(["c", "a"])
    assert np.array_equal(out, np.array([[6.0, 7.0, 8.0], [0.0, 1.0, 2.0]]))


def test_matrix_for_raises_on_missing_item():
    emb = ItemEmbeddings(
        items=["a", "b"], vectors=np.zeros((2, 3)),
        backend="x", model_name="m", slug="s",
    )
    with pytest.raises(ValueError):
        emb.matrix_for(["a", "zzz"])
```
