## Supplementary material for "Auditable cross-instrument detection of unusual multivariate psychiatric response configurations using a semantically aligned covariance subspace": Code for tool: test_io.py

```
import tempfile
import unittest
from pathlib import Path

from survey_semantics.io import infer_item_columns, read_survey_table
from survey_semantics.prompts import load_prompt_dictionary, load_prompt_sources


class SurveyIOTests(unittest.TestCase):
    def test_reads_nda_dictionary_and_infers_items(self):
        with tempfile.TemporaryDirectory() as tmpdir:
            path = Path(tmpdir) / "toy.tsv"
            path.write_text(
                "\t".join(["subjectkey", "interview_age", "q1", "q2", "total_score"]) + "\n"
                + "\t".join(["GUID", "Age in months", "Feels calm", "Has trouble sleeping", "Total score"]) + "\n"
                + "\t".join(["A", "120", "yes", "0", "1"]) + "\n"
                + "\t".join(["B", "130", "no", "1", "1"]) + "\n"
                + "\t".join(["C", "140", "yes", "2", "3"]) + "\n"
                + "\t".join(["D", "150", "no", "1", "1"]) + "\n",
                encoding="utf-8",
            )
            table = read_survey_table(path)
            items = infer_item_columns(table, min_nonmissing=2)
            self.assertEqual(table.dictionary["q1"], "Feels calm")
            self.assertEqual(set(items), {"q1", "q2"})

    def test_prompt_dictionary_overrides_item_text(self):
        with tempfile.TemporaryDirectory() as tmpdir:
            root = Path(tmpdir)
            path = root / "scale.tsv"
            path.write_text(
                "\n".join(
                    [
                        "\t".join(["subjectkey", "q1", "q2"]),
                        "\t".join(["GUID", "Original calm wording", "Original sleep wording"]),
                        "\t".join(["A", "0", "1"]),
                        "\t".join(["B", "1", "0"]),
                    ]
                ),
                encoding="utf-8",
            )
            prompt_file = root / "prompts.txt"
            prompt_file.write_text(
                "{'scale__q1': 'Notebook prompt for calmness', 'q2': 'Notebook prompt for sleep'}",
                encoding="utf-8",
            )

            prompts = load_prompt_dictionary(prompt_file)
            table = read_survey_table(path, prompt_dictionary=prompts)

            self.assertEqual(table.dictionary["q1"], "Notebook prompt for calmness")
            self.assertEqual(table.dictionary["q2"], "Notebook prompt for sleep")

    def test_prompt_directory_namespaces_bare_item_keys_by_file_stem(self):
        with tempfile.TemporaryDirectory() as tmpdir:
            root = Path(tmpdir)
            prompt_dir = root / "prompts"
            prompt_dir.mkdir()
            (prompt_dir / "scale_prompts.csv").write_text(
                "item,prompt\nq1,Scale-specific prompt for q1\nq2,Scale-specific prompt for q2\n",
                encoding="utf-8",
            )
            (prompt_dir / "other_prompts.csv").write_text(
                "item,prompt\nq1,Other instrument prompt for q1\n",
                encoding="utf-8",
            )
            prompts = load_prompt_sources(prompt_dir=prompt_dir)

            scale_path = root / "scale.tsv"
            scale_path.write_text(
                "\n".join(
                    [
                        "\t".join(["subjectkey", "q1", "q2"]),
                        "\t".join(["GUID", "Original q1", "Original q2"]),
                        "\t".join(["A", "0", "1"]),
                        "\t".join(["B", "1", "0"]),
                    ]
                ),
                encoding="utf-8",
            )
            other_path = root / "other.tsv"
            other_path.write_text(
                "\n".join(
                    [
                        "\t".join(["subjectkey", "q1"]),
                        "\t".join(["GUID", "Original other q1"]),
                        "\t".join(["A", "0"]),
                        "\t".join(["B", "1"]),
                    ]
                ),
                encoding="utf-8",
            )

            scale = read_survey_table(scale_path, prompt_dictionary=prompts)
            other = read_survey_table(other_path, prompt_dictionary=prompts)

            self.assertEqual(scale.dictionary["q1"], "Scale-specific prompt for q1")
            self.assertEqual(scale.dictionary["q2"], "Scale-specific prompt for q2")
            self.assertEqual(other.dictionary["q1"], "Other instrument prompt for q1")


if __name__ == "__main__":
    unittest.main()
```
