## Supplementary material for "Auditable cross-instrument detection of unusual multivariate psychiatric response configurations using a semantically aligned covariance subspace": Code for tool: test_pipeline.py

```
import tempfile
import unittest
from pathlib import Path

import numpy as np

from survey_semantics.io import read_survey_table
from survey_semantics.pipeline import AnalysisConfig, analyze_survey_table, select_component_count
from survey_semantics.embedding import (
    OFFLINE_ENV,
    enforce_local_ai_offline_policy,
    install_outbound_socket_blocker,
)


class PipelineTests(unittest.TestCase):
    def test_offline_embedding_env_policy(self):
        enforce_local_ai_offline_policy()
        for key, value in OFFLINE_ENV.items():
            self.assertEqual(__import__("os").environ[key], value)

    def test_socket_blocker_blocks_outbound_connections(self):
        install_outbound_socket_blocker()
        with self.assertRaises(RuntimeError):
            __import__("socket").create_connection(("example.com", 443), timeout=0.01)

    def test_component_count_uses_max_when_threshold_not_reached(self):
        selected, reached = select_component_count(
            np.asarray([0.18, 0.31, 0.42]),
            variance_threshold=0.80,
            n_components=3,
        )
        self.assertEqual(selected, 3)
        self.assertFalse(reached)

    def test_component_count_uses_first_threshold_crossing(self):
        selected, reached = select_component_count(
            np.asarray([0.18, 0.81, 0.91]),
            variance_threshold=0.80,
            n_components=3,
        )
        self.assertEqual(selected, 2)
        self.assertTrue(reached)

    def test_analyzes_small_semantic_survey(self):
        with tempfile.TemporaryDirectory() as tmpdir:
            path = Path(tmpdir) / "survey.tsv"
            lines = [
                ["subjectkey", "interview_age", "sex", "sad", "sleep", "worry", "focus", "social"],
                [
                    "GUID",
                    "Age in months",
                    "Sex of subject",
                    "Feels sad or down",
                    "Has trouble sleeping",
                    "Worries too much",
                    "Has trouble focusing",
                    "Avoids social contact",
                ],
            ]
            rows = [
                ["S1", "100", "M", "0", "0", "1", "0", "1"],
                ["S2", "110", "F", "1", "1", "1", "0", "1"],
                ["S3", "120", "M", "0", "1", "0", "1", "0"],
                ["S4", "130", "F", "2", "2", "2", "1", "2"],
                ["S5", "140", "M", "0", "0", "0", "0", "0"],
                ["S6", "150", "F", "1", "0", "1", "1", "1"],
                ["S7", "160", "M", "2", "2", "1", "2", "1"],
                ["S8", "170", "F", "0", "1", "0", "0", "0"],
                ["S9", "180", "M", "1", "2", "2", "1", "2"],
                ["S10", "190", "F", "0", "0", "0", "1", "0"],
            ]
            text = "\n".join("\t".join(row) for row in lines + rows)
            path.write_text(text, encoding="utf-8")

            table = read_survey_table(path)
            result = analyze_survey_table(
                table,
                AnalysisConfig(
                    compute_umap=False,
                    min_rows=5,
                    min_items=5,
                    covariates=["interview_age", "sex"],
                ),
            )

            self.assertEqual(result.summary["n_rows"], 10)
            self.assertEqual(result.summary["n_items"], 5)
            self.assertEqual(result.summary["embedding"], "sentence-transformers")
            self.assertIn("sentence-transformers", result.summary["embedding_slug"])
            self.assertIn("Mahalanobis_Dist", result.scores.columns)
            self.assertGreaterEqual(result.summary["optimal_d"], 1)
            self.assertIn("PC1", result.prompt_loadings.columns)
            self.assertIn("prompt", result.prompt_loadings.columns)
            self.assertFalse(result.item_weights.empty)
            self.assertFalse(result.stability.empty)
            self.assertFalse(result.dimension_selection.empty)
            self.assertFalse(result.dimension_methods.empty)
            self.assertIn("parallel_analysis", set(result.dimension_methods["method"]))
            self.assertIn("d_parallel_analysis", result.summary)
            self.assertFalse(result.drivers.empty)
            self.assertTrue(result.case_studies)
            self.assertFalse(result.case_study_label_map.empty)
            first_report = next(iter(result.case_studies.values()))
            self.assertIn("Semantic Survey Case Study", first_report)
            self.assertIn("Outlier label: Outlier #", first_report)
            self.assertNotIn("Subject ID:", first_report)
            self.assertNotIn("S1", first_report)
            self.assertTrue(all("outlier" in filename for filename in result.case_studies))

    def test_embed_column_restricts_analyzed_item_set(self):
        # Six declared items; the scale file marks one embed=false. Only the five
        # embed=true items must be analyzed (embed=false is documented, excluded).
        with tempfile.TemporaryDirectory() as tmpdir:
            path = Path(tmpdir) / "survey.tsv"
            lines = [
                ["subjectkey", "interview_age", "sex", "sad", "sleep", "worry", "focus", "social", "height"],
                ["GUID", "Age in months", "Sex", "Feels sad", "Trouble sleeping",
                 "Worries too much", "Trouble focusing", "Avoids social contact", "Body height"],
            ]
            rng = np.random.default_rng(0)
            rows = [["S{}".format(i), str(100 + i), "M" if i % 2 else "F"]
                    + [str(int(v)) for v in rng.integers(0, 3, size=6)]
                    for i in range(12)]
            path.write_text("\n".join("\t".join(r) for r in lines + rows), encoding="utf-8")

            scale_path = Path(tmpdir) / "survey_scales.csv"
            scale_path.write_text(
                "item,min,max,sentinels,reverse,embed\n"
                "sad,0,2,,false,true\n"
                "sleep,0,2,,false,true\n"
                "worry,0,2,,false,true\n"
                "focus,0,2,,false,true\n"
                "social,0,2,,false,true\n"
                "height,0,2,,false,false\n",  # documented but excluded
                encoding="utf-8",
            )
            from survey_semantics.scales import load_scale_sources
            table = read_survey_table(path)
            result = analyze_survey_table(
                table,
                AnalysisConfig(
                    compute_umap=False, min_rows=5, min_items=5,
                    covariates=["interview_age", "sex"],
                    item_scales=load_scale_sources(scale_file=scale_path),
                ),
            )
            self.assertEqual(result.summary["n_items"], 5)
            self.assertNotIn("height", set(result.item_weights["item"]))

    def test_max_d_selection_uses_all_evaluated_components(self):
        with tempfile.TemporaryDirectory() as tmpdir:
            path = Path(tmpdir) / "survey.tsv"
            lines = [
                ["subjectkey", "q1", "q2", "q3", "q4", "q5"],
                ["GUID", "Sad mood", "Poor sleep", "Excess worry", "Inattention", "Social avoidance"],
            ]
            rows = [
                ["S1", "0", "0", "1", "0", "1"],
                ["S2", "1", "1", "1", "0", "1"],
                ["S3", "0", "1", "0", "1", "0"],
                ["S4", "2", "2", "2", "1", "2"],
                ["S5", "0", "0", "0", "0", "0"],
                ["S6", "1", "0", "1", "1", "1"],
                ["S7", "2", "2", "1", "2", "1"],
                ["S8", "0", "1", "0", "0", "0"],
                ["S9", "1", "2", "2", "1", "2"],
                ["S10", "0", "0", "0", "1", "0"],
            ]
            path.write_text("\n".join("\t".join(row) for row in lines + rows), encoding="utf-8")

            result = analyze_survey_table(
                read_survey_table(path),
                AnalysisConfig(
                    compute_umap=False,
                    min_rows=5,
                    min_items=5,
                    d_selection_method="max",
                ),
            )

            self.assertEqual(result.summary["d_selection_method"], "max")
            self.assertEqual(result.summary["optimal_d"], result.summary["components_evaluated"])
            selected = result.dimension_methods[result.dimension_methods["method"] == "all_components"]
            self.assertTrue(bool(selected.iloc[0]["used_for_scores"]))


if __name__ == "__main__":
    unittest.main()
```
