## Supplementary material for "Auditable cross-instrument detection of unusual multivariate psychiatric response configurations using a semantically aligned covariance subspace": Code for tool: test_scales_weights.py

```
"""Tests for the scale-file loader and weighted statistics (Part 1 features)."""

import numpy as np
import pandas as pd
from sklearn.covariance import LedoitWolf

from survey_semantics.io import coerce_response_frame, load_weights_file
from survey_semantics.pipeline import (
    _declared_range,
    _observed_item_ranges,
    ceiling_flags,
    ceiling_item_mask,
    mahalanobis_distances,
    normalize_responses,
    residualize,
)
from survey_semantics.scales import (
    is_item_embedded,
    load_scale_sources,
    resolve_scale,
    scales_use_embed,
)


def _write(tmp_path, name, text):
    path = tmp_path / name
    path.write_text(text)
    return path


# ── scale loader ────────────────────────────────────────────────────────────

def test_scale_csv_roundtrip_and_per_item_sentinels(tmp_path):
    path = _write(
        tmp_path,
        "x_scales.csv",
        "item,min,max,sentinels,reverse\n"
        "SAD_A,1,5,7;8;9,true\n"
        "LASTDR_A,1,8,0;97;98;99,false\n",
    )
    scales = load_scale_sources(scale_file=path)
    sad = resolve_scale(scales, "x", "SAD_A")
    last = resolve_scale(scales, "x", "LASTDR_A")
    assert sad == {"min": 1.0, "max": 5.0, "sentinels": {7.0, 8.0, 9.0},
                   "reverse": True, "ceiling": None, "embed": None}
    # 7/8 are *valid* answers for LASTDR_A — its sentinels differ per item.
    assert last["sentinels"] == {0.0, 97.0, 98.0, 99.0}
    assert last["reverse"] is False


def test_scale_json_dict_of_dicts_list_sentinels(tmp_path):
    path = _write(tmp_path, "x_scales.json", '{"FOO": {"min": 1, "max": 4, "sentinels": [7, 8, 9], "reverse": true}}')
    scales = load_scale_sources(scale_file=path)
    assert resolve_scale(scales, "x", "FOO")["sentinels"] == {7.0, 8.0, 9.0}


def test_embed_column_absent_means_all_declared_analyzed(tmp_path):
    # No `embed` column -> embed is None everywhere, allowlist off, all kept.
    path = _write(tmp_path, "x_scales.csv",
                  "item,min,max,sentinels,reverse\nA,1,5,9,false\nB,1,5,9,false\n")
    scales = load_scale_sources(scale_file=path)
    assert scales_use_embed(scales) is False
    assert resolve_scale(scales, "x", "A")["embed"] is None
    assert is_item_embedded(resolve_scale(scales, "x", "A"), uses_embed=False) is True


def test_embed_column_is_an_allowlist(tmp_path):
    path = _write(tmp_path, "x_scales.csv",
                  "item,min,max,sentinels,reverse,embed\n"
                  "A,1,5,9,false,true\n"
                  "B,1,5,9,false,false\n"
                  "C,1,5,9,false,\n")  # blank embed, file otherwise uses it
    scales = load_scale_sources(scale_file=path)
    assert scales_use_embed(scales) is True
    uses = True
    assert is_item_embedded(resolve_scale(scales, "x", "A"), uses) is True   # embed=true
    assert is_item_embedded(resolve_scale(scales, "x", "B"), uses) is False  # embed=false
    assert is_item_embedded(resolve_scale(scales, "x", "C"), uses) is False  # blank -> excluded
    # An item with no declared scale at all is never embedded.
    assert is_item_embedded(resolve_scale(scales, "x", "ZZZ"), uses) is False


def test_scale_directory_namespaces_by_stem(tmp_path):
    d = tmp_path / "scales"
    d.mkdir()
    (d / "srs02_scales.csv").write_text("item,min,max,sentinels,reverse\nQ1,1,5,9,true\n")
    scales = load_scale_sources(scale_dir=d)
    assert resolve_scale(scales, "srs02", "Q1") is not None
    assert "srs02__Q1" in scales


# ── per-item sentinel coercion ──────────────────────────────────────────────

def test_coerce_frame_per_column_sentinels():
    df = pd.DataFrame({"A": [1, 2, 7, 8], "B": [7, 1, 2, 3]})
    out = coerce_response_frame(df, ["A", "B"], sentinels={"A": {7, 8}, "B": set()})
    assert np.isnan(out["A"].iloc[2]) and np.isnan(out["A"].iloc[3])
    assert out["B"].tolist() == [7, 1, 2, 3]  # empty sentinel set keeps 7


# ── declared ranges ─────────────────────────────────────────────────────────

def test_declared_range_prefers_valid_min_max():
    assert _declared_range({"min": 1, "max": 5}) == (1.0, 5.0)
    assert _declared_range({"min": 3, "max": 3}) is None  # non-increasing -> fallback
    assert _declared_range(None) is None


def test_observed_ranges_uses_declared_when_present():
    raw = pd.DataFrame({"X": [1.0, 2.0, 3.0]})
    assert _observed_item_ranges(raw, raw, [(1.0, 5.0)]) == [(1.0, 5.0)]
    assert _observed_item_ranges(raw, raw, [None]) == [(1.0, 3.0)]


# ── weights loader ──────────────────────────────────────────────────────────

def test_load_weights_file_with_and_without_header(tmp_path):
    with_header = _write(tmp_path, "w1.csv", "weight\n5423.3\n3832.2\n3422.7\n")
    no_header = _write(tmp_path, "w2.csv", "5423.3\n3832.2\n3422.7\n")
    a = load_weights_file(with_header)
    b = load_weights_file(no_header)
    assert a.shape == (3,) and b.shape == (3,)
    assert np.allclose(a, [5423.3, 3832.2, 3422.7])
    assert np.allclose(a, b)


# ── weighted statistics match the reference formulas ────────────────────────

def test_weighted_residualize_matches_wls():
    rng = np.random.RandomState(0)
    n, d, k = 200, 4, 2
    y = rng.normal(size=(n, d))
    cov = rng.normal(size=(n, k))
    w = np.abs(rng.gamma(2.0, 1000.0, size=n)) + 1.0

    wn = w / w.sum()
    design = np.column_stack([np.ones(n), cov])
    sw = np.sqrt(wn)[:, None]
    beta, _, _, _ = np.linalg.lstsq(design * sw, y * sw, rcond=None)
    expected = y - design @ beta

    assert np.allclose(residualize(y, cov, weights=w), expected, atol=1e-10)


def test_weighted_mahalanobis_matches_weighted_scatter():
    rng = np.random.RandomState(1)
    n, d = 300, 5
    p = rng.normal(size=(n, d))
    w = np.abs(rng.gamma(2.0, 500.0, size=n)) + 1.0

    wn = w / w.sum()
    mu = (wn[:, None] * p).sum(axis=0)
    diff = p - mu
    n_eff = 1.0 / (wn ** 2).sum()
    scatter = (diff * wn[:, None]).T @ diff * (n_eff / (n_eff - 1.0))
    alpha = LedoitWolf().fit(p).shrinkage_
    target = (np.trace(scatter) / d) * np.eye(d)
    cov_inv = np.linalg.inv((1 - alpha) * scatter + alpha * target)
    expected = np.sqrt(np.sum(diff @ cov_inv * diff, axis=1))

    assert np.allclose(mahalanobis_distances(p, weights=w), expected, atol=1e-9)


def test_unweighted_paths_unchanged():
    rng = np.random.RandomState(2)
    p = rng.normal(size=(120, 4))
    lw = LedoitWolf().fit(p)
    expected = np.sqrt(np.maximum(lw.mahalanobis(p), 0.0))
    assert np.allclose(mahalanobis_distances(p), expected, atol=1e-9)


# ── pan-mild / ceiling audit ────────────────────────────────────────────────

def test_ceiling_mask_polytomous_fallback_excludes_binary():
    # No scales declare a ceiling flag -> fall back to polytomous items (>=3 levels).
    cols = ["poly", "binary"]
    ranges = [(1.0, 5.0), (0.0, 1.0)]
    mask = ceiling_item_mask(cols, ranges, item_scales=None, min_levels=3)
    assert mask.tolist() == [True, False]


def test_ceiling_mask_explicit_allowlist_overrides_polytomy():
    # Both items are polytomous, but the scale file declares which are valid.
    cols = ["sad", "lastdr"]
    ranges = [(1.0, 5.0), (1.0, 8.0)]
    scales = {
        "sad": {"min": 1, "max": 5, "sentinels": set(), "reverse": True, "ceiling": True},
        "lastdr": {"min": 1, "max": 8, "sentinels": set(), "reverse": False, "ceiling": False},
    }
    mask = ceiling_item_mask(cols, ranges, item_scales=scales, min_levels=3)
    assert mask.tolist() == [True, False]  # lastdr excluded despite being polytomous


def test_ceiling_flags_severity_oriented():
    response_norm = np.array([
        [1.0, 1.0],    # item 0 at +1 -> at ceiling
        [0.5, 1.0],    # item 0 below; item 1 not in mask
        [-1.0, -1.0],  # neither
        [0.999, 0.0],  # exactly at cutoff -> at ceiling
    ])
    mask = np.array([True, False])  # only item 0 is checked
    assert ceiling_flags(response_norm, mask).tolist() == [True, False, False, True]


def test_ceiling_matches_reverse_then_normalize_for_reverse_items():
    # A reverse-scored 1..5 item: max *symptom* severity is the RAW MIN (=1).
    raw_min, raw_max = 1.0, 5.0
    responses = np.array([[raw_min], [raw_max], [3.0]])
    norm = normalize_responses(responses, [(raw_min, raw_max)])
    norm[:, 0] *= -1.0  # reverse -> severity-oriented
    flags = ceiling_flags(norm, np.array([True]))
    assert flags.tolist() == [True, False, False]


def test_ceiling_flags_empty_mask():
    response_norm = np.ones((3, 2))
    assert ceiling_flags(response_norm, np.array([False, False])).tolist() == [False, False, False]
```
